# Comparative efficacy of different eating patterns in the management of type 2 diabetes and prediabetes: An arm-based Bayesian network meta-analysis

**DOI:** 10.1101/2022.05.30.22275766

**Authors:** Ben-tuo Zeng, Hui-qing Pan, Feng-dan Li, Zhen-yu Ye, Yang Liu, Ji-wei Du

## Abstract

**Aims/Introduction:** Diet therapy is a vital approach to manage type 2 diabetes and prediabetes. However, the comparative efficacy of different eating patterns is not clear enough. We aimed to compare the efficacy of various eating patterns for glycemic control, anthropometrics, and serum lipid profiles in the management of type 2 diabetes and prediabetes.

**Materials and Methods:** We conducted a network meta-analysis using arm-based Bayesian methods and random effect models, and drew the conclusions using the partially contextualized framework. We searched twelve databases and yielded 9,534 related references, where 107 studies were eligible, comprising 8,909 participants.

**Results:** Eleven diets were evaluated for fourteen outcomes. Caloric restriction was ranked as the best pattern for weight loss (SUCRA 86.8%) and waist circumference (82.2%), low-carbohydrate diets for body mass index (81.6%) and high-density lipoprotein (84.0%), and low-glycemic-index diets for total cholesterol (87.5%) and low-density lipoprotein (86.6%). Other interventions showed some superiorities, but were of imprecision due to insufficient participants and needed further investigation. The attrition rates of interventions were similar. Meta-regression suggested that macronutrients, energy intake, and weight may modify outcomes differently. The evidence was of moderate-to-low quality, and 38.2% of the evidence items met the minimal clinically important differences.

**Conclusions:** The selection and development of dietary strategies for diabetic/prediabetic patients should depend on their holistic conditions, i.e., serum lipids profiles, glucometabolic patterns, weight and blood pressures. It is recommended to identify the most critical and urgent metabolic indicator to control for one specific patient, and then choose the most appropriate eating pattern accordingly.

## 1. Introduction

It was estimated that 10.5% of people aged 20-75 suffered from diabetes mellitus globally, where over 90% were type 2 diabetes (T2DM) ^1^. They spend about 966 billion US dollars of health expenditures per year^1^. Since T2DM has proven to be preventable and controllable^2^, the remission of a prediabetic state (PreD), or impaired glucose tolerance (IGT), was also concerned and included in the comprehensive prevention of T2DM incidence.

Beyond medications, lifestyle management is more cost-effective for T2DM/PreD patients with strong clinical evidence^3-5^, where eating patterns play the leading role. Various patterns of different nutrients and food groups have been investigated and applied to T2DM/PreD treatment and management, from the very high-fat diet in the 18^th^ century^6^ to the pattern recommended by American Diabetes Association (ADA) in 2003^7^. From an evidence-based perspective, hundreds of random controlled trials (RCT), cohorts, and related systematic reviews have quantified the efficacy of popular and widely-used eating patterns^8-13^.

However, there are variances in the effectiveness of the diets across different outcomes, e.g., blood glucose, weight, and cardiovascular risk factors. Diabetes Canada guidelines^4^ summarized the properties of dietary interventions, pointing out the differences among diets. Consequently, current guidelines strongly recommend an individualized medical nutrition therapy under the supervision of dietitians and multidisciplinary professionals^3-5^. However, how to choose and apply appropriate dietary patterns for professionals remains to be a question, due to the lack of direct evidence comparing relative efficacy of the interventions. Whether a specific diet is proper for an individual with specific laboratory profiles and situations is not clear enough, though high-quality evidence of several patterns has been drawn.

It is not cost-effective to carry out multi-arm trials directly comparing several diets. Thus, it is crucial to conduct a network meta-analysis to synthesize current evidence. Previous network meta-analyses^14, 15^ have assessed a number of patterns, but the authors only included a limited number of studies and outcomes. Furthermore, short-term trials were not considered in the analyses, but a short-term effect may be more common for some patterns^16^. Therefore, this study aimed to evaluate the relative efficacy of different eating patterns on glycemic control, anthropometrics and serum lipid profiles in the management of T2DM/PreD patients, and conclude evidence to promote clinical decision-making.

## 2. Materials & Methods

### 2.1. Study Design

We conducted an arm-based Bayesian network meta-analysis of randomized controlled trials, following the Cochrane Handbook^17^. We reported results according to the Preferred Reporting Items for Systematic reviews and Meta-Analyses Incorporating Network Meta-analysis (PRISMA-NMA)^18^. A protocol was prepared and registered a priori in PROSPERO (CRD42021278268).

### 2.2. Eligibility Criteria

We selected peer-reviewed articles and thesis according to the PICOS principle. Eligibility criteria are displayed in Table 1.

**Table 1.** Eligibility criteria

| Inclusion |  | Exclusion |  |
| --- | --- | --- | --- |
| Type | Criteria | Type | Criteria |
| P | Adults with type 2 diabetes mellitus or prediabetes | I | Any prescribed between-group |
| I | Contain at least one arm of the interventions as follows: caloric restriction (CR), high-fiber diet (fiber), Dietary Approaches to Stop Hypertension (DASH), high-protein diet (HPD), high-fat diet (HFD), low-carbohydrate diet (LCD), low-glycemic-index diet (LGID), Mediterranean diet (Med), Nordic diet (ND), Paleolithic diet (Paleo), Portfolio diet (Pfd), and vegetarian/vegan/plant-based diet (VD). The macronutrients and food group intake can be as prescribed or as actual. |  | difference on exercise, antihyperglycemic medications, insulin injection, or other co-interventions; added a single supplement, or single specified food which did not provide macronutrients; or use meal replacement to provide an appreciable percentage of energy intake; or total energy intake (TEI) < 800 kcal/d (3.3 MJ/d); or the adjustment of intervention during the trial; |
| C | Contained standard diabetes diet, e.g. ADA 2003 diet<sup>7</sup>; *ad libitum*; general nutrition counselling; or placebo (no intervention); or contain two or more intervention arms. |  |  |
| O | Reported at least one outcome as follows, where fasting plasma glucose (FPG) was the primary outcome of this meta-analysis: glycemic control, including FPG, glycated hemoglobin (HbA<sub>1c</sub>), fasting insulin (FIns) and insulin resistance (IR); anthropometrics, including weight, body mass index (BMI), waist circumference (WC), waist-to-hip | Durati<br>on | less than four weeks or one month for parallel RCTs or any phase of crossover RCTs; or intermittent intervention |
| Type | Criteria | Type | Criteria |
|  | ratio (WHR), and body fat rate (BFR), systolic blood pressure (SBP) and diastolic blood pressure (DBP); serum lipid profiles, including triacylglycerol (TG), total cholesterol (TC), low-density lipoprotein cholesterol (LDL) and high-density lipoprotein cholesterol (HDL); renal function, including serum creatinine, serum urea, serum uric acid and (estimated) glomerular filtration rate; other dichotomous outcomes, including attrition rate, remission of T2DM, incidence of hypoglycemia, incidence of drug or insulin discontinuation, incidence of T2DM from PreD. | S | single-arm or self-controlled trials |
|  |  | Other | Data availability: trials not completed, or without data analysis and published reports; or articles with inappropriate or insufficient data |
| S | Randomized controlled trials (RCT) |  |  |
| Other | Language: English or Chinese |  |  |

P, participants; I, interventions; C, comparators; O, outcomes; S, study types.

### 2.3. Search Strategy

We conducted searches of databases and trial registers, including PubMed, Web of Science, Embase, CINAHL and Open Dissertation, ProQuest, Scopus, Global Index Medicus, Cochrane Central Register of Controlled Trials, Clinicaltrials.gov, SinoMed, WanFang Med, and CNKI. All publications from the inception to 13 October 2021 were initially retrieved. An updated search was conducted on March 17, 2022 using Scopus and Google Scholar to identify the latest relevant articles. Full search strategy can be found in File S1.

### 2.4. Data Selection and Extraction

All references identified from the search were imported into EndNote 20 (Clarivate, PA, USA) to move duplicates. After automatic exclusion by filtering title using excluding terms, reviewers (B.-T.Z., H.-Q.P., and F.-D.L.) assessed the eligibility in the order of title, abstract and full text. Each reference was decided independently by at least two reviewers, and arisen discrepancies were discussed and decided by the authors together.

We used MySQL 8.0 (Oracle Corporation, TX, USA) for data extraction and management, and critical information was extracted (see File S2 for fields in MySQL tables). Two authors (B.-T.Z. and Z.-Y.Y.) independently extracted the data and checked the consistency. R 4.1.3 (R Foundation, Vienna, Austria) and Microsoft Excel 2019 (Microsoft Corporation, WA, USA) were used for data conversion and imputation. For continuous outcomes, we calculated the change from baseline and its standard deviation (SD) if not reported by the article. Correlation coefficients for changes from baseline and for crossover RCTs were estimated using reported SDs from included studies (File S3). Median and interquartile range was converted into mean and SD using methods from Luo^19^ and Wan^20^ after testing for skewness using methods from Shi et al.^21^. WebPlot Digitizer^22^ was applied for extracting data from figures. Ultimately, R package “mice”^23^ was used for the imputation of missing values of covariates for meta-regression.

### 2.5. Risk of Bias Assessment

The Risk of Bias 2 tool^24^ and Risk of Bias 2 for crossover trials^25^ were employed to assess the risk of bias (RoB) of parallel and crossover RCTs, respectively. Two reviewers (B.-T.Z. and H.-Q.P.) assessed the RoBs independently, with all arisen divergences discussed and reached consensuses.

### 2.6. Data Synthesis

Our study synthesized evidence through an arm-based Bayesian network meta-analysis in a random effect model. We use R package “gemtc” 1.0-1 for meta-analysis, inconsistency test, heterogeneity test, meta-regression, and sensitivity analysis^26, 27^. Markov chain Monte Carlo sampling was performed using JAGS 4.3.0 via R package “rjags” 4.12^28, 29^. Comparison-adjusted funnel plots, Egger’s test, and Begg’s test were performed to detect publication bias under a frequentist framework and random effect model using R package “netmeta” 2.1-0 and “metafor” 3.4-0^30, 31^.

Continuous outcomes were presented as mean difference (MD) or difference in percentage change from baseline (Percentage MD, PMD, for fasting insulin and insulin resistance) and 95% credible intervals (95% CrI), while relative risk (RR) and 95% CrI were for dichotomous variables.

### 2.7. Quality of the Evidence

We rated the quality of evidence of comparisons of experimental diets and control diets based on the GRADE Working Group’s network meta-analysis evidence rating strategies^32^ and the GRADE handbook^33^. Conclusions were drawn according to the partially contextualized framework by the GRADE workgroup^34^, where minimal clinically important differences (MCID) and thresholds for moderate and large beneficial/harm effects were identified based on previous studies^13, 35-37^ and consensuses among reviewers.

## 3. Results

We identified 9,358 publications and registrations from the initial search, and 176 from the updated search. 111 publications^38-148^ were eligible, where 107 independent studies were identified (Figure 1). All items excluded via full-text screening and their reason for exclusion were listed in File S4.

**Figure 1.**
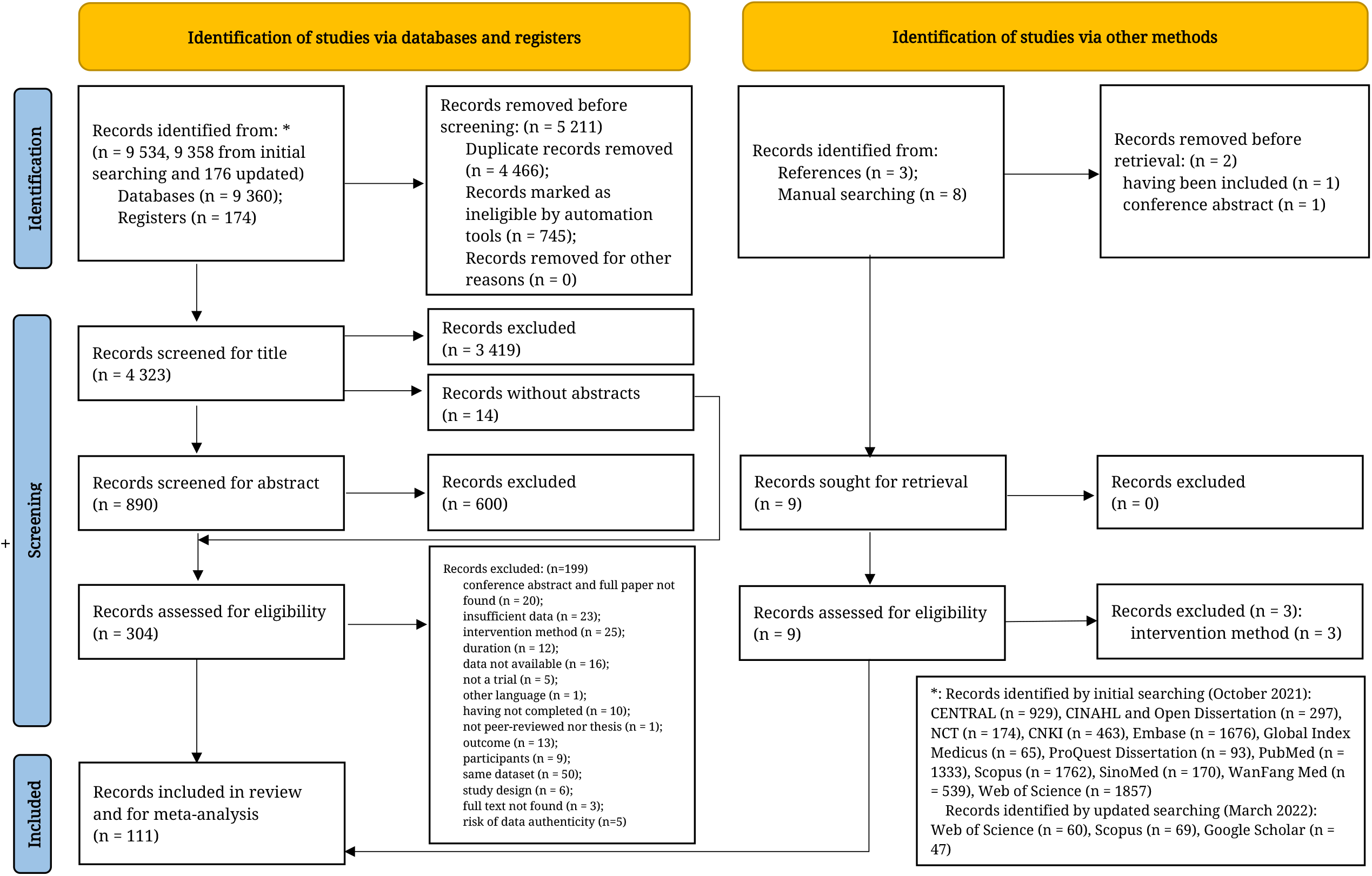
PRISMA flowchart of data selection

Among our prescribed outcomes, data of FPG, HbA_1c_, FIns, IR, weight, BMI, WC, SBP, DBP, TG, TC, HDL, LDL, and attrition rate were sufficient to form networks and perform a meta-analysis. However, other outcomes were not analyzed due to scarce data.

### 3.1. Study characteristics

The 107 included studies contained 8,909 participants for data analysis and 8,583 completers. A total of ten experimental diets and 223 arms was reported. The studies reported efficacy of CR, DASH, fiber, HFD, HPD, LCD, LGID, Med, Paleo, and VD, but ND and PfD were not included.

Characteristics of the studies are displayed in Table 2. We included 16 crossover and 91 parallel RCTs. Among them, seven were multi-arm, and six were multicenter. Four studies reported their outcomes in two or more publications. Only five studies focused on PreD population; considering that there was not significant difference among PreD and T2DM RCTs, we did not distinguish them in the meta-analysis. Fundings and conflicts of interest of the studies are listed in File S5.

**Table 2.**
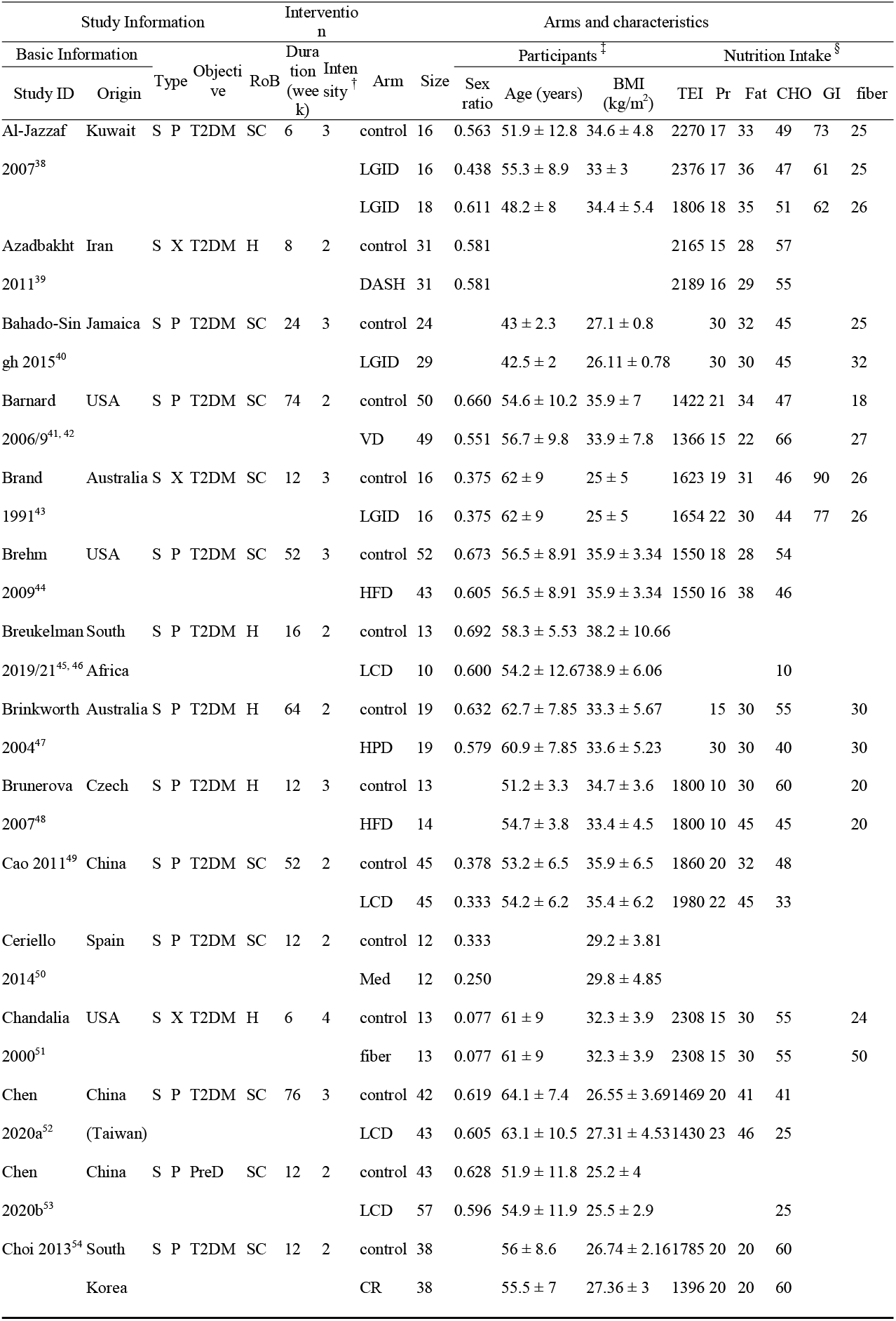
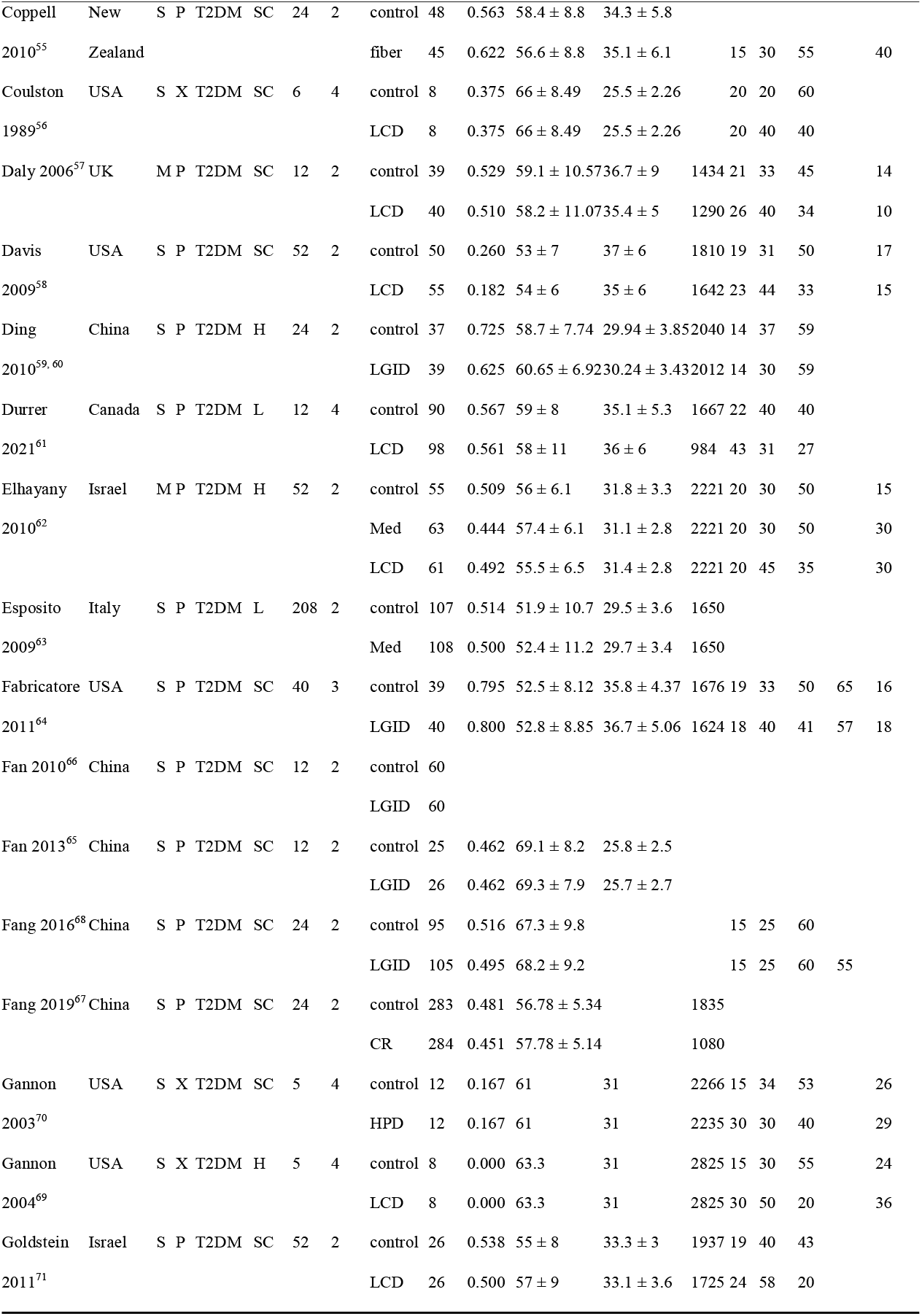
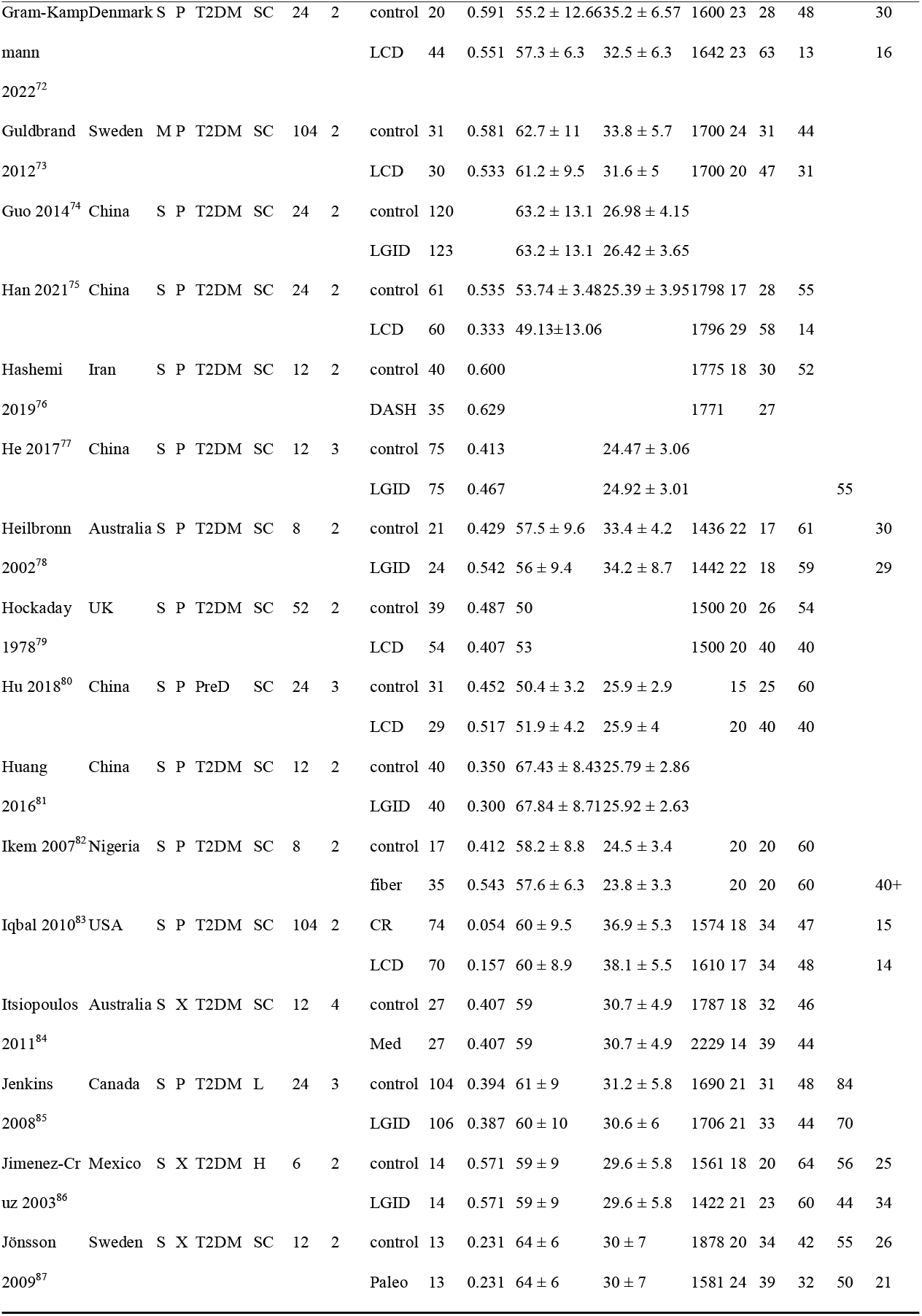
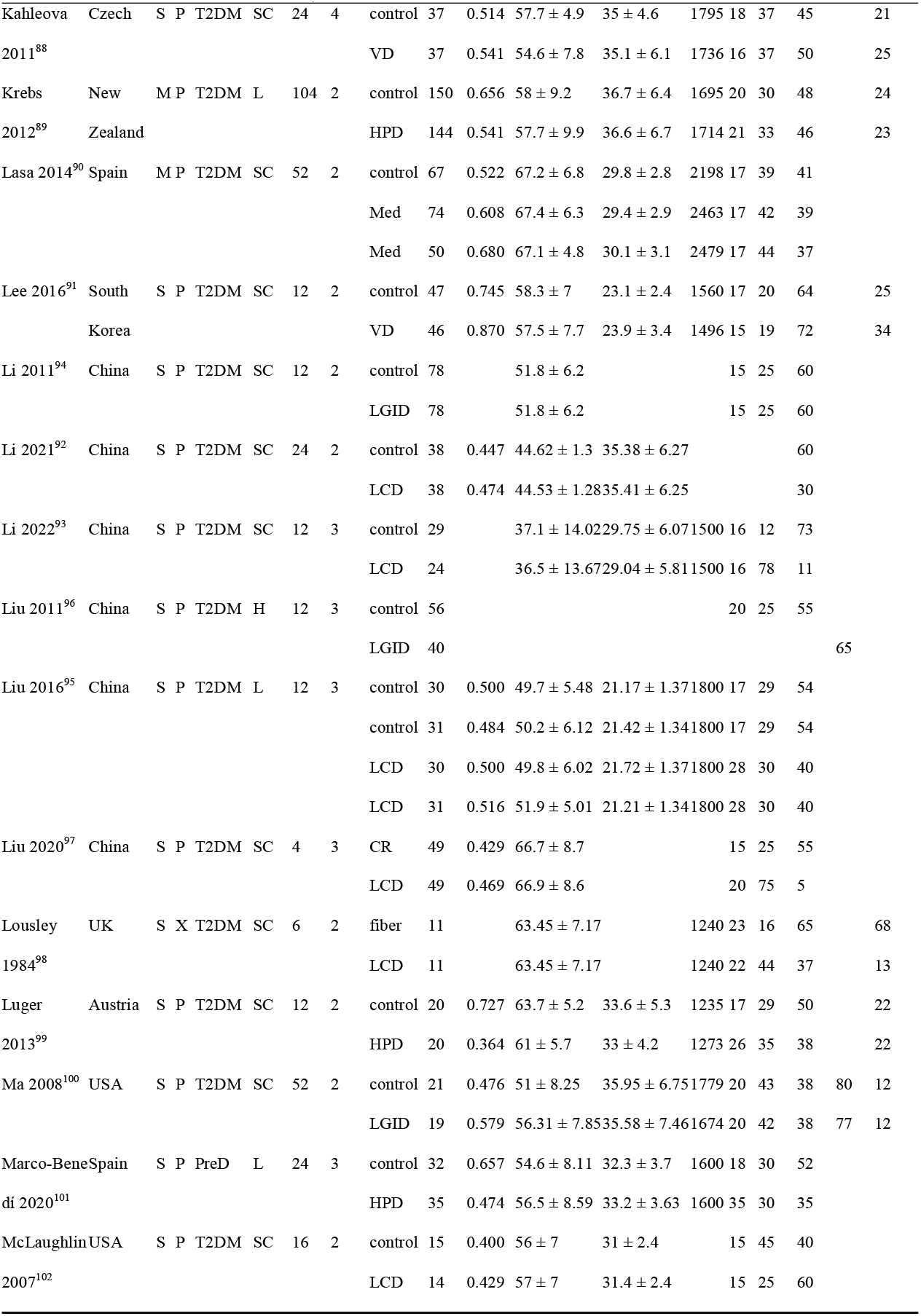
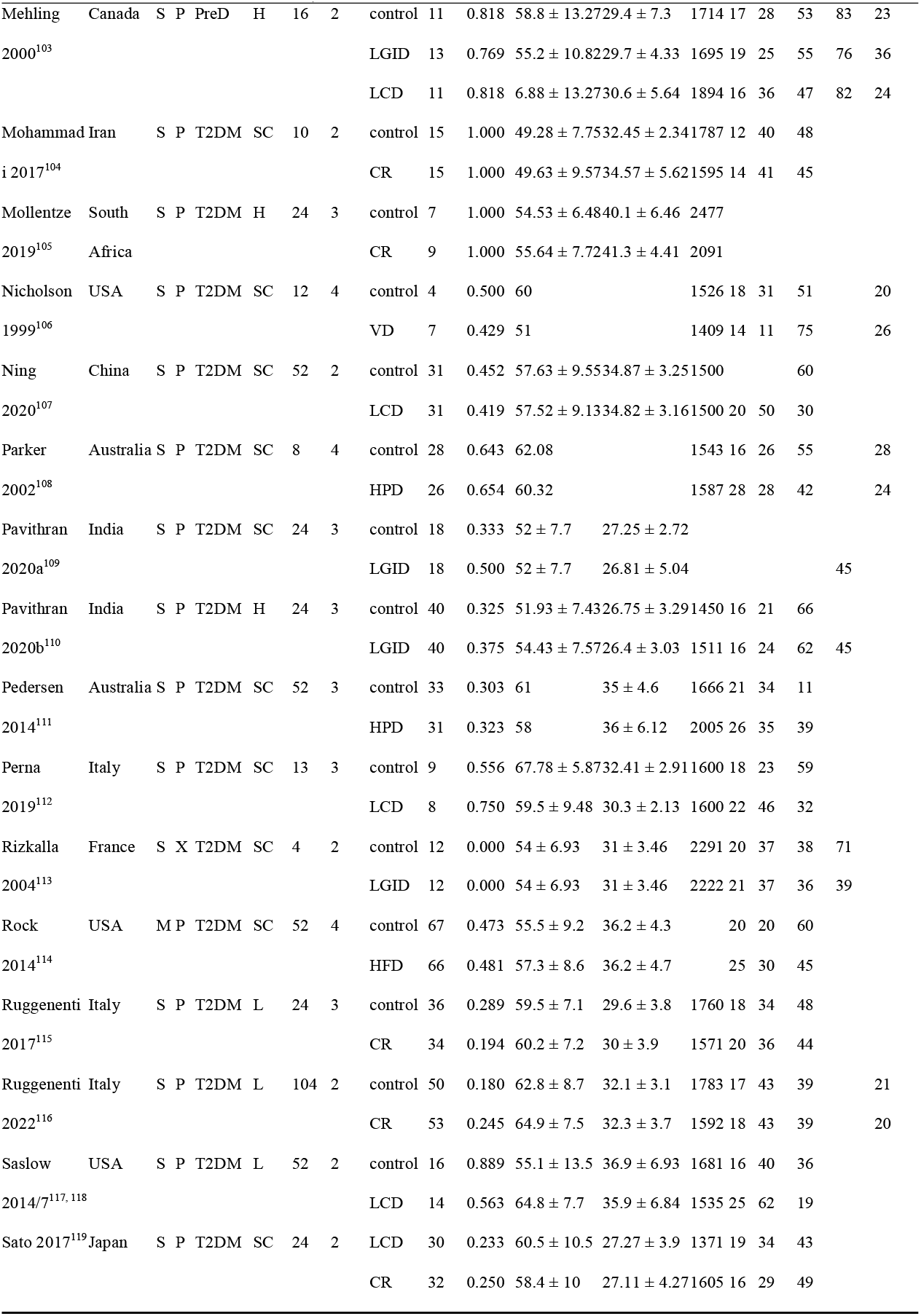
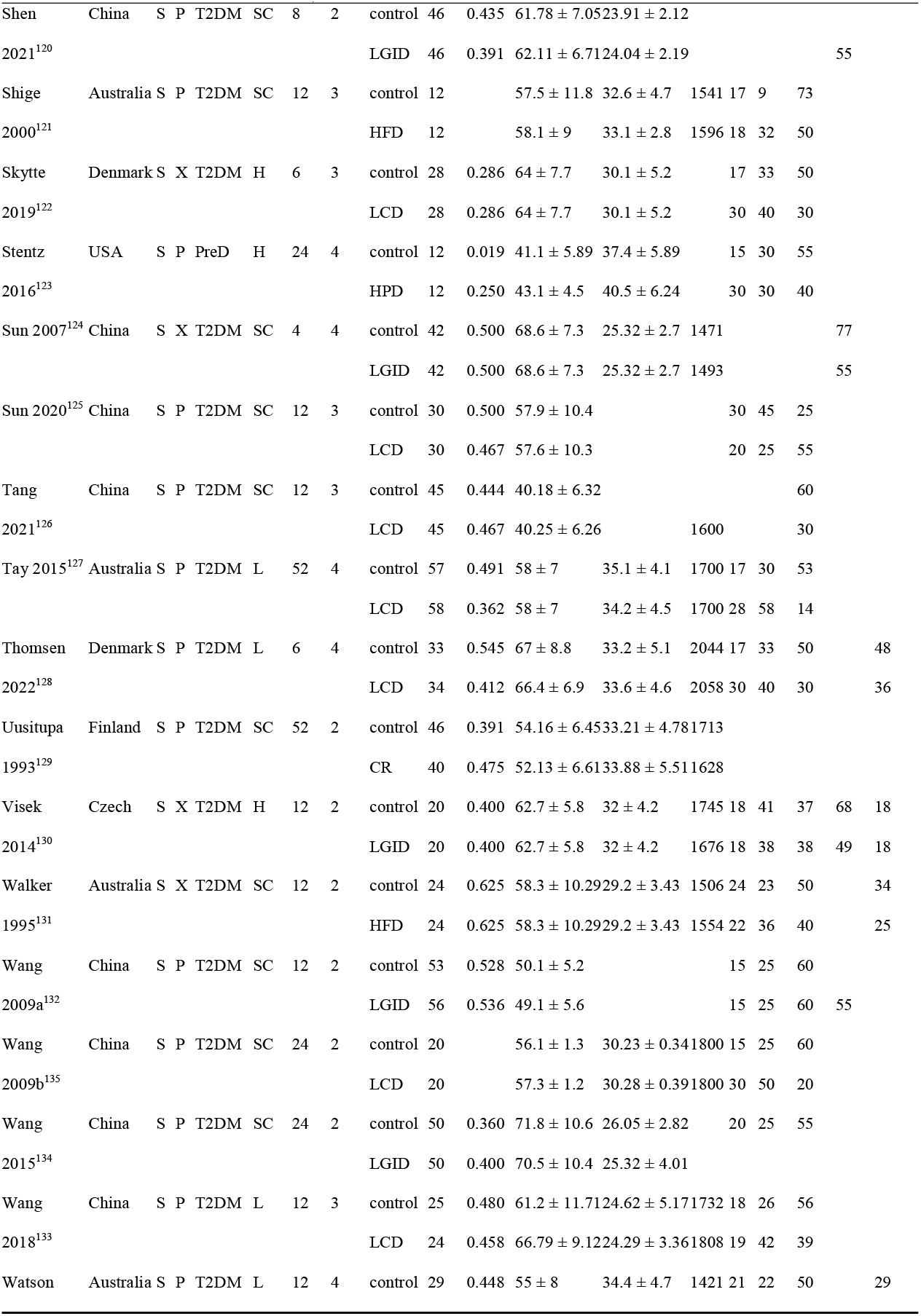
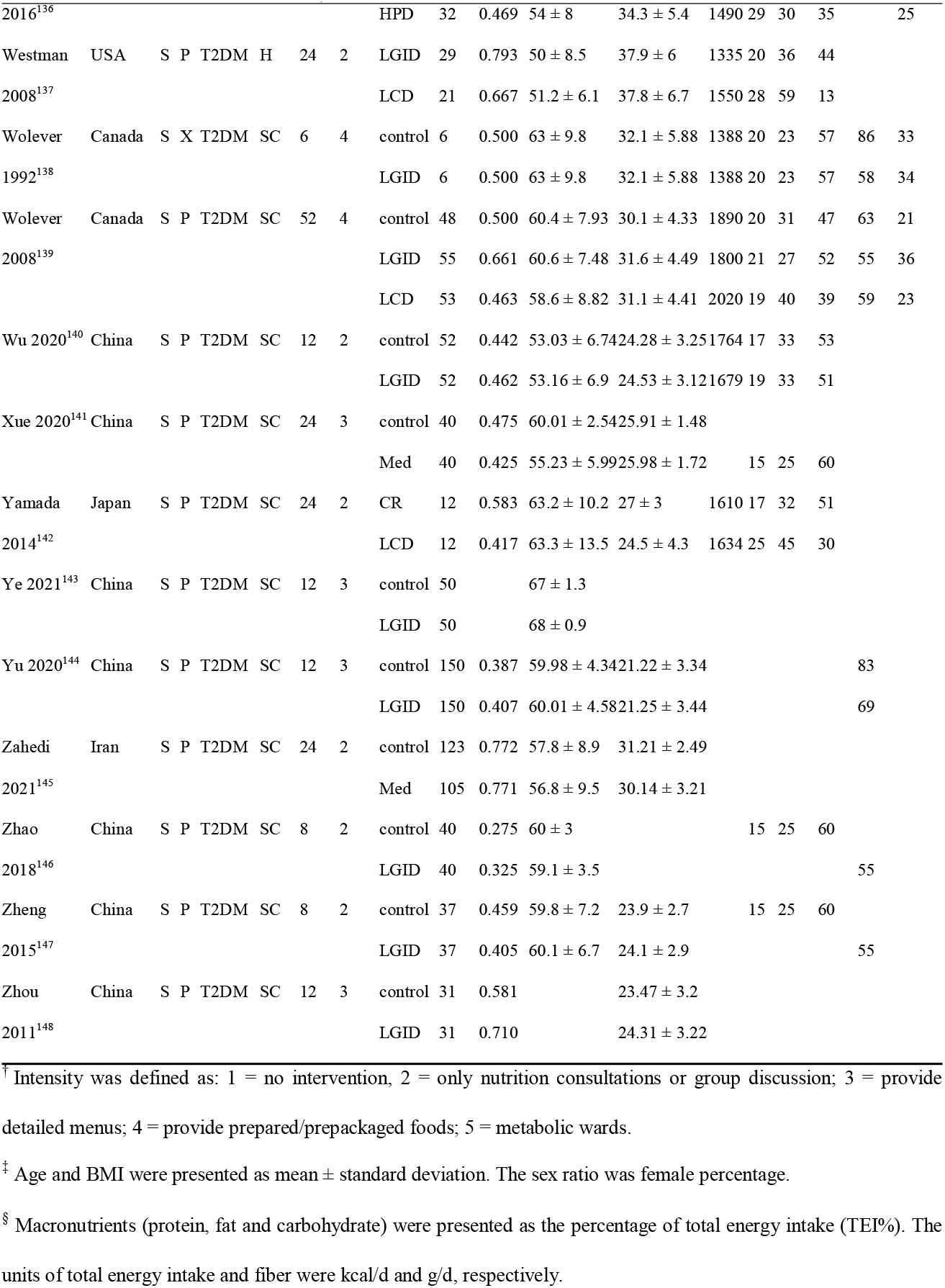
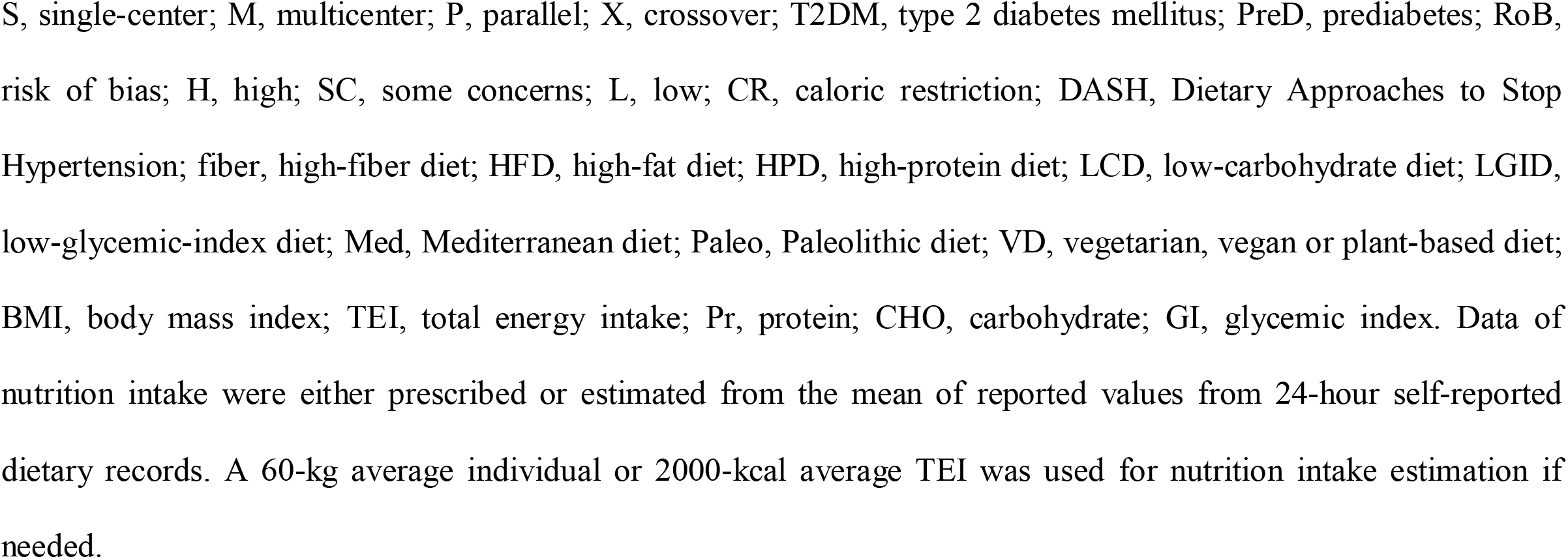
Characteristics of included studies

### 3.2. Risk of Bias Assessment

The overall risk of bias of eligible studies was acceptable, but trials of some patterns (fiber and DASH) had a relatively high risk of bias (Table 2). 15.9% of studies were at high risk of bias (Figure 2). Notably, the risk of bias of crossover RCTs was significantly higher than the parallel (*P*_0.05/2_=0.006, Mann-Whitney test), due to the period and carryover effects. Detailed risk of bias ratings of each domain are displayed in File S6.

**Figure 2.**
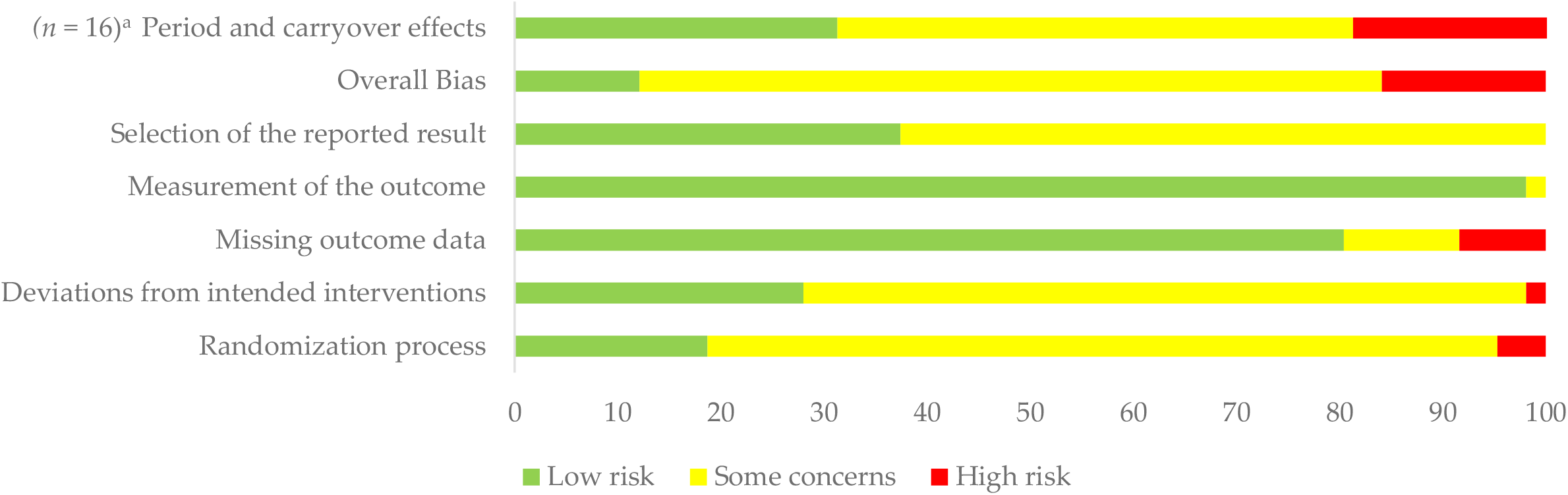
Risk of bias of included studies. ^a^ The “period and carryover effects” domain was only for crossover RCTs (*n* = 16), and other domains were for all included studies (*n* = 107).

### 3.3. Main Outcomes

The number of nodes and comparisons varied among outcomes (Figure 3 and File S7). File S8 presented all league tables and cumulative ranking curves; File S9 showed forest plots with heterogeneity and inconsistency tests of all outcomes.

**Figure 3.**
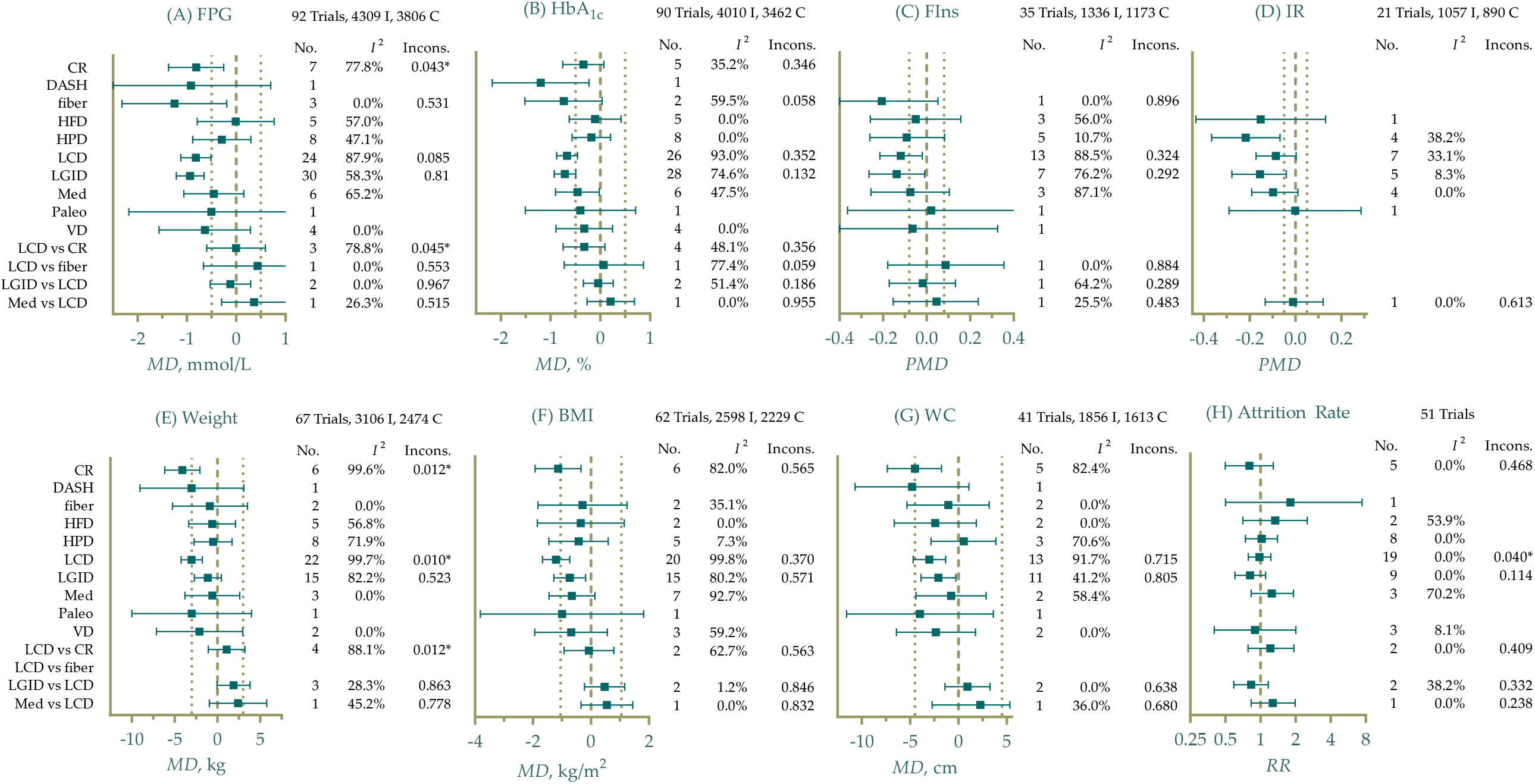
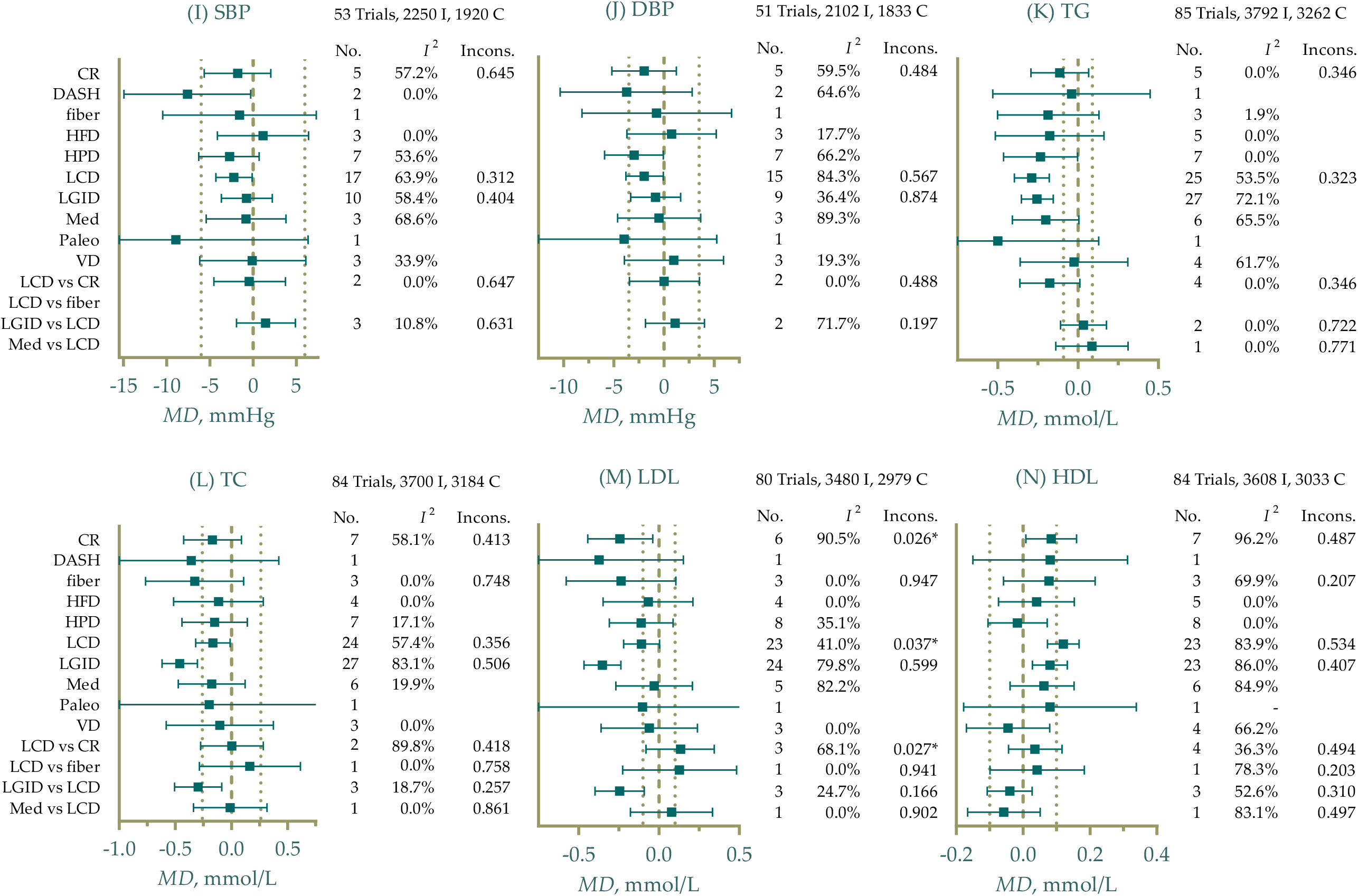
Efficacy of different eating patterns on glycemic control, anthropometrics, serum lipid profiles, and comparative attrition rate. I, intervention arm; C, control arm; No., Number of direct comparisons; Incons., *P* value of inconsistency test (node-splitting method); MD, mean difference; PMD, difference in percentage change from baseline; FPG, fasting plasma glucose; HbA_1c_, glycated hemoglobin; FIns, fasting insulin; IR, insulin resistance; BMI, body mass index; WC, waist circumference; SBP, systolic blood pressure; DBP, diastolic blood pressure; TG, triacylglycerol; TC, total cholesterol; LDL, low-density lipoprotein cholesterol; HDL, high-density lipoprotein cholesterol. Thick dashed referred to the null value, and thin dashed referred to the MCID threshold. Unless otherwise specified using “vs”, the effect sizes were experimental patterns vs control. *I*^2^ values were for network heterogeneity, including both direct and indirect comparisons.

#### 3.3.1. Glycemic Control

For glycemic control, high-fiber diet (fiber) was ranked as the best pattern for reducing FPG (MD -1.3 mmol/L, 95% CrI -2.3 to -0.22, SUCRA 82.7%) (Figure 3A). DASH (−1.2%, -2.2 to -0.23, SUCRA 90.5%) and LGID (−0.71%, -0.93 to -0.49, SUCRA 76.2%) had the highest probability of improving HbA_1c_ compared with control groups (Figure 3B). The effects on reducing FPG and HbA_1c_ were comparable.

FIns and IR were presented as PMD due to the various units reported by studies. Effects on improving insulin-related conditions were not stable and significant because of the limited sample size. High-fiber diets achieved a mean of 21% Fins reduction (95% CrI 5.2% to 46%) with a probability of 79.4% to be the best pattern (Figure 3C). IR was reported as homeostatic model assessment (HOMA)1-IR and HOMA2-IR, among which HPD showed the best beneficial effects on improving IR (−22%, -37% to -7.0%, SUCRA 86.3%) (Figure 3D).

#### 3.3.2. Anthropometrics

CR was still one of the most effective diet patterns for weight loss (−4.1 kg, -6.1 to -2.0, SUCRA 86.8%) and WC (−4.5 cm, -7.4 to -1.8, SUCRA 82.2%), and LCD was ranked as the second (−3.0 kg, -4.3 to -1.8, SUCRA 74.3%) for weight loss and the best (−1.2 kg/m^2^, -1.7 to -0.74, SUCRA 81.6%) for BMI reduction (Figure 3E-G).

As for blood pressure, DASH was found to be the best pattern for lowering SBP (−7.6 mmHg, -15 to -0.29, SUCRA 87.9%) and the second for DBP (−3.7 mmHg, -10 to 2.8, SUCRA 73.7%), while HPD was the most effective for DBP (−3.0 mmHg, -5.9 to -0.068, SUCRA 74.6%) with slight superiority to DASH (Figure 3I-J).

#### 3.3.3. Lipid Profiles

Figure 3K-N illustrated different interventions’ effects on lipid profiles comparing with control groups. LGID showed the most remarkable efficacy for lowering TC (−0.46 mmol/L, -0.62 to -0.30,

SUCRA 87.5%) and LDL (−0.35 mmol/L, -0.47 to -0.24, SUCRA 86.6%), but were not of beneficial effects on HDL. Paleo was ranked as the best pattern for improving TG (−0.50 mmol/L, -1.1 to 0.13, SUCRA 83.4%), though the outcome was not statistically significant. LCD led to an average increase of 0.12 mmol/L (95% CrI 0.073 to 0.17, SUCRA 84.0%) for HDL compared to control, thus being the best intervention with a small effect size.

#### 3.3.4. Attrition

Since a considerable number of studies did not report standardized flowcharts of follow-up, we only included trials that reported a loss in at least one arm into synthesis. An attrition rate was calculated as: the attrition number divided by the product of participant number when allocation and the duration of intervention. The meta-analysis did not find significant difference among all patterns (Figure 3H; File S8), suggesting that participants’ tolerance for each diet be similar.

### 3.4. Heterogeneity and Inconsistency Test

Generally, the included interventions were of moderate to high heterogeneity (Figure 3, File S9, and File S10), making the results less confident. LCD-control, CR-control, LGID-control, LCD-CR, and LGID-LCD pairs were of high heterogeneity in either direct or network comparison, while Med-control and HPD-control were with mild heterogeneity in lipid profiles. Significant inconsistency was observed in LCD-CR for FPG, and CR-LCD-control loop for weight and LDL using node-splitting methods. The evidence of CR, LCD and LGID showed severe incoherence and inconsistency and should be interpreted prudently.

### 3.5. Meta-regression

A random effect meta-regression model with one covariate and exchangeable coefficients was fitted for continuous outcomes. The significance of coefficients was summarized in File S11. Universally, the meta-regression denoted that weight, BMI, and macronutrient intake significantly modified the efficacy of interventions of most outcomes. On the contrary, coefficients of length, study design, medication or insulin treatment, duration of disease, and sex ratio were not significant, implying that these factors may not contribute to the effectiveness. Another notable finding that coefficients of sample size and origin (from China or not) showed significance in FPG, weight, and lipid profiles indicated potential publication or selection biases.

### 3.6. Sensitivity Analysis

Effect of Weight, BMI, and TC showed robustness, but other outcomes were not robust enough (File S12). The exclusion of several articles^51, 61, 67, 97, 98, 101, 126, 134, 135, 140, 145^ significantly changed the SUCRA and the 95% CrI of effect size, mainly in comparisons of CR, LCD, and Med vs. control, contributing to the severe heterogeneity. When testing for different models, i.e., fixed effect or unrelated study effect models, Med, HPD, and VD showed narrower 95% CrIs and became statistically significant for more outcome variables (see File S12). The analysis did not observe the sensitivity of relative effect and between-study heterogeneity priors, and correlation coefficients.

### 3.7. Publication Bias

Potential publication bias of HbA_1c_, weight and BMI existed (Egger’s test *P* = 0.002; < 0.001; and < 0.001, respectively). *P* values for all outcomes and comparison-adjusted funnel plots were listed in File S13.

### 3.8. Quality of Evidence

All MCIDs and thresholds were identified (see File S14, Figure 3, and Table 3). Of all 123 pieces of evidence comparing interventions and control groups, 49 were of moderate quality, and there was no high-quality evidence (Table 3). At the clinical level, all patterns were not significantly worse than control diets for each outcome, but most did not show moderate to large beneficial effects. All the quality of evidence should be downgraded when applying to PreD due to the indirectness, because PreD-related trials were limited.

**Table 3.**
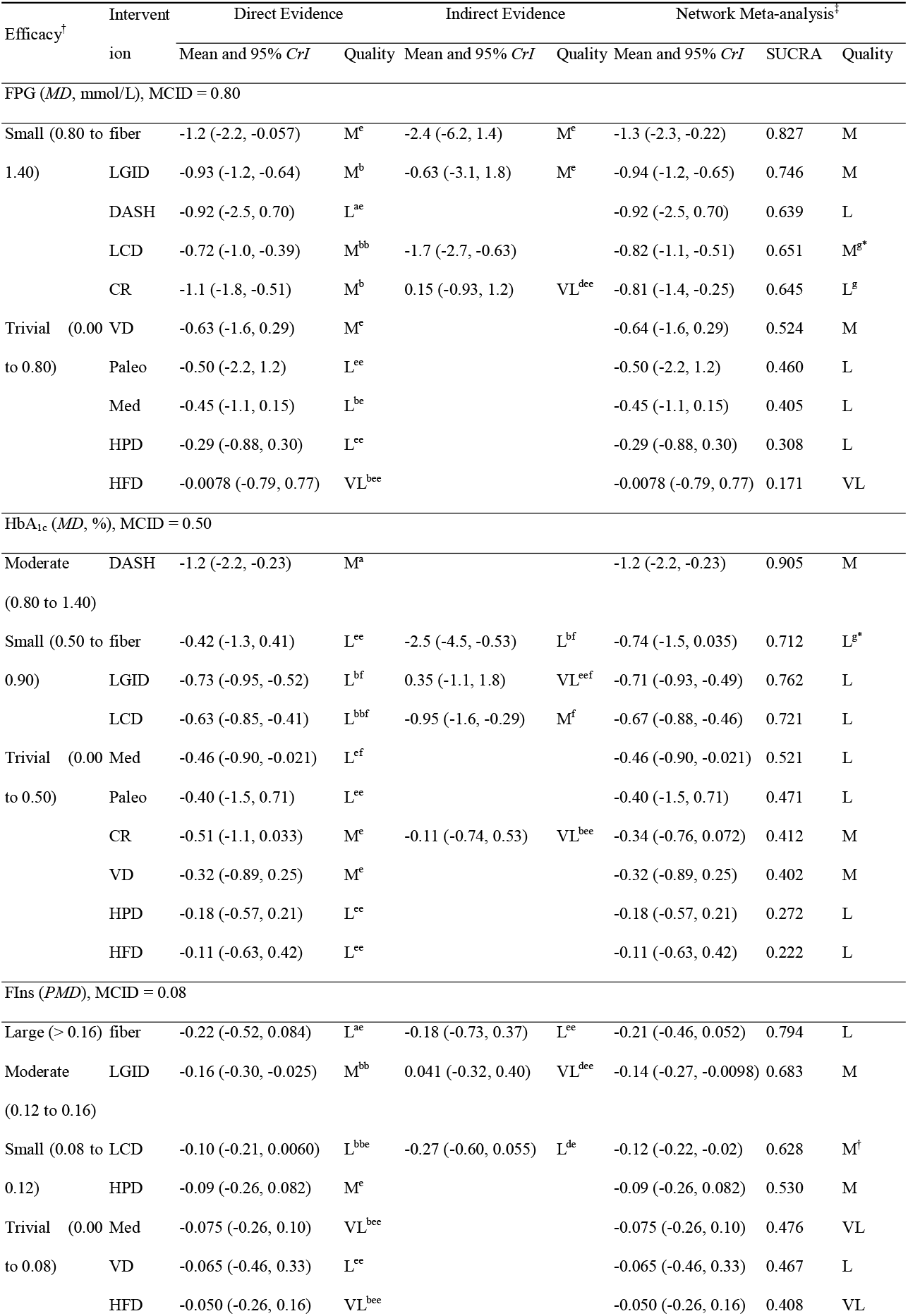
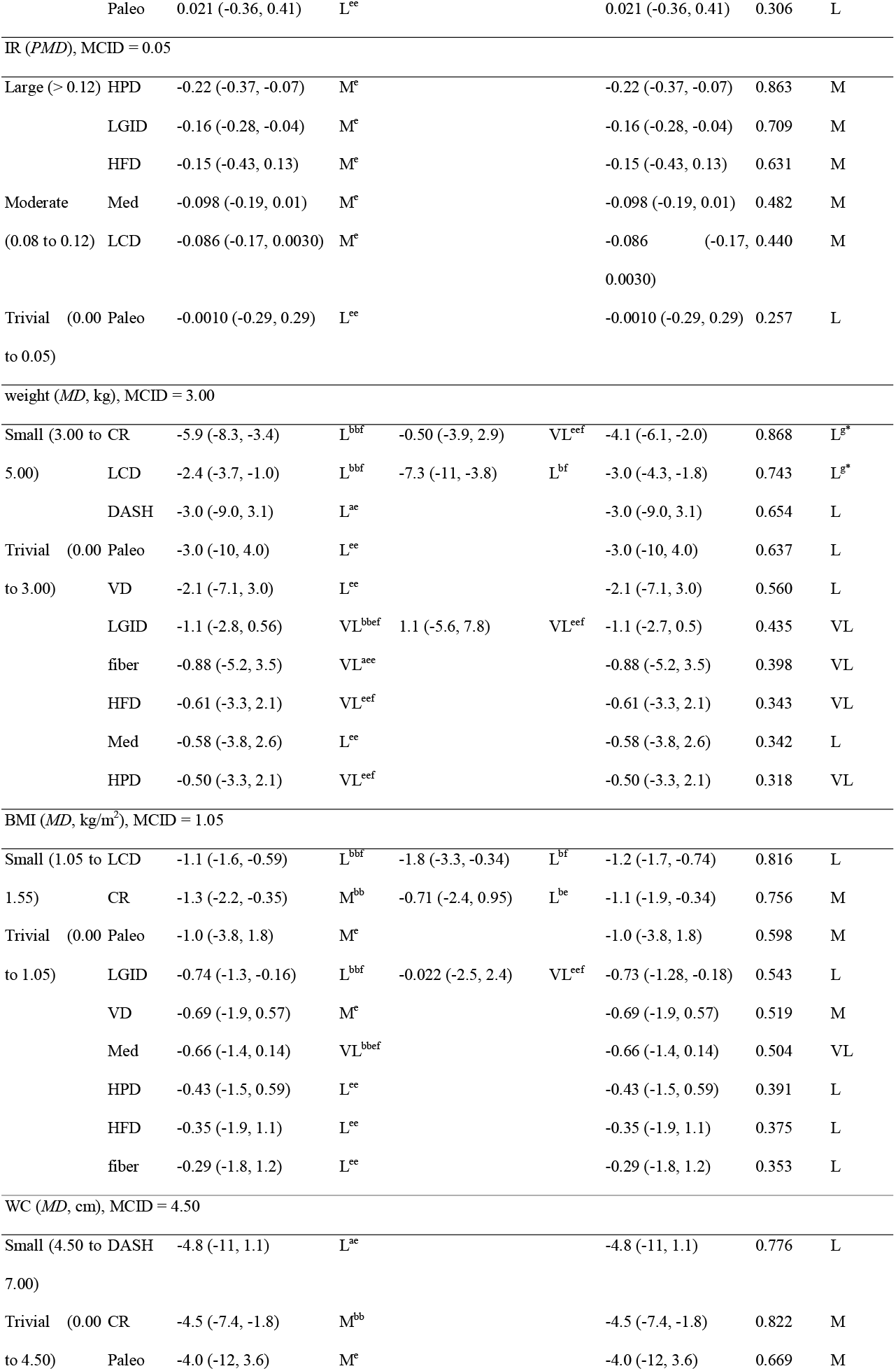
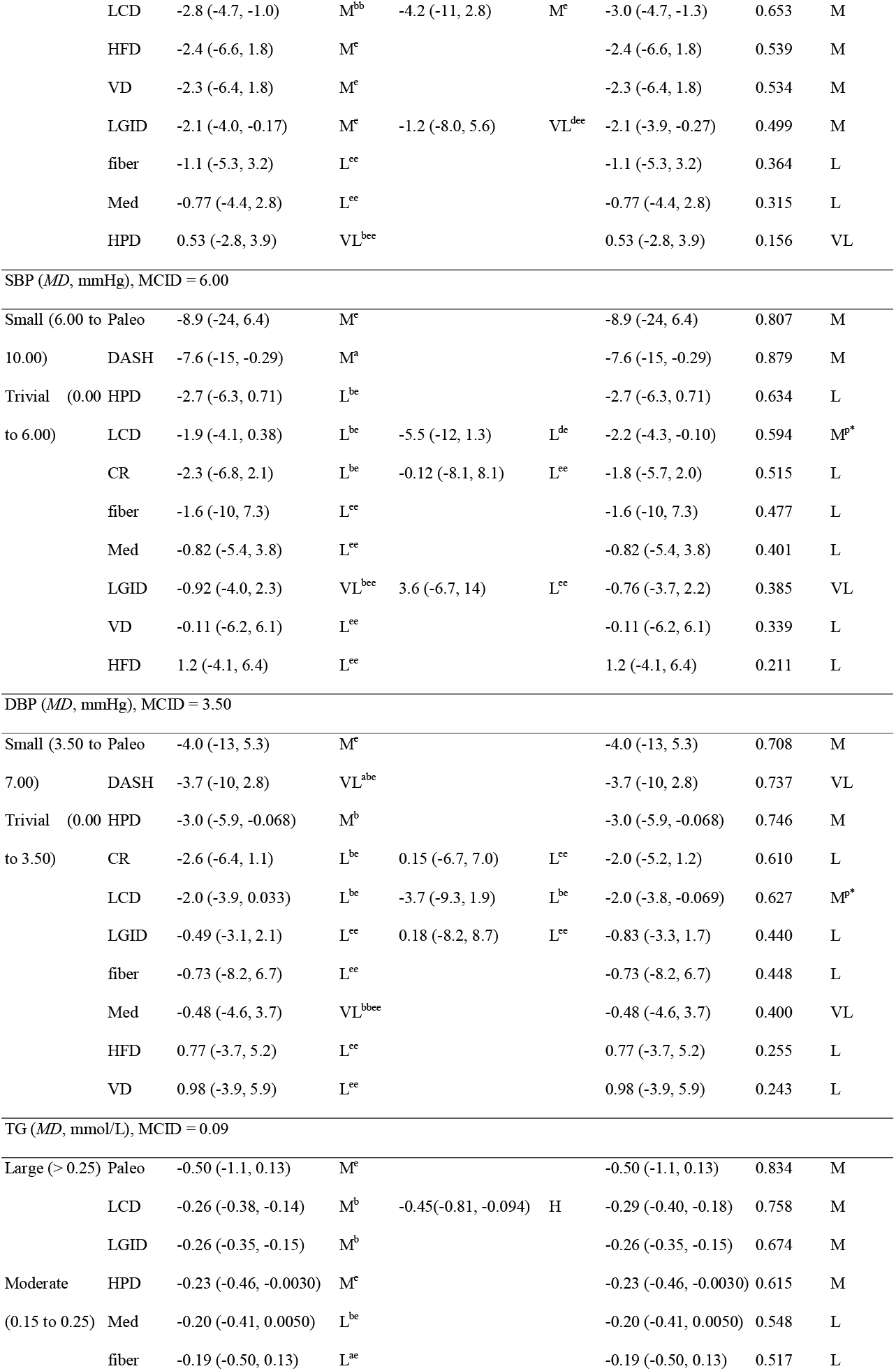
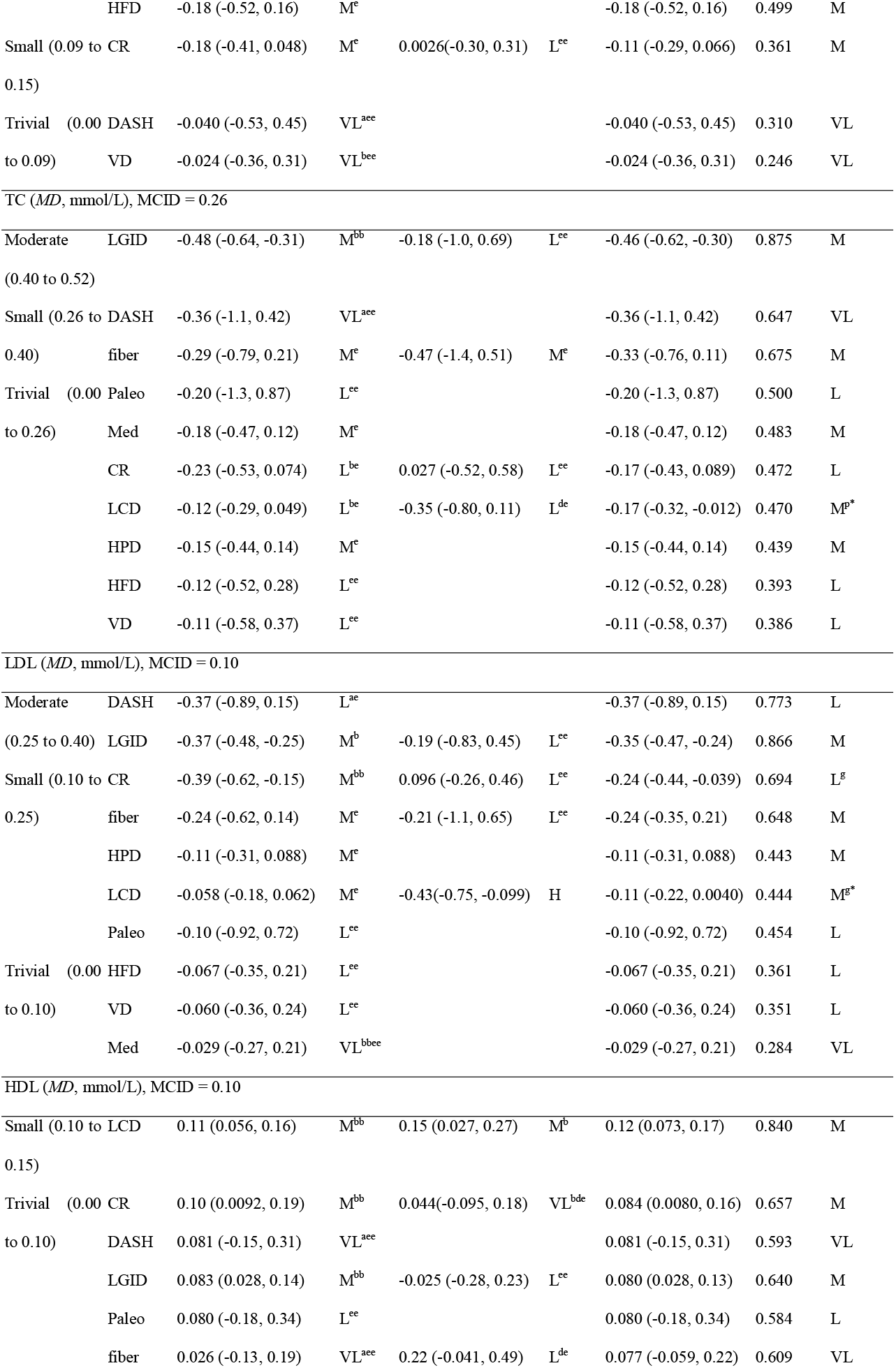
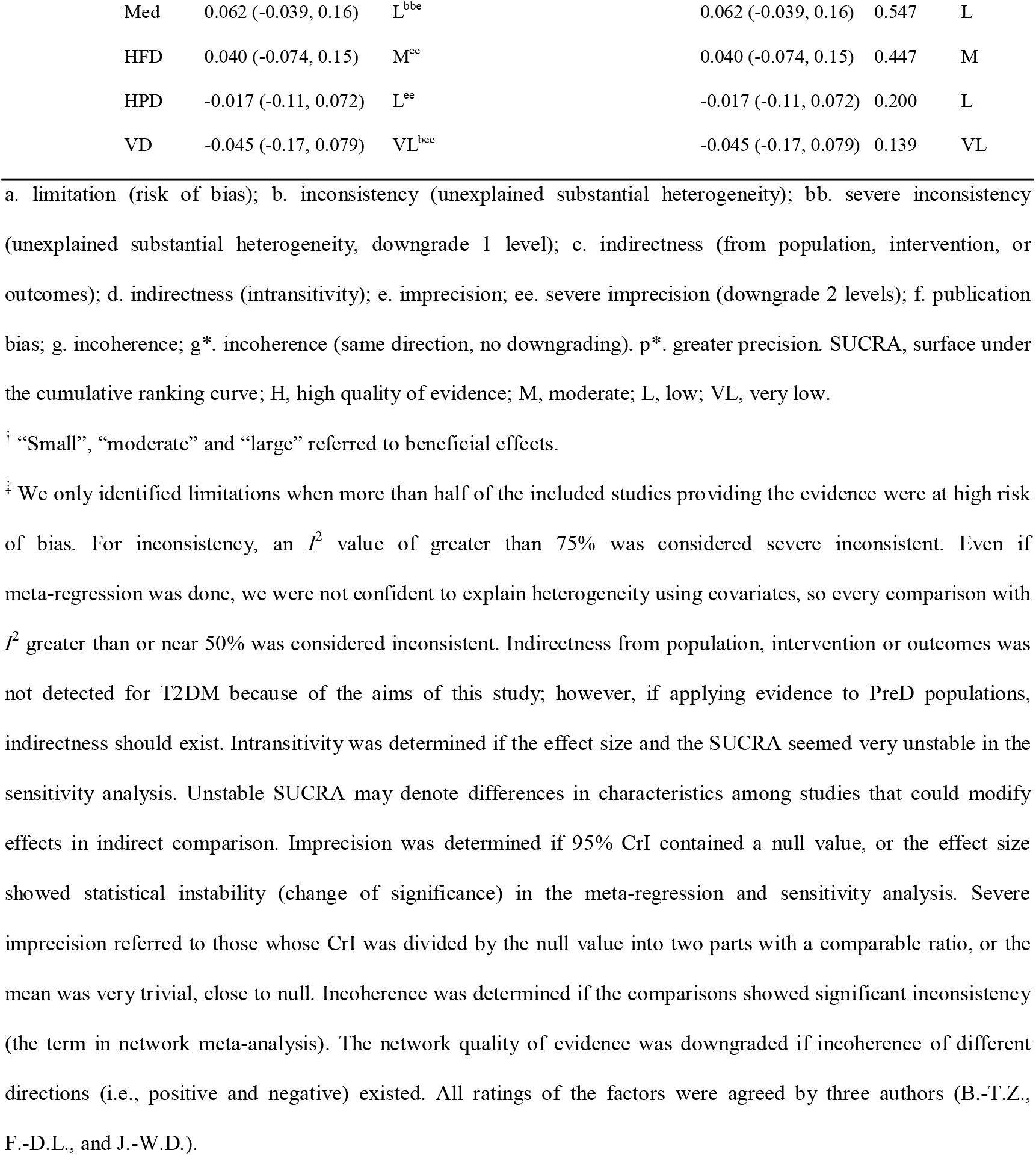
Summary of findings

## 4. Discussion

This review evaluated the comparative efficacy of ten experimental diets, and the results can provide guidance for diet selection of one specific patients. To manage patients with comorbidities and different levels of glycemic control, we concluded a dietary suggestion table derived from the evidence from the meta-analysis (Table 4). However, this table should be applied prudently because the evidence was not solid enough.

**Table 4.**
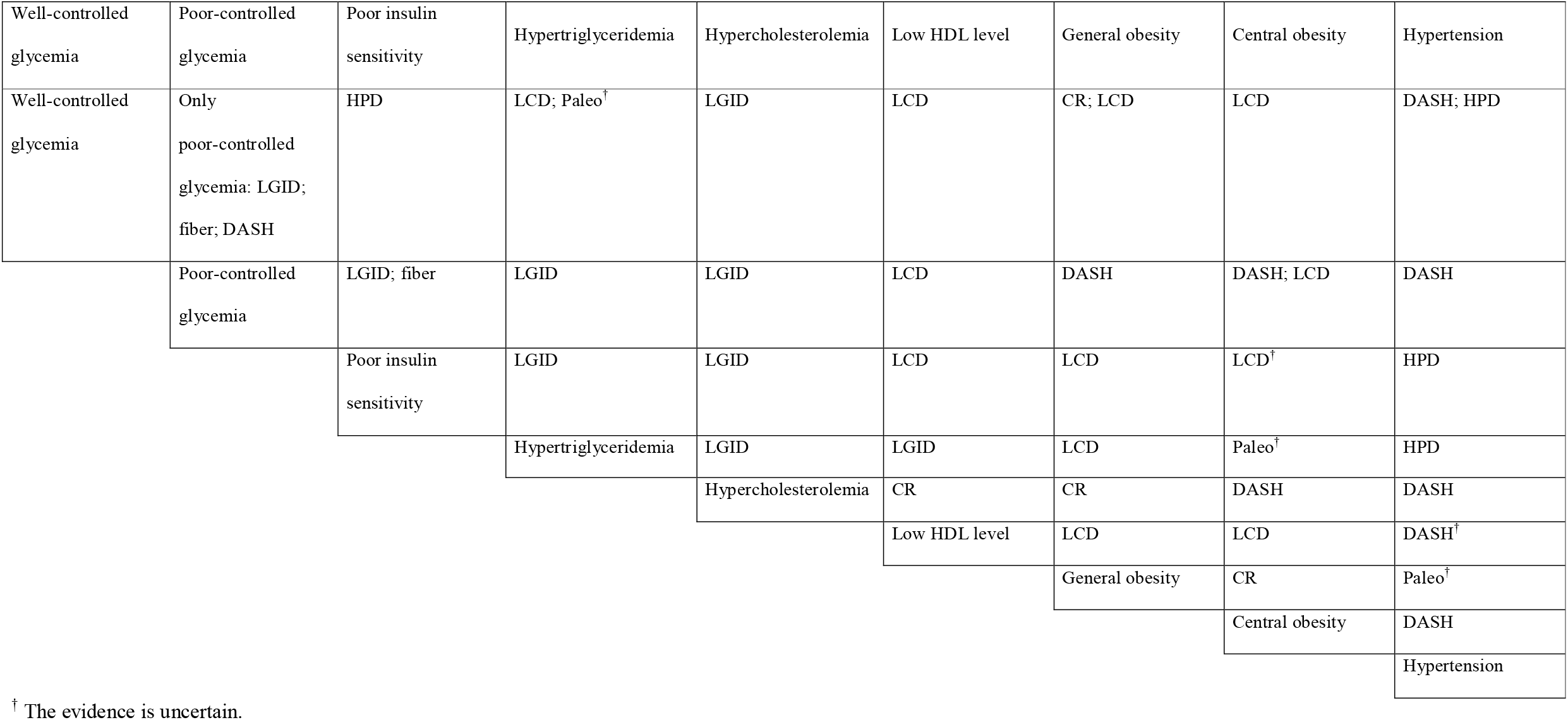
Dietary suggestions for patients with different profiles

| Well-controlled glycemia | Poor-controlled glycemia | Poor insulin sensitivity | Hypertriglyceridemia | Hypercholesterolemia | Low HDL level | General obesity | Central obesity | Hypertension |
| --- | --- | --- | --- | --- | --- | --- | --- | --- |
| Well-controlled glycemia | Only poor-controlled glycemia: LGID; fiber; DASH | HPD | LCD; Paleo<sup>†</sup> | LGID | LCD | CR; LCD | LCD | DASH; HPD |
|  | Poor-controlled glycemia | LGID; fiber | LGID | LGID | LCD | DASH | DASH; LCD | DASH |
|  |  | Poor insulin sensitivity | LGID | LGID | LCD | LCD | LCD<sup>†</sup> | HPD |
|  |  |  | Hypertriglyceridemia | LGID | LGID | LCD | Paleo<sup>†</sup> | HPD |
|  |  |  |  | Hypercholesterolemia | CR | CR | DASH | DASH |
|  |  |  |  |  | Low HDL level | LCD | LCD | DASH<sup>†</sup> |
|  |  |  |  |  |  | General obesity | CR | Paleo<sup>†</sup> |
|  |  |  |  |  |  |  | Central obesity | DASH |
|  |  |  |  |  |  |  |  | Hypertension |

<sup>†</sup> The evidence is uncertain.

### 4.1. Quantity of Macronutrients

A previous evidence basis has corroborated the efficacy of CR in weight loss, BMI and WC in patients with metabolic diseases or healthy individuals^149, 150^. However, CR did not lead to greater improvement of glycemic control, blood pressure, TG, and TC compared to standard diets. Trivial effects on these outcomes may result from weight loss but not the caloric restriction^151, 152^. The median TEI of the included CR arms was 1594 kcal/d, with a 150-to-400-kcal negative difference compared to standard diets, significantly slighter than the prescribed (−500 kcal/d). However, the deviance did not lead to the failure of trials. The phenomena were also observed in LCD and LGID.

Carbohydrate restriction acted well in weight, HbA_1c_, TG, and HDL, where improving HDL was the unique advantage of LCD. Nevertheless, other types of serum lipids, i.e., TC and LDL were not improved. The 75^th^ percentile of carbohydrate intake of the included LCD arms was 40%, indicating that nearly a quarter of included trials did not meet the low-carbohydrate criteria as prescribed. Nevertheless, the effect size was similar to previous systematic reviews^13^, and the strict following of the instruction as well as a more intensive intervention did not enhance the effects but may even lead to a decrease (File S11).

Increased protein intake without carbohydrate restriction (HPD) effectively improved IR, blood pressure and TG. Compared to other review^153^, the effectiveness of HPD on FPG, HbA_1c_ and other lipids was not observed, mainly due to the different inclusion criteria: only HPD with protein intake of more than 30% TEI and without carbohydrate restriction was included. This implied the different efficacy of protein and carbohydrate.

As for HFD, no beneficial effect was detected, and fat intake negatively modified the lipid improvement. Despite the numerical impact on specific lipids, it remained to be evaluated whether specific types of fat improved or negatively affected the overall lipoprotein profile^154^. Unfortunately, the included trials did not provide sufficient data to draw a thorough interpretation.

### 4.2. Quality of Carbohydrates

LGID and high-fiber diets emphasized more on the quality of carbohydrates. Effects of LGID and high-fiber diets were similar: both showed more excellent effects on FPG, HbA_1c_, FIns, TC, and LDL than other patterns, but did not significantly improve weight-related outcomes, consistent with other studies^155, 156^. Dietary GI and fiber of specific single food were not well-associated^157^. However, the emphasis on lowering GI may encourage participants to increase fiber intake, because the usually recommended food groups can be both low in GI and high in fiber, e.g., whole grains and nuts.

A recent high-quality meta-analysis has also denoted that dietary fiber and low-GI food were associated with a lower risk of T2DM incidence, where fiber may be a stronger protector^158^. Rather than a severe long-term restriction of carbohydrate intake which leads to higher all-cause mortality^159^, LGID and increased fiber intake can be better and sustainable approaches for T2DM patients without obesity/overweight, especially with the circumstance that most people lacked fiber intake^160^.

### 4.3. Mediterranean Diets

Even if previous cohort studies and RCTs have demonstrated the efficacy of Med in T2DM management^161^, our study failed to detect a significant improvement driven by Med. Except for HbA_1c_, IR and TG, all other outcomes were of great imprecision and trivial effects. The effect size was also more trivial than other meta-analyses^14, 162^. Small sample size compared to other interventions could be the reason when using random effects models; different calculation of effect size, i.e., MD of change from baseline or of endpoint may explain the numerical differences.

Moreover, heterogeneity was detected for almost all outcomes of Med-control comparisons, where the variance and bias of the definition of Med in different trials^163^ can be a significant reason. Though several scales have been developed to measure the adherence to Med (e.g., MedDiet Score)^164^, few trials employed it, making this problem difficult to address.

### 4.4. Vegan, Vegetarian, or Plant-based Diets

VD did not show any significant beneficial effects in our study. The mean differences of VD were similar to the previous studies^36^, thus not affecting the conclusion but lowering the quality of evidence. While using fixed effect models, the effectiveness of VD on BMI, WC, and HbA_1c_ was detected, but moderate heterogeneity made it unreasonable to employ fixed effect models.

Notably, the carbohydrate intake in VD trials was relatively high (mean 65.8%TEI). The sensitivity analysis also showed a slight improvement of SUCRA in TG after omitting Lee 2016^91^, which contained about 72%TEI of carbohydrate in VD arms. Researchers should consider a lower carbohydrate intake when conducting VD, and the effects would promise to be more significant.

### 4.5. Newly-developed diets

Evidence of the efficacy of DASH and Paleo was limited and of low quality due to the sample size, and further investigation is needed. As one of the recommended healthy patterns by Dietary Guidelines for Americans (DGA 2020-2025)^165^, many studies have addressed DASH’s benefit in blood pressure and glycemic control^166, 167^. However, related RCTs specially for T2DM/PreD patients were rare. Included studies also outlined the beneficial effects of DASH on blood pressure, TC, LDL, and HbA_1c_, and DASH was the most effective intervention for HbA_1c_ with a high probability (90.5%). As for Paleolithic diets, Tommy Jönsson and his colleagues quantified the improvement of leptin and introduced a scale (Paleolithic Diet Fraction) to measure the compliance, based on their trial^87, 168^, providing a basis for further studying.

### 4.6. Limitations

This study had several limitations. First, the heterogeneity and sensitivity lowered the quality of evidence. Second, the sample size of VD, DASH, and Paleo was limited, leading to the imprecision. Third, only five PreD trials were included, raising the indirectness of the evidence for PreD population. Moreover, there was not an adequate method to compare the longitudinal dataset of different patterns, though the data of different timepoints have been extracted.

In conclusion, Energy, carbohydrate, and dietary glycemic index (GI) restriction, as well as dietary fiber intake, were the most effective approaches with solid and abundant evidence bases. Simultaneously, DASH, Paleolithic diets, and HPD were of satisfactory efficacy in limited outcomes and worth investigation. Mediterranean diets, VD and HFD did not act well in most outcomes, mainly due to the imprecision. Heterogeneity and sensitivity should be concerned when interpreting results.

This work may eliminate some barriers on how to choose the best diet on an individualized basis. Clinicians and dietitians can choose the most important outcome that in an urgent need to control for a patient to match the most appropriate dietary pattern, according to the summary of finding table and the dietary suggestion table of this review.

## Data Availability

All data produced in the present work are contained in the manuscript and the supplementary materials.

## Acknowledgments

We acknowledge Professor Lawrence J. Cheskin from George Mason University for his kindly replying our email about the data availability of his registered trial. This research did not receive any funding.

## Disclosure of Ethical Statements

Approval of the research protocol: N/A. Informed consent: N/A.

Registry and the registration no. of the study/trial: This network meta-analysis was registered at https://www.crd.york.ac.uk/PROSPERO as CRD42021278268.

Animal studies: N/A.

## Supporting Information Legends

**File S1**. Full search strategy

**File S2**. Data extraction template

**File S3**. Correlation coefficients for estimation

**File S4**. Reason for exclusion

**File S5**. Fundings and conflicts of interest of included studies

**File S6**. Risk of bias assessment

**File S7**. Network plots

**File S8**. League tables and cumulative ranking curves

**File S9**. Forest plots

**File S10**. Heterogeneity and inconsistency test

**File S11**. Meta-regression

**File S12**. Sensitivity analysis

**File S13**. Publication bias

**File S14**. Minimal clinically important difference and thresholds for effects.

