## Supplementary material for "Comparative efficacy of different eating patterns in the management of type 2 diabetes and prediabetes: An arm-based Bayesian network meta-analysis": File S

### Supplementary Materials

for the article

***Comparative Efficacy of Different Eating Patterns***
***in the Management of Type 2 Diabetes and Prediabetes:***
***A Systematic Review and Network Meta-Analysis of Randomized Controlled Trials***

#### File S1: Full search strategy

##### S1.1 PubMed

Search time: 02:18, October 13, 2021 (GMT+8); 1 333 results

| No. | Search |
|---|---|
| #1 | "Diabetes Mellitus, Type 2"[mh] OR ("Diabetes"[tiab] AND ("Type 2"[tiab] OR "Type II"[tiab] OR "Type II"[tiab] OR "Maturity Onset"[tiab] OR "Adult Onset"[tiab] OR "Slow Onset"[tiab] OR "Noninsulin"[tiab] OR "Non-insulin"[tiab] OR "Ketosis Resistant"[tiab])) OR "NIDDM"[tiab] OR "T2DM"[tiab] |
| #2 | "Prediabetic State"[mh] OR "Prediabet* "[tiab] |
| #3 | "Diet, Carbohydrate-Restricted"[mh] OR "Diet, Ketogenic"[mh] OR ("Diet*"[tiab] AND ("Carbohydrate Restricted"[tiab] OR "Carbohydrate-Restricted"[tiab] OR "low-carbohydrate"[tiab] OR "low carbohydrate"[tiab] OR "ketogenic"[tiab])) |
| #4 | "Diet, Paleolithic"[mh] OR ("Diet*"[tiab] AND ("Paleo*"[tiab] OR "Caveman"[tiab] OR "Hunter Gatherer"[tiab] OR "Hunter-Gatherer"[tiab] OR "Stone Age"[tiab])) |
| #5 | "Diet, High-Protein"[mh] OR "Diet, High-Protein Low-Carbohydrate"[mh] OR ("Diet*"[tiab] AND ("High-Protein"[tiab] OR "High Protein"[tiab] OR "South Beach"[tiab] OR "Atkins"[tiab])) |
| #6 | "Diet, Fat-Restricted"[mh] OR ("Diet*"[tiab] AND ("Fat-Restricted"[tiab] OR "Fat Restricted"[tiab] OR "Low-Fat"[tiab] OR "Low Fat"[tiab] OR "Fat-Free"[tiab] OR "Fat Free"[tiab])) |
| #7 | "Caloric Restriction"[mh] OR ("Calori*"[tiab] AND ("Low"[tiab] OR "Restricted"[tiab]) OR "low-calorie"[tiab] OR "low calorie"[tiab]) |
| #8 | "Diet, Mediterranean"[mh] OR ("Diet*"[tiab] AND "Mediterranean"[tiab]) |
| #9 | "Diet, Vegetarian"[mh] OR (("Diet*"[tiab] AND ("Vegetarian*"[tiab] OR "Lacto-Vegetarian"[tiab])) OR (("Diet*"[tiab] OR "Nutrition"[tiab]) AND ("Plant-Based"[tiab] OR "Plant Based"[tiab]))) OR "Vegetarianism"[tiab] |
| #10 | "Diet, Vegan"[mh] OR "Vegan*"[tiab] |
| #11 | "Dietary Approaches To Stop Hypertension"[mh] OR "DASH"[tiab] OR "Dietary Approaches To Stop Hypertension"[tiab] |
| #12 | "Nordic*"[tiab] AND "diet*"[tiab] |
| #13 | "low glycemic index diet*"[tiab] OR "low-glycemic index diet*"[tiab] OR "low glycaemic index diet*"[tiab] OR "low-glycaemic index diet*"[tiab] |
| #14 | ("high fiber"[tiab] OR "high-fiber"[tiab] OR "high fibre"[tiab] OR "high-fibre"[tiab]) AND "diet*"[tiab] |
| #15 | "Portfolio diet"[tiab] |
| #16 | "randomized controlled trial"[pt] OR "controlled clinical trial"[pt] OR "random*" [tiab] OR "placebo"[tiab] OR "trial"[ti] OR ("clinical"[tiab] AND "trial"[tiab]) OR "clinical trials as topic"[mesh] OR "clinical trial"[pt] OR "random allocation"[mesh] |
| #17 | "gestation*"[ti] OR "type 1"[ti] |
| #18 | #1 OR #2 |
| #19 | #3 OR #4 OR #5 OR #6 OR #7 OR #8 OR #9 OR #10 OR #11 OR #12 OR #13 OR #14 OR |

| No. | Search |
|---|---|
| | #15 |
| #20 | #16 AND #18 AND #19 NOT #17 |

**S1.2 CENTRAL**

Search time: 02:00, October 13, 2021 (GMT+8); 929 results

| ID | Search | Hits |
|---|---|---|
| #1 | MeSH descriptor: [Diabetes Mellitus, Type 2] explode all trees | 18904 |
| #2 | "Type 2 Diabetes" | 36222 |
| #3 | #1 OR #2 | 40884 |
| #4 | MeSH descriptor: [Prediabetic State] explode all trees | 1128 |
| #5 | Prediabet* | 3016 |
| #6 | #4 OR #5 | 3016 |
| #7 | MeSH descriptor: [Diet, Carbohydrate-Restricted] explode all trees | 502 |
| #8 | MeSH descriptor: [Diet, Ketogenic] explode all trees | 89 |
| #9 | MeSH descriptor: [Diet, High-Protein] explode all trees | 101 |
| #10 | MeSH descriptor: [Diet, Paleolithic] explode all trees | 18 |
| #11 | MeSH descriptor: [Diet, Fat-Restricted] explode all trees | 996 |
| #12 | MeSH descriptor: [Caloric Restriction] explode all trees | 878 |
| #13 | MeSH descriptor: [Diet, High-Protein Low-Carbohydrate] explode all trees | 15 |
| #14 | MeSH descriptor: [Diet, Mediterranean] explode all trees | 557 |
| #15 | MeSH descriptor: [Diet, Vegetarian] explode all trees | 232 |
| #16 | MeSH descriptor: [Diet, Vegan] explode all trees | 20 |
| #17 | MeSH descriptor: [Dietary Approaches To Stop Hypertension] explode all trees | 41 |
| #18 | "Nordic Diet" | 101 |
| #19 | "low glycemic index diet" | 295 |
| #20 | "high fiber diet" | 407 |
| #21 | "low-carbohydrate diet" | 723 |
| #22 | "atkins diet" | 103 |
| #23 | "ketogenic diet" | 480 |
| #24 | "high-protein diet" | 487 |
| #25 | "low-fat diet" | 1656 |
| #26 | "portfolio diet" | 21 |
| #27 | "paleo diet" | 11 |
| #28 | "stone age diet" | 5 |
| #29 | "Hunter Gatherer diet" | 2 |
| #30 | "fat-free diet" | 4 |
| #31 | "low-calorie diet" | 1292 |
| #32 | "plant-based diet" | 104 |
| #33 | "vegan diet" | 159 |
| #34 | "DASH diet" | 432 |
| #35 | "vegetarian diet" | 298 |

| ID | Search | Hits |
|---|---|---|
| #36 | MeSH descriptor: [Diabetes, Gestational] explode all trees | 1053 |
| #37 | MeSH descriptor: [Diabetes Mellitus, Type 1] explode all trees | 5792 |
| #38 | #3 OR #6 | 42625 |
| #39 | #7 OR #8 OR #9 OR #10 OR #11 OR #12 OR #13 OR #14 OR #15 OR #16 OR #17 OR #18 OR #19 OR #20 OR #21 OR #22 OR #23 OR #24 OR #25 OR #26 OR #27 OR #28 OR #29 OR #30 OR #31 OR #32 OR #33 OR #34 OR #35 | 7598 |
| #40 | #36 OR #37 | 6816 |
| #41 | #38 AND #39 NOT #40 | 982 (for trials 929) |

**S1.3 Embase**

Search time: 03:05, October 13, 2021 (GMT+8); 1 676 results

| No. | Search |
|---|---|
| #1 | ‘non insulin dependent diabetes mellitus’/exp OR (‘Diabetes’:ti,ab AND (‘Type 2’:ti,ab OR ‘Type II’:ti,ab OR ‘Type II’:ti,ab OR ‘Maturity Onset’:ti,ab OR ‘Adult Onset’:ti,ab OR ‘Slow Onset’:ti,ab OR ‘Noninsulin’:ti,ab OR ‘Non-insulin’:ti,ab OR ‘Ketosis Resistant’:ti,ab)) OR ‘NIDDM’:ti,ab OR ‘T2DM’:ti,ab |
| #2 | ‘impaired glucose tolerance’/exp OR ‘Prediabet*’:ti,ab |
| #3 | 'low carbohydrate diet'/exp OR 'ketogenic diet'/exp OR (‘Diet*’:ti,ab AND (‘Carbohydrate Restricted’:ti,ab OR ‘Carbohydrate-Restricted’:ti,ab OR ‘low-carbohydrate’:ti,ab OR ‘low carbohydrate’:ti,ab OR ‘ketogenic’:ti,ab)) |
| #4 | 'paleolithic diet'/exp OR (‘Diet*’:ti,ab AND (‘Paleo*’:ti,ab OR ‘Caveman’:ti,ab OR ‘Hunter Gatherer’:ti,ab OR ‘Hunter-Gatherer’:ti,ab OR ‘Stone Age’:ti,ab)) |
| #5 | 'protein diet'/exp OR 'Atkins diet'/exp OR (‘Diet*’:ti,ab AND (‘High-Protein’:ti,ab OR ‘High Protein’:ti,ab OR ‘South Beach’:ti,ab OR ‘Atkins’:ti,ab)) |
| #6 | 'low fat diet'/exp OR (‘Diet*’:ti,ab AND (‘Fat-Restricted’:ti,ab OR ‘Fat Restricted’:ti,ab OR ‘Low-Fat’:ti,ab OR ‘Low Fat’:ti,ab OR ‘Fat-Free’:ti,ab OR ‘Fat Free’:ti,ab)) |
| #7 | ‘low calorie diet’/exp OR (‘Calori*’:ti,ab AND (‘Low’:ti,ab OR ‘Restricted’:ti,ab) OR ‘low-calorie’:ti,ab OR ‘low calorie’:ti,ab) |
| #8 | 'Mediterranean diet'/exp OR (‘Diet*’:ti,ab AND ‘Mediterranean’:ti,ab) |
| #9 | 'vegetarian diet'/exp OR ((‘Diet*’:ti,ab AND (‘Vegetarian*’:ti,ab OR ‘Lacto-Vegetarian’:ti,ab)) OR ((‘Diet*’:ti,ab OR ‘Nutrition’:ti,ab) AND (‘Plant-Based’:ti,ab OR ‘Plant Based’:ti,ab))) OR ‘Vegetarianism’:ti,ab |
| #10 | 'vegan diet'/exp OR ‘Vegan*’:ti,ab |
| #11 | 'DASH diet'/exp OR ‘Dietary Approaches To Stop Hypertension’:ti,ab |
| #12 | 'Nordic diet'/exp |
| #13 | 'low glycemic index diet'/exp |
| #14 | 'high fiber diet'/exp |
| #15 | ‘Portfolio diet’:ti,ab |
| #16 | 'randomized controlled trial'/exp OR 'controlled clinical trial'/exp OR random*:ti,ab OR |

| No. | Search |
|---|---|
| | placebo:ti,ab OR trial:ti |
| #17 | 'gestation*':ti OR 'type 1':ti OR 'pregnan*':ti |
| #18 | #1 OR #2 |
| #19 | #3 OR #4 OR #5 OR #6 OR #7 OR #8 OR #9 OR #10 OR #11 OR #12 OR #13 OR #14 OR 15 |
| #20 | #18 AND #19 AND #16 AND [embase]/lim NOT #17 |

**S1.4 Web of Science**

Search time: 22:02, October 13, 2021 (GMT+8); 1 857 results
Search link: https://www.webofscience.com/wos/alldb/summary/3f2ec4e8-0625-4c0f-bd3c-caa3e34cea57-0cdcec9f/relevance/1

| No. | Search |
|---|---|
| #1 | TS=("Diabetes Mellitus, Type 2" OR ("Diabetes" AND ("Type 2" OR "Type II" OR "Type II" OR "Maturity Onset" OR "Adult Onset" OR "Slow Onset" OR "Noninsulin" OR "Non-insulin" OR "Ketosis Resistant")) OR "NIDDM" OR "T2DM") |
| #2 | TS=("Prediabetic State" OR "Prediabet*" OR "impaired glucose tolerance") |
| #3 | TS=("Diet, Carbohydrate-Restricted" OR "Diet, Ketogenic" OR ("Diet*" AND ("Carbohydrate Restricted" OR "Carbohydrate-Restricted" OR "low-carbohydrate" OR "low carbohydrate" OR "ketogenic"))) |
| #4 | TS=("Diet, Paleolithic" OR ("Diet*" AND ("Paleo*" OR "Caveman" OR "Hunter Gatherer" OR "Hunter-Gatherer" OR "Stone Age"))) |
| #5 | TS=("Diet, High-Protein" OR "Diet, High-Protein Low-Carbohydrate" OR ("Diet*" AND ("High-Protein" OR "High Protein" OR "South Beach" OR "Atkins"))) |
| #6 | TS=("Diet, Fat-Restricted" OR ("Diet*" AND ("Fat-Restricted" OR "Fat Restricted" OR "Low-Fat" OR "Low Fat" OR "Fat-Free" OR "Fat Free"))) |
| #7 | TS=("Caloric Restriction" OR ("Calori*" AND ("Low" OR "Restricted") OR "low-calorie" OR "low calorie")) |
| #8 | TS=("Diet, Mediterranean" OR ("Diet*" AND "Mediterranean")) |
| #9 | TS=("Diet, Vegetarian" OR (("Diet*" AND ("Vegetarian*" OR "Lacto-Vegetarian")) OR (("Diet*" OR "Nutrition") AND ("Plant-Based" OR "Plant Based"))) OR "Vegetarianism") |
| #10 | TS=("Diet, Vegan" OR "Vegan*") |
| #11 | TS=("DASH" OR "Dietary Approaches To Stop Hypertension") |
| #12 | TS=("Nordic*" AND "diet*") |
| #13 | TS=("low glycemic index diet*" OR "low-glycemic index diet*" OR "low glycaemic index diet*" OR "low-glycaemic index diet*") |
| #14 | TS=(("high fiber" OR "high-fiber" OR "high fibre" OR "high-fibre") AND "diet*") |
| #15 | TS=("Portfolio diet") |
| #16 | TS=("randomise*" OR "randomize*" OR ("random*" AND ("allocat*" OR "assign*")) OR ("blind*" AND ("single" OR "double" OR "treble" OR "triple"))) |
| #17 | TI=("gestation*" OR "pregnan*" OR "type 1") |
| #18 | #1 OR #2 |
| #19 | #3 OR #4 OR #5 OR #6 OR #7 OR #8 OR #9 OR #10 OR #11 OR #12 OR #13 OR #14 OR |

| No. | Search |
|---|---|
| | #15 |
| #20 | #16 AND #18 AND #19 NOT #17 |

**S1.5 CINAHL and OpenDissertations**

Search time: 00:25, October 14, 2021 (GMT+8); 297 results

| No. | Search |
|---|---|
| S1 | MH ("Diabetes Mellitus, Type 2") OR ( SU (("Diabetes" AND ("Type 2" OR "Type II" OR "Type II" OR "Maturity Onset" OR "Adult Onset" OR "Slow Onset" OR "Noninsulin" OR "Non-insulin" OR "Ketosis Resistant")) OR "NIDDM" OR "T2DM") ) |
| S2 | MH ("Prediabetic State") OR (SU ("Prediabet*" OR "impaired glucose tolerance") ) |
| S3 | MH ("Diet, Carbohydrate-Restricted" OR "Diet, Ketogenic") OR (SU (("Diet*" AND ("Carbohydrate Restricted" OR "Carbohydrate-Restricted" OR "low-carbohydrate" OR "low carbohydrate" OR "ketogenic"))) ) |
| S4 | MH ("Diet, Paleolithic") OR (SU (("Diet*" AND ("Paleo*" OR "Caveman" OR "Hunter Gatherer" OR "Hunter-Gatherer" OR "Stone Age"))) ) |
| S5 | MH ("Diet, High-Protein" OR "Diet, High-Protein Low-Carbohydrate") ) OR (SU (("Diet*" AND ("High-Protein" OR "High Protein" OR "South Beach" OR "Atkins"))) |
| S6 | MH ("Diet, Fat-Restricted") OR ( SU (("Diet*" AND ("Fat-Restricted" OR "Fat Restricted" OR "Low-Fat" OR "Low Fat" OR "Fat-Free" OR "Fat Free"))) ) |
| S7 | MH ("Caloric Restriction") OR (SU (("Calori*" AND ("Low" OR "Restricted") OR "low-calorie" OR "low calorie"))) |
| S8 | MH ("Diet, Mediterranean") OR (SU ("Diet*" AND "Mediterranean") ) |
| S9 | MH ("Diet, Vegetarian") OR (SU ((("Diet*" AND ("Vegetarian*" OR "Lacto-Vegetarian")) OR (("Diet*" OR "Nutrition") AND ("Plant-Based" OR "Plant Based"))) OR "Vegetarianism") ) |
| S10 | MH ("Diet, Vegan") OR (SU ("Diet*" AND "Vegan*")) |
| S11 | MH ("Dietary Approaches To Stop Hypertension") OR (SU ("DASH" OR "Dietary Approaches To Stop Hypertension")) |
| S12 | SU ("Nordic*" AND "diet*") |
| S13 | SU ("low glycemic index diet*" OR "low-glycemic index diet*" OR "low glycaemic index diet*" OR "low-glycaemic index diet*") |
| S14 | SU (("high fiber" OR "high-fiber" OR "high fibre" OR "high-fibre") AND "diet*") |
| S15 | SU ("Portfolio diet") |
| S16 | ( TX ("rct" OR "versus" OR "vs" OR "control group*" OR "treatment arm" OR (("blind*" OR "mask*" ) and ("single" OR "double" OR "triple" OR "treble")) OR ("trial" and ("control*" OR "comparative")) OR ("phase" and ("three" OR "III" OR "III")) OR "cross over" OR "random*" OR "factorial*" OR "placebo*" OR "assign*" OR "allocat*" OR "crossover*") ) OR ( MH ("Placebos" OR "Quantitative Studies" OR "Random Assignment" OR "Clinical Trials+") ) |
| S17 | ((MH ("Animals+" OR "Animal Studies")) OR (TI ("Animal Model")) ) NOT MH ("Human") |
| S18 | TI ("Gestation*" OR "Pregnan*" OR "Type 1") |
| S19 | TI ("Protocol" OR "Review") |
| S20 | S1 OR S2 |

| No. | Search |
| --- | --- |
| S21 | S3 OR S4 OR S5 OR S6 OR S7 OR S8 OR S9 OR S10 OR S11 OR S12 OR S13 OR S14 OR S15 |
| S22 | S16 NOT S17 |
| S23 | S18 OR S19 |
| S24 | S20 AND S21 AND S22 NOT S23 |

**S1.6 ProQuest**

Search time: 01:45, October 16, 2021 (GMT+8); 93 results
Search strategy:
(((MESH("Diabetes Mellitus, Type 2") OR AB,TI(("Diabetes" AND ("Type 2" OR "Type II" OR "Type II" OR "Maturity Onset" OR "Adult Onset" OR "Slow Onset" OR "Noninsulin" OR "Non-insulin" OR "Ketosis Resistant")) OR "NIDDM" OR "T2DM")) OR (MESH("Prediabetic State") OR AB,TI("Prediabet*" OR "impaired glucose tolerance"))) AND ((MESH("Diet, Carbohydrate-Restricted" OR "Diet, Ketogenic") OR AB,TI(("Diet*" AND ("Carbohydrate Restricted" OR "Carbohydrate-Restricted" OR "low-carbohydrate" OR "low carbohydrate" OR "ketogenic")))) OR (MESH("Diet, Paleolithic") OR AB,TI("Diet*" AND ("Paleo*" OR "Caveman" OR "Hunter Gatherer" OR "Hunter-Gatherer" OR "Stone Age"))) OR (MESH("Diet, High-Protein" OR "Diet, High-Protein Low-Carbohydrate") OR AB,TI(("Diet*" AND ("High-Protein" OR "High Protein" OR "South Beach" OR "Atkins")))) OR (MESH("Diet, Fat-Restricted") OR AB,TI(("Diet*" AND ("Fat-Restricted" OR "Fat Restricted" OR "Low-Fat" OR "Low Fat" OR "Fat-Free" OR "Fat Free")))) OR (MESH("Caloric Restriction") OR AB,TI(("Calori*" AND ("Low" OR "Restricted") OR "low-calorie" OR "low calorie"))) OR (MESH("Diet, Mediterranean") OR AB,TI(("Diet*" AND "Mediterranean"))) OR (MESH("Diet, Vegetarian") OR AB,TI((("Diet*" AND ("Vegetarian*" OR "Lacto-Vegetarian")) OR (("Diet*" OR "Nutrition") AND ("Plant-Based" OR "Plant Based"))) OR "Vegetarianism")) OR (MESH("Diet, Vegan") OR AB,TI("Vegan*")) OR (MESH("Dietary Approaches To Stop Hypertension") OR AB,TI("DASH" OR "Dietary Approaches To Stop Hypertension")) OR AB,TI("Nordic*" AND "diet*") OR AB,TI("low glycemic index diet*" OR "low-glycemic index diet*" OR "low glycaemic index diet*" OR "low-glycaemic index diet*") OR AB,TI(("high fiber" OR "high-fiber" OR "high fibre" OR "high-fibre") AND "diet*") OR AB,TI("Portfolio diet")) AND stype.exact("Dissertations & Theses")) AND (MESH("Placebos" OR "Quantitative Studies" OR "Random Assignment" OR "Clinical Trials") OR ALL("rct" OR "versus" OR "vs" OR "control group*" OR "treatment arm" OR (("blind*" OR "mask*" ) and ("single" OR "double" OR "triple" OR "treble")) OR ("trial" and ("control*" OR "comparative")) OR ("phase" and ("three" OR "III" OR "III")) OR "cross over" OR "random*" OR "factorial*" OR "placebo*" OR "assign*" OR "allocat*" OR "crossover*"))

**S1.7 Scopus**

Search time: 01:16, October 15, 2021 (GMT+8); 1 762 results
Search strategy:
( ( TITLE-ABS-KEY-AUTH ( "Diabetes Mellitus, Type 2" OR ( "Diabetes" AND ( "Type 2" OR "Type II" OR "Type II" OR "Maturity Onset" OR "Adult Onset" OR "Slow Onset" OR "Noninsulin" OR "Non-insulin" OR "Ketosis Resistant" ) ) OR "NIDDM" OR

"T2DM" ) OR TITLE-ABS-KEY ( "Prediabetic State" OR "Prediabet*" OR "impaired glucose tolerance" ) ) AND ( TITLE-ABS-KEY ( "Diet, Carbohydrate-Restricted" OR "Diet, Ketogenic" OR ( "Diet*" AND ( "Carbohydrate Restricted" OR "Carbohydrate-Restricted" OR "low-carbohydrate" OR "low carbohydrate" OR "ketogenic" ) ) ) OR TITLE-ABS-KEY ( "Diet, Paleolithic" OR ( "Diet*" AND ( "Paleo*" OR "Caveman" OR "Hunter Gatherer" OR "Hunter-Gatherer" OR "Stone Age" ) ) ) OR TITLE-ABS-KEY ( "Diet, High-Protein" OR "Diet, High-Protein Low-Carbohydrate" OR ( "Diet*" AND ( "High-Protein" OR "High Protein" OR "South Beach" OR "Atkins" ) ) ) OR TITLE-ABS-KEY ( "Diet, Fat-Restricted" OR ( "Diet*" AND ( "Fat-Restricted" OR "Fat Restricted" OR "Low-Fat" OR "Low Fat" OR "Fat-Free" OR "Fat Free" ) ) ) OR TITLE-ABS-KEY ( "Caloric Restriction" OR ( "Calori*" AND ( "Low" OR "Restricted" ) OR "low-calorie" OR "low calorie" ) ) OR TITLE-ABS-KEY ( "Caloric Restriction" OR ( "Calori*" AND ( "Low" OR "Restricted" ) OR "low-calorie" OR "low calorie" ) ) OR TITLE-ABS-KEY ( "Diet, Mediterranean" OR ( "Diet*" AND "Mediterranean" ) ) OR TITLE-ABS-KEY ( "Diet, Vegetarian" OR ( ( "Diet*" AND ( "Vegetarian*" OR "Lacto-Vegetarian" ) ) OR ( ( "Diet*" OR "Nutrition" ) AND ( "Plant-Based" OR "Plant Based" ) ) ) OR "Vegetarianism" ) OR TITLE-ABS-KEY ( "Diet, Vegan" OR "Vegan*" ) OR TITLE-ABS-KEY ( "DASH" OR "Dietary Approaches To Stop Hypertension" ) OR TITLE-ABS-KEY ( "Nordic*" AND "diet*" ) OR TITLE-ABS-KEY ( "low glycemic index diet*" OR "low-glycemic index diet*" OR "low glycaemic index diet*" OR "low-glycaemic index diet*" ) OR TITLE-ABS-KEY ( ( "high fiber" OR "high-fiber" OR "high fibre" OR "high-fibre" ) AND "diet*" ) OR TITLE-ABS-KEY ( "Portfolio diet" ) ) ) AND ( TITLE-ABS-KEY ( "randomise*" OR "randomize*" OR ( "random*" AND ( "allocat*" OR "assign*" ) ) OR ( "blind*" AND ( "single" OR "double" OR "treble" OR "triple" ) ) ) AND NOT TITLE ( "Protocol" OR "Review" ) ) AND NOT ( TITLE ( "gestation*" OR "pregnan*" OR "type 1" ) )

**S1.8 Clinicaltrials.gov**

Search time: 02:34, October 13, 2021 (GMT+8); 174 results
Search strategy:
Active, not recruiting, Completed, Terminated, Unknown status Studies | Interventional Studies | "Diabetes Mellitus, Type 2" OR "Prediabetic State" | "Diet, Carbohydrate-Restricted" OR "Diet, Paleolithic" OR "Diet, High-Protein" OR "Diet, High-Protein Low-Carbohydrate" OR "Diet, Fat-Restricted" OR "Caloric Restriction" OR "Diet, Mediterranean" OR "Diet, Vegetarian" OR "Diet, Vegan" OR "Dietary Approaches To Stop Hypertension" OR "Nordic" OR "low glycemic index diet" OR "high fiber diet" OR "Portfolio diet"
Also searched for Type 2 diabetes, Diabetes, Dietary and more.

**S1.9 Global Index Medicus**

Search time: October 13, 2021 (GMT+8); 65 results
Search strategy:
"diabetes" AND "diet" AND "randomized" AND ( type_of_study:("clinical_trials") AND la:("en" OR "zh"))

**S1.10 SinoMed (in Chinese)**

Search time: October 13, 2021 (GMT+8); 170 results

| No. | Search |
|---|---|
| #1 | "糖尿病, 2 型"[不加权:扩展] OR "糖尿病前期"[不加权:扩展] |
| #2 | "膳食, 低碳水化合物"[不加权:扩展] OR "生酮膳食"[不加权:扩展] OR "膳食，旧石器时代"[不加权:扩展] OR "膳食，限制脂肪"[不加权:扩展] OR "热量限制"[不加权:扩展] OR "膳食，地中海"[不加权:扩展] OR "膳食，素食者"[不加权:扩展] OR "膳食，纯素"[不加权:扩展] |
| #3 | "低碳水化合物"[常用字段:智能] OR "高蛋白"[常用字段:智能] OR "生酮"[常用字段:智能] OR "高蛋白低碳水化合物"[常用字段:智能] OR "旧石器时代"[常用字段:智能] OR "低卡路里"[常用字段:智能] OR "低热量"[常用字段:智能] OR "地中海"[常用字段:智能] OR "素食"[常用字段:智能] OR "纯素"[常用字段:智能] OR "北欧"[常用字段:智能] OR "低血糖指数"[常用字段:智能] OR "低血糖生成指数"[常用字段:智能] OR "低血糖负荷"[常用字段:智能] OR "高纤维"[常用字段:智能] OR "组合"[常用字段:智能] |
| #4 | "临床试验"[文献类型] OR "随机对照试验"[文献类型] |
| #5 | #2 OR #3 |
| #6 | #1 AND #4 AND #5 |

**S1.11 WanFang Med (in Chinese)**

Search time: October 13, 2021 (GMT+8); 539 results
Search strategy:
主题=("2 型糖尿病" OR "糖尿病前期") AND 主题=("膳食, 低碳水化合物" OR "生酮膳食" OR "膳食，旧石器时代" OR "膳食，限制脂肪" OR "热量限制" OR "膳食，地中海" OR "膳食，素食者" OR "膳食，纯素" OR "低碳水化合物" OR "高蛋白" OR "生酮" OR "高蛋白低碳水化合物" OR "旧石器时代" OR "低卡路里" OR "低热量" OR "地中海" OR "素食" OR "纯素" OR "北欧" OR "低血糖指数" OR "低血糖生成指数" OR "低血糖负荷" OR "高纤维" OR "组合") AND 主题=("随机对照试验" OR "随机试验" OR "对照")

**S1.12 CNKI (in Chinese)**

Search time: 02:03, October 14, 2021 (GMT+8); 463 results
Search strategy:
TKA=('2 型糖尿病'+'糖尿病前期') AND TKA=('膳食, 低碳水化合物'+'生酮膳食'+'膳食，旧石器时代'+'膳食，限制脂肪'+'热量限制'+'膳食，地中海'+'膳食，素食者'+'膳食，纯素'+'低碳水化合物'+'高蛋白'+'生酮'+'高蛋白低碳水化合物'+'旧石器时代'+'低卡路里'+'低热量'+'地中海'+'素食'+'纯素'+'北欧'+'低血糖指数'+'低血糖生成指数'+'低血糖负荷'+'高纤维'+'组合') AND TKA=('随机对照试验'+'随机试验'+'对照')

**S1.13 Google Scholar**

allintitle: ("diet" "type 2 diabetes") -"meta-analysis"
allintitle: ("diet" "prediabetes") -"meta-analysis"

**S1.14 Filters used for identifying RCTs**

1. https://ent.cochrane.org/sites/ent.cochrane.org/files/public/uploads/rct_filters.pdf
2. https://community.cochrane.org/sites/default/files/uploads/inline-files/Glanville_2019_HTAi_CINAHL-filter-poster.pdf
3. https://guides.library.harvard.edu/c.php?g=309982&p=2070466

#### File S2: Data extraction template

The data extraction template was made of five tables for characteristics of studies and arms, and various tables for continuous and dichotomous outcome variables, managed in MySQL. Structure of the database was:

**Table S2.1** Structure of the database for data extraction

| Table | Columns |
|---|---|
| basicinfo [a] | studyid, pubid, id, author_en, author_zh, name_en, name_zh, pub_year, comp_year, origin, type, fundings, conflicts, registry, reg_number |
| design [b] | studyid, id, center, crossover, masking, setting, patient_status, intensity, inclusion, exclusion, period, times, medication, insulin, exercise, objective, comorbidity, bias |
| arms [c] | studyid, arm_id, arm_label, arm_label_2, arm_label_3, size_initial, size |
| participants | studyid, arm_id, size_initial, size, ethnicity, sex_M, sex_F, MFR, age_mean, age_sd, weight_mean, weight_sd, bmi_mean, bmi_sd, duration_mean, duration_sd |
| nutrition [d] | studyid, arm_id, size, endpoint, energy, protein, fat, carb, GI, GL, MUFA, PUFA, fiber |
| continuous variables [e] | studyid, arm_id, size, endpoint, baseline_mean, baseline_sd, end_mean, end_sd, change_mean, change_sd, missing, cov<br>(For each variable there was a separate table) |
| dichotomous variables [f] | studyid, arm_id, size, endpoint, attrition, med_baseline_not, med_end_not, med_higher, med_lower, remission, notremission, hypoglycemia |

a. “id” was surname of the first author + year(s) of publication. “type” can be “thesis” or “journal”
b. “center” can be “single” or “multi”. “crossover” can be “parallel” or “crossover”. “times” referred to the times of measurement. “objective” can be “T2DM” or “PreD”. “comorbidity” can be “0” or “1”.
c. “arm_label” can be names of interventions or “control”. “arm_label_2” was for extra caloric restriction. “arm_label_3” was for other characteristics.
d. “carb” referred to carbohydrate.
e. “missing” was used for identifying if this study reported standard deviations of changes from baseline. “0” for yes and “1” for no. “cov” was for calculated correlation coefficients.
f. “med_xxx” for medication use; “remission” for the remission of T2DM/PreD.

#### File S3: Correlation coefficients for estimation

**Table S3.1** Correlation coefficients

| | FPG | $HbA_{1c}$ | FIns | IR | weight | BMI | WC | SBP | DBP | TG | TC | LDL | HDL |
|---|---|---|---|---|---|---|---|---|---|---|---|---|---|
| Corr | 0.4044 | 0.5048 | 0.6701 | 0.5962 | 0.9663 | 0.9517 | 0.8833 | 0.5872 | 0.5588 | 0.5465 | 0.5649 | 0.5743 | 0.7849 |

The correlation coefficients were the mean (unweighted) of all coefficients calculated from studies which reported standard deviations of changes from baseline, and the outliers were excluded using IBM SPSS Statistics 25.

#### File S4: Reason for exclusion

| Reference | Reason for Exclusion |
|---|---|
| 1. Chen C, Su Y, Yan L, Huang D, Xia M, Li F, et al., editors. [Application of low-glycemic index diets in the prevention and treatment of diabetes mellitus in the community and evaluation of their effects]. [8th Annual Academic Conference of Danone Nutrition Center]; 2005; Chengdu, Sichuan, China. [Chinese] | conference abstract, full text not found |
| 2. Chen M, editor. [Effects of low-glycemic-index diets replacement in the prevention and treatment of type 2 diabetes mellitus]. 2011. [Chinese] | |
| 3. Chandrakala G, Arpana G, Sreenivas T, Rao PV. Low-fat (<20%) diets prevent type 2 diabetes mellitus. Diabetes. 2012;61:A190. doi: 10.2337/db12-656-835. | |
| 4. Della Pepa G, Bozzetto L, Vetrani C, Monti S, Vitale M, Izzo A, et al. Effect of a portfolio diet targeting multiple dietary components on liver fat content in individuals with type 2 diabetes: an 8-week randomised controlled clinical trial. Diabetologia. 2019;62:S92-S. PubMed PMID: WOS:000485303800181. | |
| 5. Gryka A, Rolland C, Broom I. The effects of two low-carbohydrate, high-protein diets on body composition of obese patients with type 2 diabetes. Obesity Reviews. 2010;11:246. doi: 10.1111/j.1467-789X.2010.00763-7.x. | |
| 6. Hallberg S, McKenzie A, Creighton B, Volk B, Link T, Abner M, et al. Improvement in atherogenic dyslipidemia at 70 days following a reduced carbohydrate intervention for treatment of type 2 diabetes. Journal of Clinical Lipidology. 2016;10(3):665. | |
| 7. Huet D, Rizkalla SW, Rigoir A, Veronese A, Pacher N, Slama G. Metabolic effects of chronic low glycemic index diet in type 2 diabetes. Diabetes. 2001;50(Suppl 2):A367. PubMed PMID: CN-00725733. | |
| 8. Kabisch S, Hustig A, Dambeck U, Kemper M, Gerbracht C, Honsek C, et al. Investigation of sex-dependent effects of a one-year low-carb- vs low-fat intervention in patients with high-risk prediabetes - a randomised controlled trial. Diabetologia. 2021;64(SUPPL 1):62-. PubMed PMID: WOS:000696550100115. | |
| 9. Khazrai Y, Di Rosa C, Lattanzi G, Spiezia C, Beato I, Benvenuto D, et al. Effectiveness of a very low calorie ketogenic diet with meal replacements on body weight and composition. Obesity reviews. 2020;21(SUPPL 1). doi: 10.1111/obr.13118. PubMed PMID: CN-02230054. | |
| 10. Koutsovasilis A, Vlachos D, Diakoumopoulou E, Ganotopoulou A, Stathi C, Doulgerakis D, et al. A very low carbohydrate ketogenic diet compared with a low glycemic index reduced calorie diet in obese type 2 diabetic patients. Obesity Facts. 2012;5:196. doi: 10.1159/000258190. | |
| 11. Neelima GR, Chandrakala G, Arpana G, Jain AK, Rao PV. Long-term (3-year) effects of a reduced-fat diet in type 2 diabetes. Diabetes. 2009;58. | |
| 12. Nicholson AS. Effect of a low-fat, unrefined, vegan diet on type 2 diabetes. American journal of clinical nutrition. 1999;70(35):624S-5S. PubMed PMID: CN-00495013. | |
| 13. Pavithran N, Kumar H, Menon A, Ragasudha P, Pillai M, Sundaram K. 24-WEEK, LOW GI DIET DECREASES TRUNCAL FAT MASS IN SOUTH INDIANS WITH TYPE 2 DIABETES: a RANDOMIZED STUDY. Clinical nutrition (Edinburgh, Scotland). 2019;38:S222. doi: 10.1016/S0261-5614(19)32275-7. PubMed PMID: CN-01989007. | |
| 14. Skytte MGJ, Samkani AA, Petersen AD, Thomsen MN, Astrup A, Chabanova E, et al. A carbohydrate-reduced high-protein diet significantly reduces postprandial plasma and diurnal blood glucose in subjects with type 2 diabetes. Diabetes. 2017;66:A204. | |
| 15. Smiraglia M, Bott-a G, Orsi E. Differences in anthropometric parameters and glycemic control among individuals with type 2-diabetes and obesity following two different nutritional treatments. Journal of diabetes. 2013;5:86. doi: 10.1111/1753-0407.12032. PubMed PMID: CN-01027684. | |
| 16. Srichaikul K, Hertzog V, Dutton H, Kendall C, Sievenpiper J, Jenkins D. The effect of a low glycemic index diet on diabetic nephropathy. FASEB journal. 2015;29(1 Meeting Abstracts). PubMed PMID: CN-01080531. | |
| 17. Stentz FB, Ammons A. Remission of type 2 diabetes and metabolic parameters changes with a high-protein diet. Diabetes. 2020;69. doi: 10.2337/db20-1862-P. PubMed PMID: CN-02203745. | |
| 18. Tramontana F, Maddaloni E, Greci S, Defeudis G, Strollo R, Pozzilli P, et al. The effect of dietary fiber in combination with metformin therapy in type 2 diabetes. Diabetes. 2020;69. doi: 10.2337/db20-227-OR. PubMed PMID: CN-02203916. | |

| Reference | Reason for Exclusion |
|---|---|
| 19. Tucker S, Stentz F. Effect of macronutrients on metabolic parameters and remission of type 2 diabetes. Journal of Investigative Medicine. 2020;68(2):656-7. doi: 10.1136/jim-2020-SRM.544.<br>20. Vlachos D, Ganotopoulou A, Stathi C, Koutsovasilis A, Diakoumopoulou E, Doulgerakis D, et al. A low-carbohydrate protein sparing modified fast diet compared with a low glycaemic index reduced calorie diet in obese type 2 diabetic patients. Diabetologia. 2011;54:S355. doi: 10.1007/s00125-011-2276-4. | |
| 1. Liu Y. [Effect of dietary interventions on metabolic indicators of patients with type 2 diabetes mellitus]. 2015. [Chinese]<br>2. Te Morenga LA, McAuley KA, Docherty PD, Williams SM, Mann J. The effect of a high protein, high fibre diet on insulin sensitivity measured using the Dynamic Insulin Sensitivity and Secretion Test (DISST). Australasian medical journal. 2011;4(12):780. PubMed PMID: CN-01034669.<br>3. Kenneally S. How to reverse type 2 diabetes with plant-based diet. | full text not found |
| 1. Bai Y, Lü Q, Ma X. [Study on the effect of high dietary fiber and low glycemic index diet on gut microbiota and blood glucose in type 2 diabetic patients]. Zhongguo Quan Ke Yi Xue [Chinese General Practice]. 2016;19(20). [Chinese]<br>2. Li Z. [Effectiveness of a 30% low-carbohydrate diet combined with ligliptin in the treatment of elderly patients with T2DM]. Jian Kang Bi Du [Health Must-read]. 2021;6. [Chinese]<br>3. Liang S. [Effect of low glycemic index diet on glycolipid metabolism in type 2 diabetic patients]. Tang Niao Bing Lin Chuang [Diabetes World]. 2013;5(7). [Chinese]<br>4. Lin H. [Effect of low glycemic index diet on metabolic indicators in patients with type 2 diabetes]. Lin Chuang Yi Xue [Clinical Medicine]. 2019(9). [Chinese]<br>5. Ma H. [Effect of low glycemic index dietary substitution on the prevention and treatment of type 2 diabetes mellitus patients]. Zhongguo Wei Sheng Chan Ye [China Health Industry]. 2012;9(25). [Chinese]<br>6. Mo Z. [Low glycemic index diet in the dietary treatment of type 2 diabetes]. Yin Shi Bao Jian. 2018;47(5). [Chinese]<br>7. Sun F. [Effect of a low glycemic index diabetic diet on the metabolic and nutritional status of patients with type 2 diabetes]. Yi Yao Qian Yan [Journal of Frontiers of Medicine]. 2014;32. [Chinese]<br>8. Xu L, Li M, Zheng X, Xiao L, Chen X, Wei L. [Effect of dietary guidance with high dietary fiber and low glycemic index on glycemic and BMI in patients with type 2 diabetes]. Hu Li Shi Jian Yu Yan Jiu [Nursing Practice and Research]. 2019;16(19). [Chinese]<br>9. Zhang X, Sun L, Wang Z, Qi W, Sun W, Qi B, et al. [Effect of low glycemic index diabetic diet on glycolipid metabolism in type 2 diabetic patients]. Zhongguo Xian Dai Yao Wu Ying Yong [Chinese journal of modern drug application]. 2016;10(9). [Chinese]<br>10. Zhu X. [Effectiveness of low glycemic index diet in the nutritional treatment of newly-diagnosed type 2 diabetes mellitus patients]. Yang Sheng Bao Jian Zhi Nan [Health Guide]. 2020;47. [Chinese]<br>11. Díez-Espino J, Buil-Cosiales P, Serrano-Martínez M, Toledo E, Salas-Salvadó J, Martínez-González MÁ. Adherence to the Mediterranean Diet in Patients with Type 2 Diabetes Mellitus and HbA1c Level. Annals of Nutrition & Metabolism. 2011;58(1):74-8. doi: 10.1159/000324718. PubMed PMID: 104876703. Language: English. Entry Date: 20110428. Revision Date: 20200708. Publication Type: Journal Article.<br>12. Irct2017021432571N. The effect of the DASH diet in patients with type 2 diabetes. http://wwwwhoint/trialsearch/Trial2aspx?TrialID=IRCT2017021432571N1. 2017. PubMed PMID: CN-01894201.<br>13. Practice Based Nutrition Intervention. https://ClinicalTrials.gov/show/NCT01222429; 2010.<br>14. Nielsen JV, Jönsson E, Nilsson AK. Lasting improvement of hyperglycaemia and bodyweight: low-carbohydrate diet in type 2 diabetes. A brief report. Ups J Med Sci. 2005;110(2):179-83. Epub 2005/08/04. PubMed PMID: 16075898.<br>15. Reisin E. The benefit of the mediterranean-style diet in patients with newly diagnosed diabetes. Current Hypertension Reports. 2010;12(2):56-8. doi: 10.1007/s11906-010-0102-x.<br>16. Roncero-Ramos I, Alcala-Diaz JF, Rangel-Zuñiga OA, Gomez-Delgado F, Jimenez-Lucena R, García-Rios A, et al. Prediabetes diagnosis criteria, type 2 diabetes risk and | insufficient data |

| Reference | Reason for Exclusion |
|---|---|
| dietary modulation: the CORDIOPREV study. Clinical nutrition (Edinburgh, Scotland). 2020;39(2):492-500. doi: 10.1016/j.clnu.2019.02.027. PubMed PMID: CN-02127300.<br>17. Low Glycemic Index Diet for Type 2 Diabetics. https://ClinicalTrials.gov/show/NCT01063374; 2010.<br>18. A Very High Fiber Diet Versus a Low-carbohydrate Diet for Weight Loss. https://ClinicalTrials.gov/show/NCT01051674; 2010.<br>19. Dyson PA. Dietary advice for people with diabetes: the role of carbohydrate in dietary treatment and an assessment of video education [Ph.D.]. Ann Arbor: Oxford Brookes University (United Kingdom); 2010.<br>20. Larsen RN, Mann NJ, Maclean E, Shaw JE. The effect of high-protein, low-carbohydrate diets in the treatment of type 2 diabetes: a 12 month randomised controlled trial. Diabetologia. 2011;54(4):731-40. Epub 2011/01/20. doi: 10.1007/s00125-010-2027-y. PubMed PMID: 21246185.<br>21. Zainordin NA, Warman NAE, Mohamad AF, Abu Yazid FA, Ismail NH, Chen XW, et al. Safety and efficacy of very low carbohydrate diet in patients with diabetic kidney disease-A randomized controlled trial. Plos One. 2021;16(10). doi: 10.1371/journal.pone.0258507. PubMed PMID: WOS:000732519500056.<br>22. Wang R, Lou J, Hu Y, Liu J, Zhang Q. [Effect of different medical nutrition treatments on blood glucose volatility in elderly patients with type 2 diabetes mellitus]. Chongqing Yixue. 2015;44(28). [Chinese]<br>23. Liu S. [Effect of low-carbohydrate diets in women with impaired glucose regulation] [Master's degree]. Shanghai: Shanghai University of Sport; 2015. [Chinese] | |
| 1. Chen M, Chen Y, Hua L, Zong M, Xiao F, Yi Q, et al. [The effect of low glycemic index dietary substitution on the prevention and treatment of type 2 diabetes mellitus patients]. Zhong Hua Nei Fen Mi Dai Xie Za Zhi [Chinese Journal of Endocrinology and Metabolism]. 2012;1(28). [Chinese]<br>2. Chen Q, Zhang S, Wang W, Du Y, Zhang Z. editors. [Effect of high-fiber complex diet on body composition of obese patients with type 2 diabetes mellitus]. [The 7th National Academic Conference on Integrative Medicine and Nutrition]; 2016; Zhoushan, Zhejiang, China. [Chinese]<br>3. Liu H, Wei B, Ruan H. [Effect of 30% low-carbohydrate diet combined with liraglutide on glycolipid metabolism and body mass index in diabetic patients]. Yi Xue Shi Liao Yu Jian Kang [Medical Diet and Health]. 2021;12(19). [Chinese]<br>4. Liu L, Jiao L. [Analysis of the effects of different medical nutritional treatments on elderly patients with type 2 diabetes mellitus]. Zhongguo Yi Yao Zhi Nan [Guide of China Medicine]. 2018;16(20). [Chinese]<br>5. Wang R. [Effects of different medical nutrition treatments on blood glucose volatility and insulin resistance in elderly patients with type 2 diabetes mellitus]. 2015. [Chinese]<br>6. Pharmacist-led Therapeutic Nutritional Intervention in Type 2 Diabetes. https://ClinicalTrials.gov/show/NCT03181165; 2017.<br>7. Goday A, Bellido D, Sajoux I, Crujeiras AB, Burguera B, García-Luna PP, et al. Short-term safety, tolerability and efficacy of a very low-calorie-ketogenic diet interventional weight loss program versus hypocaloric diet in patients with type 2 diabetes mellitus. Nutr Diabetes. 2016;6(9):e230. Epub 2016/09/20. doi: 10.1038/nutd.2016.36. PubMed PMID: 27643725; PubMed Central PMCID: PMC5048014 Pronokal Protein Supplies Spain.<br>8. Hsu WC, Lau KHK, Matsumoto M, Moghazy D, Keenan H, King GL. Improvement of Insulin Sensitivity by Isoenergy High Carbohydrate Traditional Asian Diet: A Randomized Controlled Pilot Feasibility Study. Plos One. 2014;9(9). doi: 10.1371/journal.pone.0106851. PubMed PMID: WOS:000344317700020.<br>9. Lucotti P, Setola E, Monti LD, Galluccio E, Costa S, Sandoli EP, et al. Beneficial effects of a long-term oral L-arginine treatment added to a hypocaloric diet and exercise training program in obese, insulin-resistant type 2 diabetic patients. American Journal of Physiology - Endocrinology and Metabolism. 2006;291(5):E906-E12. doi: 10.1152/ajpendo.00002.2006. PubMed Central PMCID: Damor(Italy).<br>10. Mårtensson A, Stomby A, Tellström A, Ryberg M, Waling M, Otten J, et al. Using a Paleo Ratio to Assess Adherence to Paleolithic Dietary Recommendations in a Randomized Controlled Trial of Individuals with Type 2 Diabetes. Nutrients. 2021;13(3):969. doi: 10.3390/nu13030969. PubMed PMID: 149577539. Language: English. Entry Date: 20210408. Revision Date: 20210408. Publication Type: Article.<br>11. Milne RM, Mann JI, Chisholm AW, Williams SM. LONG-TERM COMPARISON OF 3 DIETARY PRESCRIPTIONS IN THE TREATMENT AT NIDDM. Diabetes Care. | wrong intervention |

| Reference | Reason for Exclusion |
|---|---|
| 1994;17(1):74-80. doi: 10.2337/diacare.17.1.74. PubMed PMID: WOS:A1994MP56300012. | |
| 12. Parillo M, Rivellese AA, Ciardullo AV, Capaldo B, Giacco A, Genovese S, et al. A high-monounsaturated-fat/low-carbohydrate diet improves peripheral insulin sensitivity in non-insulin-dependent diabetic patients. Metabolism. 1992;41(12):1373-8. Epub 1992/12/01. doi: 10.1016/0026-0495(92)90111-m. PubMed PMID: 1461145. | |
| 13. Pomerleau J, Verdy M, Garrel DR, Nadeau MH. EFFECT OF PROTEIN-INTAKE ON GLYCEMIC CONTROL AND RENAL-FUNCTION IN TYPE-2 (NON-INSULIN-DEPENDENT) DIABETES-MELLITUS. Diabetologia. 1993;36(9):829-34. doi: 10.1007/bf00400358. PubMed PMID: WOS:A1993LT54900008. | |
| 14. Raben A, Vestentoft PS, Brand-Miller J, Jalo E, Drummen M, Simpson L, et al. The PREVIEW intervention study: Results from a 3-year randomized 2 x 2 factorial multinational trial investigating the role of protein, glycaemic index and physical activity for prevention of type 2 diabetes. Diabetes Obes Metab. 2021;23(2):324-37. Epub 2020/10/08. doi: 10.1111/dom.14219. PubMed PMID: 33026154; PubMed Central PMCID: PMC8120810. | |
| 15. Ramal E, Champlin A, Bahjri K. Impact of a Plant-Based Diet and Support on Mitigating Type 2 Diabetes Mellitus in Latinos Living in Medically Underserved Areas. Am J Health Promot. 2018;32(3):753-62. Epub 2017/05/16. doi: 10.1177/0890117117706793. PubMed PMID: 28503930. | |
| 16. Rodríguez-Villar C, Manzanares JM, Casals E, Pérez-Heras A, Zambón D, Gomis R, et al. High-monounsaturated fat, olive oil-rich diet has effects similar to a high-carbohydrate diet on fasting and postprandial state and metabolic profiles of patients with type 2 diabetes. Metabolism. 2000;49(12):1511-7. Epub 2001/01/06. doi: 10.1053/meta.2000.18573. PubMed PMID: 11145109. | |
| 17. Toobert DJ, Glasgow RE, Strycker LA, Barrera Jr M, Radcliffe JL, Wander RC, et al. Biologic and quality-of-life outcomes from the Mediterranean Lifestyle Program: A randomized clinical trial. Diabetes Care. 2003;26(8):2288-93. doi: 10.2337/diacare.26.8.2288. | |
| 18. Low Glycemic Index Diets (With Pulses) in Type 2 Diabetes. https://ClinicalTrials.gov/show/NCT01063361; 2010. | |
| 19. Examining the Effects of Diet on Health With an Online Program. https://ClinicalTrials.gov/show/NCT01967992; 2013. | |
| 20. Yusof BNM, Talib RA, Kamaruddin NA, Karim NA, Chinna K, Gilbertson H. A low-GI diet is associated with a short-term improvement of glycaemic control in Asian patients with type 2 diabetes. Diabetes, Obesity and Metabolism. 2009;11(4):387-96. doi: 10.1111/j.1463-1326.2008.00984.x. PubMed Central PMCID: 19175374. | |
| 21. Andrews RC, Cooper AR, Montgomery AA, Norcross AJ, Peters TJ, Sharp DJ, et al. Diet or diet plus physical activity versus usual care in patients with newly diagnosed type 2 diabetes: the Early ACTID randomised controlled trial. The Lancet. 2011;378(9786):129-39. doi: 10.1016/S0140-6736(11)60442-X. | |
| 22. Liu H, Zhang M, Wu X, Wang C, Li Z. Effectiveness of a public dietitian-led diabetes nutrition intervention on glycemic control in a community setting in China. Asia Pac J Clin Nutr. 2015;24(3):525-32. doi: 10.6133/apjcn.2015.24.3.07. PubMed PMID: 26420196. | |
| 23. Komiyama N, Saito T, Hosaka Y, Aida K, Kaneko T, Sato A, et al. Effects of a 4-week 70% high carbohydrate/15% low fat diet on glucose tolerance and on lipid profiles. Diabetes research and clinical practice. 2004;64(1):11-8. doi: 10.1016/j.diabres.2003.10.002. PubMed PMID: CN-01759448. | |
| 24. Papakonstantinou E, Triantafillidou D, Panagiotakos DB, Koutsovasilis A, Saliaris M, Manolis A, et al. A high-protein low-fat diet is more effective in improving blood pressure and triglycerides in calorie-restricted obese individuals with newly diagnosed type 2 diabetes. Eur J Clin Nutr. 2010;64(6):595-602. Epub 2010/03/11. doi: 10.1038/ejcn.2010.29. PubMed PMID: 20216558. | |
| 25. Huang M-C, Hsu C-C, Wang H-S, Shin S-J. Prospective Randomized Controlled Trial to Evaluate Effectiveness of Registered Dietitian–Led Diabetes Management on Glycemic and Diet Control in a Primary Care Setting in Taiwan. Diabetes Care. 2009;33(2):233-9. doi: 10.2337/dc09-1092. | |
| 1. Chen J. [Analysis of the effect of low glycemic index diabetic diet on the metabolism and nutritional status of type 2 diabetic patients in 106 cases]. Dong Fang Yao Shan [Oriental Medicated Diet]. 2021;6. [Chinese] | wrong intervention duration |
| 2. Fan J, Wang Y, Zhang S. [The efficacy of hypoglycemic load diet in diabetic patients]. | |

| Reference | Reason for Exclusion |
|---|---|
| Chang Wai Yu Chang Nei Ying Yang [Parenteral and Enteral Nutrition]. 2015;22(2). [Chinese] | |
| 3. Li G, Pan Y, Lu Y. [Effect of low glycemic index food diet on blood glucose in patients with type 2 diabetes]. Jian Kang Bi Du [Health Must-read]. 2019;31. [Chinese] | |
| 4. Tao J, Xu R, Zhang J. [Low glycemic index diet in the dietary treatment of type 2 diabetes]. Hu Li Yan Jiu [Chinese Nursing Research]. 2009(8B). [Chinese] | |
| 5. Wang J. [Effectiveness of 30% low-carbohydrate diet combined with metformin in the treatment of type 2 diabetes mellitus]. Te Bie Jian Kang. 2020;32. [Chinese] | |
| 6. Zhang Q, Zhou J, Zhao Y, Xu Z. [Effect of a low glycemic index diabetic diet on the metabolic and nutritional status of patients with type 2 diabetes]. Zhongguo Quan Ke Yi Xue [Chinese General Practice]. 2012;15(12). [Chinese] | |
| 7. Zhang W. [Effectiveness of low glycemic index diet in the nutritional treatment of newly-diagnosed type 2 diabetes patients]. Lin Chuang Yi Yao Wen Xian Dian Zi Za Zhi [Electronic Journal of Clinical Medical Literature]. 2019;79(6). [Chinese] | |
| 8. Jarvi AE, Karlstrom BE, Granfeldt YE, Bjorck IE, Asp NGL, Vessby BOH. Improved glycemic control and lipid profile and normalized fibrinolytic activity on a low-glycemic index diet in type 2 diabetic patients. Diabetes Care. 1999;22(1):10-8. doi: 10.2337/diacare.22.1.10. PubMed PMID: WOS:000077689400003. | |
| 9. Effects of low-carbohydrate diet on glycemic control in type 2 diabetes. http://wwwwhoint/trialsearch/Trial2aspx?TrialID=JPRN-UMIN000000873. 2007. PubMed PMID: CN-01867027. | |
| 10. Diet Composition and Physical Inactivity on Insulin Sensitivity and β-cell Function. https://clinicaltrialsgov/show/NCT03013764. 2017. PubMed PMID: CN-02042344. | |
| 11. Samkani A, Skytte MJ, Kandel D, Kjaer S, Astrup A, Deacon CF, et al. A carbohydrate-reduced high-protein diet acutely decreases postprandial and diurnal glucose excursions in type 2 diabetes patients. British journal of nutrition. 2018;119(8):910-7. doi: 10.1017/S0007114518000521. PubMed PMID: CN-01913686. | |
| 12. Yang F, Yang Y, Zhu J, Zhang Z, Gu R, Hong J. [Clinical application of low glycemic index diet in hospitalized type 2 diabetic patients]. Zhongguo Shi Wu Yu Ying Yang [Food and Nutrition in China]. 2018;24(4). [Chinese] | |
| 1. The EDGe (End Diabetes Gisborne) trial. Using the whole-foods, plant-based diet in a community programme for people with obesity and diabetes. http://wwwwhoint/trialsearch/Trial2aspx?TrialID=ACTRN12617000541303. 2017. PubMed PMID: CN-01853790. | data not available |
| 2. The effect of different types of diet on the body weight and glycemic control of diabetics. http://wwwwhoint/trialsearch/Trial2aspx?TrialID=CTRI/2018/10/015896. 2018. PubMed PMID: CN-01947381. | |
| 3. Lifestyle Modification for Obesity-Related Type 2 Diabetes. https://ClinicalTrials.gov/show/NCT00415688; 2004. | |
| 4. Effects of a Mediterranean Style Diet on Vascular Health in Type 2 Diabetes. https://ClinicalTrials.gov/show/NCT00163683; 2003. | |
| 5. Low-Carbohydrate Dietary Pattern on Glycemic Outcomes Trial. https://ClinicalTrials.gov/show/NCT03675360; 2018. | |
| 6. Irct20170214032571N. The effect of a low calorie diet in patients with type 2 diabetes. http://wwwwhoint/trialsearch/Trial2aspx?TrialID=IRCT20170214032571N10. 2019. PubMed PMID: CN-01971677. | |
| 7. Irct20180724040579N. effect of aerobic training with ketogenic diet on insulin resistance and metabolic risk indices in middle-aged men with metabolic syndrome. http://wwwwhoint/trialsearch/Trial2aspx?TrialID=IRCT20180724040579N3. 2021. PubMed PMID: CN-02282419. | |
| 8. Prospective Interventional Study on Reversibility of Type 2 Diabetes Mellitus With Hypocaloric Diet. https://ClinicalTrials.gov/show/NCT04363710; 2017. | |
| 9. Low Carbohydrate Diet in Diabetic Kidney Disease. https://ClinicalTrials.gov/show/NCT04931030; 2019. | |
| 10. Practice Based Nutrition Intervention-2. https://ClinicalTrials.gov/show/NCT01700868; 2014. | |
| 11. Long Term Free Living Study With Modified Foods and Type 2 Diabetics. https://clinicaltrialsgov/show/NCT00198913. 2005. PubMed PMID: CN-01510782. | |
| 12. Effect of Low Glycaemic Index Diet on Blood Glucose Control in Chinese Type 2 Diabetic Patients. https://clinicaltrialsgov/show/NCT01542554. 2012. PubMed PMID: CN-01535834. | |
| 13. The Interaction Between Protein Intake, Gut Microbiota and Type 2 Diabetes in Subjects | |

| Reference | Reason for Exclusion |
|---|---|
| With Different Ethnic Backgrounds. https://ClinicalTrials.gov/show/NCT03732690; 2018.<br>14. RAcute Glycemic Effects of a Very Low Fat Diet in Type 2 Diabetes. https://ClinicalTrials.gov/show/NCT00006432; 2000.<br>15. Dietary Pattern and Metabolic Health Study. https://ClinicalTrials.gov/show/NCT03856762; 2019.<br>16. Contrasting Ketogenic and Mediterranean Diets in Individuals With Type 2 Diabetes and Prediabetes: The Keto-Med Trial. https://ClinicalTrials.gov/show/NCT03810378; 2019. | |
| 1. Clinical digest. Strict vegetarians at lower risk of developing diabetes. Nursing Standard. 2009;23(42):17-. doi: 10.7748/ns.23.42.17.s25. PubMed PMID: 105366754. Language: English. Entry Date: 20090724. Revision Date: 20200619. Publication Type: Journal Article.<br>2. Xu Y, Ji Q. [Translation of: A low–fat vegan diet and a conventional diabetes diet in the treatment of type 2 diabetes: a randomized, controlled, 74–wk clinical trial]. Zhong Hua Tang Niao Bing Za Zhi [Chinese Journal of Diabetes Mellitus]. 2009(2): 150. [Chinese] (This is a translation).<br>3. King A. The benefits of a Mediterranean diet. Nature Reviews Cardiology. 2013;10(5):239. doi: 10.1038/nrcardio.2013.36.<br>4. Singh M, Hung ES, Cullum A, Allen RE, Aggett PJ, Dyson P, et al. Lower carbohydrate diets for adults with type 2 diabetes. Diabetic Medicine. 2022;39(3). doi: 10.1111/dme.14674. PubMed Central PMCID: 34850972.<br>5. Conlon JM, Flatt PR, Bailey CJ. Recent advances in peptide-based therapy for Type 2 diabetes and obesity. Peptides. 2021;145. doi: 10.1016/j.peptides.2021.170652. PubMed Central PMCID: 34555424. | not a trial |
| 1. Numazawa R, Morohoshi M, Ozaku S, Yamazaki K, Nanba H, Uchida M, et al. Study on the effectiveness of a mild low-carbohydrate diet for improving glycemic control and relieving psychological burden in patients with type 2 diabetes. Journal of the Japan Diabetes Society. 2019;62(8):477-86. doi: 10.11213/tonyobyo.62.477. | not Chinese nor English |
| 1. Web-based low carbohydrate dietary intervention for adults with type 2 diabetes. http://wwwwhoint/trialsearch/Trial2aspx?TrialID=ACTRN12621000096853. 2021. PubMed PMID: CN-02242235.<br>2. Comparing High and Normal Protein Diets for the Dietary Remission of Type 2 Diabetes. https://ClinicalTrials.gov/show/NCT03832933; 2019.<br>3. A randomized control trial for a moderate carbohydrate diet compared to a low carbohydrate diet in overweight or obese individuals with type 2 diabetes mellitus or prediabetes. http://wwwwhoint/trialsearch/Trial2aspx?TrialID=ChiCTR1900022930. 2019. PubMed PMID: CN-01973152.<br>4. Ketogenic Diet Treatment of Obesity With Co-morbid Type 2 Diabetes Mellitus and/or Obstructive Sleep Apnea. https://ClinicalTrials.gov/show/NCT02069197; 2014.<br>5. Blood Pressure and Glucose Lowering Diet for Taiwanese. https://ClinicalTrials.gov/show/NCT01364337; 2010.<br>6. The LoBAG Diet and Type 2 Diabetes Mellitus. https://clinicaltrialsgov/show/NCT02717078. 2016. PubMed PMID: CN-01556787.<br>7. Effect of a Dietary Intervention on Intracellular Lipid, Insulin Sensitivity, and Glycemic Control in Type 2 Diabetes. https://clinicaltrialsgov/show/NCT04088981. 2019. PubMed PMID: CN-01968741.<br>8. Dietary Approaches to Stop Hypertension for Diabetes. https://clinicaltrialsgov/show/NCT04286555. 2020. PubMed PMID: CN-02088565.<br>9. REmote SUpport for Low-Carbohydrate Treatment of Type 2 Diabetes. https://clinicaltrialsgov/show/NCT04916314. 2021. PubMed PMID: CN-02278201.<br>10. Effects of Caloric Restriction in Obesity and Type 2 Diabetes. https://ClinicalTrials.gov/show/NCT01930136; 2013. | Trial not completed |
| 1. Safety of the Pronokal method in obese diabetic patients: the Diaprokal study. http://wwwwhoint/trialsearch/Trial2aspx?TrialID=ISRCTN55835754. 2014. PubMed PMID: CN-01822504. | paper not peer-reviewed |
| 1. Shao L, Zhao S. [Effect of nutritional therapy on patients with type 2 diabetes mellitus combined with pulmonary tuberculosis]. Zhongguo Xian Dai Yao Wu Ying Yong [Chinese journal of modern drug application]. 2018;12(14). [Chinese] | wrong outcome |

| Reference | Reason for Exclusion |
|---|---|
| 2. de Paula TP, Steemburgo T, de Almeida JC, Dall'Alba V, Gross JL, de Azevedo MJ. The role of Dietary Approaches to Stop Hypertension (DASH) diet food groups in blood pressure in type 2 diabetes. British Journal of Nutrition. 2012;108(1):155-62. doi: 10.1017/S0007114511005381. PubMed PMID: 104471052. Language: English. Entry Date: 20120713. Revision Date: 20200708. Publication Type: Journal Article. | |
| 3. Golan R, Tirosh A, Schwarzfuchs D, Harman-Boehm I, Thiery J, Fiedler GM, et al. Dietary intervention induces flow of changes within biomarkers of lipids, inflammation, liver enzymes, and glycemic control. Nutrition. 2012;28(2):131-7. doi: 10.1016/j.nut.2011.04.001. PubMed PMID: WOS:000299603400005. | |
| 4. Guldbrand H, Lindström T, Dizdar B, Bunjaku B, Östgren CJ, Nystrom FH, et al. Randomization to a low-carbohydrate diet advice improves health related quality of life compared with a low-fat diet at similar weight-loss in Type 2 diabetes mellitus. Diabetes Res Clin Pract. 2014;106(2):221-7. Epub 2014/10/02. doi: 10.1016/j.diabres.2014.08.032. PubMed PMID: 25271116. | |
| 5. Investigating the limits of reversibility of type 2 diabetes. http://wwwwhoint/trialsearch/Trial2aspx?TrialID=ISRCTN88634530. 2012. PubMed PMID: CN-01852328. | |
| 6. Kahleova H, Klemetova M, Herynek V, Kolarova M, Herynek S, Hill M, et al. The effect of a vegetarian vs. Conventional hypocaloric diabetic diet on thigh adipose tissue distribution in subjects with type 2 diabetes. Diabetes. 2017;66:A202-. PubMed PMID: CN-01724298. | |
| 7. Luscombe ND, Clifton PM, Noakes M, Parker B, Wittert G. Effects of energy-restricted diets containing increased protein on weight loss, resting energy expenditure, and the thermic effect of feeding in type 2 diabetes. Diabetes Care. 2002;25(4):652-7. Epub 2002/03/29. doi: 10.2337/diacare.25.4.652. PubMed PMID: 11919120. | |
| 8. A Randomized Cross-over Trial of the Postprandial Effects of Three Different Diets in Patients With Type 2 Diabetes. https://ClinicalTrials.gov/show/NCT01522157; 2012. | |
| 9. Diabetes Dietary Study- Low Carbohydrate and Low-Fat Diets in Type 2 Diabetes. https://ClinicalTrials.gov/show/NCT00795691; 2004. | |
| 10. Turner-McGrievy GM, Barnard ND, Cohen J, Jenkins DJ, Gloede L, Green AA. Changes in nutrient intake and dietary quality among participants with type 2 diabetes following a low-fat vegan diet or a conventional diabetes diet for 22 weeks. J Am Diet Assoc. 2008;108(10):1636-45. Epub 2008/10/18. doi: 10.1016/j.jada.2008.07.015. PubMed PMID: 18926128. | |
| 11. Wolever TMS, Chiasson J-L, Josse RG, Leiter LA, Maheux P, Rabasa-Lhoret R, et al. Effects of Changing the Amount and Source of Dietary Carbohydrates on Symptoms and Dietary Satisfaction Over a 1-Year Period in Subjects with Type 2 Diabetes: Canadian Trial of Carbohydrates in Diabetes (CCD). Canadian Journal of Diabetes. 2017;41(2):164-76. doi: 10.1016/j.jcjd.2016.08.223. PubMed PMID: 122370178. Language: English. Entry Date: 20180117. Revision Date: 20190202. Publication Type: Article. | |
| 12. Chen Y-S, Chen Y-H. [Effect of dietary education on low glycemic index foods on blood glucose in patients with type 2 diabetes]. Hu Li Xue Za Zhi [Journal of Nursing Science]. 2007;22(19). [Chinese] | |
| 13. Kakoschke N, Zajac IT, Tay J, Luscombe-Marsh ND, Thompson CH, Noakes M, et al. Effects of very low-carbohydrate vs. high-carbohydrate weight loss diets on psychological health in adults with obesity and type 2 diabetes: a 2-year randomized controlled trial. Eur J Nutr. 2021;60(8):4251-62. doi: 10.1007/s00394-021-02587-z. PubMed Central PMCID: 34018052. | |
| 1. Bradley U, Spence M, Courtney CH, McKinley MC, Ennis CN, McCance DR, et al. Low-fat versus low-carbohydrate weight reduction diets: effects on weight loss, insulin resistance, and cardiovascular risk: a randomized control trial. Diabetes. 2009;58(12):2741-8. Epub 2009/09/02. doi: 10.2337/db09-0098. PubMed PMID: 19720791; PubMed Central PMCID: PMC2780863. | wrong patients |
| 2. Debont AJ, Baker IA, Stleger AS, Sweetnam PM, Wragg KG, Stephens SM, et al. A RANDOMIZED CONTROLLED TRIAL OF THE EFFECT OF LOW FAT DIET ADVICE ON DIETARY-RESPONSE IN INSULIN INDEPENDENT DIABETIC WOMEN. Diabetologia. 1981;21(6):529-33. doi: 10.1007/bf00281543. PubMed PMID: WOS:A1981MW24500004. | |
| 3. Paleolithic Diet in the Treatment of Glucose Intolerance. https://ClinicalTrials.gov/show/NCT00419497; 2003. | |
| 4. Kahleova H, Petersen KF, Shulman GI, Alwarith J, Rembert E, Tura A, et al. A dietary | |

| Reference | Reason for Exclusion |
| --- | --- |
| intervention to alter insulin sensitivity, intramyocellular and hepatocellular lipids, postprandial metabolism, and body weight: a 16-week randomised trial. Diabetologia. 2020;63(SUPPL 1):S16-S7. doi: 10.1007/s00125-020-05221-5. PubMed PMID: CN-02230001. | |
| 5. Liese AD, Bortsov A, Günther AL, Dabelea D, Reynolds K, Standiford DA, et al. Association of DASH diet with cardiovascular risk factors in youth with diabetes mellitus: the SEARCH for Diabetes in Youth study. Circulation. 2011;123(13):1410-7. doi: 10.1161/CIRCULATIONAHA.110.955922. PubMed PMID: 104866023. Language: English. Entry Date: 20110617. Revision Date: 20161117. Publication Type: journal article. | |
| 6. McAuley KA, Hopkins CM, Smith KJ, McLay RT, Williams SM, Taylor RW, et al. Comparison of high-fat and high-protein diets with a high-carbohydrate diet in insulin-resistant obese women. Diabetologia. 2005;48(1):8-16. Epub 2004/12/24. doi: 10.1007/s00125-004-1603-4. PubMed PMID: 15616799. | |
| 7. An 18-month Trial of a Low Glycemic Load Diet. https://clinicaltrialsgov/show/NCT00130299. 2005. PubMed PMID: CN-02016755. | |
| 8. Sacks FM, Carey VJ, Anderson CA, Miller ER, 3rd, Copeland T, Charleston J, et al. Effects of high vs low glycemic index of dietary carbohydrate on cardiovascular disease risk factors and insulin sensitivity: the OmniCarb randomized clinical trial. Jama. 2014;312(23):2531-41. Epub 2014/12/17. doi: 10.1001/jama.2014.16658. PubMed PMID: 25514303; PubMed Central PMCID: PMC4370345. | |
| 9. Shai I. The effect of low-carb, mediterranean and low-fat diets on renal function; a 2-year dietary intervention randomized controlled trial (direct). Obesity facts. 2012;5:19-. doi: 10.1159/000171026. PubMed PMID: CN-01786377. | |
| 1. Ding H, Shao J, Zhe W, Zhang Y. [Application of low glycemic index diet in nutritional intervention for Uyghur people with diabetes]. Zhongguo Man Xing Bing Yu Fang Yu Kong Zhi [Chinese Journal of Prevention and Control of Chronic Diseases]. 2010(2). | same dataset of included studies |
| 2. Huang J, Xia J, Zhou W, Tang D, Zhang S, Cheng F, et al. [Effect of combined low-glycemic index and low-glycemic load diets on glucolipid metabolism in patients with type 2 diabetes]. Zhong Hua Tang Niao Bing Za Zhi [Chinese Journal of Diabetes Mellitus]. 2014(7). [Chinese] | |
| 3. Low Carbohydrate Diet Compared to Calorie and Fat Restricted Diet in Patients With Obesity and Type II Diabetes. https://ClinicalTrials.gov/show/NCT00108459; 2004. | |
| 4. Barnard N, Cohen J, Jenkins DJ, Turner-McGrievy G, Ferdowsian H. Randomized clinical trial of a plant-based diet for glycemic, lipid, and weight control in type 2 diabetes: Follow-up results. Diabetes. 2007;56:A448-A. PubMed PMID: WOS:000246930203091. | |
| 5. Barnard N, Cohen J, Jenkins DJA, Turner-McGrievy G. Effect of a plant-based diet on glycemic control and cardiovascular risk factors in individuals with type 2 diabetes: A randomized clinical trial. Diabetes. 2006;55:A8-A. PubMed PMID: WOS:000238055800036. | |
| 6. A randomized controlled study to observe the effect of loosely low carbohydrate diet on metabolism with type 2 diabetes. http://wwwwhoint/trialsearch/Trial2aspx?TrialID=ChiCTR-TRC-14004277. 2014. PubMed PMID: CN-01871372. | |
| 7. Elhayany A, Lustman A, Abel R, Attal-Singer J, Vinker S. A low carbohydrate Mediterranean diet improves cardiovascular risk factors and diabetes control among overweight patients with type 2 diabetes mellitus: a 1-year prospective randomized intervention study. Diabetes Obes Metab. 2010;12(3):204-9. Epub 2010/02/16. doi: 10.1111/j.1463-1326.2009.01151.x. PubMed PMID: 20151996. | |
| 8. Fogelholm M, Larsen TM, Westerterp-Plantenga M, MacDonald I, Martinez JA, Handjieva-Darlenska T, et al. PREVIEW-Design, methods and baseline participant description of an international intervention to prevent type-2 diabetes. Annals of nutrition and metabolism. 2015;67:414. doi: 10.1159/000440895. PubMed PMID: CN-01160326. | |
| 9. Hospital B, Copenhagen Uo, Aarhus Uo. Cut Down on Carbohydrate Usage in the Diet of Type 2 Diabetes. https://ClinicalTrials.gov/show/NCT02764021; 2016. | |
| 10. Paleolithic Diet in the Treatment of Diabetes Type 2 in Primary Health Care. https://ClinicalTrials.gov/show/NCT00435240; 2005. | |
| 11. The Effectiveness of Low Carbohydrate Diet in Reducing Polypharmacy for Patients With Type 2 Diabetes Mellitus. https://ClinicalTrials.gov/show/NCT03176056; 2016. | |
| 12. A Reduced-carbohydrate Diet High in Monounsaturated Fats in Type 2 Diabetes. | |

| Reference | Reason for Exclusion |
| --- | --- |
| https://ClinicalTrials.gov/show/NCT03068078; 2016. | |
| 13. Hrachovinová T, Kahleová H, Hackerová P, Pelikánová T. Changes in eating behavior, depression and quality of life in patients with type 2 diabetes after a 3-months-intervention with low-fat vegetarian diet and conventional diabetic diet. Diabetologia. 2009;52(S1):S392. doi: 10.1007/s00125-009-1445-1. | |
| 14. Irct2014011416223N. The effect of diet therapy as epigenetic factors in the control of diabetic complications. http://wwwwhoint/trialsearch/Trial2aspx?TrialID=IRCT2014011416223N1. 2014. PubMed PMID: CN-01815229. | |
| 15. Kitabchi AE, Brewer A, Wan J, Sands C, Stentz FB. Remission of impaired glucose tolerance (IGT) to normal glucose tolerance (NGT) in obese adults with high protein vs. High carbohydrate diet. Diabetes. 2015;64:A23-A4. doi: 10.2337/db151385. | |
| 16. Krebs JD, Elley CR, Parry-Strong A, Lunt H, Drury PL, Bell DA, et al. Two year randomised controlled trial of high-protein versus high-carbohydrate diet in type 2 diabetes: Diabetes excess weight loss (DEWL). Diabetes. 2011;60:A213. doi: 10.2337/db11-716-867. | |
| 17. Lindström T, Bachrach-Lindström M, Guldbrand H, Dizdar B, Bunjaku B, Östgren C, et al. Randomisation to a low-carbohydrate diet improves health related quality of life compared with a low-fat diet at similar weight loss in type 2 diabetes. Diabetologia. 2013;56:S347. doi: 10.1007/s00125-013-3012-z. | |
| 18. Lyckorna V. Comparison Between Low Carbohydrate Diet and Traditionally Recommended Diabetic Diet in the Treatment of Diabetes Mellitus Type 2. https://ClinicalTrials.gov/show/NCT01005498; 2009. | |
| 19. McLaughlin T, Carter S, Abbasi F, Lamendola C, Reaven G. Comparison of moderately low-fat vs low-carbohydrate diets for weight loss in patients with diet-controlled diabetes: A randomized trial. Diabetes. 2006;55:A77-A8. PubMed PMID: WOS:000238055800332. | |
| 20. Plant-Based Dietary Intervention in Type 2 Diabetes. https://ClinicalTrials.gov/show/NCT00276939; 2003. | |
| 21. Canadian Trial of Dietary Carbohydrates in Diabetes. https://clinicaltrialsgov/show/NCT00223574. 2005. PubMed PMID: CN-01511621. | |
| 22. Dietary Control of Type 2 Diabetes: low-Carbohydrate Mediterranean Diet Versus Low-Fat Diet. https://clinicaltrialsgov/show/NCT00725257. 2008. PubMed PMID: CN-02014585. | |
| 23. The Effect of DASH Diet on the Cardiometabolic Risks and Hepatic Function Among Type 2 Diabetic Patients. https://clinicaltrialsgov/show/NCT01049321. 2010. PubMed PMID: CN-02015977. | |
| 24. NJ D, N T, C S. [Effect of a 1-year low-carbohydrate or low-fat dietary intervention on weight and blood glucose in patients with type 2 diabetes]. Yao Pin Ping Jia [Drug Evaluation]. 2009(10). Epub 20091215. | |
| 25. Nystrom FH, Östgren CJ, Lindström T, Bachrach-Lindstrom M, Schöld AK, Dizdar B, et al. A high fat diet improves glycaemic control compared with low fat diet: A 24-month randomised prospective study of patients with type 2 diabetes in primary health care. Diabetologia. 2011;54:S358. doi: 10.1007/s00125-011-2276-4. | |
| 26. Pedersen E, Jesudason D, Clifton P. High protein weight loss diets in obese subjects with type 2 diabetes mellitus. Obesity Reviews. 2014;15:172. doi: 10.1111/obr.12151. | |
| 27. Preventing Renal Functional Abnormalities With Calorie Restriction in Subjects With Abdominal Obesity and Type 2 Diabetes at Increased Renal and Cardiovascular Risk. https://ClinicalTrials.gov/show/NCT01213212; 2009. | |
| 28. A Study to Assess the Effect of a Normal vs. High Protein Diets in Carbohydrates Metabolism in Obese Subjects With Diabetes or Prediabetes. https://ClinicalTrials.gov/show/NCT02559479; 2015. | |
| 29. Saslow L, Moskowitz JT, Mason AE, Kim S, Goldman V, Hecht F. A RANDOMIZED TRIAL OF AN INTERNET-DELIVERED INTERVENTION COMPARING VERY LOW-CARBOHYDRATE AND MYPLATE DIETS IN TYPE 2 DIABETES. Annals of Behavioral Medicine. 2016;50:S253-S. PubMed PMID: WOS:000526998301181. | |
| 30. Sato J, Kanazawa A, Makita S, Hatae C, Komiya K, Shimizu T, et al. A randomized controlled trial of 130 g/day low-carbohydrate diet in type 2 diabetes with poor glycemic control. Clin Nutr. 2017;36(4):992-1000. Epub 2016/07/31. doi: 10.1016/j.clnu.2016.07.003. PubMed PMID: 27472929. | |
| 31. Sato J, Kanazawa A, Makita S, Hatae C, Komiya K, Shimizu T, et al. The efficacy and safety of 130g/day low-carbohydrate diet in Japanese type 2 diabetes patients with poor glycemic control. Diabetes. 2016;65:A394. doi: 10.2337/db16-1375-1656. | |

| Reference | Reason for Exclusion |
|---|---|
| 32. Skytte MJ, Samkani AA, Petersen AD, Thomsen MN, Astrup A, Chabanova E, et al. A carbohydrate-reduced high-protein diet significantly reduces HbA1c, diurnal and prandial plasma glucose in weight stable subjects with type 2 diabetes. Diabetologia. 2018;61:S332-S3. doi: 10.1007/s00125-018-4693-0. PubMed PMID: CN-01647098. | |
| 33. Stentz FB, Ammons A, Christman JV. Remission of type 2 diabetes and decreased inflammatory markers with a high-protein diet. Diabetes. 2021;70(SUPPL 1). doi: 10.2337/db21-201-OR. | |
| 34. Stentz FB, Kitabchi A. Remission of prediabetes and improved metabolic parameters with a high-protein diet. Diabetes. 2016;65:A509. doi: 10.2337/db16-1771-2041. | |
| 35. Stentz FB, Mikhael A, Kineish O, Christman J, Sands C. High protein diet leads to prediabetes remission and positive changes in incretins and cardiovascular risk factors. Nutr Metab Cardiovasc Dis. 2021;31(4):1227-37. Epub 2021/02/08. doi: 10.1016/j.numecd.2020.11.027. PubMed PMID: 33549435. | |
| 36. Taghrid L, Rizkalla SW, Huet D, Rigoir A, Veronese A, Slama G. Improvement of glycemic control and plasma lipid levels by chronic low glycemic index diet in type 2 diabetes. Diabetes. 2003;52(Suppl 1):A72. PubMed PMID: CN-00725873. | |
| 37. Tay J, Luscombe-Marsh N, Thompson C, Noakes M, Buckley J, Wittert G, et al. LONG-TERM EFFECTS OF A LOW CARBOHYDRATE, LOW SATURATED FAT DIET VERSUS A CONVENTIONAL HIGH CARBOHYDRATE, LOW FAT DIET IN TYPE 2 DIABETES: A RANDOMISED TRIAL. Diabetes Research and Clinical Practice. 2014;106:S34-S. doi: 10.1016/s0168-8227(14)70272-4. PubMed PMID: WOS:000361124300067. | |
| 38. Tay J, Thompson CH, Luscombe-Marsh ND, Noakes M, Buckley JD, Wittert GA, et al. Long-Term Effects of a Very Low Carbohydrate Compared With a High Carbohydrate Diet on Renal Function in Individuals With Type 2 Diabetes: A Randomized Trial. Medicine (Baltimore). 2015;94(47):e2181. Epub 2015/12/04. doi: 10.1097/md.0000000000002181. PubMed PMID: 26632754; PubMed Central PMCID: PMC5059023. | |
| 39. Low Glycemic Index Diets vs. High Cereal Fibre Diets in Type 2 Diabetes. https://ClinicalTrials.gov/show/NCT00438698; 2004. | |
| 40. Tramontana F, Maddaloni E, Greci S, Defeudis G, Strollo R, Pozzilli P, et al. The effect of dietary fiber on glycaemic control in patients with type 2 diabetes on metformin monotherapy. Diabetologia. 2020;63(SUPPL 1):S105-. doi: 10.1007/s00125-020-05221-5. PubMed PMID: CN-02231149. | |
| 41. Turner-McGrievy GM, Barnard ND, Cohen J, Jenkins DJA, Gloede L, Green AA. Changes in Nutrient Intake and Dietary Quality among Participants with Type 2 Diabetes Following a Low-Fat Vegan Diet or a Conventional Diabetes Diet for 22 Weeks. Journal of the American Dietetic Association. 2008;108(10):1636-45. doi: 10.1016/j.jada.2008.07.015. PubMed PMID: WOS:000259873600020. | |
| 42. Visek J, Lacigova S, Cechurova D, Blaha V, Rusavy Z. Comparison of the impact of a low-glycemic index diet and a commonly used diabetic diet a randomized crossover study. Clinical nutrition, supplement. 2011;6(1):117. PubMed PMID: CN-01034620. | |
| 43. Watson N, Dyer K, Buckley J, Brinkworth G, Coates A, Parfitt G, et al. Effects of Low-Fat Diets Differing in Protein and Carbohydrate Content on Cardiometabolic Risk Factors during Weight Loss and Weight Maintenance in Obese Adults with Type 2 Diabetes. Nutrients. 2016;8(5). Epub 2016/05/18. doi: 10.3390/nu8050289. PubMed PMID: 27187457; PubMed Central PMCID: PMC4882702. | |
| 44. Westman E, Yancy W, Marquart M, Hepburn J. A randomized, controlled trial of a low-carbobydrate ketogenic diet vs. a low-glycemic index diet for type 2 diabetes. Obesity Research. 2005;13:A140-A. PubMed PMID: WOS:000232088800541. | |
| 45. Westman EC, Yancy WS, Jr., Marquart ML, Hepburn J. A randomized, controlled trial of a low-glycemic index vs. a low-carbohydrate, ketogenic diet for type 2 diabetes. Diabetes. 2006;55:A8-A. PubMed PMID: WOS:000238055800035. | |
| 46. Wycherley TP, Thompson CH, Buckley JD, Luscombe-Marsh ND, Noakes M, Wittert GA, et al. Long-term effects of weight loss with a very-low carbohydrate, low saturated fat diet on flow mediated dilatation in patients with type 2 diabetes: A randomised controlled trial. Atherosclerosis. 2016;252:28-31. Epub 2016/08/06. doi: 10.1016/j.atherosclerosis.2016.07.908. PubMed PMID: 27494448. | |
| 47. Yamada S, Yamada Y, Irie J. A non-calorie-restricted non-ketogenic low-carbohydrate diet is effective as an alternative therapy for patients with type 2 diabetes. Diabetes. 2013;62:A192-A3. doi: 10.2337/db13-680-858. | |
| 48. Wolever TMS, Mehling C. High-carbohydrate-low-glycaemic index dietary advice improves glucose disposition index in subjects with impaired glucose tolerance. British | |

| Reference | Reason for Exclusion |
|---|---|
| Journal of Nutrition. 2002;87(5):477-87. doi: 10.1079/bjn2002568. PubMed PMID: WOS:000175540000009. | |
| 49. Sun J, Chen Y, Zong M, Zhang X, Sun S, Wu Y, et al. [Effects of dietary nutrition intervention on glucolipid metabolism and antioxidant capacity in elderly diabetic patients]. Lao Nian Yi Xue Yu Bao Jian [Geriatrics & Health Care]. 2008;14(2). [Chinese] | |
| 50. Zhang T, Zhang F, Cao Y, Ma Y. [Effect of low-carbohydrate diet in patients with newly-diagnosed type 2 diabetes]. Yi Xue Xin Xi – Shang Xun Kan [Medical Information]. 2016;29(35). | |
| 1. Alzahrani AH, Skytte MJ, Samkani A, Thomsen MN, Astrup A, Ritz C, et al. Effects of a Self-Prepared Carbohydrate-Reduced High-Protein Diet on Cardiovascular Disease Risk Markers in Patients with Type 2 Diabetes. Nutrients. 2021;13(5):1694. doi: 10.3390/nu13051694. PubMed PMID: 150499135. Language: English. Entry Date: 20210603. Revision Date: 20210603. Publication Type: Article.<br>2. The Effects of a Novel Lifestyle Intervention Program on Insulin Sensitivity in Type 2 Diabetes. https://ClinicalTrials.gov/show/NCT04509245; 2018.<br>3. Effects of a low-carbohydrate diet on carotid artery intima-media thickness,urinary albumin excretion and eGFR in patients with type 2 diabetes: a 2-year intervention study. http://wwwwhoint/trialsearch/Trial2aspx?TrialID=JPRN-UMIN000004717. 2011. PubMed PMID: CN-01797807.<br>4. Hussain TA, Mathew TC, Dashti AA, Asfar S, Al-Zaid N, Dashti HM. Effect of low-calorie versus low-carbohydrate ketogenic diet in type 2 diabetes. Nutrition. 2012;28(10):1016-21. Epub 2012/06/08. doi: 10.1016/j.nut.2012.01.016. PubMed PMID: 22673594.<br>5. Sang D, Lu Z, Feng X, Zeng L, Liu L, Jiang F, et al. [Effect of a moderate low-carbohydrate diet on cardiovascular risk factors in overweight/obese patients with newly diagnosed type 2 diabetes mellitus]. Zhongguo Xun Zheng Xin Xue Guan Yi Xue Za Zhi [Chinese Journal of Evidence-Based Cardiovascular Medicine]. 2018;6(10). [Chinese]<br>6. Malinska H, Klementová M, Kudlackova M, Veleba J, Hoskova E, Oliyarnyk O, et al. A plant-based meal reduces postprandial oxidative and dicarbonyl stress in men with diabetes or obesity compared with an energy- and macronutrient-matched conventional meal in a randomized crossover study. Nutr Metab. 2021;18(1). doi: 10.1186/s12986-021-00609-5. | wrong study design |
| 1. Shao M, Qiang N, Zhang M. [Effects of low-carbohydrate diets on energy and glucose metabolism in newly diagnosed diabetic patients]. Taishan Yi Xue Yuan Xue Bao [Journal of Taishan Medical College]. 2018;39(4). [Chinese]<br>2. Xie X, Li Z, You Y. [Observations on the effects of low-carbohydrate diets on energy and glucose metabolism in newly diagnosed diabetic patients]. Ji Ceng Yi Xue Lun Tan [The Medical Forum]. 2019;23(5). [Chinese]<br>3. Wang R, Gao Y, Zhang J, Hou S. [Effect of low glycemic index diet on serum lipids in diabetic patients]. Zhongguo Yi Kan [Chinese Journal of Medicine]. 2012;47(3). [Chinese]<br>4. Long Y, Long Q, Tao X. [Effect of "Mediterranean diets" in patients with type 2 diabetes]. Qiqihar Yi Xue Yuan Xue Bao [Journal of Qiqihar University of Medicine]. 2015;14. [Chinese]<br>5. Wang Y, Song Y, Kang W, Song D. [Benefit assessment of low glycemic index diets in elderly diabetic patients]. Zhongguo Lao Nian Xue Za Zhi [Chinese Journal of Gerontology]. 2013;33(16). [Chinese] | data mistake or concerns on data authenticity |

#### File S5: Fundings and conflicts of interest of included studies

| Study | Fundings | Conflicts of Interest | Reg. No. |
|---|---|---|---|
| Al-Jazzaf 2007 | NR | NR | NR |
| Azadbakht 2011 | NR | N | NCT01049321 |
| Bahado-Singh 2015 | NR | NR | NR |
| Barnard 2006 | grant R01 DK059362-01A2 from the National Institute of Diabetes and Digestive and Kidney Diseases and by the Diabetes Action Research and Education Foundation | NR | NCT00276939 |
| Barnard 2009 | grant R01 DK059362-01A2 from the National Institute of Diabetes and Digestive and Kidney Diseases and by the Diabetes Action Research and Education Foundation | N | NCT00276939 |
| Brand 1991 | the Sydney University Nutrition Research Foundation, CSL-Novo Pty., Ltd., and the Apex-Australian Diabetes Foundation | NR | NR |
| Brehm 2009 | supported by the American Diabetes Association, U.S. Public Health Service (PHS) Grant DK57900, and the Cincinnati Children’s Hospital Medical Center Clinical Research Center (supported by U.S. PHS General Clinical Research Grant M01 RR 08084, General Clinical Research Centers Program, National Center for Research Resources, National Institutes of Health) | N | NR |
| Breukelman 2019 | N | N | NR |
| Breukelman 2021 | N | N | NR |
| Brinkworth 2004 | NR | N | NR |
| Brunerova 2007 | VZ MSM 0021620814 | N | NR |
| Cao 2011 | NR | NR | NR |
| Ceriello 2014 | NR | N | NR |
| Chandalia 2000 | grants (M01-RR00633 and HL-29252) from the National Institutes of Health and by research grants from the Bundesministerium für Bildung, Forschung, Wissenschaft und Technologie (01EC9402) and the Deutsche Forschungsgemeinschaft (BE 1673/1-1) | NR | NR |
| Chen 2020a | NR | NR | NCT03176056 |
| Chen 2020b | N | N | ChiCTR1900024880 |

| Study | Fundings | Conflicts of Interest | Reg. No. |
|---|---|---|---|
| Choi 2013 | Brain Korea 21 Project of the Ministry of Education and Human Resources Development, Republic of Korea (A102065-10111070100) | N | NR |
| Coppell 2010 | the Health Research Council of New Zealand (06/352) and the Southern Trust, New Zealand | N | NCT00124553 |
| Coulston 1989 | NIH Research Grants RR7022 and HL-08506 and the Nora Eccles Treadwell Foundation | NR | NR |
| Daly 2006 | Diabetes UK | Mark Daly has received research funding from the Sugar Bureau. He has worked on grants funded by National Starch and the erstwhile Ministry of Agriculture, Food and Fisheries; in addition he has received speaker fees from the Sugar Bureau | NR |
| Davis 2009 | the Robert C. Atkins Foundation and the Diabetes Research and Training Center (P60 DK020541) and by Clinical and Translational Science Award UL1 RR025750 | N | NCT00795691 |
| Ding 2010a | Key Projects of Scientific Research Program for Universities in Xinjiang Uygur Autonomous Region No.XJEDU2006I32 | NR | N |
| Ding 2010b | Key Projects of Scientific Research Program for Universities in Xinjiang Uygur Autonomous Region No.XJEDU2006I32 | NR | N |
| Durrer 2021 | Peer-reviewed funding was obtained from the Mitacs Accelerate program (Grant No. IT08605). Matching funds for the Mitacs Accelerate fellowship to C.D. were provided by industry partner Pharmasave Drugs (Pacific) Ltd. Further funding support was provided through salary support to J.P.L. from the Canadian Institutes for Health Research (MSH-141980) and the Michael Smith Foundation for Health Research (Scholar Award #16890). Food products were provided in-kind to pharmacies by Ideal Protein. | J.P.L. holds founder shares and advises for Metabolic Insights Inc., and is volunteer Chief ScientificOfficer for the not-for-profit Institute for Personalized Therapeutic Nutrition. S.M. is employed as Chief Executive Officer for the not-for-profit Institute for Personalized Therapeutic Nutrition. J.W. is a member of the Scientific Advisory Board, and has received travel support and speaker’s honoraria, from Atkins Nutritionals Inc. J.D.J. is Chair of the Board for the Institute for Personalized Therapeutic Nutrition and receives no compensation. C.D., J.S., A.M.B., and K.G. have nothing to declare. | NCT03181165 |
| Elhayany 2010 | NR | NR | NCT00520182 |
| Esposito 2009 | In part by the Second University of Naples | N | NCT00725257 |

| Study | Fundings | Conflicts of Interest | Reg. No. |
|---|---|---|---|
| Fabricatore 2011 | grant K23DK070777 from the National Institute of Diabetes and Digestive and Kidney Diseases (NIDDK) to Dr. Fabricatore. In addition, this project was supported by grant K24DK065018 from NIDDK to Dr. Wadden, by grant K24DK082730 from NIDDK to Dr. Ludwig, ... | Dr. Fabricatore has received research funding from Merck,Dr. Fabricatore is currently employed by Nutrisystem, Inc. Dr. Wadden has received research support from Novo Nordisk | NCT00729196 |
| Fan 2010 | Project of Science Development Plan of Datong Science and Technology Bureau, Tongkeifa [2004] No. 70, Tongcaijiaozi [2004] No. 65 | NR | NR |
| Fan 2013 | NR | NR | NR |
| Fang 2016 | NR | NR | NR |
| Fang 2019 | NR | NR | NR |
| Gannon 2003 | grants from the American Diabetes Association, the Minnesota Beef Council, and the Colorado and Nebraska Beef Councils and by Merit Review Funds from the Medical Research Service, Department of Veterans Affairs, Minneapolis | MCG, FQN, and HH are full-time employees of the Department of Veterans Affairs, Minneapolis. MCG and FQN are members of the American Diabetes Association Professional Society | NR |
| Gannon 2004 | he American Diabetes Association, the Minnesota Beef Council, and the Colorado and Nebraska Beef Councils | NR | NR |
| Goldstein 2011 | N | N | NCT00552890 |
| Gram-Kampmann 2022 | AP Møller Foundation; Danish Diabetes Academy funded by the Novo Nordisk Foundation; Novo Nordisk Fonden; Odense Universitetshospital; Overlæge Johan Boserup og Lise Boserups Legat; Region of Southern Denmark; Syddansk Universitet | N | NCT03068078 |
| Guldbrand 2012 | NR | N | NCT01005498 |
| Guo 2014 | NR | NR | NR |
| Han 2021 | the National Natural Science Foundation of China (82070799) | NR | NR |
| Hashemi 2019 | NR | N | IRCT20170214325 71N1 |
| He 2017 | Natural Science Foundation of Xinjiang Uygur Autonomous Region (2015211C061) | NR | NR |
| Heilbronn 2002 | NR | NR | NR |
| Hockaday 1978 | NR | NR | NR |
| Hu 2018 | Scientific Research Project of Shanghai Yangpu District Science and Technology Commission and Health and Family Planning Commission (YP15Q10) | NR | NR |

| Study | Fundings | Conflicts of Interest | Reg. No. |
|---|---|---|---|
| Huang 2016 | Nantong Health Bureau Youth (China) Fund (WQ2014033) | NR | NR |
| Ikem 2007 | NR | NR | NR |
| Iqbal 2010 | VA Merit Review Entry program | N | NCT00108459 |
| Itsiopoulos 2011 | grants from the NHMRC in Australia, Diabetes Australia, and Diabetes CCRE | NR | ACTRN012607000394448 |
| Jenkins 2008 | the Canadian Institutes of Health Research, Canada Research Chair Endowment of the Federal Government of Canada, and Barilla (Italy) | Dr Jenkins reported serving on the Scientific Advisory Board of Unilever, the Sanitarium Company, and the California Strawberry Commission; receiving honoraria for scientific advice from the Almond Board of California, Barilla, and Unilever Canada; being on the speaker's panel for the Almond Board of California; and receiving research grants from Loblaws, Unilever, Barilla, and the Almond Board of California. His wife is a director of Glycemic Index Laboratories, Toronto, Ontario, Canada. Dr Kendall reported having been on the speaker's panel for the Almond Board of California and receiving partial salary funding from research grants provided by Unilever, Loblaws, and the Almond Board of California. Mr Vidgen reported receiving partial salary funding from research grants provided by Unilever, Loblaws, and the Almond Board of California. No other authors reported any financial disclosures. | NCT00438698 |
| Jimenez-Cruz 2003 | Omnilife-Conacyt | NR | NR |
| Jönsson 2009 | Crafoordska stiftelsen, Region Skåne and Lund University | N | 00435240 |
| Kahleova 2011 | grant IGA MZCR NS ⁄10534-3 from Ministry of Health, Prague, Czech Republic | N | NCT00883038 |
| Krebs 2012 | The Health Research Council of New Zealand (06/337) | N | ACTRN12606000490572 |
| Lasa 2014 | Spanish Ministry of Health (PI1001407, AGL2009130906-C02-02, AGL2010-22319-C03-02, G03/140, RD06/0045), by Carlos III Health Institute (PREDIMED; CIBERobn), by Public Health | N | ISRCTN35739639 |

| Study | Fundings | Conflicts of Interest | Reg. No. |
|---|---|---|---|
| | Division of the Department of Health of the Autonomous Government of Catalonia i... | | |
| Lee 2016 | Korea Health Industry Development Institute, funded by the Ministry of Health & Welfare (A111716-12020000100), as well as the Korean Health Technology R&D Project, funded by the Ministry of Health and Welfare, Republic of Korea (HI13C0715 and HI11C1300) | NR | CRiS KCT0001771 |
| Li 2011 | NR | NR | NR |
| Li 2021 | NR | NR | NR |
| Li 2022 | Putian Science and Technology Bureau, Fujian province, China | N | NR |
| Liu 2011 | NR | NR | NR |
| Liu 2016 | NR | NR | NR |
| Liu 2020 | Key Project of Medical Science Research in Hebei Province (20191737); Project of Science and Technology Research and Development Program of Shijiazhuang City, Hebei Province (181460923) | NR | NR |
| Lousley 1984 | British Diabetic Association and the Simon Broome Heart Research Trust for financial support | NR | NR |
| Luger 2013 | NR | N | NR |
| Ma 2008 | grant 5 P30 DK032520 from the National Institute of Diabetes and Digestive and Kidney Diseases | NR | NCT00473811 |
| Marco-Benedí 2020 | CIBERCV (co-supported by the European Regional Development Fund (ERDF) which is allocated by the European Union; IIS16/0114), PI13/02507 and PI15/01983; and a grant from INTEROVIC | N | NCT02559479 |
| McLaughlin 2007 | National Institutes of Health Grants RR2HLL406 and RR 000070 | NR | NCT00168459 |
| Mehling 2000 | the Canadian Diabetes Association and Olive Oil Council of Canada | NR | NR |
| Mohammadi 2017 | NR | NR | IRCT2014011416223N1 |
| Mollentze 2019 | an unrestricted educational grant by Mr. Christo Strydom, Bloemfontein, South Africa | Author WF Mollentze was the manager of the Christo Strydom Metabolic Research Unit, University of the Free State, at the time research was conducted. The University of the Free State | NR |

| Study | Fundings | Conflicts of Interest | Reg. No. |
|---|---|---|---|
| | | received partial funding for the Christo Strydom Metabolic Research Unit from CSN, the company that manufactures and distributes CSN products in South Africa. Author WFMollentze received a speaker honorarium from Novartis within the last 3 years. Author G Joubert declares that she has no conflict of interest. Author S van der Linde declares that she has no conflict ofinterest. Author A Prins declares she has no conflict of interest. Author GM Marx declares she has no conflict of interest. Author KG Tsie declares that she has no conflict of interest. | |
| Nicholson 1999 | NR | NR | NR |
| Ning 2020 | NR | NR | NR |
| Parker 2002 | Meadow Lea Foods | NR | NR |
| Pavithran 2020a | N | N | CTRI/2019/12/022 425 |
| Pavithran 2020b | N | N | CTRI/2019/12/022 425 |
| Pedersen 2014 | NR | PMC is the co-author of The CSIRO Total Wellbeing Diet Book | ACTRN126080000 45314 |
| Perna 2019 | Deanship of Scientific Research, University of Bahrain (Project No. 19/2011). | N | NR |
| Rizkalla 2004 | grants from INSERM, from Pierre and Marie Curie University, from Danone Vitapole, from Nestle France, from the Association Benjamin Delessert, and from the Association of Young Diabetic Individuals, France. | NR | NR |
| Rock 2014 | unding was provided through a clinical trial contract to the coordinating center (School of Medicine, UCSD). Jenny Craig, Inc. (Carlsbad, CA) | This study was supported by Jenny Craig, Inc. By contractual agreement, scientists at UCSD and the University of Minnesota have responsibility and independence regarding data management, analysis, and publication. The sponsor contributed to the development of the design and protocol through discussions with the investigators during | NCT01345500 |

| Study | Fundings | Conflicts of Interest | Reg. No. |
|---|---|---|---|
| | | the development phase of the study. The funding sponsor had no role in the conduct of the study; collection, management, analysis, and interpretation of the data; preparation, review, or approval of the manuscript (except for verifying the specific weight loss program activities that comprised the intervention); and decision to submit the manuscript for publication. No other potential conflicts of interest relevant to this article were reported. | |
| Ruggenenti 2017 | grants from the Istituto Superiore di Sanita/National Institutes of Health Collaborative Projects of the Italian Ministry of Health, the Bakewell Foundation, and the Longer Life Foundation | N | NCT01213212 |
| Ruggenenti 2022 | grant of the Italian Ministry of Health (Project Code: RF-2010-2309734) and was partially internally funded by the Istituto di Ricerche Farmacologiche Mario Negri IRCCS, Bergamo, Italy | N | NCT01930136 |
| Saslow 2014 | The research was supported by a grant from the William K. Bowes, Jr. Foundation. Laura Saslow was supported by NIH grant T32AT003997 from the National Center for Complementary and Alternative Medicine (NCCAM). Judith Moskowitz was supported by NIH grant K24 MH093225 from the National Institute of Mental Health. Frederick Hecht was supported by NIH grant K24 AT007827 from NCCAM. The funders had no role in study design, data collection and analysis, decision to publish, or preparation of the manuscript. | N | NCT01713764 |
| Saslow 2017 | The research was supported by a grant from the William K. Bowes, Jr. Foundation and the Mount Zion Health Fund. Laura Saslow and Ashley E. Mason were supported by National Institutes of Health grant T32AT003997 from the National Center for Complementary and Integrative Health. Laura Saslow was also supported by a K01 from the National Institute of Diabetes and Digestive and Kidney Diseases (DK107456). Ashley E. Mason was also supported by a K23 from the National Heart, Lung, and Blood Institute (HL133442). Judith Moskowitz was | Stephen Phinney is a paid member of the Atkins Scientific Advisory Board, a founder of Virta Health, and has authored books on low-carbohydrate, high fat diets: New Atkins and You, The Art and Science of Low Carbohydrate Living, and The Art and Science of Low Carbohydrate Performance. Frederick Hecht is on the Scientific Advisory Board for Virta Health. The other authors declare no competing financial interests. | NCT01713764 |

| Study | Fundings | Conflicts of Interest | Reg. No. |
|---|---|---|---|
| | supported by National Institutes of Health grant K24 MH093225 from the National Institute of Mental Health. Frederick Hecht was supported by National Institutes of Health grant K24 AT007827 from National Center for Complementary and Integrative Health. | | |
| Sato 2017 | Mishima Kaiun Memorial Foundation | JS has received lecture fees from Novartis Pharmaceuticals, Novo Nordisk Pharma, Sanofi, and Takeda Pharmaceutical Co. AK has received lecture fees from Kissei Pharma, Sanofi and Takeda Pharmaceutical Co. YT has received lecture fees from Takeda Pharmaceutical Co., MSD, Eli Lilly, Kissei Pharma and AstraZeneca. TM has received lecture fees from MSD, Takeda Pharmaceutical Co., and Eli Lilly. YF has received lecture fees from Novartis Pharmaceuticals and Eli Lilly, research funds from Novartis Pharmaceuticals, MSD and Takeda Pharmaceutical Co. HW has received lecture fees from Asteras, Astrazeneca, Boehringer Ingelheim, Daiichi Sankyo Inc., Eli Lilly and Company, Kissei Pharmaceutical Co., Kowa Pharmaceutical CO., Kyowa Hakko Kirin Co., MSD, Novartis Pharmaceuticals, Novo Nordisk Pharma, Ono Pharmaceutical Co., Mitsubishi Tanabe Pharma, Sanofi-Aventis, Sanwakagaku Kenkyusho, and Takeda Pharmaceutical Co. and research funds from Asteras, Astrazeneca, Bristol-Myers Squibb, Boehringer Ingelheim, Daiichi Sankyo Inc., Dainippon Sumitomo Pharma, Eli Lilly, Johnson and Johnson, Kissei Pharmaceutical Co., Kowa Pharmaceutical Co., Kyowa Hakko Kirin Co. MSD, Mitsubishi Tanabe Pharma, Mochida Pharmaceutical Co., Novartis Pharmaceuticals, Novo Nordisk Pharma, Pfizer, Sanwakagaku | UMIN000010663 |

| Study | Fundings | Conflicts of Interest | Reg. No. |
|---|---|---|---|
| | | Kenkyusho, Sanofi, and Takeda Pharmaceutical Co. All the other authors report no conflict of interest. | |
| Shen 2021 | Zhejiang Provincial Medical and Health Science and Technology Plan Project (2020ZH079) | NR | NR |
| Shige 2000 | Grains Research & Development and Meadow Lea Foods (Sydney, Australia) provided the specially prepared foods | NR | NR |
| Skytte 2019 | The study was funded by grants from: Arla Food for Health; the Novo Nordisk Foundation Center for Basic Metabolic Research, University of Copenhagen; the Department of Clinical Medicine, Aarhus University; the Department of Nutrition, Exercise and Sports, University of Copenhagen; and Copenhagen University Hospital, Bispebjerg. Dietary ingredients were partly provided by Arla Foods, JAN Import A/S, Royal Greenland and Danish Crown. The study sponsors, Arla Food for Health, Arla Foods, JAN Import A/S, Royal Greenland and Danish Crown, were informed, but not involved in the design of the study; the collection, analysis, and interpretation of data; writing the report; or the decision to submit the report for publication. Coauthors from the University ofCopenhagen and Aarhus University were involved in the study, as detailed in the contribution statement. | AA is a member of advisory boards/consultant for: BioCare Copenhagen, Denmark; the Dutch Beer Institute, the Netherlands; Gelesis, USA; Groupe Éthique et Santé, France; McCain Foods Limited, USA; Navamedic, Denmark; Novo Nordisk, Denmark; Pfizer, USA; Saniona, Denmark; Weight Watchers, USA; and is a recipient of travel grants and honoraria as speaker for a wide range ofDanish and international concerns. AA is co-owner and a member ofthe board of the consultancy company Dentacom ApS, Denmark; co-founder and coowner of UCPH spin-outs Mobile Fitness A/S, Flaxslim ApS and Personalized Weight Management Research Consortium ApS (Glucodiet.dk). AA is co-inventor of a number of patents owned by UCPH, in accordance with Danish law. AA is co-author of a number of diet and cookery books, including books on personalised diet. AA is not an advocate or activist for specific diets, and is not strongly committed to any specific diet, e.g. veganism, Atkins diet, gluten-free diet, high-animal protein diet or dietary supplements. TML is an advisor for 'Sense' diet programme. None of the other authors have conflicts of interest to declare. | NCT02764021 |
| Stentz 2016 | the American Diabetes Association (7-12-CT-41) and the AD Baskin Research Fund (PIs FBS and AEK). | N | NCT0164284 |
| Sun 2007 | National Natural Science Foundation of China (30371221), | NR | NR |

| Study | Fundings | Conflicts of Interest | Reg. No. |
|---|---|---|---|
| | Shanghai Science and Technology Commission Fund (034119855) | | |
| Sun 2020 | NR | NR | NR |
| Tang 2021 | NR | NR | NR |
| Tay 2015 | the National Health and Medical Research Council of Australia (project grant 103415) and the Agency for Science, Technology and Research, Singapore (postgraduate research scholarship to JT | N | ACTRN126120003 69820 |
| Thomsen 2022 | grants from Arla Foods amba, The Danish Dairy Research Foundation, and Copenhagen University Hospital Bispebjerg Frederiksberg | AA is currently employed by The Novo Nordisk Foundation to establish a National Centre for Healthy Weight and is a member of the advisory board/consultant for Gelesis (USA), Groupe Éthique et Santé (France) and Weight Watchers (USA). AA is co-owner of the University of Copenhagen spin-off Flax-Slim ApS and is co-inventor on a pending provisional patent application for the use of biomarkers to predict responses to weight-loss diets and other related patents and patent applications that are all owned by the University of Copenhagen in accordance with Danish law. AA is co-author ofa number ofdiet and cookery books, including books on personalised diet. AA is not an advocate or activist for specific diets and is not strongly committed to any specific diet (e.g. veganism, Atkins diet, gluten-free diet, high animal protein diet or dietary supplements). TML is an advisor for the 'Sense' diet programme. The remaining authors declare that there are no relationships or activities that might bias, or be perceived to bias, their work. | NCT03814694 |
| Uusitupa 1993 | financially supported by the Medical Research Council of the Academy of Finland, the Emil Aaltonen Foundation, The North Savo Regional Fund of the Finnish Cultural Foundation,the Finnish Foundation Research and Nordisk Insulin of Diabetes Foundation,Denm... | N | NR |

| Study | Fundings | Conflicts of Interest | Reg. No. |
|---|---|---|---|
| Visek 2014 | grant of the Medical Faculty in Pilsen, Charles University in Prague, MSM 0021620814 | N | NR |
| Walker 1995 | grant from Diabetes Australia. We are grateful for products supplied by the International Olive Oil Council and Meadow Lea Foods Australia | NR | NR |
| Wang 2009a | NR | NR | NR |
| Wang 2009b | NR | NR | NR |
| Wang 2015 | NR | NR | NR |
| Wang 2018 | Suzhou Science and Technology Project, China (Grant number SYS201513) | N | NR |
| Watson 2016 | grant from the Pork Co-operative Research Centre (Pork CRC), an Australian Government funding initiative | NAW is supported by a post-graduate research scholarship from the Pork CRC | ACTRN12613000008729 |
| Westman 2008 | the Robert C. Atkins Foundation | N | NR |
| Wolever 1992 | grant from the Bristol Myers Company, New York. | NR | NR |
| Wolever 2008 | the Canadian Institutes of Health Research (CIHR-MCT44205) | NR | ISRCTN81151522 |
| Wu 2020 | NR | NR | N |
| Xue 2020 | NR | NR | NR |
| Yamada 2014 | NR | N | NR |
| Ye 2021 | NR | NR | NR |
| Yu 2020 | Henan University Intramural Fund (Project No. 2013YBZR028) | NR | NR |
| Zahedi 2021 | NR | N | IRCT2017011131875N1 |
| Zhao 2018 | NR | NR | NR |
| Zheng 2015 | Clinical Research Fund of Zhejiang Medical Association (2012ZYC-A101) | NR | NR |
| Zhou 2011 | NR | NR | NR |

N, not declared / no funding; NR, not reported.

### File S6: Risk of bias assessment

#### S6.1 Risk of bias graph

| Study ID | Reference | D1 | D2 | D3 | D4 | D5 | DS | Overall |
|---|---|---|---|---|---|---|---|---|
| 1 | Ding 2010 | - | ! | + | ! | ! | | - |
| 2 | Wu 2020 | ! | ! | + | ! | ! | | ! |
| 3 | He 2017 | ! | ! | + | + | ! | | ! |
| 4 | Yu 2020 | ! | ! | + | + | + | | ! |
| 5 | Liu 2016 | + | + | + | + | + | | + |
| 6 | Liu 2011 | - | ! | + | + | ! | | - |
| 7 | Liu 2020 | ! | ! | + | + | ! | | ! |
| 8 | Ye 2021 | ! | ! | + | + | ! | | ! |
| 9 | Zhou 2011 | ! | ! | + | + | ! | | ! |
| 10 | Sun 2020 | ! | ! | + | + | ! | | ! |
| 11 | Sun 2007 | ! | ! | + | + | ! | + | ! |
| 12 | Ning 2020 | ! | ! | + | + | ! | | ! |
| 13 | Fang 2019 | ! | ! | + | + | ! | | ! |
| 14 | Fang 2016 | ! | ! | + | + | ! | | ! |
| 15 | Cao 2011 | ! | ! | + | + | ! | | ! |
| 16 | Li 2021 | ! | ! | + | + | ! | | ! |
| 17 | Li 2011 | ! | ! | + | + | ! | | ! |
| 18 | Fan 2013 | ! | ! | + | + | ! | | ! |
| 19 | Tang 2021 | ! | ! | + | + | ! | | ! |
| 20 | Shen 2021 | ! | ! | + | + | ! | | ! |
| 21 | Wang 2009 | ! | ! | + | + | ! | | ! |
| 22 | Wang 2009b | ! | ! | + | + | ! | | ! |
| 23 | Wang 2015 | ! | ! | + | + | ! | | ! |
| 24 | Hu 2018 | ! | ! | + | + | ! | | ! |
| 25 | Fan 2010 | ! | ! | + | + | ! | | ! |
| 26 | Xue 2020 | ! | ! | + | + | ! | | ! |
| 27 | Zhao 2018 | ! | ! | + | + | ! | | ! |
| 28 | Zheng 2015 | ! | ! | + | + | ! | | ! |
| 29 | Guo 2014 | ! | ! | + | + | ! | | ! |
| 30 | Huang 2016 | ! | ! | + | + | ! | | ! |
| 31 | Al-Jazzaf 2007 | ! | ! | + | + | ! | | ! |
| 32 | Azadbakht 2011 | ! | - | - | + | ! | + | - |
| 33 | Bahado-Singh 2015 | ! | ! | + | + | + | | ! |
| 34 | Barnard 2006+9 | ! | + | + | + | + | | ! |
| 35 | Brand 1991 | ! | ! | + | + | ! | + | ! |
| 36 | Brehm 2009 | ! | + | + | + | ! | | ! |
| 37 | Breukelman 2019/21 | - | + | + | + | ! | | - |
| 38 | Brinkworth 2004 | - | ! | - | + | + | | - |
| 39 | Brunerova 2007 | - | ! | + | + | ! | | - |
| 40 | Ceriello 2014 | ! | ! | + | + | ! | | ! |
| 41 | Chandalia 2000 | ! | ! | + | + | + | - | - |
| 42 | Chen 2020a | ! | + | + | + | + | | ! |
| 43 | Chen 2020b | + | ! | ! | + | + | | ! |
| 44 | Choi 2013 | ! | ! | + | + | ! | | ! |
| 45 | Coppell 2010 | ! | + | + | + | ! | | ! |
| 46 | Coulston 1989 | ! | ! | + | + | ! | ! | ! |
| 47 | Daly 2006 | ! | ! | ! | + | + | | ! |
| 48 | Davis 2009 | ! | + | + | + | + | | ! |
| 49 | Durrer 2021 | + | + | + | + | + | | + |
| 50 | Elhayany 2010 | ! | ! | - | + | + | | - |
| 51 | Esposito 2009 | + | + | + | + | + | | + |
| 52 | Fabricatore 2011 | ! | ! | + | + | ! | | ! |
| 53 | Gannon 2004 | ! | ! | - | + | ! | ! | - |
| 54 | Gannon 2003 | ! | ! | + | + | ! | ! | ! |
| 55 | Goldstein 2011 | ! | + | + | + | + | | ! |
| 56 | Gram-Kampmann 2022 | ! | + | + | + | + | | ! |
| 57 | Guldbrand 2012 | ! | + | + | + | + | | ! |
| 58 | Han 2021 | + | ! | + | + | ! | | ! |
| 59 | Hashemi 2019 | ! | ! | + | + | ! | | ! |
| 60 | Heilbronn 2002 | ! | + | ! | + | ! | | ! |
| 61 | Hockaday 1978 | ! | ! | + | + | ! | | ! |
| 62 | Ikem 2007 | + | ! | + | + | ! | | ! |
| 63 | Iqbal 2010 | ! | + | + | + | ! | | ! |
| 64 | Itsiopoulos 2011 | ! | ! | ! | + | ! | + | ! |
| 65 | Jenkins 2008 | + | + | + | + | + | | + |
| 66 | Jimenez-Cruz 2003 | ! | ! | - | + | ! | ! | - |
| 67 | Jönsson 2009 | + | + | + | + | ! | + | ! |
| 68 | Kahleova 2011 | ! | + | + | + | + | | ! |
| 69 | Krebs 2012 | + | + | + | + | + | | + |
| 70 | Lasa 2014 | + | ! | + | + | + | | ! |
| 71 | Lee 2016 | ! | + | + | + | + | | ! |
| 72 | Li 2022 | ! | ! | + | + | ! | | ! |
| 73 | Lousley 1984 | ! | ! | ! | + | ! | ! | ! |
| 74 | Luger 2013 | ! | ! | + | + | + | | ! |
| 75 | Ma 2008 | ! | ! | ! | + | + | | ! |
| 76 | Marco-Benedí 2020 | + | + | + | + | + | | + |
| 77 | McLaughlin 2007 | ! | ! | + | + | ! | | ! |
| 78 | Mehling 2007 | ! | - | + | + | ! | | - |
| 79 | Mohammadi 2017 | ! | ! | + | + | + | | ! |
| 80 | Mollentze 2019 | ! | ! | - | + | + | | - |
| 81 | Nicholson 1999 | ! | ! | + | + | ! | | ! |
| 82 | Parker 2002 | ! | ! | ! | + | ! | | ! |
| 83 | Pavithran 2020a | ! | ! | + | + | ! | | ! |
| 84 | Pavithran 2020b | ! | ! | - | + | ! | | - |
| 85 | Pedersen 2014 | + | ! | ! | + | ! | | ! |
| 86 | Perna 2019 | ! | ! | ! | + | ! | | ! |

+ Low risk
! Some concerns
- High risk

D1 Randomisation process
D2 Deviations from the intended interventions
D3 Missing outcome data
D4 Measurement of the outcome
D5 Selection of the reported result
DS Bias arising from period and carryover effects

| Study ID | Reference | D1 | D2 | D3 | D4 | D5 | DS | Overall |
|---|---|---|---|---|---|---|---|---|
| 87 | Rizkalla 2004 | ! | ! | + | + | + | ! | ! |
| 88 | Rock 2014 | ! | + | + | + | + | | ! |
| 89 | Ruggenenti 2017 | + | + | + | + | + | | + |
| 90 | Ruggenenti 2022 | + | + | + | + | + | | + |
| 91 | Saslow 2014/2017 | + | + | + | + | + | | + |
| 92 | Sato 2017 | ! | ! | ! | + | + | | ! |
| 93 | Shige 2000 | ! | ! | + | + | ! | | ! |
| 94 | Skytte 2019 | ! | ! | + | + | + | - | - |
| 95 | Stentz 2016 | ! | ! | - | + | ! | | - |
| 96 | Tay 2015 | + | + | + | + | + | | + |
| 97 | Thomsen 2022 | + | + | + | + | + | | + |
| 98 | Uusitupa 1993 | ! | ! | + | + | + | | ! |
| 99 | Visek 2014 | ! | + | + | + | ! | - | - |
| 100 | Walker 1995 | ! | ! | + | + | + | ! | ! |
| 101 | Wang 2018 | + | + | + | + | + | | + |
| 102 | Watson 2016 | + | + | + | + | + | | + |
| 103 | Westman 2008 | ! | ! | - | + | ! | | - |
| 104 | Wolever 1992 | ! | ! | + | + | ! | ! | ! |
| 105 | Wolever 2008 | + | + | ! | + | + | | ! |
| 106 | Yamada 2014 | ! | ! | + | + | ! | | ! |
| 107 | Zahedi 2021 | ! | ! | ! | + | ! | | ! |

#### S6.2 Detailed ratings of each item

**Table S6.1** Risk of bias assessment of parallel randomized controlled trials

| Study ID | Reference | Randomization process | | | | Deviations from intended interventions | | | | | | | | Missing outcome data | | | | | Measurement of the outcome | | | | | | Selection of the reported result | | | | Overall |
|---|---|---|---|---|---|---|---|---|---|---|---|---|---|---|---|---|---|---|---|---|---|---|---|---|---|---|---|---|---|
| | | 1.1 | 1.2 | 1.3 | RoB 1 | 2.1 | 2.2 | 2.3 | 2.4 | 2.5 | 2.6 | 2.7 | RoB 2 | 3.1 | 3.2 | 3.3 | 3.4 | RoB 3 | 4.1 | 4.2 | 4.3 | 4.4 | 4.5 | RoB 4 | 5.1 | 5.2 | 5.3 | RoB 5 | |
| 1 | Ding 2010 | NI | N | N | H | Y | Y | PY | PN | NA | N | PN | SC | PY | NA | NA | NA | L | N | N | NI | PY | PN | SC | NI | PN | N | SC | H |
| 2 | Wu 2020 | PY | NI | N | SC | Y | Y | NI | NA | NA | N | PN | SC | Y | NA | NA | NA | L | N | N | NI | NI | PN | SC | NI | N | N | SC | SC |
| 3 | He 2017 | PY | NI | N | SC | Y | Y | PN | NA | NA | N | N | SC | Y | NA | NA | NA | L | PN | N | NI | PN | NA | L | NI | PN | N | SC | SC |
| 4 | Yu 2020 | PY | NI | PN | SC | Y | PY | PN | NA | NA | N | PN | SC | Y | NA | NA | NA | L | PN | N | NI | N | NA | L | NI | N | N | L | SC |
| 5 | Liu 2016 | Y | Y | N | L | PN | PN | NA | NA | NA | PN | N | L | PN | PY | NA | NA | L | N | N | PN | NA | NA | L | PY | PN | N | L | L |
| 6 | Liu 2011 | NI | NI | PY | H | NI | NI | PN | NA | NA | N | PN | SC | Y | NA | NA | NA | L | PN | N | PY | PN | NA | L | NI | N | N | SC | H |
| 7 | Liu 2020 | PY | NI | PN | SC | NI | PY | NI | NA | NA | N | PN | SC | Y | NA | NA | NA | L | N | N | NI | PN | NA | L | NI | N | N | SC | SC |
| 8 | Ye 2021 | NI | NI | PN | SC | NI | NI | PN | NA | NA | N | PN | SC | Y | NA | NA | NA | L | PN | N | NI | PN | NA | L | NI | N | N | SC | SC |
| 9 | Zhou 2011 | Y | NI | N | SC | NI | NI | PN | NA | NA | N | N | SC | PN | N | PN | NA | L | PN | N | NI | PN | NA | L | NI | N | N | SC | SC |
| 10 | Sun 2020 | NI | NI | PN | SC | NI | NI | PN | NA | NA | N | PN | SC | Y | NA | NA | NA | L | PN | N | NI | PN | NA | L | NI | N | PN | SC | SC |
| 12 | Ning 2020 | PY | NI | PN | SC | NI | NI | PN | NA | NA | N | PN | SC | Y | NA | NA | NA | L | N | N | NI | PN | NA | L | NI | N | N | SC | SC |

| Study ID | Reference | Randomization process | | | | Deviations from intended interventions | | | | | | | | Missing outcome data | | | | | Measurement of the outcome | | | | | | Selection of the reported result | | | | Overall |
|---|---|---|---|---|---|---|---|---|---|---|---|---|---|---|---|---|---|---|---|---|---|---|---|---|---|---|---|---|---|
| | | 1.1 | 1.2 | 1.3 | RoB 1 | 2.1 | 2.2 | 2.3 | 2.4 | 2.5 | 2.6 | 2.7 | RoB 2 | 3.1 | 3.2 | 3.3 | 3.4 | RoB 3 | 4.1 | 4.2 | 4.3 | 4.4 | 4.5 | RoB 4 | 5.1 | 5.2 | 5.3 | RoB 5 | |
| 13 | Fang 2019 | NI | NI | PN | SC | NI | NI | PN | NA | NA | N | PN | SC | Y | NA | NA | NA | L | N | N | NI | PN | NA | L | NI | N | N | SC | SC |
| 14 | Fang 2016 | NI | NI | PN | SC | NI | NI | PN | NA | NA | N | N | SC | Y | NA | NA | NA | L | N | N | NI | PN | NA | L | NI | N | N | SC | SC |
| 15 | Cao 2011 | NI | NI | N | SC | NI | NI | N | NA | NA | N | N | SC | Y | NA | NA | NA | L | N | N | NI | N | NA | L | NI | N | N | SC | SC |
| 16 | Li 2021 | PY | NI | N | SC | NI | NI | N | NA | NA | N | PN | SC | Y | NA | NA | NA | L | N | N | NI | PN | NA | L | NI | N | N | SC | SC |
| 17 | Li 2011 | PN | NI | N | SC | NI | NI | PN | NA | NA | N | PN | SC | Y | NA | NA | NA | L | N | N | NI | N | NA | L | NI | N | N | SC | SC |
| 18 | Fan 2013 | Y | NI | N | SC | NI | NI | N | NA | NA | N | PN | SC | Y | NA | NA | NA | L | N | N | NI | PN | NA | L | NI | N | N | SC | SC |
| 19 | Tang 2021 | Y | NI | PN | SC | NI | NI | PN | NA | NA | NI | PN | SC | Y | NA | NA | NA | L | N | N | NI | N | NA | L | NI | N | N | SC | SC |
| 20 | Shen 2021 | Y | NI | PN | SC | NI | NI | PN | NA | NA | NI | PN | SC | Y | NA | NA | NA | L | N | N | NI | N | NA | L | NI | N | N | SC | SC |
| 21 | Wang 2009 | PN | NI | PN | SC | NI | NI | N | NA | NA | NI | PN | SC | Y | NA | NA | NA | L | N | N | NI | N | NA | L | NI | N | N | SC | SC |
| 22 | Wang 2009b | PN | NI | N | SC | NI | NI | PN | NA | NA | NI | N | SC | Y | NA | NA | NA | L | N | N | NI | N | NA | L | NI | N | N | SC | SC |
| 23 | Wang 2015 | PN | NI | PN | SC | NI | NI | N | NA | NA | NI | PN | SC | Y | NA | NA | NA | L | N | N | NI | N | NA | L | NI | N | N | SC | SC |
| 24 | Hu 2018 | Y | NI | PN | SC | NI | NI | PN | NA | NA | NI | N | SC | PN | PN | PN | NA | L | N | N | NI | N | NA | L | NI | N | N | SC | SC |
| 25 | Fan 2010 | NI | NI | N | SC | NI | NI | N | NA | NA | NI | N | SC | Y | NA | NA | NA | L | N | N | NI | N | NA | L | NI | N | N | SC | SC |
| 26 | Xue 2020 | NI | NI | PN | SC | NI | NI | N | NA | NA | NI | N | SC | Y | NA | NA | NA | L | N | N | NI | N | NA | L | NI | N | N | SC | SC |
| 27 | Zhao 2018 | NI | NI | PN | SC | NI | NI | PN | NA | NA | NI | PN | SC | Y | NA | NA | NA | L | N | N | NI | N | NA | L | NI | N | N | SC | SC |
| 28 | Zheng 2015 | Y | NI | N | SC | NI | NI | N | NA | NA | NI | N | SC | Y | NA | NA | NA | L | N | N | NI | N | NA | L | NI | N | N | SC | SC |
| 29 | Guo 2014 | PN | NI | N | SC | NI | NI | N | NA | NA | NI | PN | SC | Y | NA | NA | NA | L | N | N | NI | N | NA | L | NI | N | N | SC | SC |
| 30 | Huang 2016 | PN | NI | PN | SC | NI | NI | N | NA | NA | NI | PN | SC | Y | NA | NA | NA | L | PN | N | NI | N | NA | L | NI | PN | PN | SC | SC |
| 31 | Al-Jazzaf 2007 | PN | NI | PN | SC | NI | NI | N | NA | NA | N | N | SC | PN | PY | NA | NA | L | N | N | NI | PN | NA | L | NI | N | N | SC | SC |
| 33 | Bahado-Singh 2015 | PN | NI | N | SC | PY | NI | N | NA | NA | NI | N | SC | Y | NA | NA | NA | L | N | N | NI | N | NA | L | PY | N | N | L | SC |
| 34 | Barnard 2006+9 | Y | NI | N | SC | NI | NI | N | NA | NA | Y | NA | L | PN | PN | PN | NA | L | N | N | NI | N | NA | L | Y | N | N | L | SC |
| 36 | Brehm 2009 | PN | NI | PN | SC | NI | NI | N | NA | NA | Y | NA | L | N | PY | NA | NA | L | N | N | NI | N | NA | L | NI | N | N | SC | SC |
| 37 | Breukelman 2019/21 | PN | NI | PY | H | NI | NI | PN | NA | NA | Y | NA | L | PN | PN | PN | NA | L | N | N | NI | N | NA | L | NI | N | N | SC | H |
| 38 | Brinkworth 2004 | Y | PN | PY | H | NI | NI | PN | NA | NA | NI | N | SC | N | PN | NI | NI | H | N | N | NI | N | NA | L | PY | N | N | L | H |
| 39 | Brunerova 2007 | PN | N | PN | H | Y | Y | N | NA | NA | NI | N | SC | NI | N | PN | NA | L | N | N | Y | N | NA | L | NI | N | N | SC | H |
| 40 | Ceriello 2014 | Y | NI | N | SC | NI | NI | N | NA | NA | N | N | SC | Y | NA | NA | NA | L | N | N | NI | N | NA | L | NI | N | N | SC | SC |
| 42 | Chen 2020a | Y | NI | N | SC | Y | Y | N | NA | NA | Y | NA | L | PN | Y | NA | NA | L | N | N | Y | N | NA | L | Y | N | N | L | SC |

| Study ID | Reference | Randomization process | | | | Deviations from intended interventions | | | | | | | | Missing outcome data | | | | | Measurement of the outcome | | | | | | Selection of the reported result | | | | Overall |
|---|---|---|---|---|---|---|---|---|---|---|---|---|---|---|---|---|---|---|---|---|---|---|---|---|---|---|---|---|---|
| | | 1.1 | 1.2 | 1.3 | RoB 1 | 2.1 | 2.2 | 2.3 | 2.4 | 2.5 | 2.6 | 2.7 | RoB 2 | 3.1 | 3.2 | 3.3 | 3.4 | RoB 3 | 4.1 | 4.2 | 4.3 | 4.4 | 4.5 | RoB 4 | 5.1 | 5.2 | 5.3 | RoB 5 | |
| 43 | Chen 2020b | Y | PY | N | L | Y | Y | N | NA | NA | N | N | SC | N | PN | PY | N | SC | N | N | Y | N | NA | L | Y | N | N | L | SC |
| 44 | Choi 2013 | PY | NI | N | SC | NI | NI | PN | NA | NA | NI | PN | SC | Y | NA | NA | NA | L | N | N | N | NA | NA | L | NI | N | N | SC | SC |
| 45 | Coppell 2010 | Y | NI | PN | SC | NI | NI | PN | NA | NA | Y | NA | L | PN | PY | NA | NA | L | N | N | N | NA | NA | L | NI | N | N | SC | SC |
| 47 | Daly 2006 | PY | NI | N | SC | Y | Y | N | NA | NA | N | PN | SC | PN | PN | NI | PN | SC | N | N | NI | N | NA | L | Y | N | N | L | SC |
| 48 | Davis 2009 | PY | NI | N | SC | Y | Y | N | NA | NA | Y | NA | L | Y | NA | NA | NA | L | N | N | Y | N | NA | L | PY | N | N | L | SC |
| 49 | Durrer 2021 | Y | PY | PN | L | Y | Y | N | NA | NA | Y | NA | L | N | Y | NA | NA | L | N | N | Y | N | NA | L | Y | N | N | L | L |
| 50 | Elhayany 2010 | PY | NI | N | SC | NI | NI | PN | NA | NA | PN | N | SC | N | PN | PY | NI | H | N | N | NI | N | NA | L | PY | N | N | L | H |
| 51 | Esposito 2009 | Y | Y | N | L | Y | Y | N | NA | NA | Y | NA | L | PY | NA | NA | NA | L | N | N | N | NA | NA | L | Y | N | N | L | L |
| 52 | Fabricatore 2011 | PY | NI | PN | SC | Y | Y | PN | NA | NA | NI | PN | SC | PN | N | PN | NA | L | N | N | NI | N | NA | L | NI | N | N | SC | SC |
| 55 | Goldstein 2011 | PY | NI | PN | SC | N | NI | PN | NA | NA | Y | NA | L | PY | NA | NA | NA | L | N | PN | NI | N | NA | L | Y | N | N | L | SC |
| 56 | Gram-Kampmann 2022 | PY | NI | PN | SC | Y | Y | PN | NA | NA | Y | NA | L | PN | PY | NA | NA | L | N | N | NI | N | NA | L | Y | N | N | L | SC |
| 57 | Guldbrand 2012 | Y | NI | PN | SC | NI | NI | PN | NA | NA | Y | NA | L | PN | N | PN | NA | L | N | N | NI | N | NA | L | PY | N | N | L | SC |
| 58 | Han 2021 | Y | PY | PN | L | Y | Y | PN | NA | NA | N | PN | SC | PY | NA | NA | NA | L | N | N | NI | PN | NA | L | PN | PN | N | SC | SC |
| 59 | Hashemi 2019 | PY | NI | PN | SC | NI | NI | PN | NA | NA | PN | PN | SC | Y | NA | NA | NA | L | N | N | NI | N | NA | L | NI | N | N | SC | SC |
| 60 | Heilbronn 2002 | NI | NI | PN | SC | NI | NI | PN | NA | NA | PY | NA | L | PN | PN | NI | PN | SC | N | N | NI | PN | NA | L | N | N | N | SC | SC |
| 61 | Hockaday 1978 | PY | NI | N | SC | NI | NI | PN | NA | NA | NI | PN | SC | Y | NA | NA | NA | L | N | N | NI | N | NA | L | NI | PN | N | SC | SC |
| 62 | Ikem 2007 | Y | PY | PN | L | NI | NI | PN | NA | NA | NI | PN | SC | Y | NA | NA | NA | L | N | N | NI | N | NA | L | NI | N | N | SC | SC |
| 63 | Iqbal 2010 | PY | NI | PN | SC | NI | NI | PN | NA | NA | Y | NA | L | N | PN | PN | NA | L | N | N | NI | N | NA | L | NI | N | N | SC | SC |
| 65 | Jenkins 2008 | PY | PY | PN | L | Y | Y | PN | NA | NA | Y | NA | L | N | PY | NA | NA | L | N | N | N | NA | NA | L | Y | N | N | L | L |
| 68 | Kahleova 2011 | PY | NI | N | SC | Y | Y | N | NA | NA | Y | NA | L | PN | N | PN | NA | L | N | N | Y | N | NA | L | PY | N | N | L | SC |
| 69 | Krebs 2012 | Y | Y | N | L | Y | Y | N | NA | NA | Y | NA | L | N | Y | NA | NA | L | N | N | N | NA | NA | L | Y | N | N | L | L |
| 70 | Lasa 2014 | Y | PY | N | L | Y | NI | PN | NA | NA | NI | PN | SC | NI | PY | NA | NA | L | N | N | Y | N | NA | L | PY | N | N | L | SC |
| 71 | Lee 2016 | Y | NI | N | SC | Y | Y | PN | NA | NA | Y | NA | L | N | PY | NA | NA | L | N | N | NI | N | NA | L | Y | N | N | L | SC |
| 72 | Li 2022 | PY | NI | PN | SC | NI | NI | PN | NA | NA | NI | PN | SC | PY | NA | NA | NA | L | N | N | NI | N | NA | L | NI | N | N | SC | SC |
| 74 | Luger 2013 | PY | NI | PN | SC | NI | NI | PN | NA | NA | NI | PN | SC | PY | NA | NA | NA | L | N | N | PN | NA | NA | L | PY | N | N | L | SC |
| 75 | Ma 2008 | Y | NI | N | SC | NI | NI | PN | NA | NA | NI | PN | SC | NI | N | NI | PN | SC | N | N | NI | N | NA | L | PY | N | N | L | SC |
| 76 | Marco-Benedí 2020 | Y | Y | N | L | NI | NI | PN | NA | NA | Y | NA | L | N | PY | NA | NA | L | N | N | NI | N | NA | L | PY | N | N | L | L |

| Study ID | Reference | Randomization process | | | | Deviations from intended interventions | | | | | | | | Missing outcome data | | | | | Measurement of the outcome | | | | | | Selection of the reported result | | | | Overall |
|---|---|---|---|---|---|---|---|---|---|---|---|---|---|---|---|---|---|---|---|---|---|---|---|---|---|---|---|---|---|
| | | 1.1 | 1.2 | 1.3 | RoB 1 | 2.1 | 2.2 | 2.3 | 2.4 | 2.5 | 2.6 | 2.7 | RoB 2 | 3.1 | 3.2 | 3.3 | 3.4 | RoB 3 | 4.1 | 4.2 | 4.3 | 4.4 | 4.5 | RoB 4 | 5.1 | 5.2 | 5.3 | RoB 5 | |
| 77 | McLaughlin 2007 | NI | NI | N | SC | NI | NI | N | NA | NA | NI | PN | SC | Y | NA | NA | NA | L | N | N | NI | N | NA | L | NI | N | N | SC | SC |
| 78 | Mehling 2007 | Y | NI | N | SC | Y | Y | NI | NA | NA | NI | PY | H | PY | NA | NA | NA | L | N | N | Y | N | NA | L | NI | N | N | SC | H |
| 79 | Mohammadi 2017 | Y | NI | N | SC | NI | NI | PN | NA | NA | NI | PN | SC | PY | NA | NA | NA | L | N | N | NI | N | NA | L | PY | N | N | L | SC |
| 80 | Mollentze 2019 | Y | NI | N | SC | Y | Y | NI | NA | NA | NI | PN | SC | N | N | NI | PY | H | N | N | Y | N | NA | L | PY | N | N | L | H |
| 81 | Nicholson 1999 | PY | NI | N | SC | NI | NI | NI | NA | NA | N | PN | SC | PN | PY | NA | NA | L | N | N | NI | N | NA | L | NI | N | N | SC | SC |
| 82 | Parker 2002 | PY | NI | N | SC | NI | NI | NI | NA | NA | NI | PN | SC | N | N | NI | PN | SC | N | N | NI | N | NA | L | NI | N | N | SC | SC |
| 83 | Pavithran 2020a | PN | NI | N | SC | NI | NI | PN | NA | NA | N | PN | SC | PY | NA | NA | NA | L | N | N | NI | N | NA | L | NI | N | N | SC | SC |
| 84 | Pavithran 2020b | NI | NI | N | SC | NI | NI | PN | NA | NA | NI | PN | SC | N | N | NI | NI | H | N | N | NI | N | NA | L | NI | N | N | SC | H |
| 85 | Pedersen 2014 | Y | Y | N | L | NI | NI | PN | NA | NA | N | PN | SC | N | PN | NI | PN | SC | N | N | NI | N | NA | L | NI | N | N | SC | SC |
| 86 | Perna 2019 | NI | NI | PN | SC | NI | NI | PN | NA | NA | NI | PN | SC | NI | N | NI | PN | SC | N | N | NI | N | NA | L | NI | N | N | SC | SC |
| 88 | Rock 2014 | Y | NI | N | SC | NI | Y | PN | NA | NA | Y | NA | L | PY | NA | NA | NA | L | N | N | NI | N | NA | L | PY | N | N | L | SC |
| 89 | Ruggenenti 2017 | Y | PY | N | L | Y | Y | PN | NA | NA | Y | NA | L | Y | NA | NA | NA | L | N | N | N | NA | NA | L | PY | N | N | L | L |
| 90 | Ruggenenti 2022 | Y | Y | N | L | Y | Y | PN | NA | NA | Y | NA | L | PN | Y | NA | NA | L | N | N | NI | N | NA | L | Y | N | N | L | L |
| 91 | Saslow 2014/2017 | Y | Y | N | L | Y | Y | PN | NA | NA | Y | NA | L | Y | NA | NA | NA | L | N | N | NI | N | NA | L | Y | N | N | L | L |
| 92 | Sato 2017 | Y | NI | N | SC | Y | Y | PN | NA | NA | N | PN | SC | PN | PN | PY | PN | SC | N | N | NI | N | NA | L | Y | N | N | L | SC |
| 93 | Shige 2000 | NI | NI | PN | SC | NI | NI | N | NA | NA | NI | PN | SC | Y | NA | NA | NA | L | N | N | NI | N | NA | L | NI | N | N | SC | SC |
| 95 | Stentz 2016 | Y | NI | N | SC | NI | NI | NI | NA | NA | N | PN | SC | N | N | PY | NI | H | N | N | NI | N | NA | L | NI | N | N | SC | H |
| 96 | Tay 2015 | Y | Y | N | L | Y | Y | N | NA | NA | Y | NA | L | N | Y | NA | NA | L | N | N | N | NA | NA | L | PY | N | N | L | L |
| 97 | Thomsen 2022 | Y | Y | N | L | Y | Y | PN | NA | NA | Y | NA | L | Y | NA | NA | NA | L | N | N | N | NA | NA | L | PY | N | N | L | L |
| 98 | Uusitupa 1993 | NI | NI | N | SC | NI | NI | PN | NA | NA | NI | PN | SC | Y | NA | NA | NA | L | N | N | NI | N | NA | L | PY | N | N | L | SC |
| 101 | Wang 2018 | Y | Y | N | L | PY | PY | N | NA | NA | PY | NA | L | PN | PY | NA | NA | L | N | N | NI | N | NA | L | PY | N | N | L | L |
| 102 | Watson 2016 | Y | Y | N | L | NI | NI | PN | NA | NA | Y | NA | L | N | Y | NA | NA | L | N | N | NI | N | NA | L | Y | N | N | L | L |
| 103 | Westman 2008 | Y | NI | N | SC | Y | Y | PN | NA | NA | N | PN | SC | N | N | PY | PY | H | N | N | Y | PN | NA | L | NI | N | N | SC | H |
| 105 | Wolever 2008 | Y | Y | N | L | NI | NI | PN | NA | NA | PY | NA | L | N | PN | NI | PN | SC | N | N | NI | N | NA | L | PY | N | N | L | SC |
| 106 | Yamada 2014 | Y | NI | N | SC | Y | Y | PN | NA | NA | NI | N | SC | Y | NA | NA | NA | L | N | N | NI | N | NA | L | NI | N | N | SC | SC |
| 107 | Zahedi 2021 | Y | NI | N | SC | NI | NI | PN | NA | NA | N | PN | SC | N | N | NI | PN | SC | N | N | NI | N | NA | L | NI | N | N | SC | SC |

RoB, risk of bias; Y, yes; PY, probably yes; N, no; PN, probably no; NI, no information; NA, not applicable; H, high risk of bias; SC, some concerns of risk of bias; L, low risk of bias.

**Table S6.2** Risk of bias assessment of crossover randomized controlled trials

| Study ID | Reference | Randomization process | | | | Risk of bias arising from period and carryover effects | | | | Deviations from intended interventions | | | | | | | | Missing outcome data | | | | | Measurement of the outcome | | | | | | Selection of the reported result | | | | | Overall |
|---|---|---|---|---|---|---|---|---|---|---|---|---|---|---|---|---|---|---|---|---|---|---|---|---|---|---|---|---|---|---|---|---|---|---|
| | | **1.1** | **1.2** | **1.3** | **RoB1** | **S.1** | **S.2** | **S.3** | **RoBS** | **2.1** | **2.2** | **2.3** | **2.4** | **2.5** | **2.6** | **2.7** | **RoB2** | **3.1** | **3.2** | **3.3** | **3.4** | **RoB3** | **4.1** | **4.2** | **4.3** | **4.4** | **4.5** | **RoB4** | **5.1** | **5.2** | **5.3** | **5.4** | **RoB5** | |
| 11 | Sun 2007 | NI | NI | N | SC | Y | NA | PY | L | NI | NI | PN | NA | NA | N | PN | SC | Y | NA | NA | NA | L | PN | N | NI | PN | NA | L | NI | N | N | N | SC | SC |
| 32 | Azadbakht 2011 | PN | NI | PN | SC | Y | NA | Y | L | NI | NI | PN | NA | NA | N | PY | H | N | PN | PY | PY | H | N | N | NI | N | NA | L | NI | N | N | N | SC | H |
| 35 | Brand 1991 | PN | NI | NI | SC | Y | NA | PY | L | NI | NI | PN | NA | NA | NI | N | SC | Y | NA | NA | NA | L | N | N | NI | N | NA | L | NI | N | N | N | SC | SC |
| 41 | Chandalia 2000 | PY | NI | NI | SC | Y | NA | PN | H | NI | NI | PY | PN | NA | NI | PN | SC | PY | NA | NA | NA | L | N | N | NI | N | NA | L | Y | N | N | N | L | H |
| 46 | Coulston 1989 | PN | NI | NI | SC | Y | NA | NI | SC | NI | NI | PN | NA | NA | N | PN | SC | Y | NA | NA | NA | L | N | N | NI | PN | NA | L | NI | N | N | N | SC | SC |
| 53 | Gannon 2004 | Y | NI | NI | SC | PN | NI | PY | SC | NI | NI | PN | NA | NA | N | PN | SC | N | PN | NI | NI | H | N | N | NI | N | NA | L | NI | N | N | N | SC | H |
| 54 | Gannon 2003 | Y | NI | NI | SC | NI | NI | Y | SC | NI | NI | PN | NA | NA | N | PN | SC | Y | NA | NA | NA | L | N | N | NI | N | NA | L | NI | N | N | N | SC | SC |
| 64 | Itsiopoulos 2011 | PY | NI | NI | SC | NI | Y | PY | L | NI | NI | PN | NA | NA | NI | PN | SC | NI | N | NI | PN | SC | N | N | NI | N | NA | L | NI | N | N | N | SC | SC |
| 66 | Jimenez-Cruz 2003 | PY | NI | N | SC | NI | NI | Y | SC | NI | NI | PN | NA | NA | NI | PN | SC | N | N | PY | NI | H | N | N | NI | N | NA | L | NI | N | N | N | SC | H |
| 67 | Jönsson 2009 | Y | PY | PN | L | Y | NA | PY | L | Y | Y | PN | NA | NA | Y | NA | L | N | PY | NA | NA | L | N | N | NI | N | NA | L | NI | N | N | N | SC | SC |
| 73 | Lousley 1984 | Y | NI | PN | SC | NI | NI | NI | SC | NI | NI | PN | NA | NA | NI | PN | SC | N | N | NI | PN | SC | N | N | NI | N | NA | L | N | N | N | N | SC | SC |
| 87 | Rizkalla 2004 | NI | NI | NI | SC | NI | N | Y | SC | NI | NI | PN | NA | NA | NI | PN | SC | Y | NA | NA | NA | L | N | N | NI | N | NA | L | PY | N | N | N | L | SC |
| 94 | Skytte 2019 | Y | NI | NI | SC | Y | NA | N | H | NI | NI | PN | NA | NA | NI | PN | SC | Y | NA | NA | NA | L | N | N | NI | N | NA | L | PY | N | N | N | L | H |
| 99 | Visek 2014 | NI | NI | NI | SC | NI | NI | PN | H | Y | Y | PN | NA | NA | PY | NA | L | Y | NA | NA | NA | L | N | N | NI | N | NA | L | NI | N | N | N | SC | H |
| 100 | Walker 1995 | NI | NI | N | SC | NI | N | Y | SC | NI | NI | N | NA | NA | NI | PN | SC | Y | NA | NA | NA | L | N | N | NI | N | NA | L | PY | N | N | N | L | SC |
| 104 | Wolever 1992 | NI | NI | NI | SC | NI | N | Y | SC | NI | NI | N | NA | NA | N | N | SC | Y | NA | NA | NA | L | N | N | NI | N | NA | L | NI | N | N | N | SC | SC |

RoB, risk of bias; Y, yes; PY, probably yes; N, no; PN, probably no; NI, no information; NA, not applicable; H, high risk of bias; SC, some concerns of risk of bias; L, low risk of bias.

### File S7: Network plots

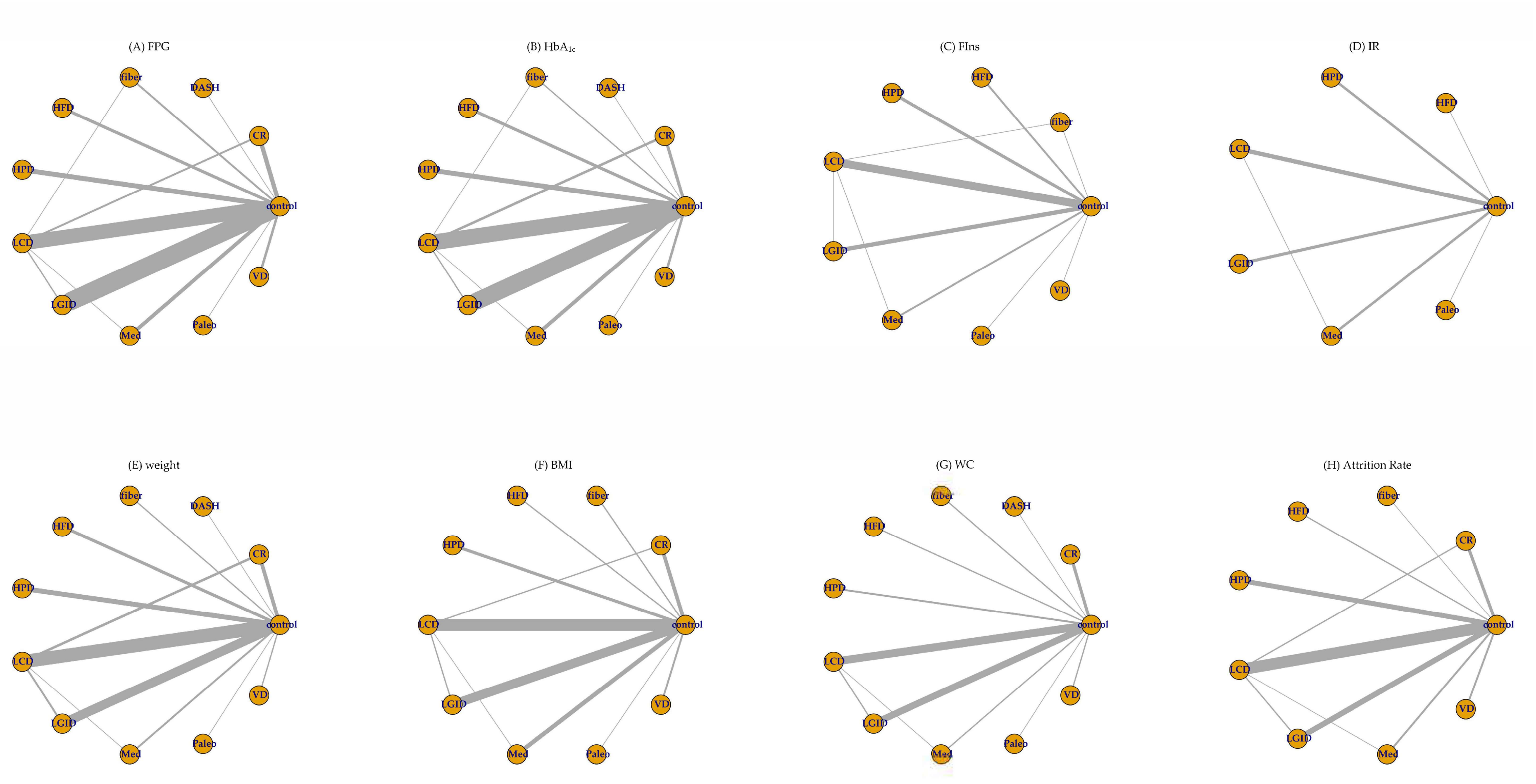

(I) SBP

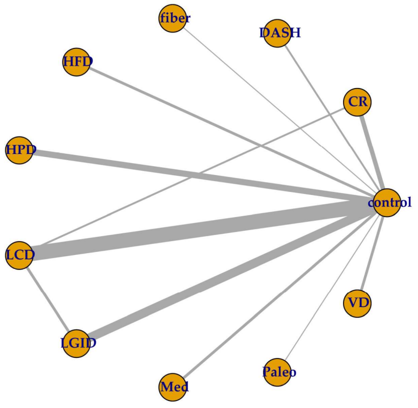

(J) DBP

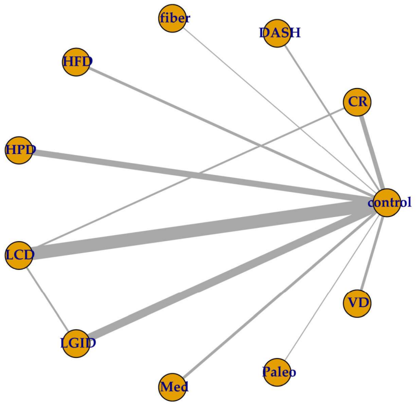

(K) TG

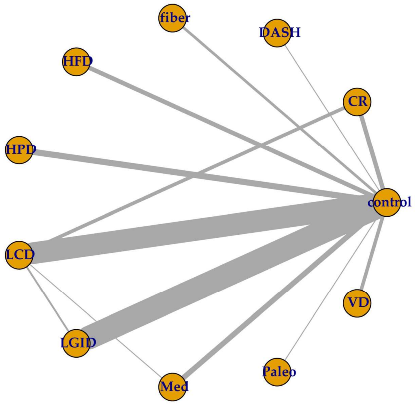

(L) TC

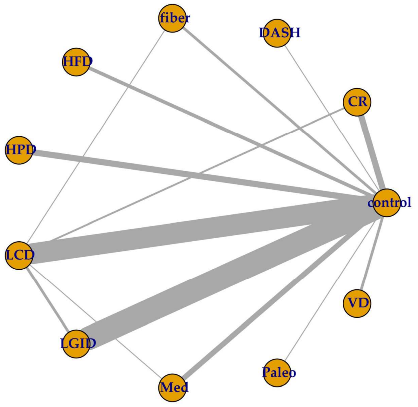

(M) LDL

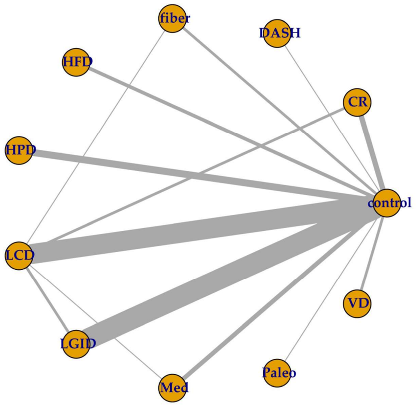

(N) HDL

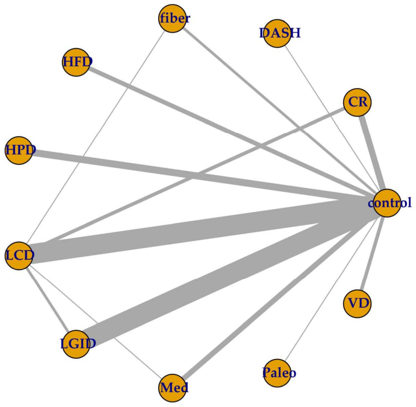

### File S8: League tables and cumulative ranking curves

#### S8.1 League tables

FPG (Right vs Left), mmol/L; $HbA_{1c}$ (Left vs Right), %; Data were *MD* [95% *CrI*]. Blue cells represented significant lower values, and red cells for significant higher values.

| | control | CR | DASH | fiber | HFD | HPD | LCD | LGID | Med | Paleo | VD |
|---|---|---|---|---|---|---|---|---|---|---|---|
| control | control | -0.814 [-1.372, -0.254] | -0.919 [-2.541, 0.702] | -1.252 [-2.319, -0.194] | -0.007 [-0.790, 0.772] | -0.294 [-0.884, 0.296] | -0.818 [-1.124, -0.510] | -0.936 [-1.218, -0.650] | -0.454 [-1.063, 0.154] | -0.502 [-2.179, 1.186] | -0.635 [-1.565, 0.291] |
| CR | -0.343 [-0.757, 0.072] | CR | -0.106 [-1.822, 1.607] | -0.437 [-1.639, 0.756] | 0.805 [-0.155, 1.765] | 0.521 [-0.296, 1.330] | -0.004 [-0.601, 0.590] | -0.122 [-0.750, 0.502] | 0.360 [-0.463, 1.182] | 0.312 [-1.460, 2.083] | 0.179 [-0.908, 1.257] |
| DASH | -1.198 [-2.175, -0.226] | -0.854 [-1.919, 0.201] | DASH | -0.332 [-2.277, 1.603] | 0.913 [-0.888, 2.709] | 0.628 [-1.106, 2.349] | 0.103 [-1.546, 1.752] | -0.017 [-1.661, 1.628] | 0.467 [-1.263, 2.192] | 0.416 [-1.916, 2.746] | 0.283 [-1.586, 2.153] |
| fiber | -0.735 [-1.516, 0.035] | -0.392 [-1.275, 0.482] | 0.463 [-0.789, 1.704] | fiber | 1.245 [-0.070, 2.560] | 0.958 [-0.253, 2.174] | 0.434 [-0.666, 1.534] | 0.316 [-0.779, 1.419] | 0.797 [-0.426, 2.020] | 0.749 [-1.237, 2.741] | 0.617 [-0.793, 2.027] |
| HFD | -0.105 [-0.626, 0.417] | 0.238 [-0.428, 0.906] | 1.093 [-0.008, 2.202] | 0.629 [-0.297, 1.573] | HFD | -0.285 [-1.267, 0.692] | -0.810 [-1.646, 0.032] | -0.929 [-1.756, -0.093] | -0.445 [-1.437, 0.543] | -0.494 [-2.346, 1.366] | -0.625 [-1.843, 0.583] |
| HPD | -0.181 [-0.570, 0.206] | 0.163 [-0.406, 0.726] | 1.017 [-0.028, 2.070] | 0.553 [-0.306, 1.425] | -0.076 [-0.730, 0.572] | HPD | -0.525 [-1.190, 0.142] | -0.642 [-1.298, 0.017] | -0.161 [-1.007, 0.693] | -0.209 [-1.979, 1.583] | -0.341 [-1.440, 0.758] |
| LCD | -0.667 [-0.879, -0.460] | -0.324 [-0.748, 0.093] | 0.531 [-0.467, 1.527] | 0.067 [-0.724, 0.866] | -0.562 [-1.127, -0.004] | -0.486 [-0.929, -0.047] | LCD | -0.118 [-0.527, 0.292] | 0.363 [-0.298, 1.027] | 0.315 [-1.390, 2.032] | 0.183 [-0.799, 1.157] |
| LGID | -0.711 [-0.929, -0.493] | -0.368 [-0.835, 0.098] | 0.486 [-0.512, 1.488] | 0.024 [-0.775, 0.835] | -0.605 [-1.172, -0.044] | -0.530 [-0.974, -0.085] | -0.044 [-0.341, 0.258] | LGID | 0.482 [-0.190, 1.154] | 0.433 [-1.268, 2.141] | 0.300 [-0.671, 1.271] |
| Med | -0.461 [-0.900, -0.021] | -0.117 [-0.718, 0.481] | 0.737 [-0.330, 1.809] | 0.273 [-0.614, 1.171] | -0.357 [-1.039, 0.325] | -0.281 [-0.865, 0.307] | 0.207 [-0.267, 0.686] | 0.250 [-0.239, 0.742] | Med | -0.048 [-1.830, 1.747] | -0.180 [-1.296, 0.927] |
| Paleo | -0.402 [-1.510, 0.708] | -0.059 [-1.243, 1.124] | 0.796 [-0.674, 2.271] | 0.331 [-1.016, 1.692] | -0.295 [-1.524, 0.924] | -0.221 [-1.395, 0.954] | 0.265 [-0.856, 1.397] | 0.309 [-0.820, 1.441] | 0.058 [-1.137, 1.251] | Paleo | -0.132 [-2.056, 1.773] |
| VD | -0.324 [-0.894, 0.246] | 0.019 [-0.686, 0.721] | 0.875 [-0.254, 2.005] | 0.412 [-0.547, 1.375] | -0.218 [-0.992, 0.552] | -0.142 [-0.830, 0.545] | 0.344 [-0.259, 0.953] | 0.388 [-0.223, 0.996] | 0.138 [-0.585, 0.856] | 0.078 [-1.170, 1.321] | VD |

| FIns (Right vs Left), *PMD* [95% *CrI*] | | | | | | | | | |
|---|---|---|---|---|---|---|---|---|---|
| | control | fiber | HFD | HPD | LCD | LGID | Med | Paleo | VD |
| control | control | -0.206 [-0.462, 0.052] | -0.050 [-0.259, 0.157] | -0.092 [-0.261, 0.082] | -0.119 [-0.215, -0.020] | -0.138 [-0.265, -0.009] | -0.075 [-0.256, 0.104] | 0.021 [-0.364, 0.407] | -0.065 [-0.456, 0.326] |
| | | fiber | 0.157 [-0.175, 0.484] | 0.115 [-0.193, 0.424] | 0.087 [-0.179, 0.355] | 0.069 [-0.218, 0.354] | 0.132 [-0.183, 0.443] | 0.228 [-0.235, 0.689] | 0.141 [-0.325, 0.608] |
| HFD | -0.152 [-0.434, 0.131] | | HFD | -0.042 [-0.308, 0.229] | -0.069 [-0.295, 0.162] | -0.088 [-0.330, 0.158] | -0.025 [-0.300, 0.250] | 0.071 [-0.366, 0.510] | -0.015 [-0.459, 0.427] |
| HPD | -0.218 [-0.366, -0.068] | | -0.066 [-0.388, 0.256] | HPD | -0.027 [-0.225, 0.168] | -0.045 [-0.262, 0.166] | 0.017 [-0.235, 0.262] | 0.113 [-0.310, 0.533] | 0.027 [-0.404, 0.451] |
| LCD | -0.086 [-0.172, 0.003] | | 0.066 [-0.230, 0.363] | 0.132 [-0.042, 0.305] | LCD | -0.018 [-0.172, 0.132] | 0.045 [-0.154, 0.237] | 0.141 [-0.258, 0.536] | 0.054 [-0.350, 0.455] |
| LGID | -0.155 [-0.278, -0.040] | | -0.004 [-0.313, 0.302] | 0.063 [-0.132, 0.248] | -0.070 [-0.220, 0.073] | LGID | 0.063 [-0.158, 0.282] | 0.159 [-0.247, 0.566] | 0.073 [-0.339, 0.483] |
| Med | -0.098 [-0.191, 0.010] | | 0.056 [-0.240, 0.359] | 0.122 [-0.055, 0.304] | -0.011 [-0.132, 0.120] | 0.058 [-0.087, 0.226] | Med | 0.096 [-0.327, 0.521] | 0.010 [-0.421, 0.440] |
| Paleo | -0.001 [-0.290, 0.286] | | 0.150 [-0.254, 0.555] | 0.217 [-0.108, 0.541] | 0.085 [-0.217, 0.384] | 0.155 [-0.153, 0.467] | 0.094 [-0.213, 0.397] | Paleo | -0.086 [-0.636, 0.463] |
| IR (Left vs Right), *PMD* [95% *CrI*] | | | | | | | | | VD |

| Weight (Right vs Left), kg, *MD* [95% *CrI*] | | | | | | | | | | | |
|---|---|---|---|---|---|---|---|---|---|---|---|
| | control | CR | DASH | fiber | HFD | HPD | LCD | LGID | Med | Paleo | VD |
| control | control | -4.086<br>[-6.13, -2.038] | -3.001<br>[-9.040, 3.087] | -0.878<br>[-5.246, 3.507] | -0.614<br>[-3.349, 2.117] | -0.497<br>[-2.712, 1.704] | -3.003<br>[-4.250, -1.760] | -1.122<br>[-2.694, 0.457] | -0.581<br>[-3.793, 2.620] | -2.988<br>[-9.995, 3.978] | -2.091<br>[-7.123, 2.955] |
| CR | -1.131<br>[-1.927, -0.343] | CR | 1.085<br>[-5.305, 7.490] | 3.211<br>[-1.617, 8.053] | 3.473<br>[0.038, 6.878] | 3.590<br>[0.574, 6.600] | 1.089<br>[-1.072, 3.221] | 2.963<br>[0.427, 5.529] | 3.505<br>[-0.283, 7.289] | 1.102<br>[-6.203, 8.364] | 1.998<br>[-3.428, 7.456] |
| DASH | | | DASH | 2.122<br>[-5.379, 9.576] | 2.387<br>[-4.284, 9.01] | 2.507<br>[-3.976, 8.932] | 0.004<br>[-6.211, 6.168] | 1.88<br>[-4.384, 8.12] | 2.417<br>[-4.468, 9.258] | 0.013<br>[-9.254, 9.240] | 0.910<br>[-6.971, 8.785] |
| fiber | -0.291<br>[-1.826, 1.249] | 0.841<br>[-0.88, 2.572] | | fiber | 0.261<br>[-4.884, 5.42] | 0.382<br>[-4.523, 5.279] | -2.126<br>[-6.69, 2.43] | -0.245<br>[-4.894, 4.419] | 0.290<br>[-5.131, 5.729] | -2.113<br>[-10.35, 6.122] | -1.214<br>[-7.879, 5.475] |
| HFD | -0.351<br>[-1.851, 1.148] | 0.781<br>[-0.920, 2.485] | | -0.059<br>[-2.213, 2.086] | HFD | 0.118<br>[-3.399, 3.633] | -2.389<br>[-5.385, 0.625] | -0.509<br>[-3.645, 2.659] | 0.028<br>[-4.182, 4.249] | -2.366<br>[-9.912, 5.100] | -1.474<br>[-7.195, 4.251] |
| HPD | -0.429<br>[-1.456, 0.594] | 0.701<br>[-0.592, 1.996] | | -0.138<br>[-1.985, 1.706] | -0.081<br>[-1.891, 1.743] | HPD | -2.505<br>[-5.032, 0.023] | -0.627<br>[-3.321, 2.110] | -0.083<br>[-3.984, 3.795] | -2.500<br>[-9.844, 4.824] | -1.592<br>[-7.068, 3.934] |
| LCD | -1.205<br>[-1.674, -0.737] | -0.073<br>[-0.928, 0.790] | | -0.917<br>[-2.524, 0.692] | -0.854<br>[-2.421, 0.716] | -0.777<br>[-1.899, 0.350] | LCD | 1.880<br>[-0.034, 3.802] | 2.423<br>[-0.950, 5.767] | 0.015<br>[-7.106, 7.077] | 0.910<br>[-4.258, 6.124] |
| LGID | -0.732<br>[-1.280, -0.183] | 0.399<br>[-0.555, 1.368] | | -0.441<br>[-2.074, 1.191] | -0.381<br>[-1.975, 1.219] | -0.303<br>[-1.462, 0.863] | 0.474<br>[-0.222, 1.167] | LGID | 0.538<br>[-3.037, 4.094] | -1.868<br>[-9.072, 5.262] | -0.966<br>[-6.243, 4.324] |
| Med | -0.656<br>[-1.448, 0.137] | 0.474<br>[-0.637, 1.593] | | -0.366<br>[-2.092, 1.363] | -0.308<br>[-2.002, 1.397] | -0.227<br>[-1.523, 1.074] | 0.549<br>[-0.347, 1.444] | 0.075<br>[-0.886, 1.037] | Med | -2.411<br>[-10.10, 5.270] | -1.506<br>[-7.453, 4.465] |
| Paleo | -0.997<br>[-3.817, 1.817] | 0.133<br>[-2.782, 3.061] | | -0.712<br>[-3.911, 2.506] | -0.65<br>[-3.837, 2.546] | -0.569<br>[-3.565, 2.434] | 0.208<br>[-2.651, 3.063] | -0.265<br>[-3.141, 2.603] | -0.34<br>[-3.275, 2.591] | Paleo | 0.911<br>[-7.683, 9.539] |
| VD | -0.685<br>[-1.941, 0.565] | 0.447<br>[-1.035, 1.937] | | -0.391<br>[-2.385, 1.592] | -0.333<br>[-2.286, 1.626] | -0.252<br>[-1.877, 1.366] | 0.521<br>[-0.825, 1.856] | 0.048<br>[-1.33, 1.416] | -0.028<br>[-1.517, 1.455] | 0.310<br>[-2.770, 3.390] | VD |
| BMI (Left vs Right), $kg/m^2$, *MD* [95% *CrI*] | | | | | | | | | | | |

| WC (Right vs Left), cm, *MD* [95% *CrI*] | | | | | | | | | | |
|---|---|---|---|---|---|---|---|---|---|---|
| control | CR | DASH | fiber | HFD | HPD | LCD | LGID | Med | Paleo | VD |
| control | -4.499 [-7.359, -1.762] | -4.801 [-10.70, 1.071] | -1.068 [-5.324, 3.179] | -2.399 [-6.635, 1.843] | 0.528 [-2.838, 3.871] | -3.013 [-4.682, -1.328] | -2.078 [-3.862, -0.272] | -0.770 [-4.379, 2.852] | -3.987 [-11.57, 3.600] | -2.347 [-6.418, 1.751] |
| | CR | -0.303 [-6.769, 6.275] | 3.426 [-1.596, 8.593] | 2.103 [-2.936, 7.256] | 5.028 [0.714, 9.450] | 1.484 [-1.723, 4.810] | 2.421 [-0.834, 5.814] | 3.724 [-0.777, 8.366] | 0.511 [-7.515, 8.635] | 2.152 [-2.728, 7.177] |
| | | DASH | 3.722 [-3.517, 11.01] | 2.406 [-4.853, 9.657] | 5.334 [-1.460, 12.11] | 1.793 [-4.306, 7.936] | 2.72 [-3.415, 8.909] | 4.033 [-2.864, 10.93] | 0.817 [-8.786, 10.44] | 2.452 [-4.729, 9.63] |
| | | | fiber | -1.324 [-7.324, 4.673] | 1.599 [-3.825, 6.993] | -1.942 [-6.511, 2.635] | -1.007 [-5.625, 3.604] | 0.302 [-5.295, 5.883] | -2.924 [-11.61, 5.758] | -1.291 [-7.150, 4.634] |
| | | | | HFD | 2.922 [-2.495, 8.311] | -0.616 [-5.181, 3.937] | 0.317 [-4.285, 4.938] | 1.632 [-3.955, 7.166] | -1.584 [-10.25, 7.083] | 0.049 [-5.835, 5.931] |
| | | | | | HPD | -3.544 [-7.266, 0.237] | -2.604 [-6.396, 1.234] | -1.301 [-6.215, 3.645] | -4.511 [-12.82, 3.777] | -2.879 [-8.146, 2.432] |
| | | | | | | LCD | 0.932 [-1.392, 3.257] | 2.244 [-1.539, 6.014] | -0.975 [-8.739, 6.803] | 0.66 [-3.764, 5.082] |
| | | | | | | | LGID | 1.313 [-2.718, 5.309] | -1.915 [-9.694, 5.903] | -0.271 [-4.74, 4.186] |
| | | | | | | | | Med | -3.227 [-11.60, 5.200] | -1.585 [-7.037, 3.867] |
| | | | | | | | | | Paleo | 1.646 [-7.017, 10.26] |
| | | | | | | | | | | VD |

| SBP (Right vs Left), mmHg, *MD* [95% *CrI*] | | | | | | | | | | | |
|---|---|---|---|---|---|---|---|---|---|---|---|
| | control | CR | DASH | fiber | HFD | HPD | LCD | LGID | Med | Paleo | VD |
| control | control | -1.760 [-5.667, 2.044] | -7.576 [-14.93, -0.289] | -1.578 [-10.45, 7.308] | 1.156 [-4.120, 6.420] | -2.727 [-6.293, 0.710] | -2.218 [-4.289, -0.101] | -0.760 [-3.657, 2.234] | -0.823 [-5.421, 3.813] | -8.942 [-24.23, 6.354] | -0.111 [-6.170, 6.108] |
| CR | -1.963 [-5.198, 1.226] | CR | -5.815 [-14.07, 2.462] | 0.191 [-9.412, 9.899] | 2.920 [-3.579, 9.500] | -0.964 [-6.201, 4.205] | -0.451 [-4.528, 3.749] | 0.998 [-3.710, 5.944] | 0.930 [-4.991, 7.029] | -7.185 [-22.96, 8.620] | 1.656 [-5.479, 9.025] |
| DASH | -3.713 [-10.33, 2.814] | -1.749 [-9.092, 5.507] | DASH | 6.003 [-5.502, 17.53] | 8.745 [-0.279, 17.77] | 4.849 [-3.298, 12.94] | 5.360 [-2.221, 13.03] | 6.820 [-0.997, 14.79] | 6.771 [-1.841, 15.42] | -1.366 [-18.36, 15.61] | 7.482 [-1.983, 17.10] |
| fiber | -0.733 [-8.166, 6.747] | 1.238 [-6.849, 9.408] | 3.002 [-6.935, 12.98] | fiber | 2.737 [-7.599, 13.07] | -1.15 [-10.76, 8.336] | -0.638 [-9.721, 8.488] | 0.816 [-8.472, 10.26] | 0.752 [-9.220, 10.79] | -7.393 [-25.00, 10.32] | 1.468 [-9.253, 12.31] |
| HFD | 0.768 [-3.696, 5.220] | 2.722 [-2.734, 8.211] | 4.470 [-3.376, 12.45] | 1.489 [-7.191, 10.13] | HFD | -3.887 [-10.27, 2.370] | -3.374 [-9.025, 2.341] | -1.917 [-7.879, 4.191] | -1.988 [-8.941, 5.065] | -10.10 [-26.25, 6.064] | -1.265 [-9.263, 6.929] |
| HPD | -2.965 [-5.920, -0.068] | -1.002 [-5.347, 3.339] | 0.747 [-6.438, 7.969] | -2.239 [-10.28, 5.733] | -3.722 [-9.083, 1.595] | HPD | 0.510 [-3.481, 4.695] | 1.973 [-2.493, 6.680] | 1.897 [-3.799, 7.805] | -6.214 [-21.87, 9.473] | 2.616 [-4.346, 9.840] |
| LCD | -1.952 [-3.783, -0.069] | 0.010 [-3.432, 3.543] | 1.770 [-5.012, 8.661] | -1.227 [-8.900, 6.447] | -2.714 [-7.511, 2.140] | 1.013 [-2.410, 4.538] | LCD | 1.455 [-1.907, 4.898] | 1.398 [-3.679, 6.437] | -6.738 [-22.19, 8.708] | 2.111 [-4.331, 8.627] |
| LGID | -0.828 [-3.310, 1.664] | 1.136 [-2.870, 5.193] | 2.892 [-4.085, 9.957] | -0.099 [-7.964, 7.728] | -1.590 [-6.693, 3.526] | 2.135 [-1.674, 6.011] | 1.120 [-1.832, 4.048] | LGID | -0.054 [-5.594, 5.338] | -8.202 [-23.78, 7.355] | 0.648 [-6.126, 7.473] |
| Med | -0.479 [-4.601, 3.660] | 1.487 [-3.710, 6.726] | 3.242 [-4.432, 11.03] | 0.243 [-8.280, 8.789] | -1.243 [-7.319, 4.847] | 2.487 [-2.556, 7.601] | 1.471 [-3.071, 5.972] | 0.347 [-4.472, 5.168] | Med | -8.143 [-24.10, 7.823] | 0.712 [-6.892, 8.457] |
| Paleo | -3.971 [-13.22, 5.260] | -2.02 [-11.78, 7.771] | -0.264 [-11.54, 11.09] | -3.257 [-15.10, 8.628] | -4.743 [-15.00, 5.551] | -1.008 [-10.70, 8.695] | -2.020 [-11.50, 7.364] | -3.147 [-12.77, 6.411] | -3.491 [-13.64, 6.642] | Paleo | 8.86 [-7.590, 25.26] |
| VD | 0.980 [-3.935, 5.946] | 2.943 [-2.908, 8.915] | 4.699 [-3.479, 12.99] | 1.713 [-7.263, 10.67] | 0.216 [-6.400, 6.919] | 3.947 [-1.744, 9.754] | 2.934 [-2.330, 8.209] | 1.815 [-3.714, 7.372] | 1.460 [-4.981, 7.942] | 4.964 [-5.469, 15.47] | VD |
| DBP (Left vs Right), mmHg, *MD* [95% *CrI*] | | | | | | | | | | | |

TG (Right vs Left), mmol/L, *MD* [95% *CrI*]

| | control | CR | DASH | fiber | HFD | HPD | LCD | LGID | Med | Paleo | VD |
|---|---|---|---|---|---|---|---|---|---|---|---|
| control | control | -0.113 [-0.293, 0.066] | -0.040 [-0.530, 0.450] | -0.186 [-0.501, 0.131] | -0.176 [-0.515, 0.162] | -0.233 [-0.464, -0.003] | -0.289 [-0.396, -0.179] | -0.255 [-0.353, -0.154] | -0.201 [-0.408, 0.005] | -0.500 [-1.128, 0.129] | -0.024 [-0.360, 0.310] |
| CR | -0.170 [-0.426, 0.089] | CR | 0.074 [-0.448, 0.594] | -0.072 [-0.436, 0.293] | -0.063 [-0.446, 0.319] | -0.120 [-0.411, 0.171] | -0.175 [-0.361, 0.012] | -0.141 [-0.344, 0.064] | -0.088 [-0.359, 0.183] | -0.387 [-1.042, 0.266] | 0.09 [-0.291, 0.468] |
| DASH | -0.357 [-1.134, 0.420] | -0.188 [-1.007, 0.633] | DASH | -0.146 [-0.729, 0.437] | -0.136 [-0.730, 0.457] | -0.194 [-0.735, 0.347] | -0.249 [-0.748, 0.254] | -0.215 [-0.713, 0.287] | -0.161 [-0.694, 0.370] | -0.461 [-1.253, 0.335] | 0.016 [-0.577, 0.608] |
| fiber | -0.328 [-0.764, 0.107] | -0.158 [-0.661, 0.345] | 0.029 [-0.862, 0.916] | fiber | 0.010 [-0.457, 0.473] | -0.048 [-0.438, 0.342] | -0.103 [-0.437, 0.233] | -0.069 [-0.400, 0.264] | -0.015 [-0.395, 0.364] | -0.315 [-1.017, 0.39] | 0.163 [-0.300, 0.623] |
| HFD | -0.115 [-0.516, 0.282] | 0.054 [-0.421, 0.526] | 0.242 [-0.638, 1.112] | 0.212 [-0.379, 0.803] | HFD | -0.058 [-0.466, 0.354] | -0.112 [-0.468, 0.245] | -0.079 [-0.431, 0.277] | -0.025 [-0.422, 0.374] | -0.325 [-1.039, 0.394] | 0.152 [-0.323, 0.628] |
| HPD | -0.150 [-0.440, 0.141] | 0.020 [-0.369, 0.408] | 0.208 [-0.628, 1.037] | 0.179 [-0.344, 0.701] | -0.033 [-0.527, 0.459] | HPD | -0.055 [-0.308, 0.201] | -0.021 [-0.271, 0.230] | 0.032 [-0.278, 0.343] | -0.267 [-0.937, 0.405] | 0.210 [-0.198, 0.618] |
| LCD | -0.165 [-0.318, -0.012] | 0.004 [-0.274, 0.283] | 0.192 [-0.600, 0.984] | 0.163 [-0.285, 0.614] | -0.050 [-0.477, 0.379] | -0.016 [-0.343, 0.313] | LCD | 0.034 [-0.109, 0.177] | 0.087 [-0.139, 0.311] | -0.212 [-0.849, 0.425] | 0.265 [-0.089, 0.615] |
| LGID | -0.461 [-0.619, -0.304] | -0.292 [-0.592, 0.009] | -0.104 [-0.898, 0.688] | -0.133 [-0.594, 0.330] | -0.346 [-0.773, 0.085] | -0.312 [-0.642, 0.019] | -0.296 [-0.506, -0.087] | LGID | 0.054 [-0.176, 0.282] | -0.246 [-0.883, 0.391] | 0.231 [-0.121, 0.578] |
| Med | -0.177 [-0.474, 0.122] | -0.008 [-0.400, 0.385] | 0.180 [-0.655, 1.014] | 0.151 [-0.376, 0.678] | -0.061 [-0.559, 0.440] | -0.028 [-0.442, 0.390] | -0.012 [-0.338, 0.315] | 0.284 [-0.052, 0.621] | Med | -0.299 [-0.96, 0.363] | 0.177 [-0.216, 0.571] |
| Paleo | -0.199 [-1.252, 0.867] | -0.030 [-1.114, 1.066] | 0.159 [-1.153, 1.477] | 0.128 [-1.009, 1.279] | -0.083 [-1.211, 1.056] | -0.050 [-1.141, 1.057] | -0.034 [-1.097, 1.044] | 0.262 [-0.803, 1.339] | -0.022 [-1.116, 1.083] | Paleo | 0.477 [-0.238, 1.186] |
| VD | -0.105 [-0.581, 0.372] | 0.065 [-0.476, 0.607] | 0.253 [-0.661, 1.164] | 0.223 [-0.422, 0.871] | 0.011 [-0.608, 0.634] | 0.046 [-0.515, 0.605] | 0.060 [-0.440, 0.561] | 0.357 [-0.146, 0.860] | 0.072 [-0.489, 0.637] | 0.095 [-1.076, 1.253] | VD |

TC (Left vs Right), mmol/L, *MD [95% CrI]*

| LDL (Right vs Left), mmol/L, *MD* [95% *CrI*] | | | | | | | | | | | |
|---|---|---|---|---|---|---|---|---|---|---|---|
| | control | CR | DASH | fiber | HFD | HPD | LCD | LGID | Med | Paleo | VD |
| control | control | -0.243 [-0.443, -0.039] | -0.372 [-0.891, 0.152] | -0.235 [-0.578, 0.106] | -0.067 [-0.347, 0.212] | -0.110 [-0.309, 0.088] | -0.108 [-0.219, 0.004] | -0.352 [-0.466, -0.237] | -0.029 [-0.268, 0.209] | -0.102 [-0.920, 0.716] | -0.060 [-0.360, 0.240] |
| CR | 0.084 [0.008, 0.160] | CR | -0.128 [-0.690, 0.432] | 0.008 [-0.390, 0.399] | 0.176 [-0.173, 0.518] | 0.133 [-0.154, 0.413] | 0.135 [-0.080, 0.345] | -0.109 [-0.342, 0.119] | 0.214 [-0.100, 0.522] | 0.141 [-0.703, 0.982] | 0.183 [-0.181, 0.543] |
| DASH | 0.081 [-0.150, 0.312] | -0.004 [-0.245, 0.240] | DASH | 0.137 [-0.487, 0.756] | 0.304 [-0.291, 0.895] | 0.262 [-0.300, 0.817] | 0.264 [-0.271, 0.798] | 0.020 [-0.517, 0.552] | 0.343 [-0.234, 0.913] | 0.269 [-0.699, 1.239] | 0.312 [-0.291, 0.912] |
| fiber | 0.077 [-0.059, 0.215] | -0.007 [-0.161, 0.150] | -0.003 [-0.271, 0.266] | fiber | 0.168 [-0.272, 0.610] | 0.124 [-0.268, 0.521] | 0.127 [-0.226, 0.483] | -0.118 [-0.475, 0.245] | 0.206 [-0.209, 0.624] | 0.132 [-0.754, 1.020] | 0.175 [-0.279, 0.632] |
| HFD | 0.040 [-0.074, 0.153] | -0.045 [-0.181, 0.093] | -0.041 [-0.298, 0.216] | -0.038 [-0.217, 0.139] | HFD | -0.044 [-0.386, 0.300] | -0.041 [-0.341, 0.262] | -0.286 [-0.586, 0.018] | 0.038 [-0.329, 0.406] | -0.035 [-0.897, 0.833] | 0.007 [-0.402, 0.418] |
| HPD | -0.017 [-0.105, 0.072] | -0.101 [-0.217, 0.016] | -0.097 [-0.345, 0.149] | -0.094 [-0.258, 0.068] | -0.056 [-0.200, 0.088] | HPD | 0.002 [-0.225, 0.230] | -0.242 [-0.470, -0.013] | 0.081 [-0.229, 0.391] | 0.009 [-0.834, 0.852] | 0.05 [-0.308, 0.411] |
| LCD | 0.120 [0.073, 0.167] | 0.035 [-0.044, 0.116] | 0.039 [-0.197, 0.275] | 0.042 [-0.099, 0.183] | 0.080 [-0.043, 0.203] | 0.137 [0.036, 0.237] | LCD | -0.244 [-0.397, -0.092] | 0.079 [-0.178, 0.334] | 0.006 [-0.821, 0.834] | 0.049 [-0.274, 0.367] |
| LGID | 0.080 [0.028, 0.132] | -0.004 [-0.095, 0.087] | -0.001 [-0.237, 0.235] | 0.003 [-0.144, 0.148] | 0.041 [-0.085, 0.165] | 0.097 [-0.006, 0.200] | -0.040 [-0.107, 0.027] | LGID | 0.323 [0.059, 0.587] | 0.251 [-0.577, 1.078] | 0.293 [-0.029, 0.614] |
| Med | 0.062 [-0.039, 0.162] | -0.023 [-0.147, 0.103] | -0.019 [-0.271, 0.233] | -0.016 [-0.185, 0.153] | 0.022 [-0.129, 0.174] | 0.078 [-0.055, 0.213] | -0.058 [-0.166, 0.051] | -0.019 [-0.131, 0.095] | Med | -0.073 [-0.926, 0.780] | -0.031 [-0.413, 0.353] |
| Paleo | 0.080 [-0.178, 0.338] | -0.004 [-0.273, 0.264] | 0.000 [-0.347, 0.345] | 0.003 [-0.291, 0.294] | 0.041 [-0.242, 0.322] | 0.097 [-0.176, 0.370] | -0.040 [-0.301, 0.223] | 0.000 [-0.263, 0.264] | 0.018 [-0.259, 0.296] | Paleo | 0.042 [-0.829, 0.913] |
| VD | -0.045 [-0.170, 0.079] | -0.129 [-0.275, 0.017] | -0.126 [-0.389, 0.136] | -0.122 [-0.309, 0.062] | -0.085 [-0.254, 0.084] | -0.028 [-0.182, 0.124] | -0.165 [-0.299, -0.031] | -0.125 [-0.261, 0.010] | -0.107 [-0.268, 0.053] | -0.125 [-0.412, 0.161] | VD |
| HDL (Left vs Right), mmol/L, *MD* [95% *CrI*] | | | | | | | | | | | |

| Average weekly attrition (Right vs Left), *RR* | | | | | | | | |
|---|---|---|---|---|---|---|---|---|
| control | CR | fiber | HFD | HPD | LCD | LGID | Med | VD |
| control | 0.803 [0.499, 1.288] | 1.810 [0.500, 7.439] | 1.338 [0.707, 2.517] | 1.023 [0.744, 1.400] | 0.980 [0.787, 1.228] | 0.815 [0.604, 1.105] | 1.252 [0.833, 1.920] | 0.901 [0.401, 2.012] |
| | CR | 2.260 [0.572, 9.812] | 1.661 [0.746, 3.712] | 1.273 [0.721, 2.256] | 1.218 [0.783, 1.934] | 1.016 [0.591, 1.771] | 1.561 [0.850, 2.917] | 1.121 [0.434, 2.877] |
| | | fiber | 0.737 [0.158, 3.104] | 0.562 [0.133, 2.139] | 0.541 [0.131, 1.994] | 0.450 [0.105, 1.717] | 0.692 [0.16, 2.670] | 0.493 [0.098, 2.303] |
| | | | HFD | 0.766 [0.376, 1.545] | 0.732 [0.375, 1.451] | 0.609 [0.303, 1.239] | 0.937 [0.444, 2.032] | 0.675 [0.241, 1.886] |
| | | | | HPD | 0.957 [0.655, 1.420] | 0.797 [0.517, 1.237] | 1.223 [0.734, 2.100] | 0.882 [0.369, 2.099] |
| | | | | | LCD | 0.832 [0.592, 1.165] | 1.279 [0.832, 1.988] | 0.921 [0.394, 2.125] |
| | | | | | | LGID | 1.537 [0.934, 2.569] | 1.104 [0.464, 2.628] |
| | | | | | | | Med | 0.720 [0.283, 1.782] |
| | | | | | | | | VD |

**S8.2 Cumulative ranking curves**

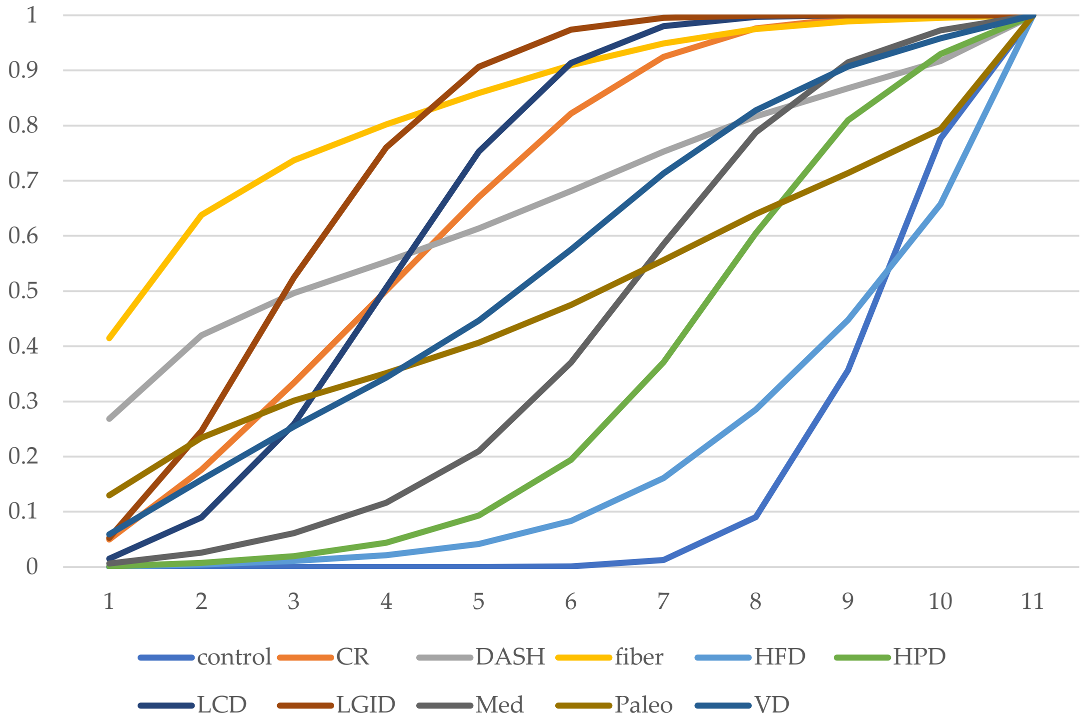

**Figure S8.1** Cumulative ranking curve of FPG

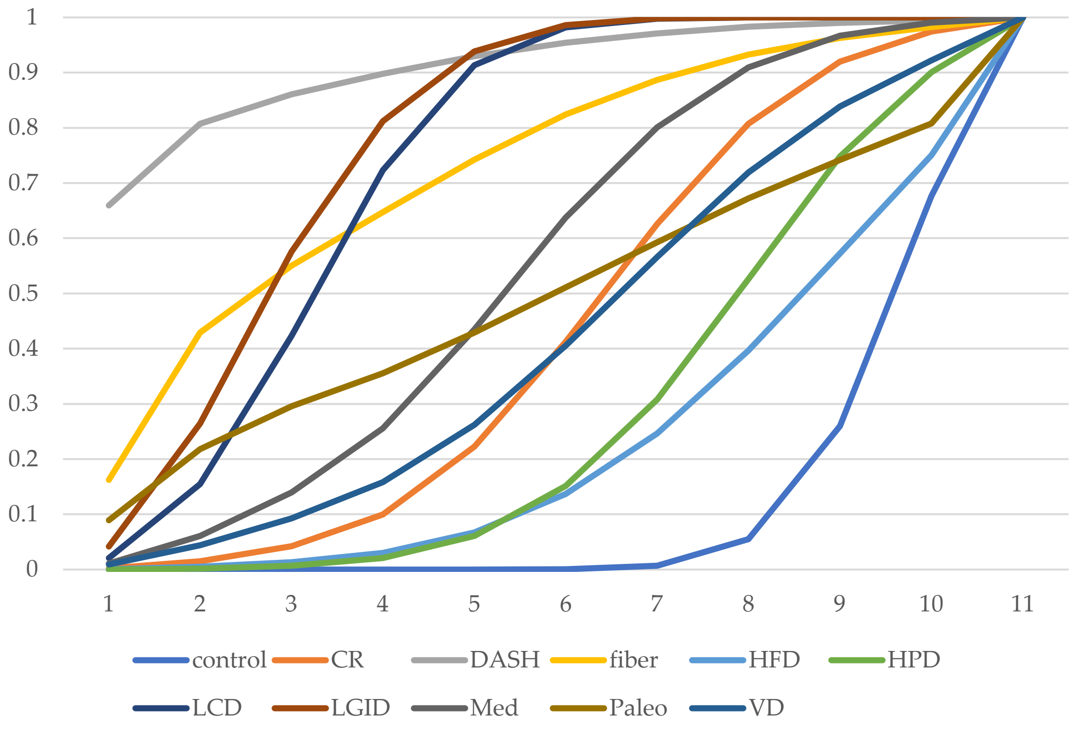

**Figure S8.2** Cumulative ranking curve of $HbA_{1c}$

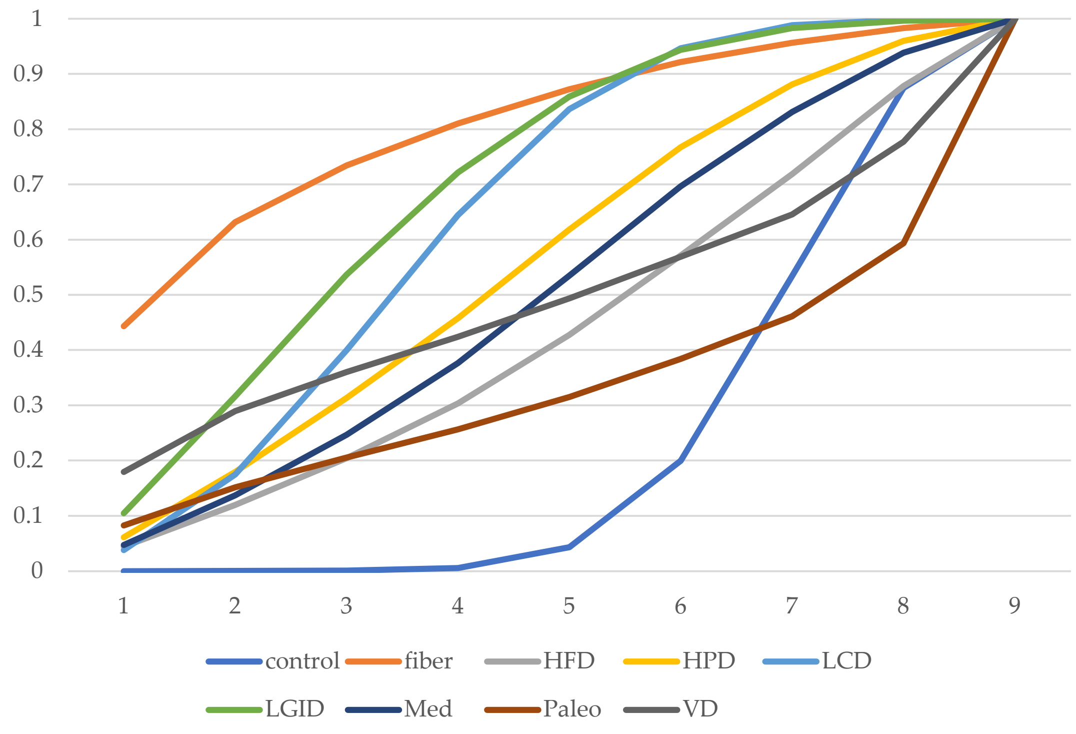

**Figure S8.3** Cumulative ranking curve of FIns

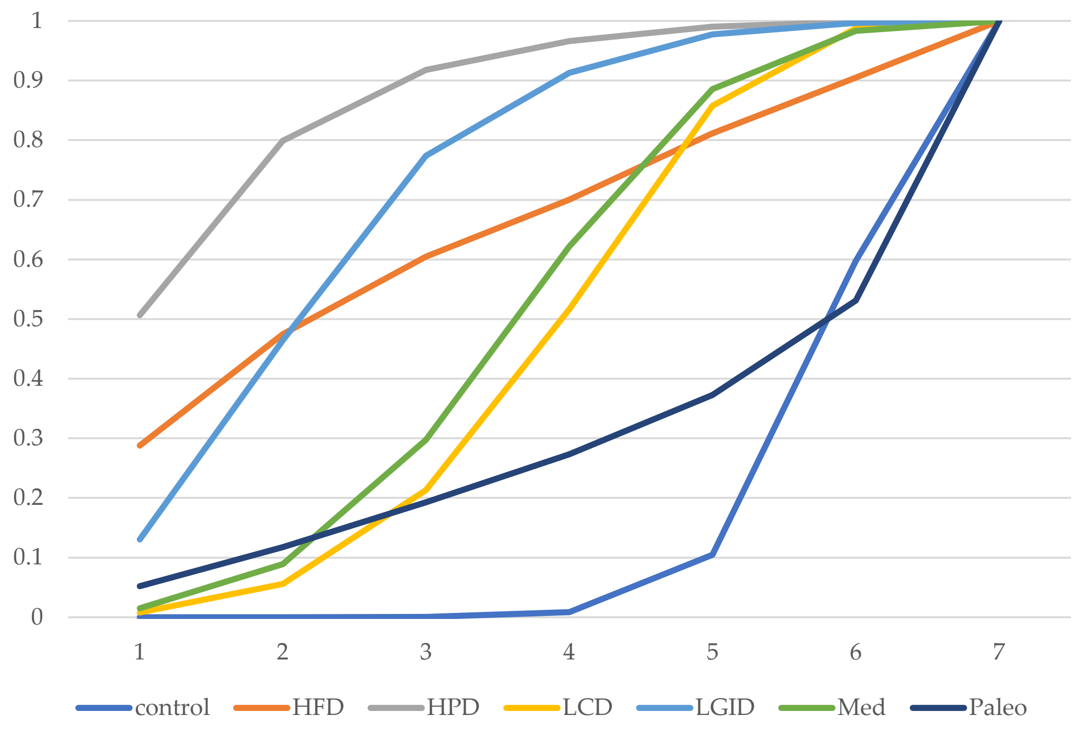

**Figure S8.4** Cumulative ranking curve of IR

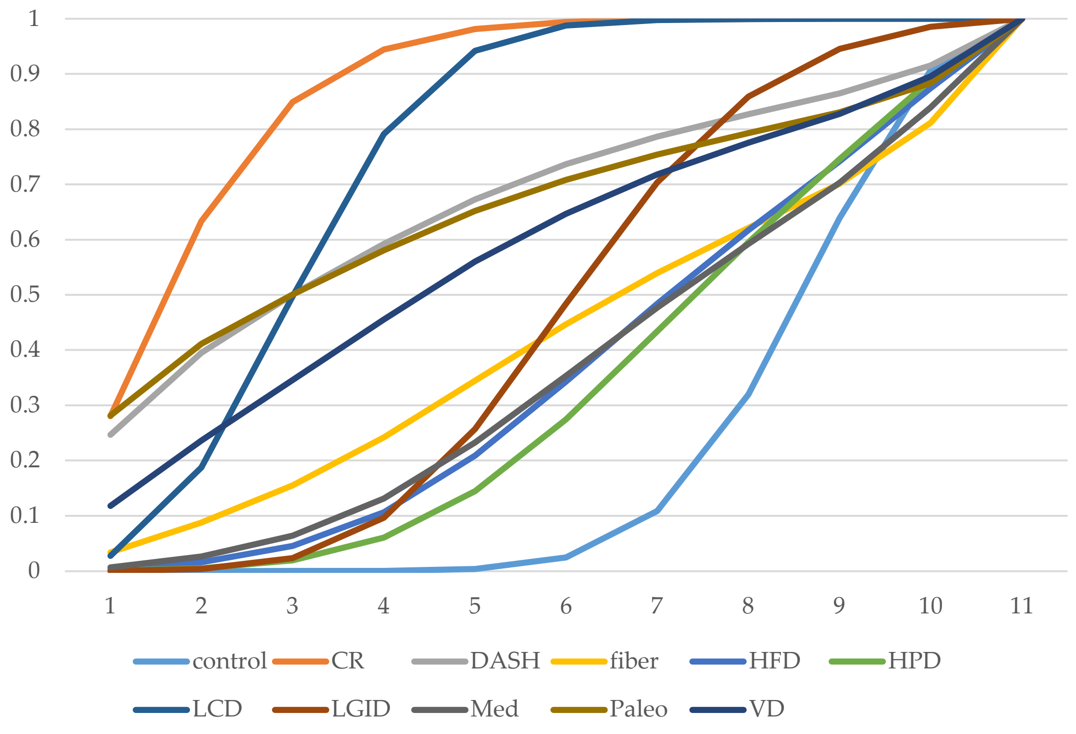

**Figure S8.5** Cumulative ranking curve of weight

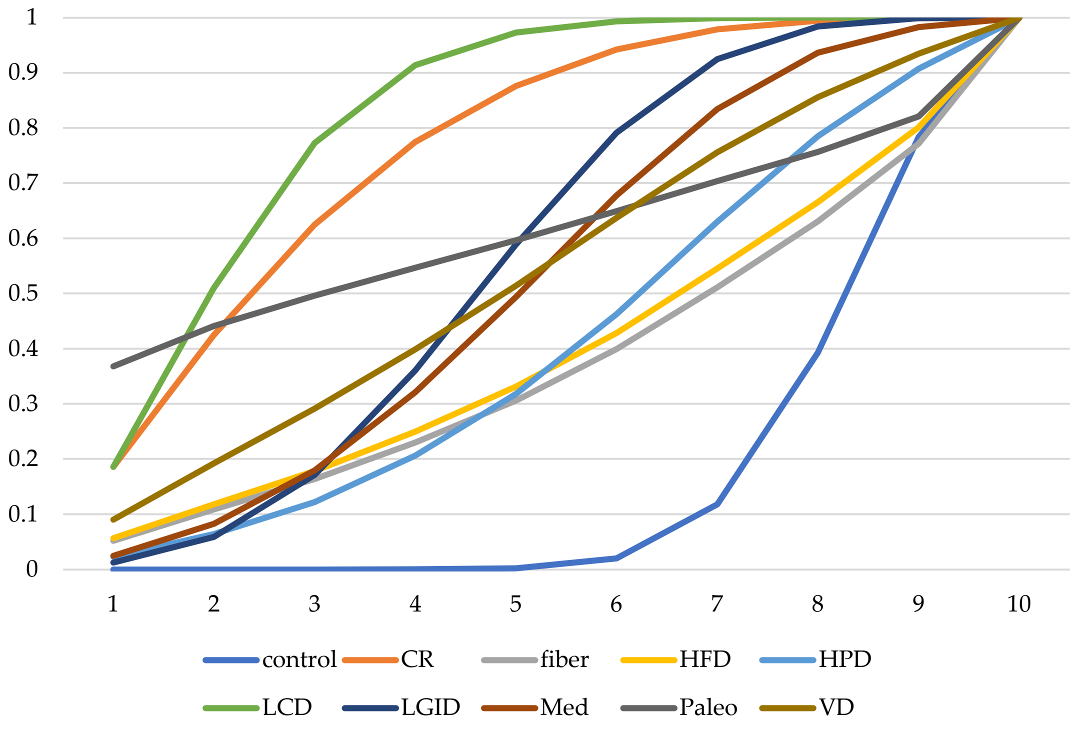

**Figure S8.6** Cumulative ranking curve of BMI

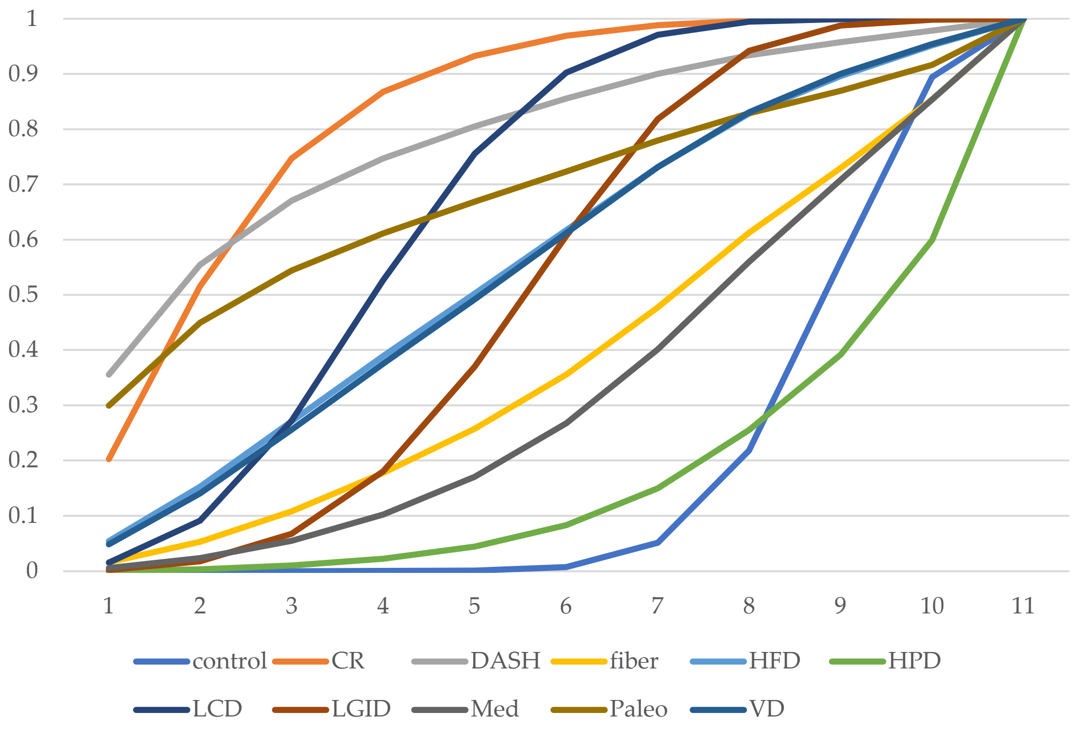

**Figure S8.7** Cumulative ranking curve of WC

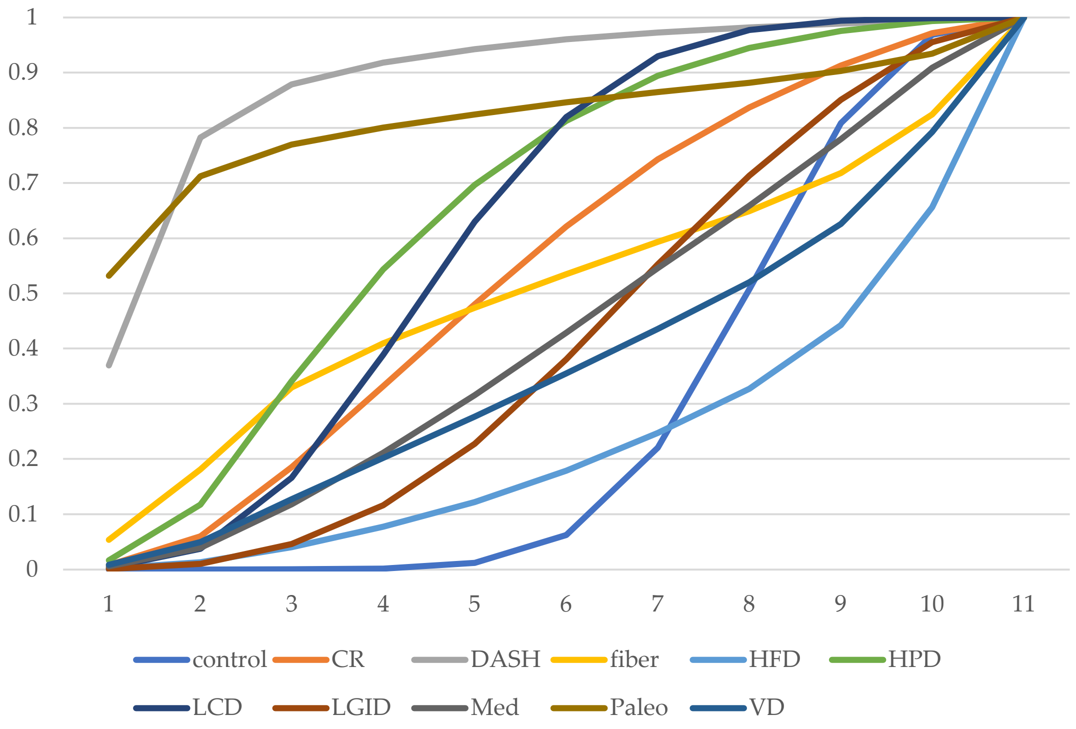

**Figure S8.8** Cumulative ranking curve of SBP

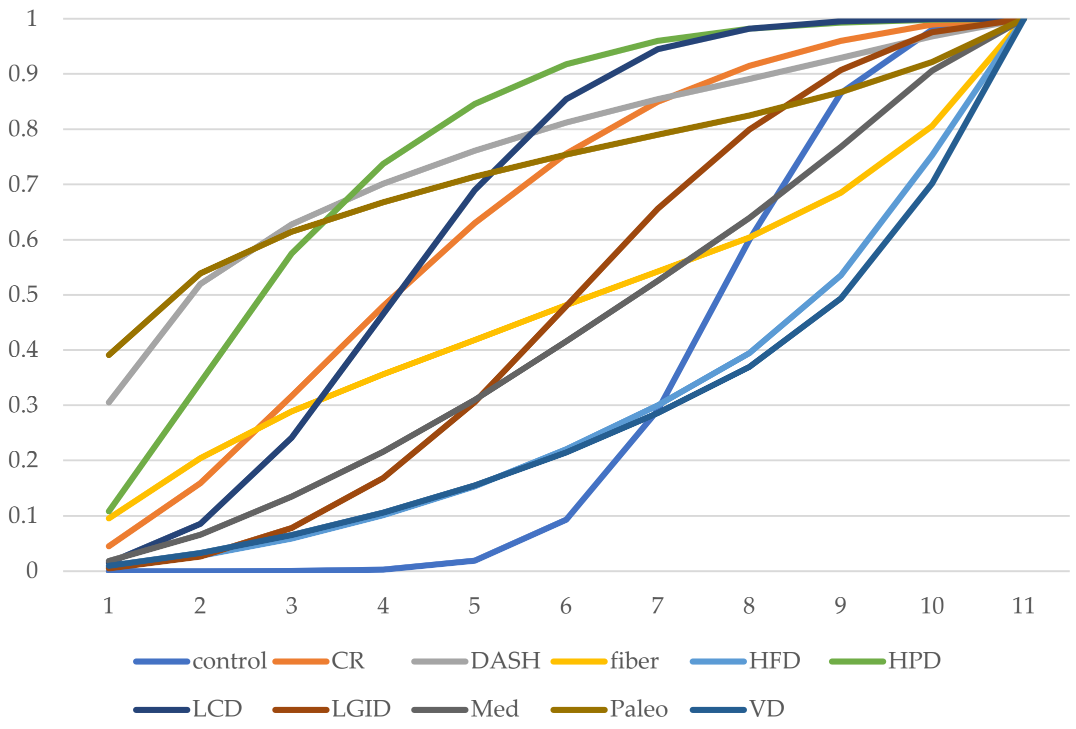

**Figure S8.9** Cumulative ranking curve of DBP

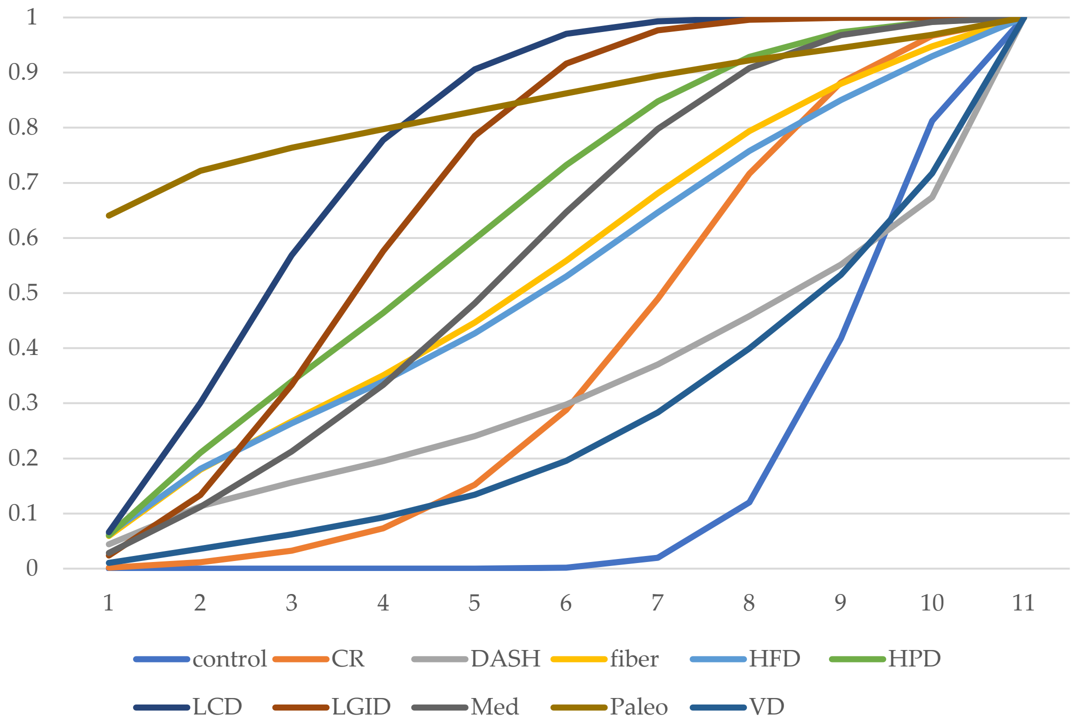

**Figure S8.10** Cumulative ranking curve of TG

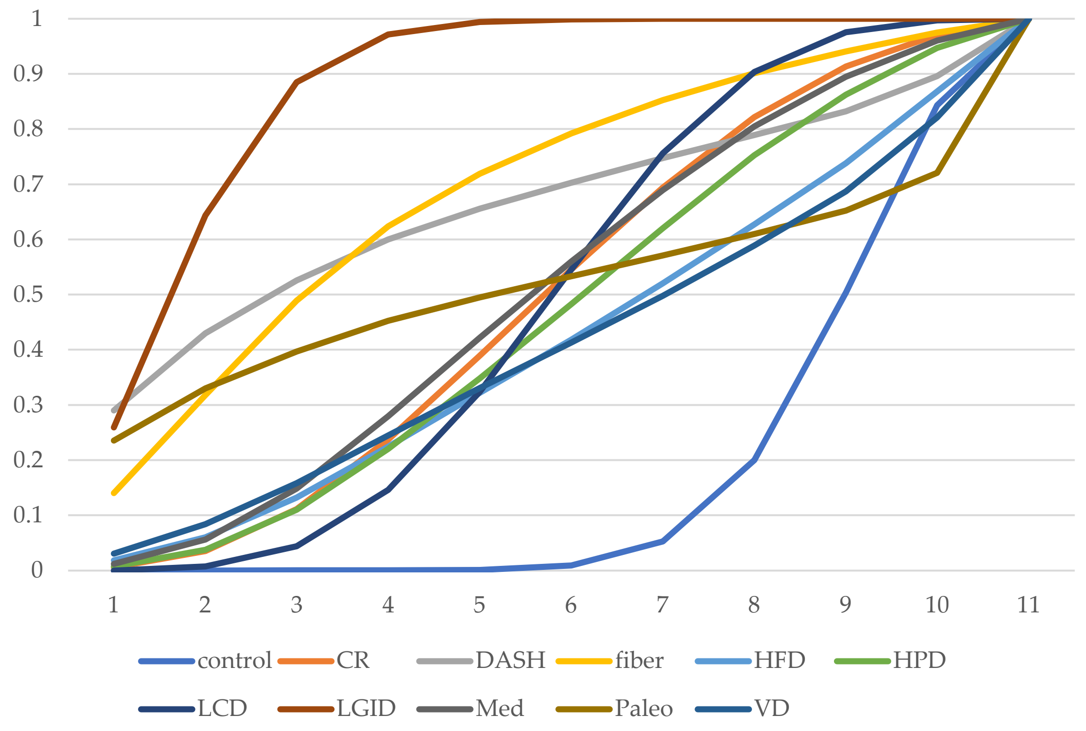

**Figure S8.11** Cumulative ranking curve of TC

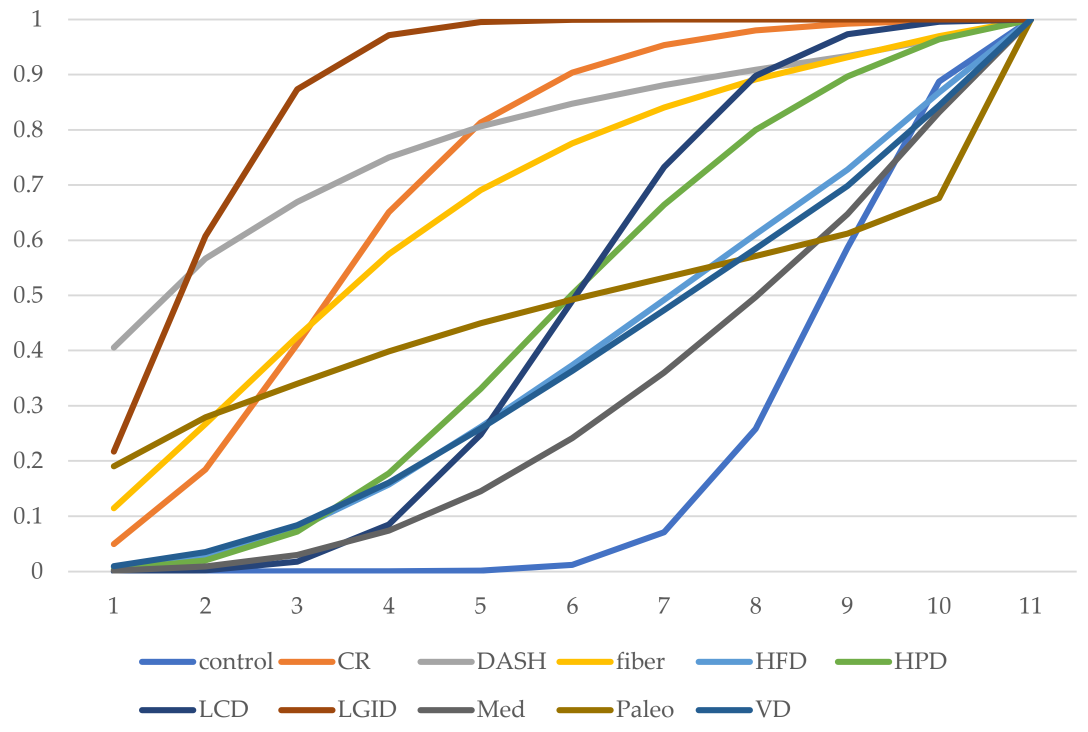

**Figure S8.12** Cumulative ranking curve of LDL

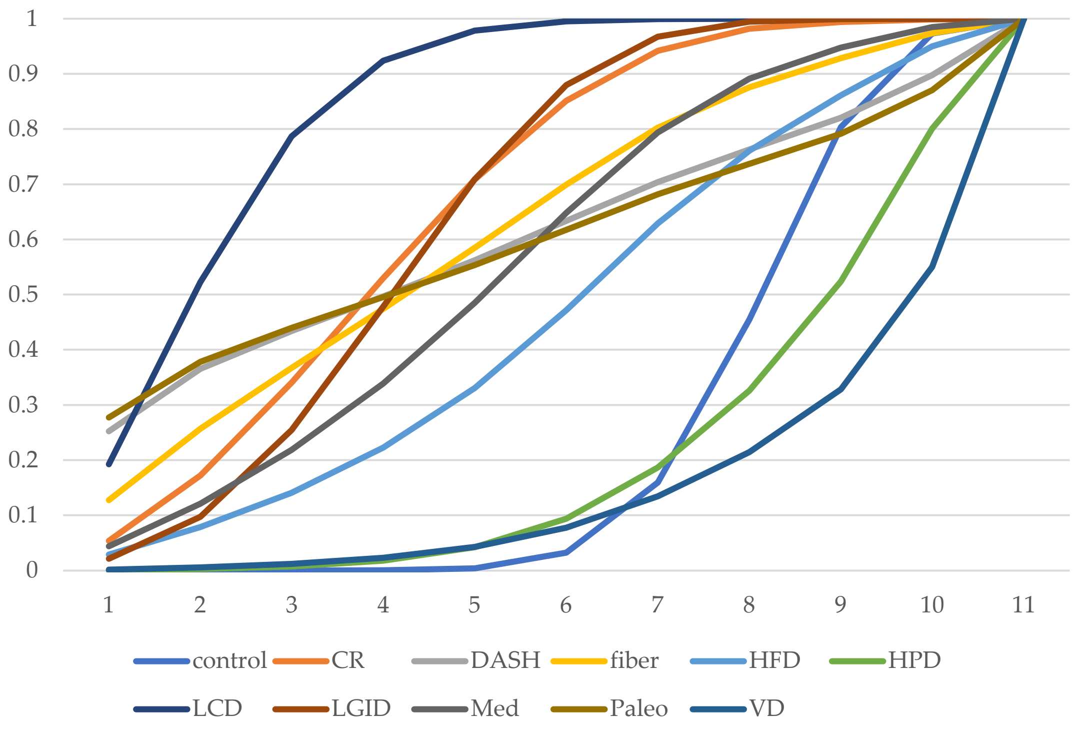

**Figure S8.13** Cumulative ranking curve of HDL

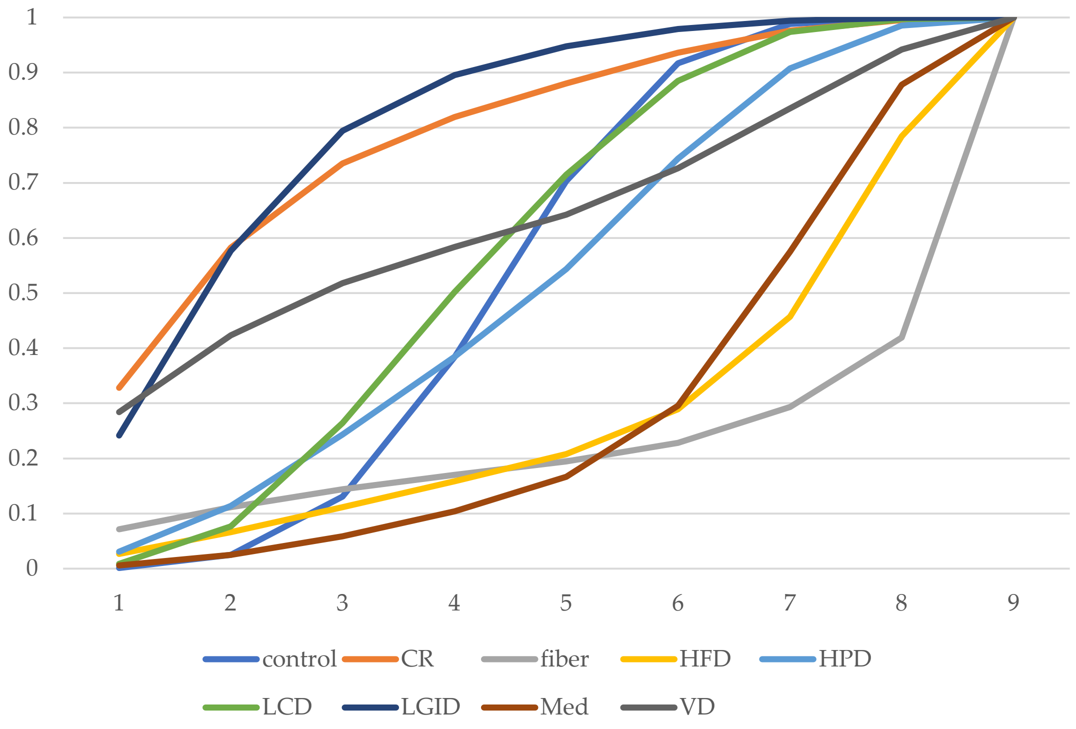

**Figure S8.14** Cumulative ranking curve of Attrition Rate

#### File S9: Forest plots

##### S9.1 Forest plots of FPG (*MD*, mmol/L)

| Study | I^2 | Mean Difference (95% CrI) |
|---|---|---|
| **CR vs control** | | |
| Choi 2013 | | −0.97 (−1.5, −0.42) |
| Fang 2019 | | −2.4 (−3.1, −1.8) |
| Mohammadi 2017 | | −2.5 (−5.5, 0.57) |
| Mollentze 2019 | | 1.0 (−2.2, 4.2) |
| Ruggenenti 2017 | | −1.4 (−2.3, −0.50) |
| Ruggenenti 2022 | | −0.39 (−1.3, 0.55) |
| Uusitupa 1993 | | −0.40 (−1.4, 0.62) |
| Pooled (pair−wise) | 72.3% | −1.1 (−1.8, −0.51) |
| Indirect (back−calculated) | | 0.29 (−0.86, 1.4) |
| Pooled (network) | 77.8% | −0.81 (−1.4, −0.25) |
| **DASH vs control** | | |
| Azadbakht 2011 | | −0.92 (−1.9, 0.084) |
| Pooled (pair−wise) | | −0.91 (−2.5, 0.67) |
| Indirect (back−calculated) | | NA |
| Pooled (network) | | −0.92 (−2.5, 0.69) |
| **fiber vs control** | | |
| Chandalia 2000 | | −0.67 (−2.4, 1.1) |
| Coppell 2010 | | −0.90 (−2., 0.17) |
| Ikem 2007 | | −2. (−3.6, −0.39) |
| Pooled (pair−wise) | 0.0% | −1.2 (−2.2, −0.071) |
| Indirect (back−calculated) | | −2.9 (−7.5, 1.6) |
| Pooled (network) | 0.0% | −1.3 (−2.3, −0.20) |
| **HFD vs control** | | |
| Brehm 2009 | | −0.16 (−1.3, 0.92) |
| Brunerova 2007 | | −0.20 (−1.5, 1.1) |
| Rock 2014 | | −0.94 (−2., 0.067) |
| Shige 2000 | | −0.0045 (−1.9, 1.9) |
| Walker 1995 | | 1.2 (0.22, 2.2) |
| Pooled (pair−wise) | 57.0% | −0.0078 (−0.78, 0.76) |
| Indirect (back−calculated) | | NA |
| Pooled (network) | 57.0% | −0.0048 (−0.79, 0.77) |
| **HPD vs control** | | |
| Brinkworth 2004 | | −1.1 (−2.9, 0.72) |
| Gannon 2003 | | 0.0016 (−0.91, 0.91) |
| Krebs 2012 | | 0.30 (−0.36, 0.96) |
| Luger 2013 | | −2.2 (−4.2, −0.22) |
| Marco−Benedi 2020 | | −0.75 (−1.2, −0.26) |
| Parker 2002 | | −0.40 (−1.3, 0.51) |
| Pedersen 2014 | | 0.20 (−0.80, 1.2) |
| Watson 2016 | | 0.50 (−1.0, 2.0) |
| Pooled (pair−wise) | 47.1% | −0.29 (−0.87, 0.29) |
| Indirect (back−calculated) | | NA |
| Pooled (network) | 47.1% | −0.29 (−0.89, 0.29) |

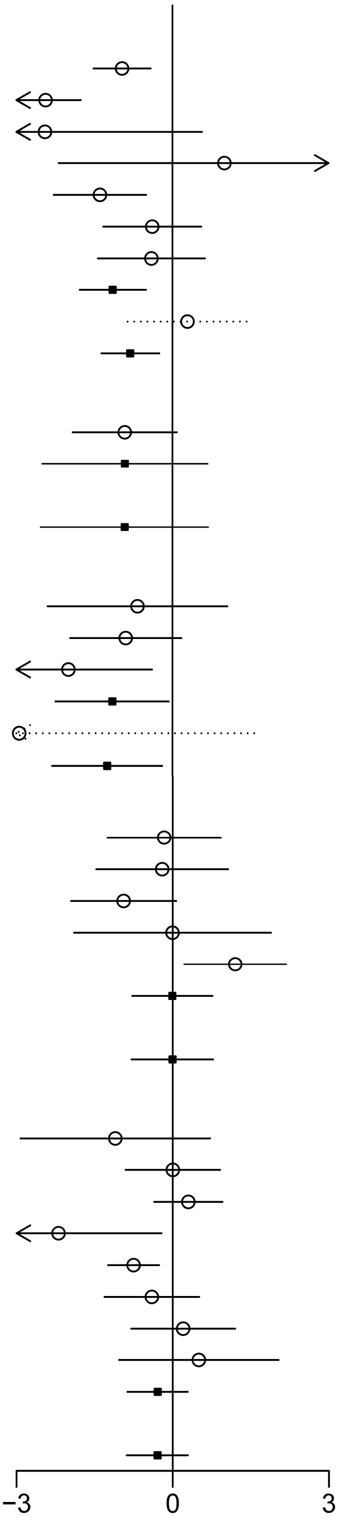

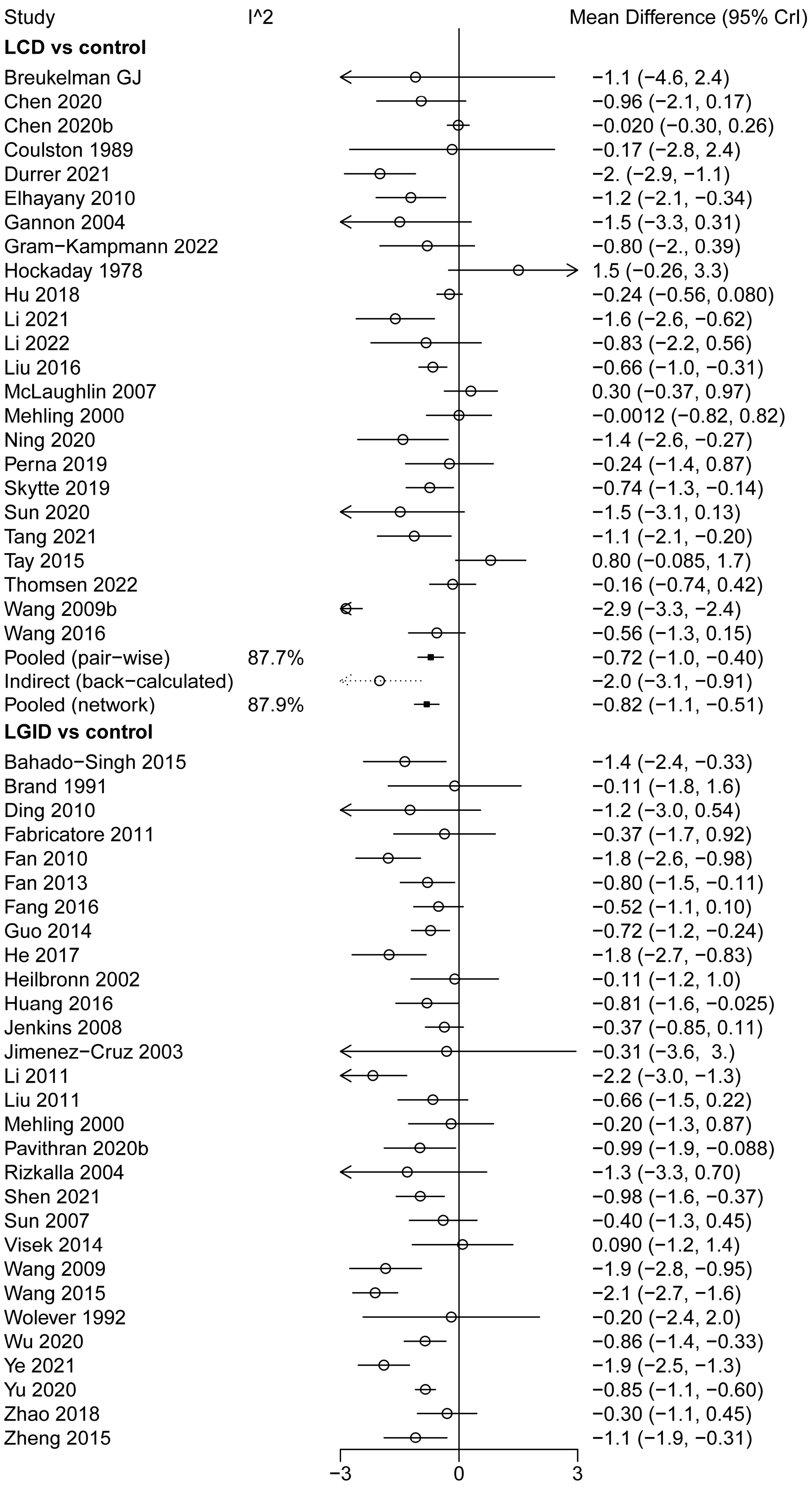
Study
I^2
Mean Difference (95% CrI)
LCD vs control
Breukelman GJ −1.1 (−4.6, 2.4)
Chen 2020 −0.96 (−2.1, 0.17)
Chen 2020b −0.020 (−0.30, 0.26)
Coulston 1989 −0.17 (−2.8, 2.4)
Durrer 2021 −2. (−2.9, −1.1)
Elhayany 2010 −1.2 (−2.1, −0.34)
Gannon 2004 −1.5 (−3.3, 0.31)
Gram−Kampmann 2022 −0.80 (−2., 0.39)
Hockaday 1978 1.5 (−0.26, 3.3)
Hu 2018 −0.24 (−0.56, 0.080)
Li 2021 −1.6 (−2.6, −0.62)
Li 2022 −0.83 (−2.2, 0.56)
Liu 2016 −0.66 (−1.0, −0.31)
McLaughlin 2007 0.30 (−0.37, 0.97)
Mehling 2000 −0.0012 (−0.82, 0.82)
Ning 2020 −1.4 (−2.6, −0.27)
Perna 2019 −0.24 (−1.4, 0.87)
Skytte 2019 −0.74 (−1.3, −0.14)
Sun 2020 −1.5 (−3.1, 0.13)
Tang 2021 −1.1 (−2.1, −0.20)
Tay 2015 0.80 (−0.085, 1.7)
Thomsen 2022 −0.16 (−0.74, 0.42)
Wang 2009b −2.9 (−3.3, −2.4)
Wang 2016 −0.56 (−1.3, 0.15)
Pooled (pair−wise) 87.7% −0.72 (−1.0, −0.40)
Indirect (back−calculated) −2.0 (−3.1, −0.91)
Pooled (network) 87.9% −0.82 (−1.1, −0.51)
LGID vs control
Bahado−Singh 2015 −1.4 (−2.4, −0.33)
Brand 1991 −0.11 (−1.8, 1.6)
Ding 2010 −1.2 (−3.0, 0.54)
Fabricatore 2011 −0.37 (−1.7, 0.92)
Fan 2010 −1.8 (−2.6, −0.98)
Fan 2013 −0.80 (−1.5, −0.11)
Fang 2016 −0.52 (−1.1, 0.10)
Guo 2014 −0.72 (−1.2, −0.24)
He 2017 −1.8 (−2.7, −0.83)
Heilbronn 2002 −0.11 (−1.2, 1.0)
Huang 2016 −0.81 (−1.6, −0.025)
Jenkins 2008 −0.37 (−0.85, 0.11)
Jimenez−Cruz 2003 −0.31 (−3.6, 3.)
Li 2011 −2.2 (−3.0, −1.3)
Liu 2011 −0.66 (−1.5, 0.22)
Mehling 2000 −0.20 (−1.3, 0.87)
Pavithran 2020b −0.99 (−1.9, −0.088)
Rizkalla 2004 −1.3 (−3.3, 0.70)
Shen 2021 −0.98 (−1.6, −0.37)
Sun 2007 −0.40 (−1.3, 0.45)
Visek 2014 0.090 (−1.2, 1.4)
Wang 2009 −1.9 (−2.8, −0.95)
Wang 2015 −2.1 (−2.7, −1.6)
Wolever 1992 −0.20 (−2.4, 2.0)
Wu 2020 −0.86 (−1.4, −0.33)
Ye 2021 −1.9 (−2.5, −1.3)
Yu 2020 −0.85 (−1.1, −0.60)
Zhao 2018 −0.30 (−1.1, 0.45)
Zheng 2015 −1.1 (−1.9, −0.31)
−3
0
3

| Study | I^2 | Mean Difference (95% CrI) |
|---|---|---|
| **LGID vs control** | | |
| Zhou 2011 | | −0.23 (−1.3, 0.80) |
| Pooled (pair−wise) | 58.3% | −0.94 (−1.2, −0.65) |
| Indirect (back−calculated) | | NA |
| Pooled (network) | 58.3% | −0.94 (−1.2, −0.65) |
| **Med vs control** | | |
| Elhayany 2010 | | −0.43 (−1.3, 0.40) |
| Esposito 2009 | | −0.90 (−1.2, −0.64) |
| Itsiopoulos 2011 | | −0.40 (−2.3, 1.5) |
| Lasa 2014 | | −0.063 (−0.72, 0.60) |
| Xue 2020 | | −0.18 (−1.1, 0.71) |
| Zahedi 2021 | | −1.0 (−2.1, 0.077) |
| Pooled (pair−wise) | 58.2% | −0.51 (−1.1, 0.11) |
| Indirect (back−calculated) | | NA |
| Pooled (network) | 65.2% | −0.45 (−1.1, 0.15) |
| **Paleo vs control** | | |
| Jonsson 2009 | | −0.50 (−1.6, 0.59) |
| Pooled (pair−wise) | | −0.50 (−2.1, 1.2) |
| Indirect (back−calculated) | | NA |
| Pooled (network) | | −0.50 (−2.2, 1.2) |
| **VD vs control** | | |
| Barnard 2006−9 | | −0.31 (−1.5, 0.87) |
| Kahleova 2011 | | −0.44 (−1.7, 0.78) |
| Lee 2016 | | −0.73 (−1.7, 0.25) |
| Nicholson 1999 | | −1.8 (−4.3, 0.78) |
| Pooled (pair−wise) | 0.0% | −0.63 (−1.5, 0.27) |
| Indirect (back−calculated) | | NA |
| Pooled (network) | 0.0% | −0.64 (−1.6, 0.28) |
| **LCD vs CR** | | |
| Iqbal 2010 | | 0.14 (−1.4, 1.6) |
| Liu 2020 | | −1.3 (−2.0, −0.53) |
| Yamada 2014 | | −1.2 (−3.1, 0.63) |
| Pooled (pair−wise) | 34.3% | −0.88 (−1.9, 0.15) |
| Indirect (back−calculated) | | 0.44 (−0.29, 1.2) |
| Pooled (network) | 78.8% | −0.0049 (−0.60, 0.59) |
| **LCD vs fiber** | | |
| Lousley 1984 | | 1.6 (−1.9, 5.1) |
| Pooled (pair−wise) | | 1.6 (−2.1, 5.3) |
| Indirect (back−calculated) | | 0.33 (−0.81, 1.5) |
| Pooled (network) | 0.0% | 0.44 (−0.66, 1.5) |
| **LGID vs LCD** | | |
| Mehling 2000 | | −0.20 (−1.0, 0.63) |
| Westman 2008 | | 0.22 (−1.9, 2.3) |
| Pooled (pair−wise) | 0.0% | −0.089 (−1.3, 1.2) |
| Indirect (back−calculated) | | −0.12 (−0.56, 0.31) |
| Pooled (network) | 0.0% | −0.12 (−0.53, 0.29) |
| **Med vs LCD** | | |
| Elhayany 2010 | | 0.79 (0.046, 1.5) |
| Pooled (pair−wise) | | 0.79 (−0.65, 2.2) |
| Indirect (back−calculated) | | 0.25 (−0.50, 0.99) |
| Pooled (network) | 26.3% | 0.36 (−0.30, 1.0) |

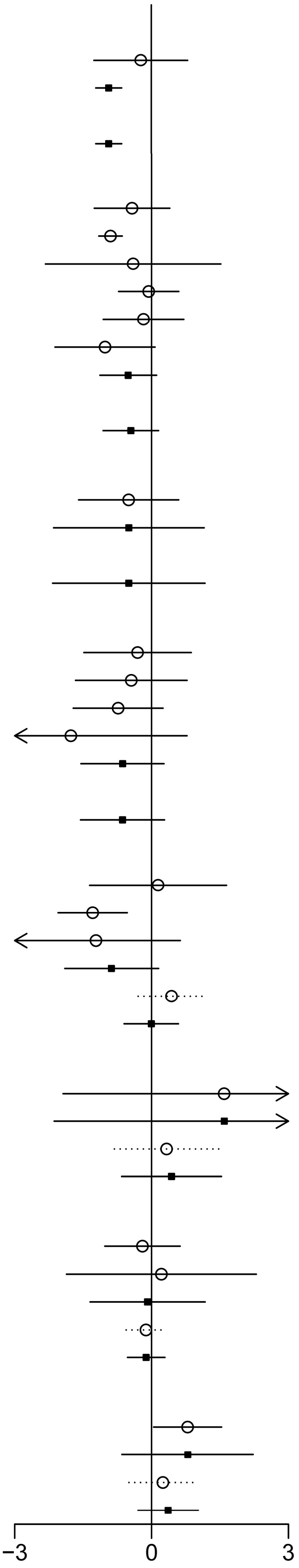

**S9.2 Forest plots of HbA1c (*MD*, %)**

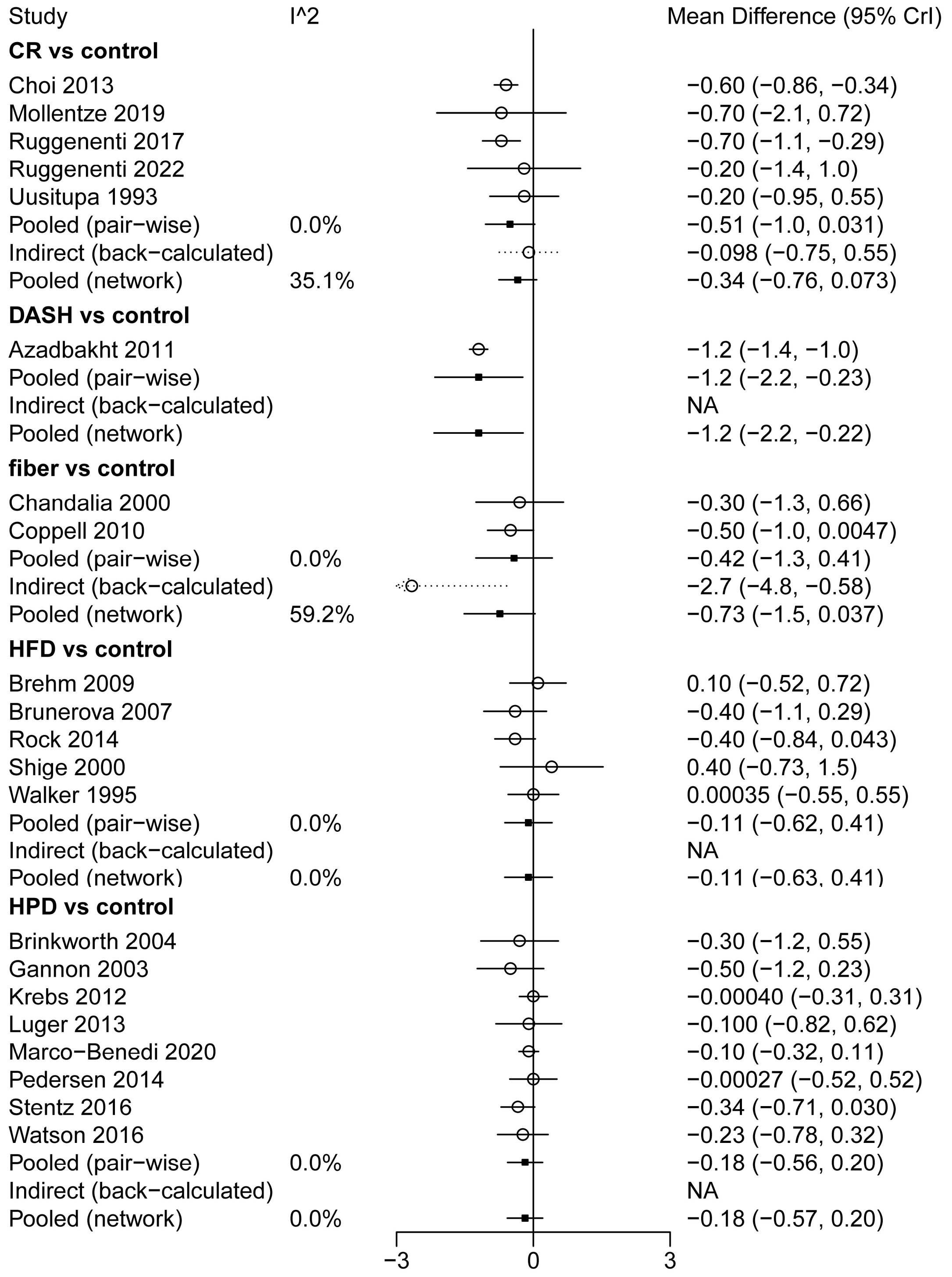

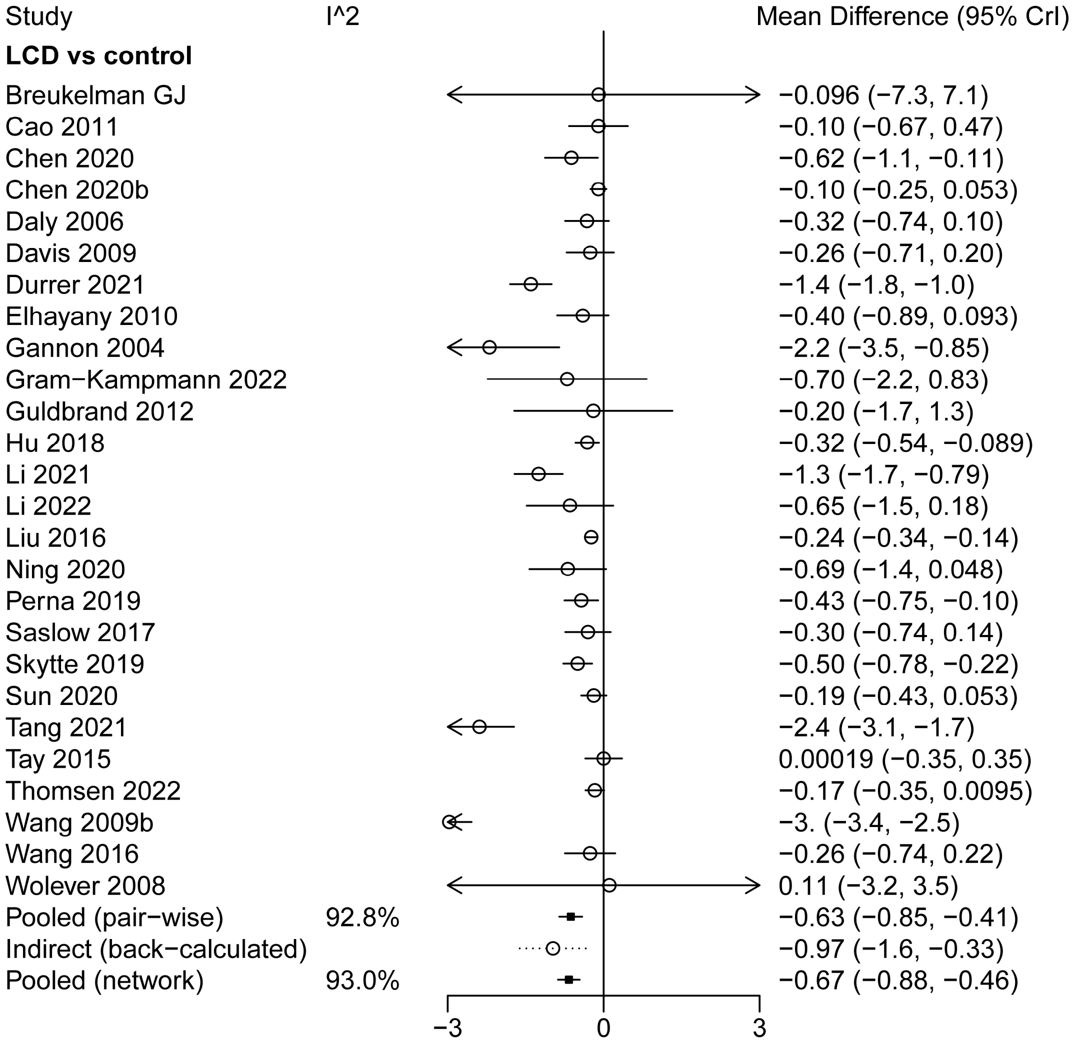
Study
I^2
Mean Difference (95% CrI)
LCD vs control
Breukelman GJ −0.096 (−7.3, 7.1)
Cao 2011 −0.10 (−0.67, 0.47)
Chen 2020 −0.62 (−1.1, −0.11)
Chen 2020b −0.10 (−0.25, 0.053)
Daly 2006 −0.32 (−0.74, 0.10)
Davis 2009 −0.26 (−0.71, 0.20)
Durrer 2021 −1.4 (−1.8, −1.0)
Elhayany 2010 −0.40 (−0.89, 0.093)
Gannon 2004 −2.2 (−3.5, −0.85)
Gram−Kampmann 2022 −0.70 (−2.2, 0.83)
Guldbrand 2012 −0.20 (−1.7, 1.3)
Hu 2018 −0.32 (−0.54, −0.089)
Li 2021 −1.3 (−1.7, −0.79)
Li 2022 −0.65 (−1.5, 0.18)
Liu 2016 −0.24 (−0.34, −0.14)
Ning 2020 −0.69 (−1.4, 0.048)
Perna 2019 −0.43 (−0.75, −0.10)
Saslow 2017 −0.30 (−0.74, 0.14)
Skytte 2019 −0.50 (−0.78, −0.22)
Sun 2020 −0.19 (−0.43, 0.053)
Tang 2021 −2.4 (−3.1, −1.7)
Tay 2015 0.00019 (−0.35, 0.35)
Thomsen 2022 −0.17 (−0.35, 0.0095)
Wang 2009b −3. (−3.4, −2.5)
Wang 2016 −0.26 (−0.74, 0.22)
Wolever 2008 0.11 (−3.2, 3.5)
Pooled (pair−wise) 92.8% −0.63 (−0.85, −0.41)
Indirect (back−calculated) −0.97 (−1.6, −0.33)
Pooled (network) 93.0% −0.67 (−0.88, −0.46)
−3
0
3

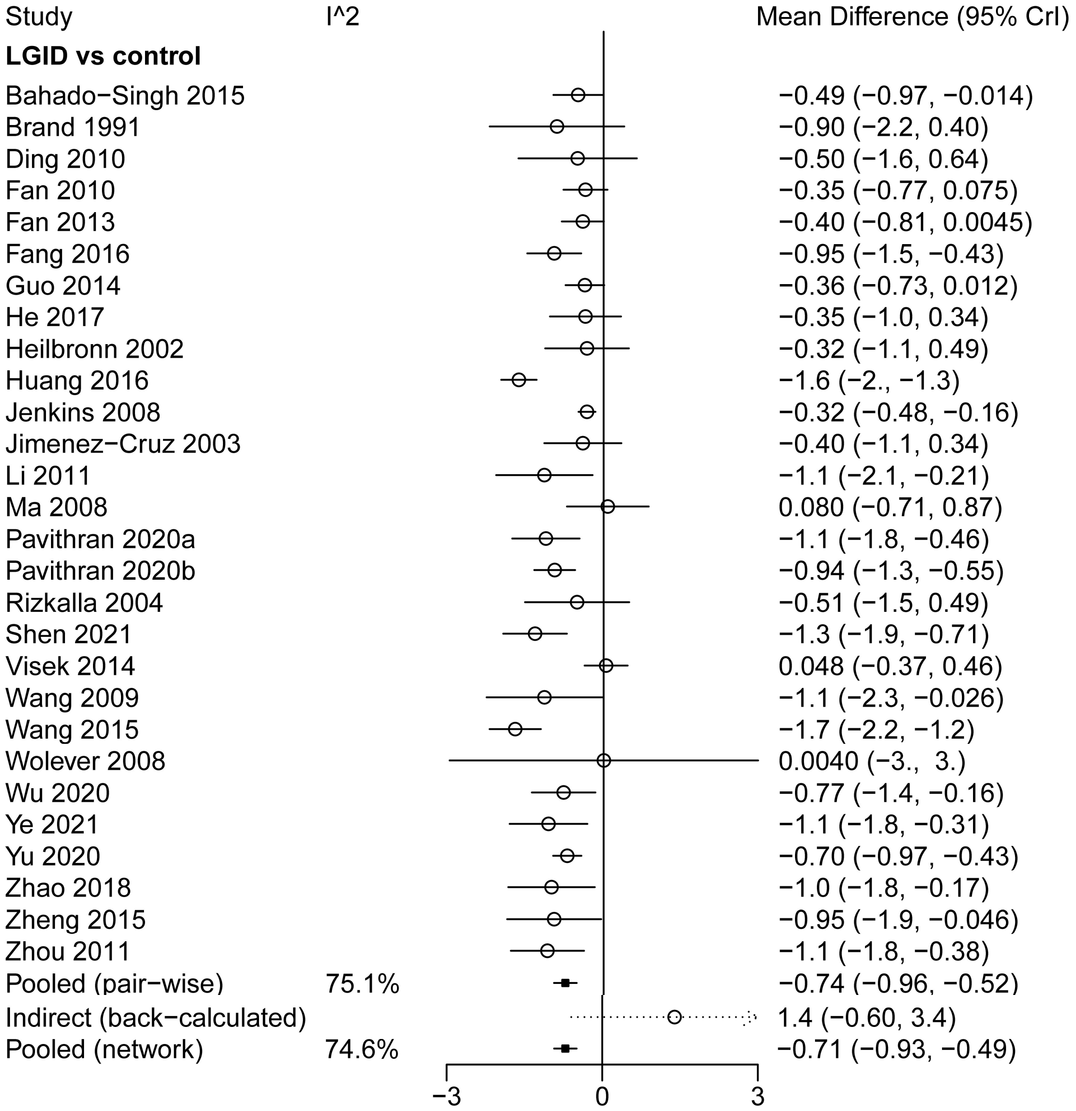
Study
I^2
Mean Difference (95% CrI)
LGID vs control
Bahado−Singh 2015 −0.49 (−0.97, −0.014)
Brand 1991 −0.90 (−2.2, 0.40)
Ding 2010 −0.50 (−1.6, 0.64)
Fan 2010 −0.35 (−0.77, 0.075)
Fan 2013 −0.40 (−0.81, 0.0045)
Fang 2016 −0.95 (−1.5, −0.43)
Guo 2014 −0.36 (−0.73, 0.012)
He 2017 −0.35 (−1.0, 0.34)
Heilbronn 2002 −0.32 (−1.1, 0.49)
Huang 2016 −1.6 (−2., −1.3)
Jenkins 2008 −0.32 (−0.48, −0.16)
Jimenez−Cruz 2003 −0.40 (−1.1, 0.34)
Li 2011 −1.1 (−2.1, −0.21)
Ma 2008 0.080 (−0.71, 0.87)
Pavithran 2020a −1.1 (−1.8, −0.46)
Pavithran 2020b −0.94 (−1.3, −0.55)
Rizkalla 2004 −0.51 (−1.5, 0.49)
Shen 2021 −1.3 (−1.9, −0.71)
Visek 2014 0.048 (−0.37, 0.46)
Wang 2009 −1.1 (−2.3, −0.026)
Wang 2015 −1.7 (−2.2, −1.2)
Wolever 2008 0.0040 (−3., 3.)
Wu 2020 −0.77 (−1.4, −0.16)
Ye 2021 −1.1 (−1.8, −0.31)
Yu 2020 −0.70 (−0.97, −0.43)
Zhao 2018 −1.0 (−1.8, −0.17)
Zheng 2015 −0.95 (−1.9, −0.046)
Zhou 2011 −1.1 (−1.8, −0.38)
Pooled (pair−wise) 75.1% −0.74 (−0.96, −0.52)
Indirect (back−calculated) 1.4 (−0.60, 3.4)
Pooled (network) 74.6% −0.71 (−0.93, −0.49)
−3
0
3

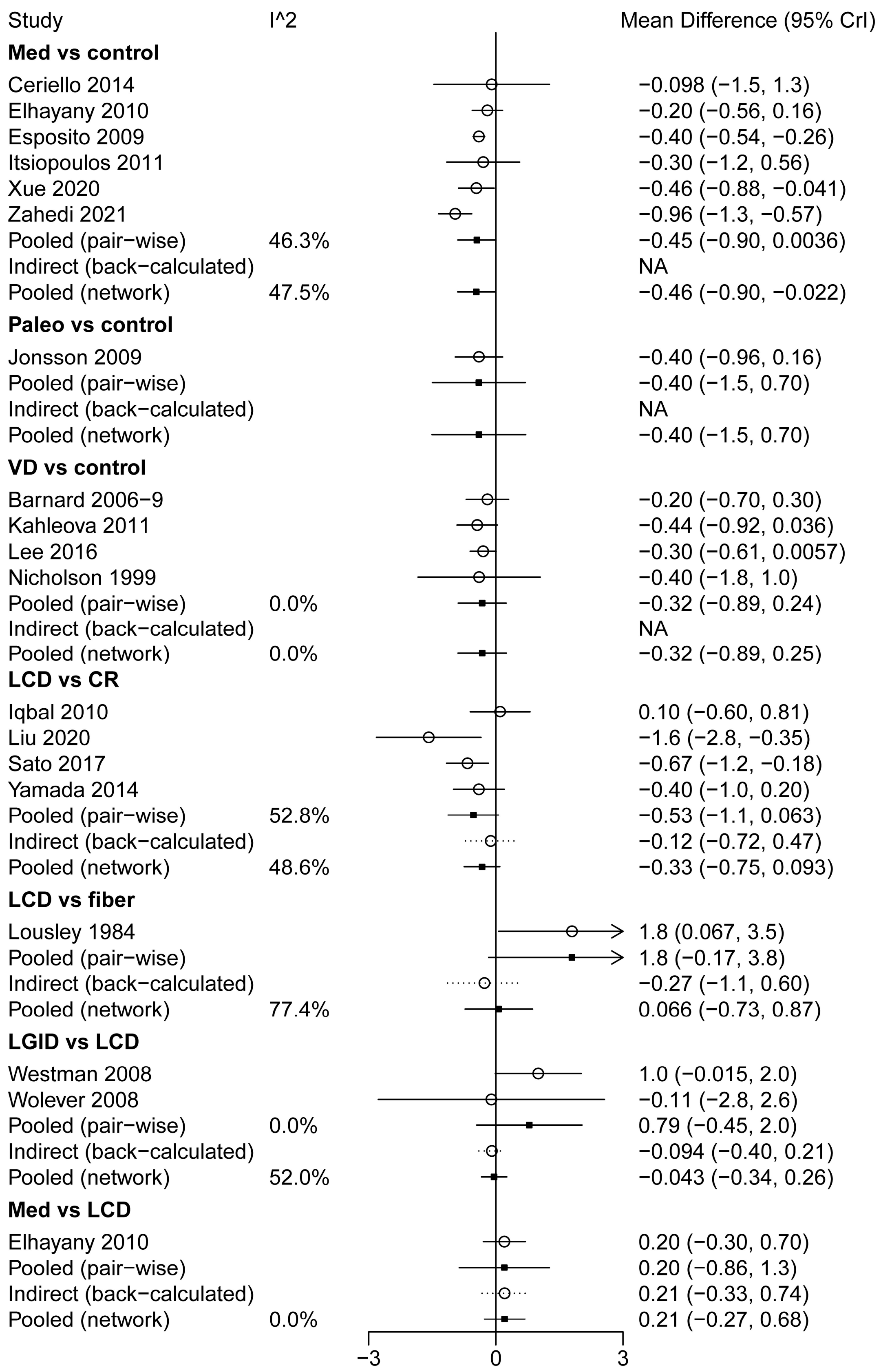

Study
I^2
Mean Difference (95% CrI)
Med vs control
Ceriello 2014 −0.098 (−1.5, 1.3)
Elhayany 2010 −0.20 (−0.56, 0.16)
Esposito 2009 −0.40 (−0.54, −0.26)
Itsiopoulos 2011 −0.30 (−1.2, 0.56)
Xue 2020 −0.46 (−0.88, −0.041)
Zahedi 2021 −0.96 (−1.3, −0.57)
Pooled (pair−wise) 46.3% −0.45 (−0.90, 0.0036)
Indirect (back−calculated) NA
Pooled (network) 47.5% −0.46 (−0.90, −0.022)
Paleo vs control
Jonsson 2009 −0.40 (−0.96, 0.16)
Pooled (pair−wise) −0.40 (−1.5, 0.70)
Indirect (back−calculated) NA
Pooled (network) −0.40 (−1.5, 0.70)
VD vs control
Barnard 2006−9 −0.20 (−0.70, 0.30)
Kahleova 2011 −0.44 (−0.92, 0.036)
Lee 2016 −0.30 (−0.61, 0.0057)
Nicholson 1999 −0.40 (−1.8, 1.0)
Pooled (pair−wise) 0.0% −0.32 (−0.89, 0.24)
Indirect (back−calculated) NA
Pooled (network) 0.0% −0.32 (−0.89, 0.25)
LCD vs CR
Iqbal 2010 0.10 (−0.60, 0.81)
Liu 2020 −1.6 (−2.8, −0.35)
Sato 2017 −0.67 (−1.2, −0.18)
Yamada 2014 −0.40 (−1.0, 0.20)
Pooled (pair−wise) 52.8% −0.53 (−1.1, 0.063)
Indirect (back−calculated) −0.12 (−0.72, 0.47)
Pooled (network) 48.6% −0.33 (−0.75, 0.093)
LCD vs fiber
Lousley 1984 1.8 (0.067, 3.5)
Pooled (pair−wise) 1.8 (−0.17, 3.8)
Indirect (back−calculated) −0.27 (−1.1, 0.60)
Pooled (network) 77.4% 0.066 (−0.73, 0.87)
LGID vs LCD
Westman 2008 1.0 (−0.015, 2.0)
Wolever 2008 −0.11 (−2.8, 2.6)
Pooled (pair−wise) 0.0% 0.79 (−0.45, 2.0)
Indirect (back−calculated) −0.094 (−0.40, 0.21)
Pooled (network) 52.0% −0.043 (−0.34, 0.26)
Med vs LCD
Elhayany 2010 0.20 (−0.30, 0.70)
Pooled (pair−wise) 0.20 (−0.86, 1.3)
Indirect (back−calculated) 0.21 (−0.33, 0.74)
Pooled (network) 0.0% 0.21 (−0.27, 0.68)
−3
0
3

**S9.3 Forest plots of FIns (*PMD*)**

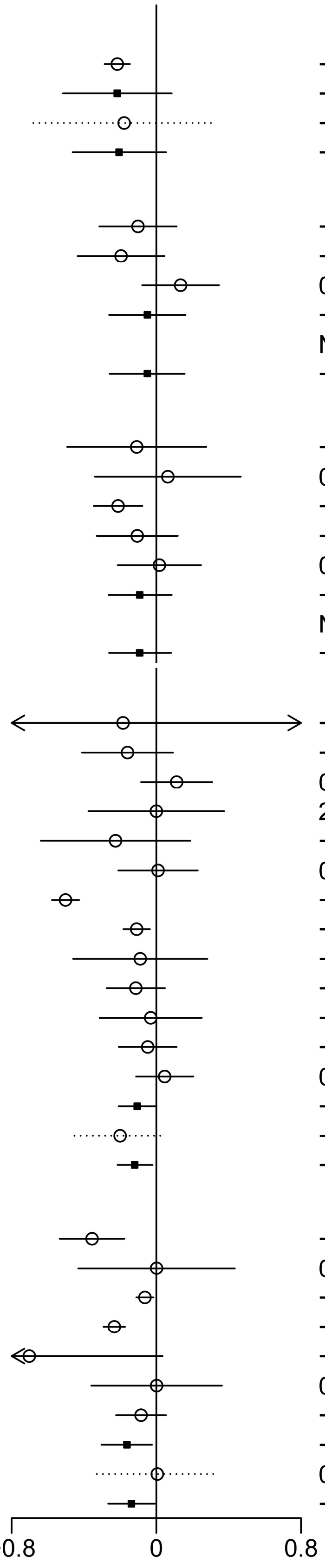

| Study | I^2 | PMD (95% CrI) |
|---|---|---|
| **fiber vs control** | | |
| Chandalia 2000 | | −0.22 (−0.29, −0.15) |
| Pooled (pair−wise) | | −0.22 (−0.52, 0.085) |
| Indirect (back−calculated) | | −0.18 (−0.68, 0.32) |
| Pooled (network) | 0.0% | −0.21 (−0.46, 0.054) |
| **HFD vs control** | | |
| Brehm 2009 | | −0.10 (−0.32, 0.11) |
| Rock 2014 | | −0.19 (−0.44, 0.045) |
| Walker 1995 | | 0.13 (−0.078, 0.35) |
| Pooled (pair−wise) | 55.6% | −0.049 (−0.26, 0.16) |
| Indirect (back−calculated) | | NA |
| Pooled (network) | 55.6% | −0.050 (−0.26, 0.16) |
| **HPD vs control** | | |
| Brinkworth 2004 | | −0.11 (−0.49, 0.28) |
| Gannon 2003 | | 0.064 (−0.34, 0.47) |
| Marco−Benedi 2020 | | −0.21 (−0.35, −0.077) |
| Parker 2002 | | −0.11 (−0.33, 0.12) |
| Watson 2016 | | 0.017 (−0.21, 0.25) |
| Pooled (pair−wise) | 11.2% | −0.092 (−0.26, 0.085) |
| Indirect (back−calculated) | | NA |
| Pooled (network) | 11.1% | −0.092 (−0.26, 0.082) |
| **LCD vs control** | | |
| Breukelman GJ | | −0.18 (−3.4, 3.) |
| Coulston 1989 | | −0.16 (−0.41, 0.092) |
| Elhayany 2010 | | 0.11 (−0.084, 0.31) |
| Gannon 2004 | | 2.5e−05 (−0.38, 0.38) |
| Gram−Kampmann 2022 | | −0.23 (−0.64, 0.19) |
| Hockaday 1978 | | 0.0094 (−0.21, 0.23) |
| Li 2022 | | −0.50 (−0.58, −0.43) |
| Liu 2016 | | −0.11 (−0.18, −0.035) |
| Saslow 2017 | | −0.089 (−0.46, 0.28) |
| Skytte 2019 | | −0.11 (−0.27, 0.048) |
| Sun 2020 | | −0.031 (−0.31, 0.25) |
| Tay 2015 | | −0.048 (−0.21, 0.11) |
| Thomsen 2022 | | 0.046 (−0.11, 0.20) |
| Pooled (pair−wise) | 89.8% | −0.11 (−0.21, 0.0015) |
| Indirect (back−calculated) | | −0.20 (−0.45, 0.052) |
| Pooled (network) | 88.5% | −0.12 (−0.21, −0.021) |
| **LGID vs control** | | |
| Bahado−Singh 2015 | | −0.36 (−0.53, −0.18) |
| Fabricatore 2011 | | 0.0014 (−0.43, 0.43) |
| Guo 2014 | | −0.063 (−0.11, −0.016) |
| Liu 2011 | | −0.23 (−0.29, −0.17) |
| Pavithran 2020b | | −0.70 (−1.4, 0.034) |
| Rizkalla 2004 | | 0.0016 (−0.36, 0.36) |
| Sun 2007 | | −0.085 (−0.22, 0.054) |
| Pooled (pair−wise) | 80.8% | −0.16 (−0.30, −0.024) |
| Indirect (back−calculated) | | 0.0054 (−0.33, 0.34) |
| Pooled (network) | 76.1% | −0.14 (−0.27, −0.0086) |

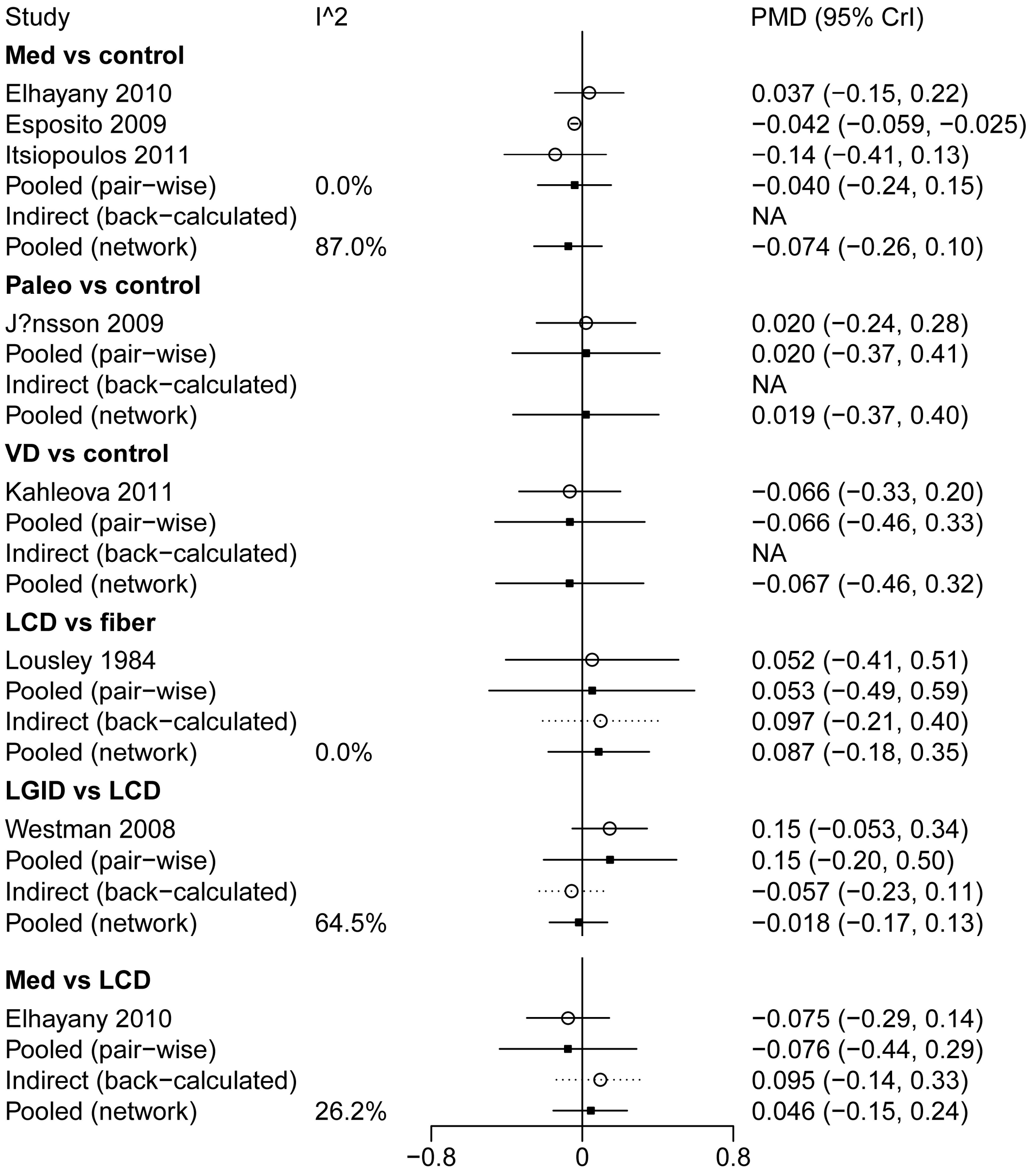
Study
I^2
PMD (95% CrI)
Med vs control
Elhayany 2010 0.037 (−0.15, 0.22)
Esposito 2009 −0.042 (−0.059, −0.025)
Itsiopoulos 2011 −0.14 (−0.41, 0.13)
Pooled (pair−wise) 0.0% −0.040 (−0.24, 0.15)
Indirect (back−calculated) NA
Pooled (network) 87.0% −0.074 (−0.26, 0.10)
Paleo vs control
J?nsson 2009 0.020 (−0.24, 0.28)
Pooled (pair−wise) 0.020 (−0.37, 0.41)
Indirect (back−calculated) NA
Pooled (network) 0.019 (−0.37, 0.40)
VD vs control
Kahleova 2011 −0.066 (−0.33, 0.20)
Pooled (pair−wise) −0.066 (−0.46, 0.33)
Indirect (back−calculated) NA
Pooled (network) −0.067 (−0.46, 0.32)
LCD vs fiber
Lousley 1984 0.052 (−0.41, 0.51)
Pooled (pair−wise) 0.053 (−0.49, 0.59)
Indirect (back−calculated) 0.097 (−0.21, 0.40)
Pooled (network) 0.0% 0.087 (−0.18, 0.35)
LGID vs LCD
Westman 2008 0.15 (−0.053, 0.34)
Pooled (pair−wise) 0.15 (−0.20, 0.50)
Indirect (back−calculated) −0.057 (−0.23, 0.11)
Pooled (network) 64.5% −0.018 (−0.17, 0.13)
Med vs LCD
Elhayany 2010 −0.075 (−0.29, 0.14)
Pooled (pair−wise) −0.076 (−0.44, 0.29)
Indirect (back−calculated) 0.095 (−0.14, 0.33)
Pooled (network) 26.2% 0.046 (−0.15, 0.24)
−0.8
0
0.8

**S9.4 Forest plots of IR (*PMD*)**

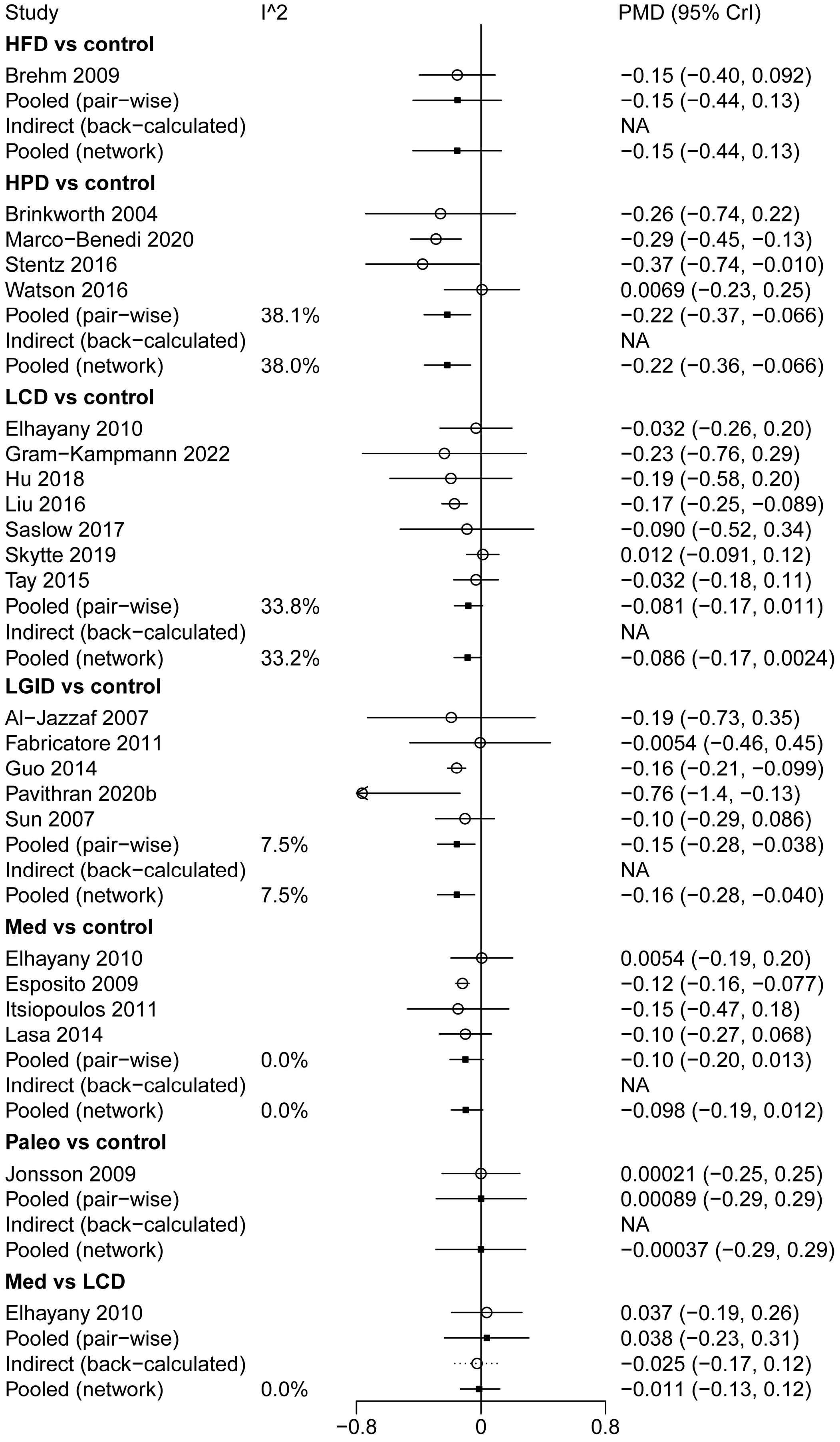

**S9.5 Forest plots of weight (*MD*, kg)**

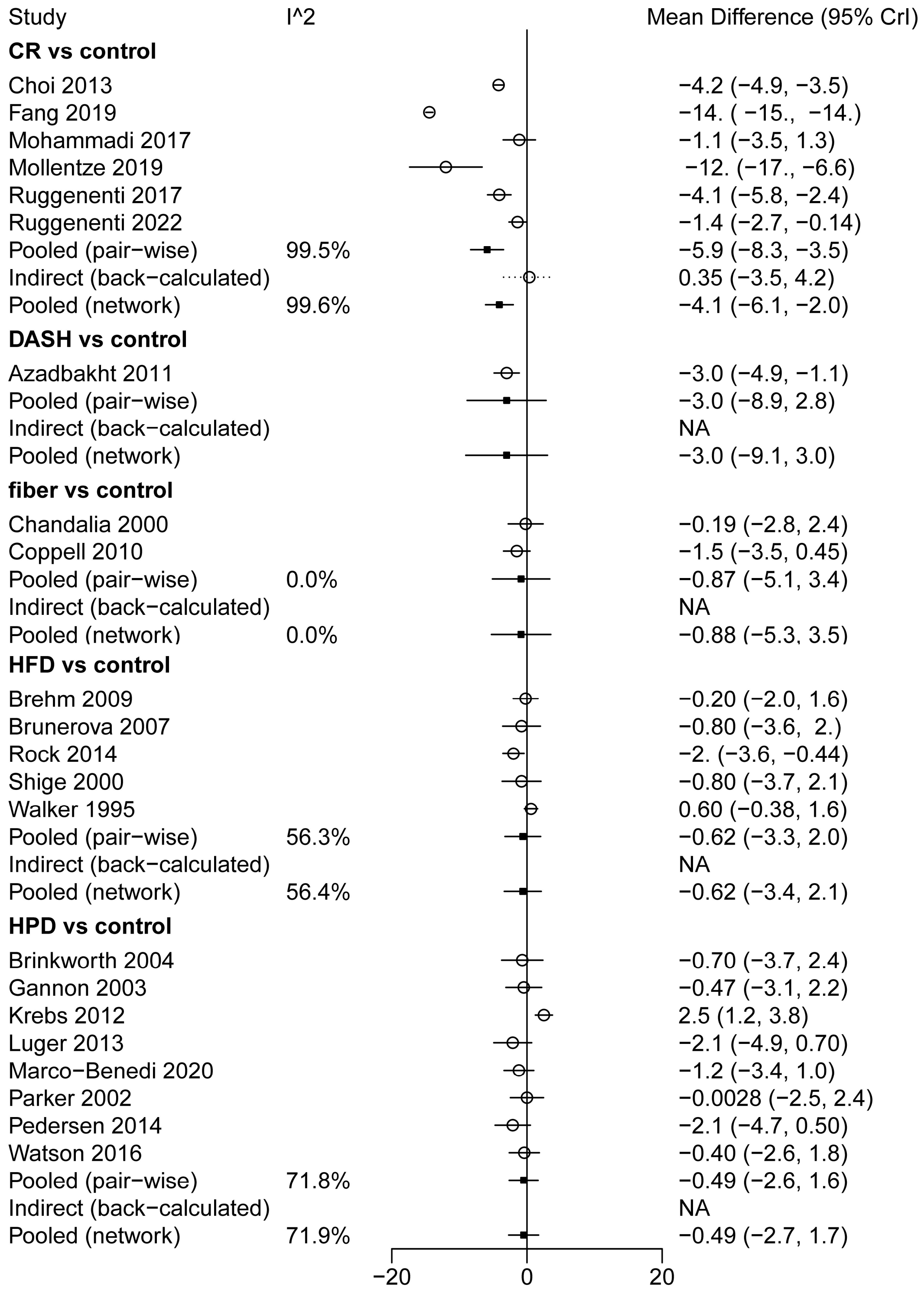

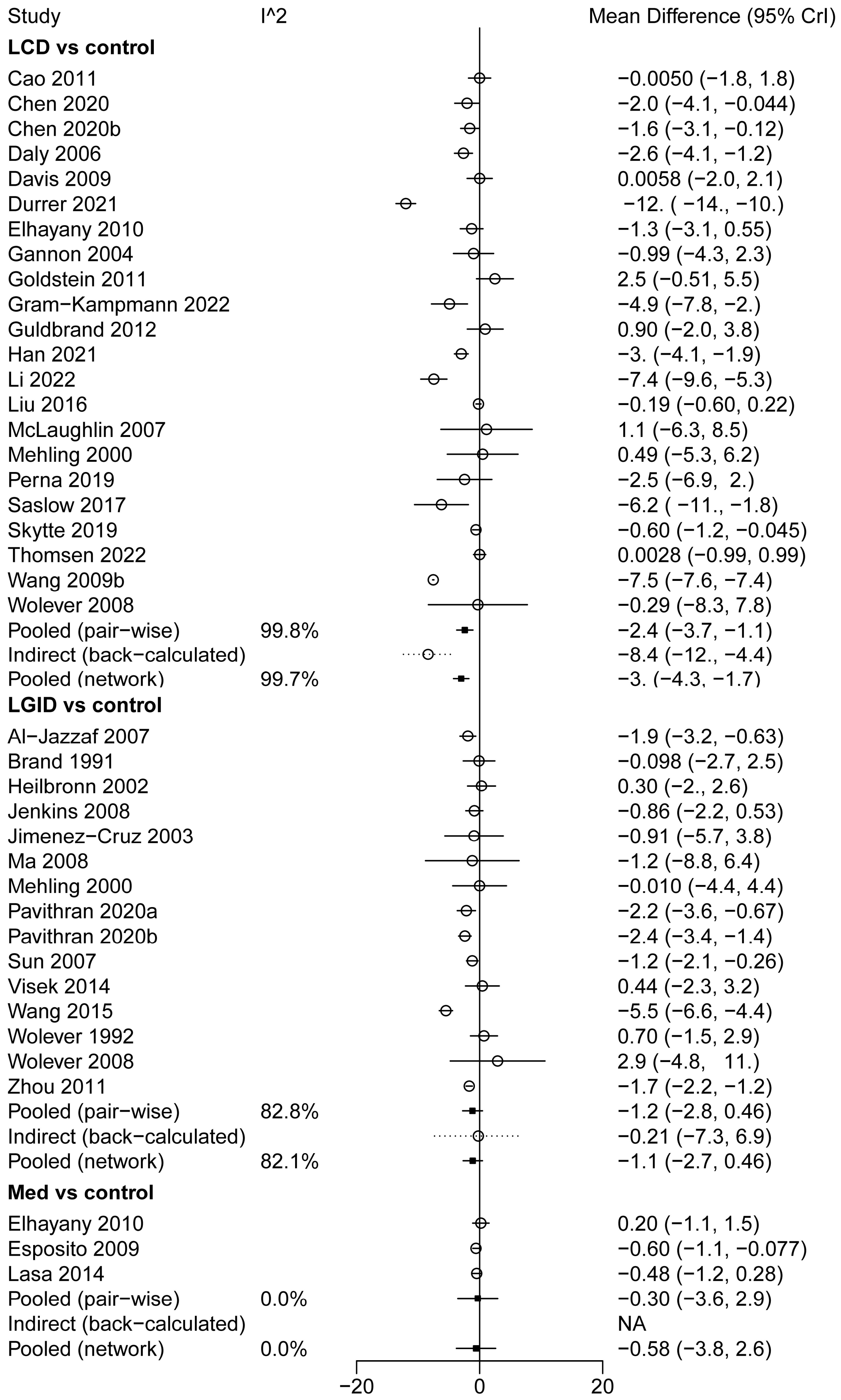
Study
I^2
Mean Difference (95% CrI)
LCD vs control
Cao 2011 −0.0050 (−1.8, 1.8)
Chen 2020 −2.0 (−4.1, −0.044)
Chen 2020b −1.6 (−3.1, −0.12)
Daly 2006 −2.6 (−4.1, −1.2)
Davis 2009 0.0058 (−2.0, 2.1)
Durrer 2021 −12. ( −14., −10.)
Elhayany 2010 −1.3 (−3.1, 0.55)
Gannon 2004 −0.99 (−4.3, 2.3)
Goldstein 2011 2.5 (−0.51, 5.5)
Gram−Kampmann 2022 −4.9 (−7.8, −2.)
Guldbrand 2012 0.90 (−2.0, 3.8)
Han 2021 −3. (−4.1, −1.9)
Li 2022 −7.4 (−9.6, −5.3)
Liu 2016 −0.19 (−0.60, 0.22)
McLaughlin 2007 1.1 (−6.3, 8.5)
Mehling 2000 0.49 (−5.3, 6.2)
Perna 2019 −2.5 (−6.9, 2.)
Saslow 2017 −6.2 ( −11., −1.8)
Skytte 2019 −0.60 (−1.2, −0.045)
Thomsen 2022 0.0028 (−0.99, 0.99)
Wang 2009b −7.5 (−7.6, −7.4)
Wolever 2008 −0.29 (−8.3, 7.8)
Pooled (pair−wise) 99.8% −2.4 (−3.7, −1.1)
Indirect (back−calculated) −8.4 (−12., −4.4)
Pooled (network) 99.7% −3. (−4.3, −1.7)
LGID vs control
Al−Jazzaf 2007 −1.9 (−3.2, −0.63)
Brand 1991 −0.098 (−2.7, 2.5)
Heilbronn 2002 0.30 (−2., 2.6)
Jenkins 2008 −0.86 (−2.2, 0.53)
Jimenez−Cruz 2003 −0.91 (−5.7, 3.8)
Ma 2008 −1.2 (−8.8, 6.4)
Mehling 2000 −0.010 (−4.4, 4.4)
Pavithran 2020a −2.2 (−3.6, −0.67)
Pavithran 2020b −2.4 (−3.4, −1.4)
Sun 2007 −1.2 (−2.1, −0.26)
Visek 2014 0.44 (−2.3, 3.2)
Wang 2015 −5.5 (−6.6, −4.4)
Wolever 1992 0.70 (−1.5, 2.9)
Wolever 2008 2.9 (−4.8, 11.)
Zhou 2011 −1.7 (−2.2, −1.2)
Pooled (pair−wise) 82.8% −1.2 (−2.8, 0.46)
Indirect (back−calculated) −0.21 (−7.3, 6.9)
Pooled (network) 82.1% −1.1 (−2.7, 0.46)
Med vs control
Elhayany 2010 0.20 (−1.1, 1.5)
Esposito 2009 −0.60 (−1.1, −0.077)
Lasa 2014 −0.48 (−1.2, 0.28)
Pooled (pair−wise) 0.0% −0.30 (−3.6, 2.9)
Indirect (back−calculated) NA
Pooled (network) 0.0% −0.58 (−3.8, 2.6)
−20
0
20

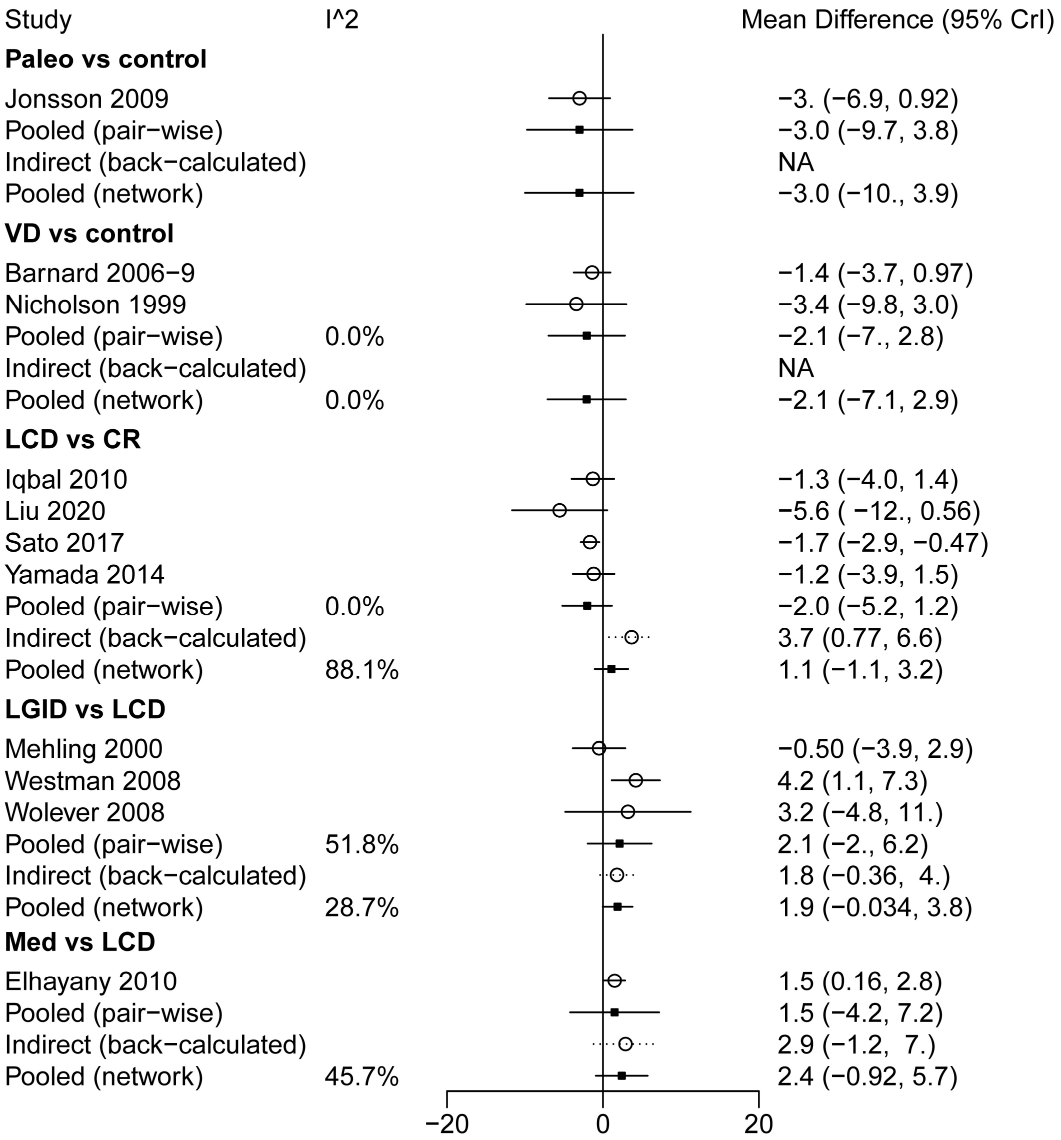
Study
I^2
Mean Difference (95% CrI)
Paleo vs control
Jonsson 2009 −3. (−6.9, 0.92)
Pooled (pair−wise) −3.0 (−9.7, 3.8)
Indirect (back−calculated) NA
Pooled (network) −3.0 (−10., 3.9)
VD vs control
Barnard 2006−9 −1.4 (−3.7, 0.97)
Nicholson 1999 −3.4 (−9.8, 3.0)
Pooled (pair−wise) 0.0% −2.1 (−7., 2.8)
Indirect (back−calculated) NA
Pooled (network) 0.0% −2.1 (−7.1, 2.9)
LCD vs CR
Iqbal 2010 −1.3 (−4.0, 1.4)
Liu 2020 −5.6 ( −12., 0.56)
Sato 2017 −1.7 (−2.9, −0.47)
Yamada 2014 −1.2 (−3.9, 1.5)
Pooled (pair−wise) 0.0% −2.0 (−5.2, 1.2)
Indirect (back−calculated) 3.7 (0.77, 6.6)
Pooled (network) 88.1% 1.1 (−1.1, 3.2)
LGID vs LCD
Mehling 2000 −0.50 (−3.9, 2.9)
Westman 2008 4.2 (1.1, 7.3)
Wolever 2008 3.2 (−4.8, 11.)
Pooled (pair−wise) 51.8% 2.1 (−2., 6.2)
Indirect (back−calculated) 1.8 (−0.36, 4.)
Pooled (network) 28.7% 1.9 (−0.034, 3.8)
Med vs LCD
Elhayany 2010 1.5 (0.16, 2.8)
Pooled (pair−wise) 1.5 (−4.2, 7.2)
Indirect (back−calculated) 2.9 (−1.2, 7.)
Pooled (network) 45.7% 2.4 (−0.92, 5.7)
−20
0
20

**S9.6 Forest plots of BMI (*MD*, kg/m$^2$)**

| Study | I^2 | Mean Difference (95% CrI) |
|---|---|---|
| **CR vs control** | | |
| Choi 2013 | | −1.7 (−2., −1.4) |
| Mohammadi 2017 | | −0.27 (−1.3, 0.73) |
| Mollentze 2019 | | −3.5 (−5.3, −1.7) |
| Ruggenenti 2017 | | −1.3 (−1.9, −0.74) |
| Ruggenenti 2022 | | −0.60 (−1.0, −0.15) |
| Uusitupa 1993 | | −0.90 (−1.6, −0.25) |
| Pooled (pair−wise) | 83.2% | −1.3 (−2.2, −0.34) |
| Indirect (back−calculated) | | −0.75 (−2.3, 0.80) |
| Pooled (network) | 82.1% | −1.1 (−1.9, −0.34) |
| **fiber vs control** | | |
| Coppell 2010 | | −0.60 (−1.4, 0.16) |
| Ikem 2007 | | 0.00016 (−0.61, 0.61) |
| Pooled (pair−wise) | 34.3% | −0.29 (−1.9, 1.3) |
| Indirect (back−calculated) | | NA |
| Pooled (network) | 34.2% | −0.29 (−1.8, 1.2) |
| **HFD vs control** | | |
| Rock 2014 | | −0.50 (−1.0, 0.018) |
| Walker 1995 | | −0.20 (−0.64, 0.24) |
| Pooled (pair−wise) | 0.0% | −0.35 (−1.9, 1.2) |
| Indirect (back−calculated) | | NA |
| Pooled (network) | 0.0% | −0.35 (−1.8, 1.2) |
| **HPD vs control** | | |
| Luger 2013 | | −0.80 (−1.7, 0.088) |
| Marco−Benedi 2020 | | −0.47 (−1.3, 0.37) |
| Pedersen 2014 | | −1.0 (−1.9, −0.13) |
| Stentz 2016 | | 0.60 (−0.93, 2.1) |
| Watson 2016 | | −0.24 (−0.95, 0.47) |
| Pooled (pair−wise) | 7.9% | −0.43 (−1.5, 0.61) |
| Indirect (back−calculated) | | NA |
| Pooled (network) | 7.9% | −0.43 (−1.4, 0.59) |
| **LCD vs control** | | |
| Chen 2020 | | −0.71 (−1.5, 0.100) |
| Chen 2020b | | −0.50 (−0.96, −0.041) |
| Durrer 2021 | | −4.4 (−5.1, −3.7) |
| Elhayany 2010 | | −0.50 (−0.94, −0.065) |
| Gram−Kampmann 2022 | | −1.5 (−2.6, −0.43) |
| Guldbrand 2012 | | 0.20 (−0.71, 1.1) |
| Han 2021 | | −1.3 (−1.6, −0.88) |
| Hu 2018 | | −0.40 (−1.3, 0.48) |
| Li 2021 | | −3.7 (−4.6, −2.7) |
| Li 2022 | | −2.5 (−3.5, −1.5) |
| Liu 2016 | | 0.033 (−0.13, 0.20) |
| McLaughlin 2007 | | 0.60 (−0.030, 1.2) |
| Mehling 2000 | | 0.20 (−2.3, 2.7) |
| Perna 2019 | | −0.78 (−2.5, 0.89) |
| Saslow 2017 | | −1.7 (−3.2, −0.19) |
| Skytte 2019 | | −0.20 (−0.48, 0.077) |
| Tay 2015 | | 0.30 (−0.62, 1.2) |
| Thomsen 2022 | | 0.00024 (−0.29, 0.29) |
| Wang 2009b | | −4.9 (−5., −4.8) |
| Wang 2016 | | 0.38 (−1.5, 2.3) |
| Pooled (pair−wise) | 99.8% | −1.1 (−1.6, −0.61) |
| Indirect (back−calculated) | | −1.6 (−2.8, −0.45) |
| Pooled (network) | 99.8% | −1.2 (−1.7, −0.74) |

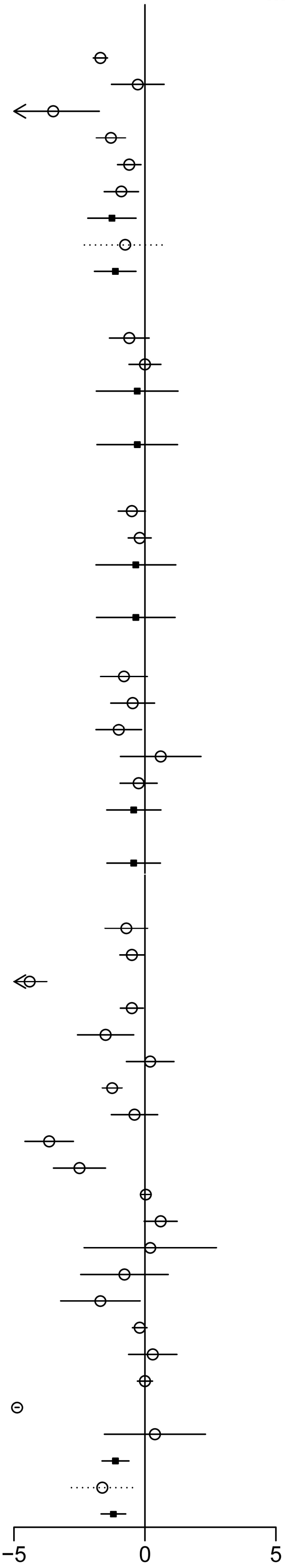

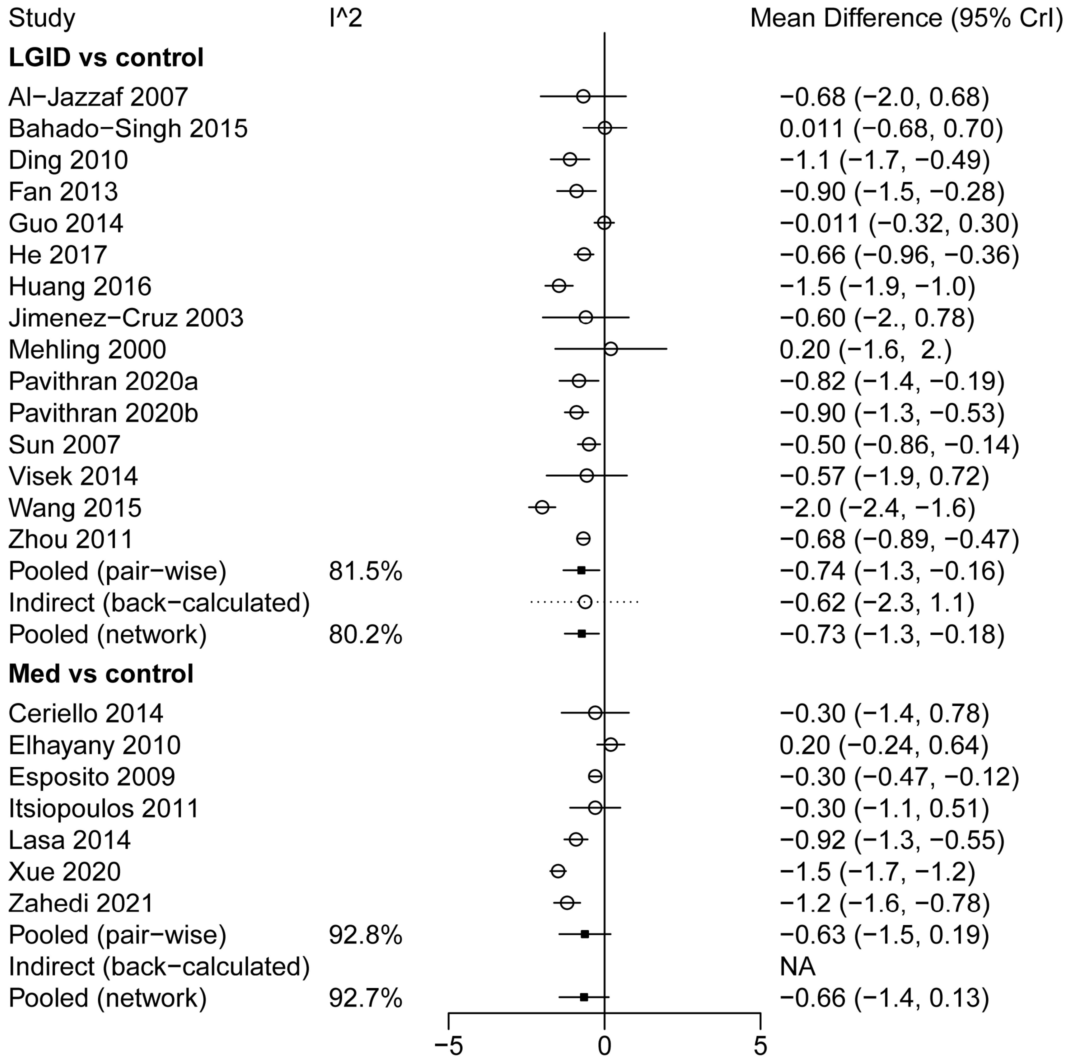
Study
I^2
Mean Difference (95% CrI)
LGID vs control
Al−Jazzaf 2007 −0.68 (−2.0, 0.68)
Bahado−Singh 2015 0.011 (−0.68, 0.70)
Ding 2010 −1.1 (−1.7, −0.49)
Fan 2013 −0.90 (−1.5, −0.28)
Guo 2014 −0.011 (−0.32, 0.30)
He 2017 −0.66 (−0.96, −0.36)
Huang 2016 −1.5 (−1.9, −1.0)
Jimenez−Cruz 2003 −0.60 (−2., 0.78)
Mehling 2000 0.20 (−1.6, 2.)
Pavithran 2020a −0.82 (−1.4, −0.19)
Pavithran 2020b −0.90 (−1.3, −0.53)
Sun 2007 −0.50 (−0.86, −0.14)
Visek 2014 −0.57 (−1.9, 0.72)
Wang 2015 −2.0 (−2.4, −1.6)
Zhou 2011 −0.68 (−0.89, −0.47)
Pooled (pair−wise) 81.5% −0.74 (−1.3, −0.16)
Indirect (back−calculated) −0.62 (−2.3, 1.1)
Pooled (network) 80.2% −0.73 (−1.3, −0.18)
Med vs control
Ceriello 2014 −0.30 (−1.4, 0.78)
Elhayany 2010 0.20 (−0.24, 0.64)
Esposito 2009 −0.30 (−0.47, −0.12)
Itsiopoulos 2011 −0.30 (−1.1, 0.51)
Lasa 2014 −0.92 (−1.3, −0.55)
Xue 2020 −1.5 (−1.7, −1.2)
Zahedi 2021 −1.2 (−1.6, −0.78)
Pooled (pair−wise) 92.8% −0.63 (−1.5, 0.19)
Indirect (back−calculated) NA
Pooled (network) 92.7% −0.66 (−1.4, 0.13)
−5
0
5

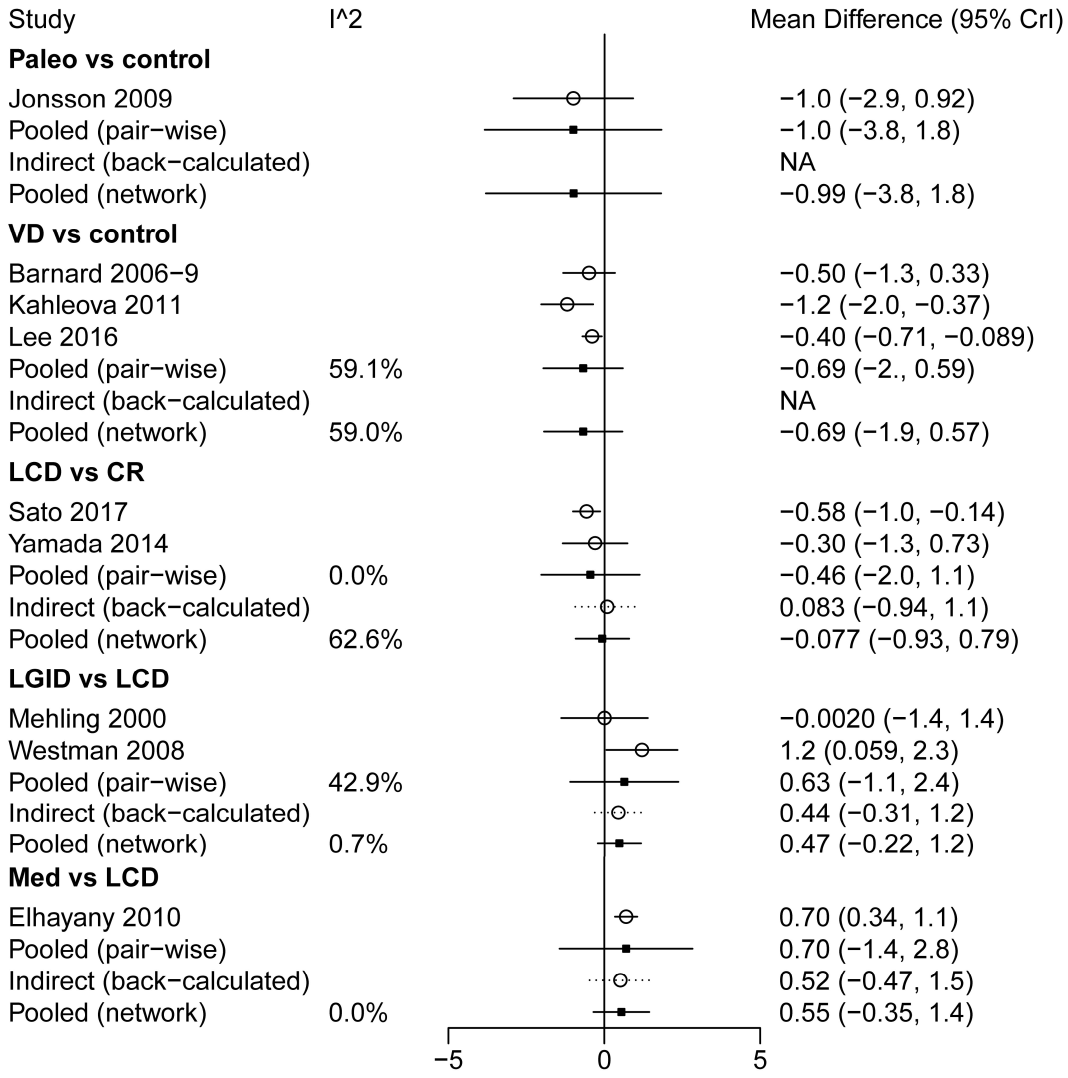
Study
I^2
Mean Difference (95% CrI)
Paleo vs control
Jonsson 2009
−1.0 (−2.9, 0.92)
Pooled (pair−wise)
−1.0 (−3.8, 1.8)
Indirect (back−calculated)
NA
Pooled (network)
−0.99 (−3.8, 1.8)
VD vs control
Barnard 2006−9
−0.50 (−1.3, 0.33)
Kahleova 2011
−1.2 (−2.0, −0.37)
Lee 2016
−0.40 (−0.71, −0.089)
Pooled (pair−wise)
59.1%
−0.69 (−2., 0.59)
Indirect (back−calculated)
NA
Pooled (network)
59.0%
−0.69 (−1.9, 0.57)
LCD vs CR
Sato 2017
−0.58 (−1.0, −0.14)
Yamada 2014
−0.30 (−1.3, 0.73)
Pooled (pair−wise)
0.0%
−0.46 (−2.0, 1.1)
Indirect (back−calculated)
0.083 (−0.94, 1.1)
Pooled (network)
62.6%
−0.077 (−0.93, 0.79)
LGID vs LCD
Mehling 2000
−0.0020 (−1.4, 1.4)
Westman 2008
1.2 (0.059, 2.3)
Pooled (pair−wise)
42.9%
0.63 (−1.1, 2.4)
Indirect (back−calculated)
0.44 (−0.31, 1.2)
Pooled (network)
0.7%
0.47 (−0.22, 1.2)
Med vs LCD
Elhayany 2010
0.70 (0.34, 1.1)
Pooled (pair−wise)
0.70 (−1.4, 2.8)
Indirect (back−calculated)
0.52 (−0.47, 1.5)
Pooled (network)
0.0%
0.55 (−0.35, 1.4)
−5
0
5

**S9.7 Forest plots of WC (*MD*, cm)**

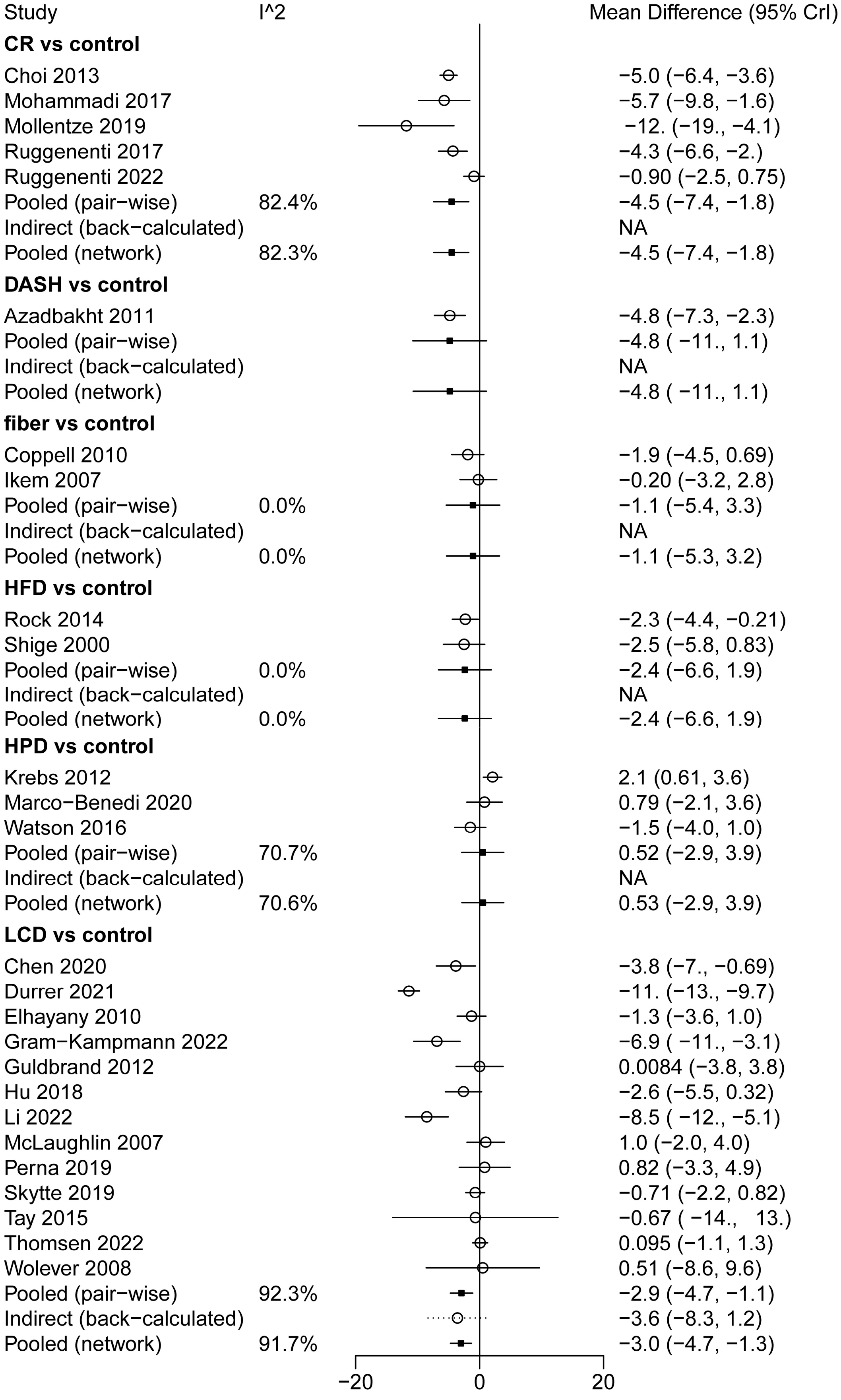

| Study | I^2 | Mean Difference (95% CrI) |
|---|---|---|
| **LGID vs control** | | |
| Al−Jazzaf 2007 | | −5.1 (−12., 1.9) |
| Ding 2010 | | −0.86 (−2.5, 0.81) |
| Fabricatore 2011 | | −0.70 (−4.6, 3.2) |
| He 2017 | | −3.6 (−5.2, −1.9) |
| Ma 2008 | | −2.5 (−7.2, 2.2) |
| Pavithran 2020a | | −0.99 (−4.5, 2.5) |
| Pavithran 2020b | | −2.1 (−4.1, −0.081) |
| Sun 2007 | | −1.8 (−3.7, 0.13) |
| Wang 2015 | | −6.3 (−9., −3.6) |
| Wolever 2008 | | 2.6 (−3.8, 9.0) |
| Zhou 2011 | | −1.4 (−3.5, 0.68) |
| Pooled (pair−wise) | 46.0% | −2.2 (−4.1, −0.24) |
| Indirect (back−calculated) | | −1.5 (−6.8, 3.9) |
| Pooled (network) | 41.1% | −2.1 (−3.9, −0.27) |
| **Med vs control** | | |
| Elhayany 2010 | | −0.20 (−2.2, 1.8) |
| Esposito 2009 | | −0.40 (−0.90, 0.097) |
| Pooled (pair−wise) | 0.0% | −0.31 (−4.3, 3.7) |
| Indirect (back−calculated) | | NA |
| Pooled (network) | 58.2% | −0.76 (−4.4, 2.9) |
| **Paleo vs control** | | |
| Jonsson 2009 | | −4.0 (−9.4, 1.4) |
| Pooled (pair−wise) | | −4. ( −12., 3.7) |
| Indirect (back−calculated) | | NA |
| Pooled (network) | | −4.0 ( −12., 3.6) |
| **VD vs control** | | |
| Barnard 2006−9 | | −2.4 (−4.9, 0.10) |
| Lee 2016 | | −2.3 (−4.2, −0.37) |
| Pooled (pair−wise) | 0.0% | −2.3 (−6.5, 1.8) |
| Indirect (back−calculated) | | NA |
| Pooled (network) | 0.0% | −2.4 (−6.5, 1.8) |
| **LGID vs LCD** | | |
| Westman 2008 | | 1.8 (−1.8, 5.4) |
| Wolever 2008 | | 2.1 (−4.3, 8.5) |
| Pooled (pair−wise) | 0.0% | 1.9 (−3.2, 7.0) |
| Indirect (back−calculated) | | 0.68 (−1.9, 3.3) |
| Pooled (network) | 0.0% | 0.93 (−1.4, 3.3) |
| **Med vs LCD** | | |
| Elhayany 2010 | | 1.1 (−0.77, 3.) |
| Pooled (pair−wise) | | 1.1 (−4.6, 6.8) |
| Indirect (back−calculated) | | 3.1 (−1.9, 8.2) |
| Pooled (network) | 36.2% | 2.2 (−1.6, 6.0) |

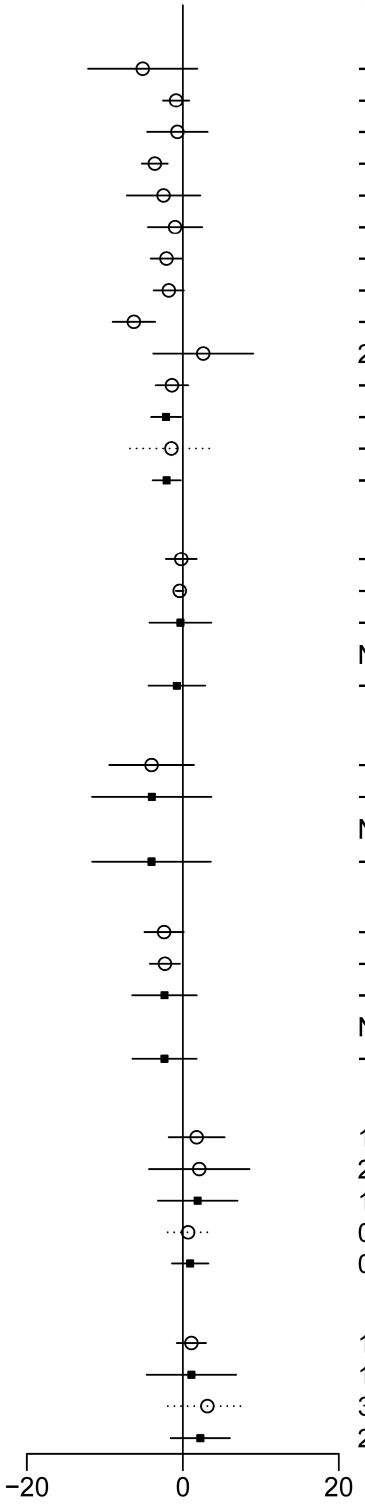

**S9.8 Forest plots of SBP (*MD*, mmHg)**

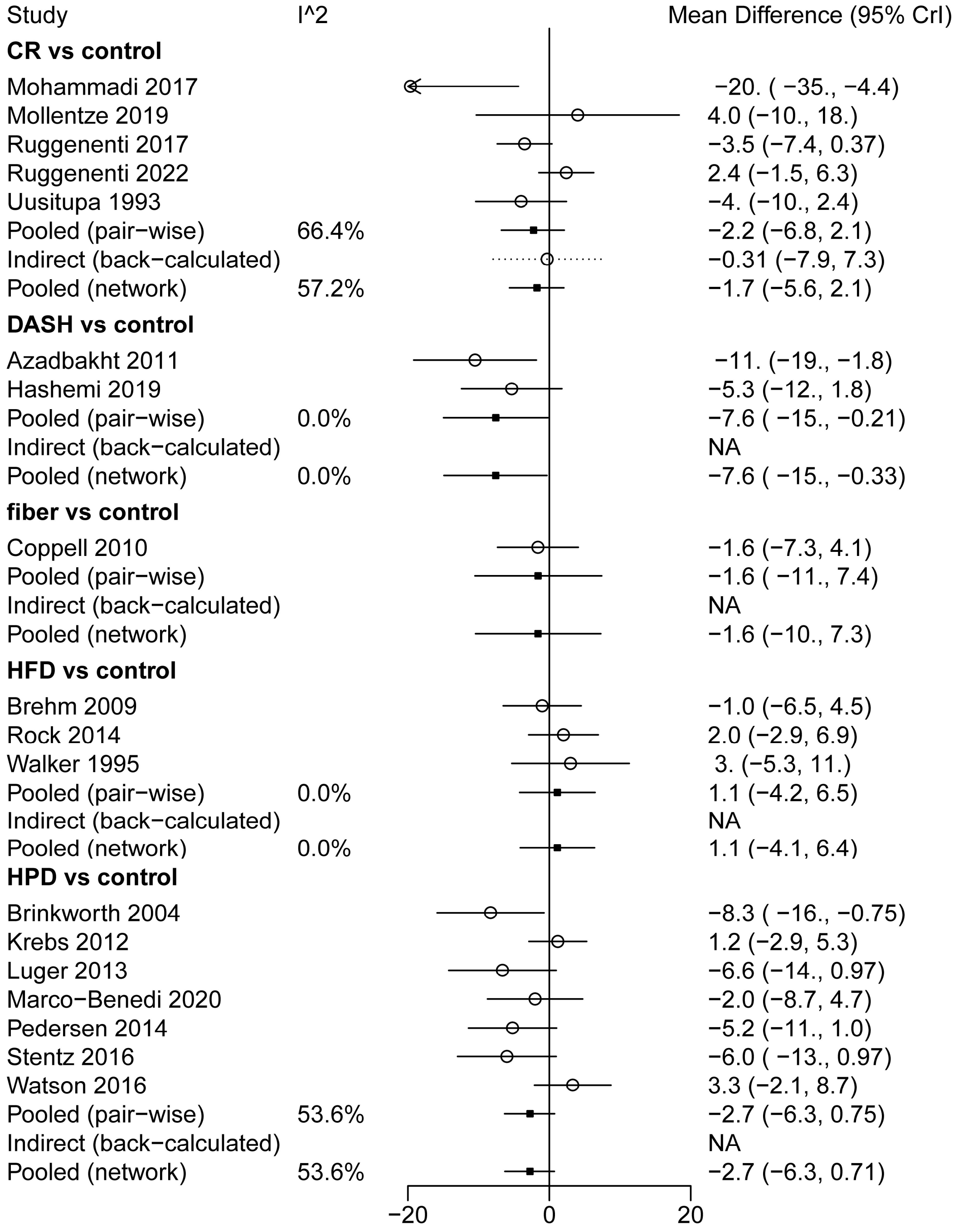

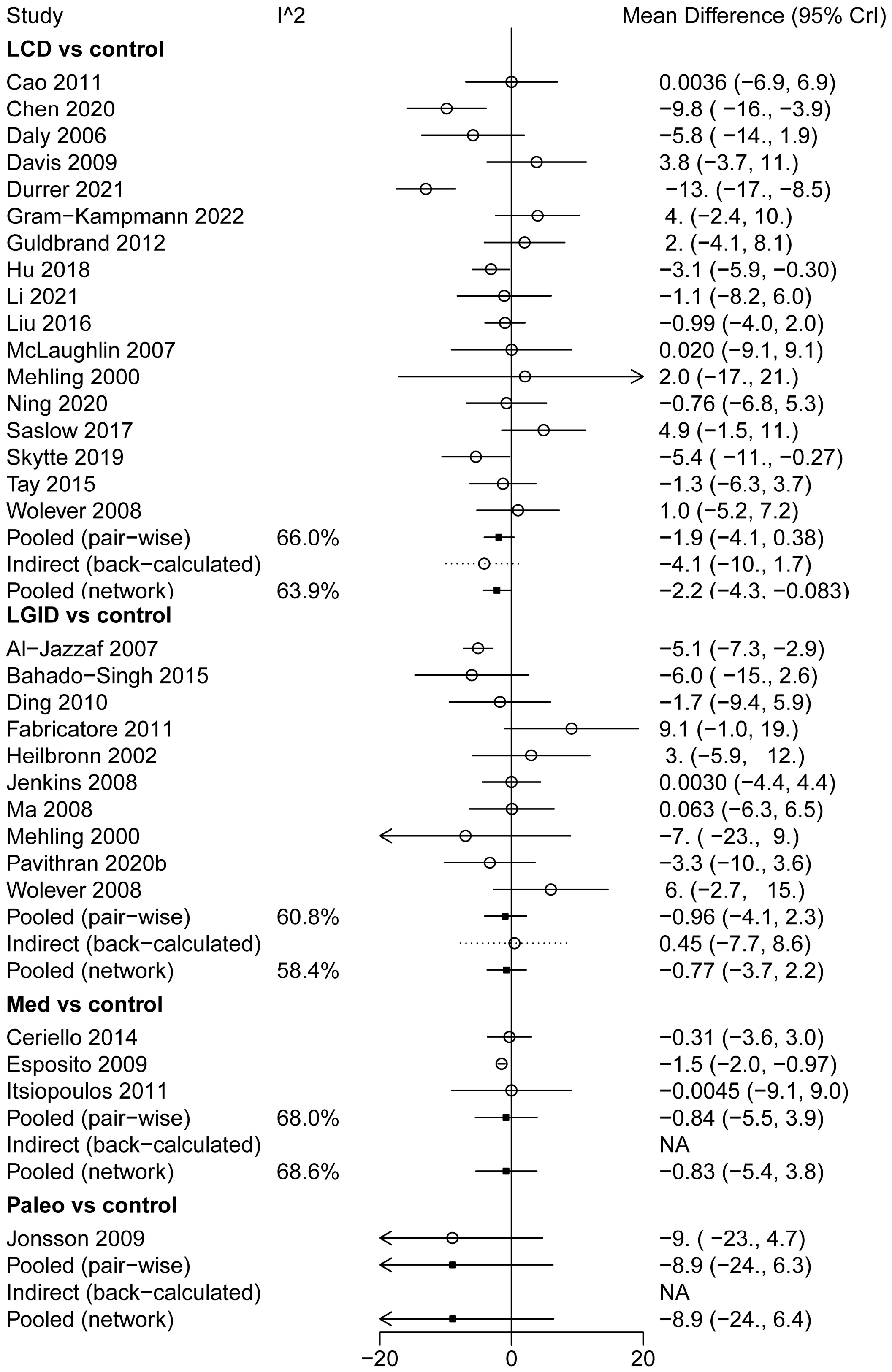
Study
I^2
Mean Difference (95% CrI)
LCD vs control
Cao 2011 0.0036 (−6.9, 6.9)
Chen 2020 −9.8 ( −16., −3.9)
Daly 2006 −5.8 ( −14., 1.9)
Davis 2009 3.8 (−3.7, 11.)
Durrer 2021 −13. (−17., −8.5)
Gram−Kampmann 2022 4. (−2.4, 10.)
Guldbrand 2012 2. (−4.1, 8.1)
Hu 2018 −3.1 (−5.9, −0.30)
Li 2021 −1.1 (−8.2, 6.0)
Liu 2016 −0.99 (−4.0, 2.0)
McLaughlin 2007 0.020 (−9.1, 9.1)
Mehling 2000 2.0 (−17., 21.)
Ning 2020 −0.76 (−6.8, 5.3)
Saslow 2017 4.9 (−1.5, 11.)
Skytte 2019 −5.4 ( −11., −0.27)
Tay 2015 −1.3 (−6.3, 3.7)
Wolever 2008 1.0 (−5.2, 7.2)
Pooled (pair−wise) 66.0% −1.9 (−4.1, 0.38)
Indirect (back−calculated) −4.1 (−10., 1.7)
Pooled (network) 63.9% −2.2 (−4.3, −0.083)
LGID vs control
Al−Jazzaf 2007 −5.1 (−7.3, −2.9)
Bahado−Singh 2015 −6.0 ( −15., 2.6)
Ding 2010 −1.7 (−9.4, 5.9)
Fabricatore 2011 9.1 (−1.0, 19.)
Heilbronn 2002 3. (−5.9, 12.)
Jenkins 2008 0.0030 (−4.4, 4.4)
Ma 2008 0.063 (−6.3, 6.5)
Mehling 2000 −7. ( −23., 9.)
Pavithran 2020b −3.3 (−10., 3.6)
Wolever 2008 6. (−2.7, 15.)
Pooled (pair−wise) 60.8% −0.96 (−4.1, 2.3)
Indirect (back−calculated) 0.45 (−7.7, 8.6)
Pooled (network) 58.4% −0.77 (−3.7, 2.2)
Med vs control
Ceriello 2014 −0.31 (−3.6, 3.0)
Esposito 2009 −1.5 (−2.0, −0.97)
Itsiopoulos 2011 −0.0045 (−9.1, 9.0)
Pooled (pair−wise) 68.0% −0.84 (−5.5, 3.9)
Indirect (back−calculated) NA
Pooled (network) 68.6% −0.83 (−5.4, 3.8)
Paleo vs control
Jonsson 2009 −9. ( −23., 4.7)
Pooled (pair−wise) −8.9 (−24., 6.3)
Indirect (back−calculated) NA
Pooled (network) −8.9 (−24., 6.4)
−20
0
20

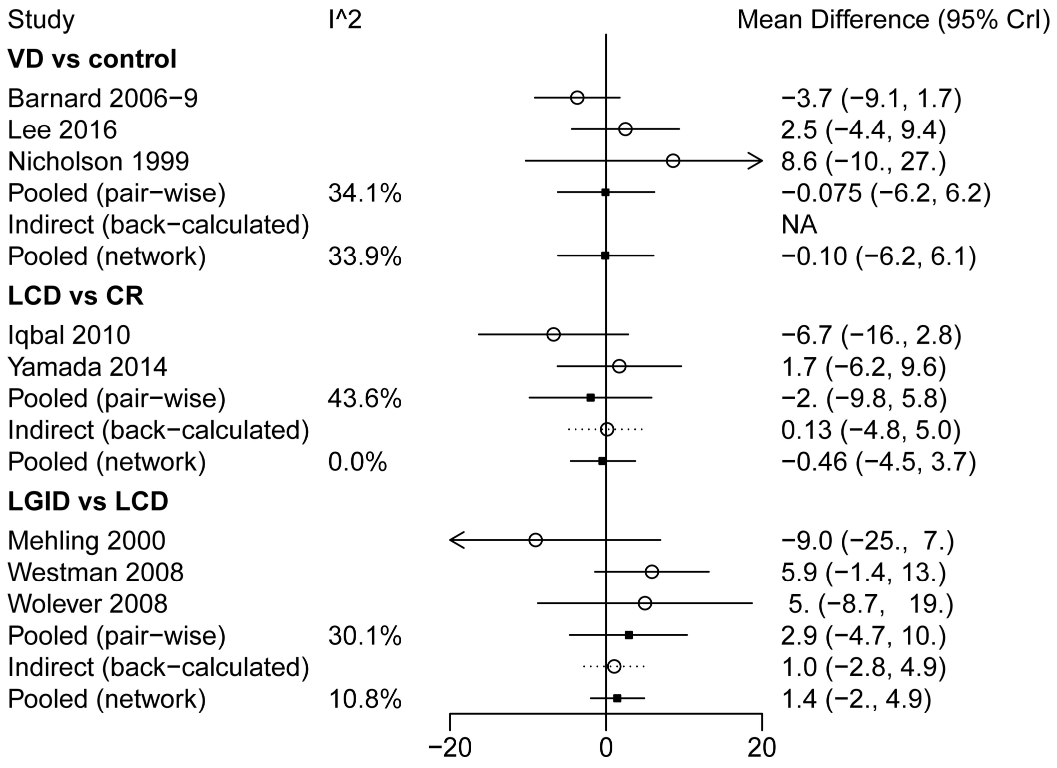
Study
I^2
Mean Difference (95% CrI)
VD vs control
Barnard 2006−9
−3.7 (−9.1, 1.7)
Lee 2016
2.5 (−4.4, 9.4)
Nicholson 1999
8.6 (−10., 27.)
Pooled (pair−wise)
34.1%
−0.075 (−6.2, 6.2)
Indirect (back−calculated)
NA
Pooled (network)
33.9%
−0.10 (−6.2, 6.1)
LCD vs CR
Iqbal 2010
−6.7 (−16., 2.8)
Yamada 2014
1.7 (−6.2, 9.6)
Pooled (pair−wise)
43.6%
−2. (−9.8, 5.8)
Indirect (back−calculated)
0.13 (−4.8, 5.0)
Pooled (network)
0.0%
−0.46 (−4.5, 3.7)
LGID vs LCD
Mehling 2000
−9.0 (−25., 7.)
Westman 2008
5.9 (−1.4, 13.)
Wolever 2008
5. (−8.7, 19.)
Pooled (pair−wise)
30.1%
2.9 (−4.7, 10.)
Indirect (back−calculated)
1.0 (−2.8, 4.9)
Pooled (network)
10.8%
1.4 (−2., 4.9)
−20
0
20

**S9.9 Forest plots of DBP (*MD*, mmHg)**

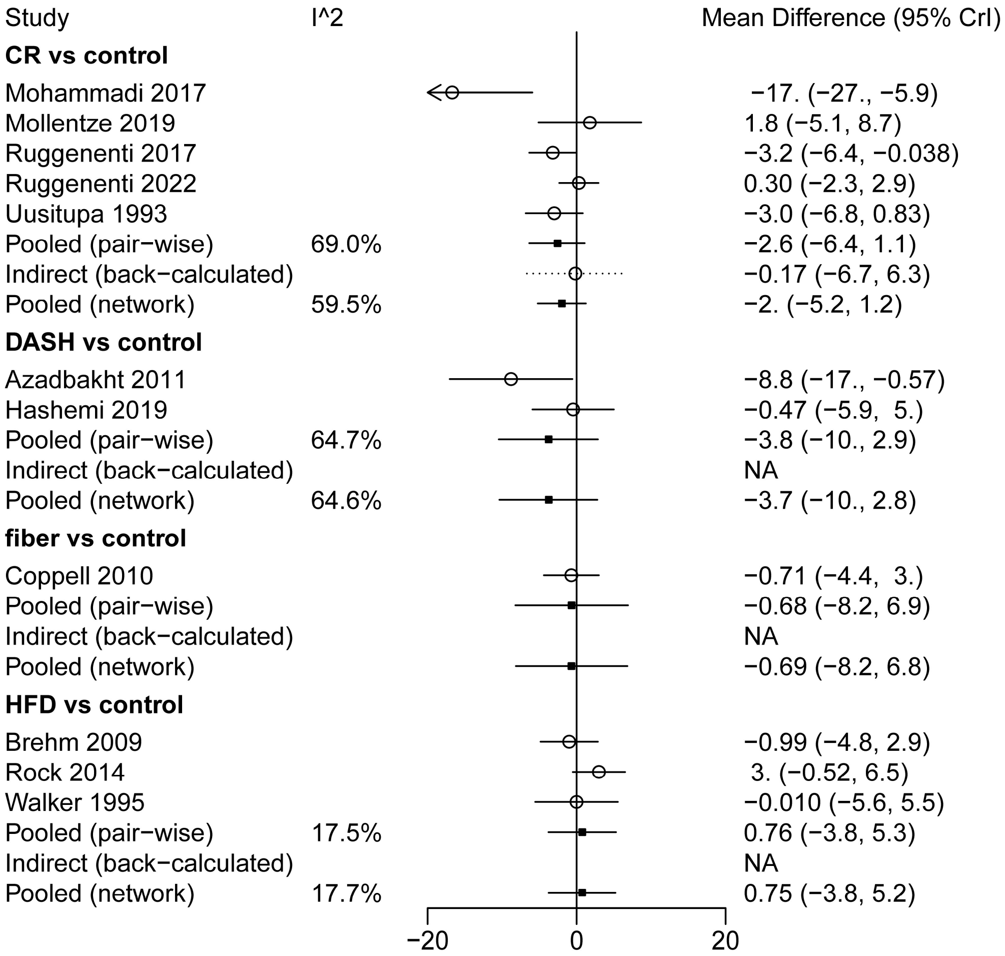

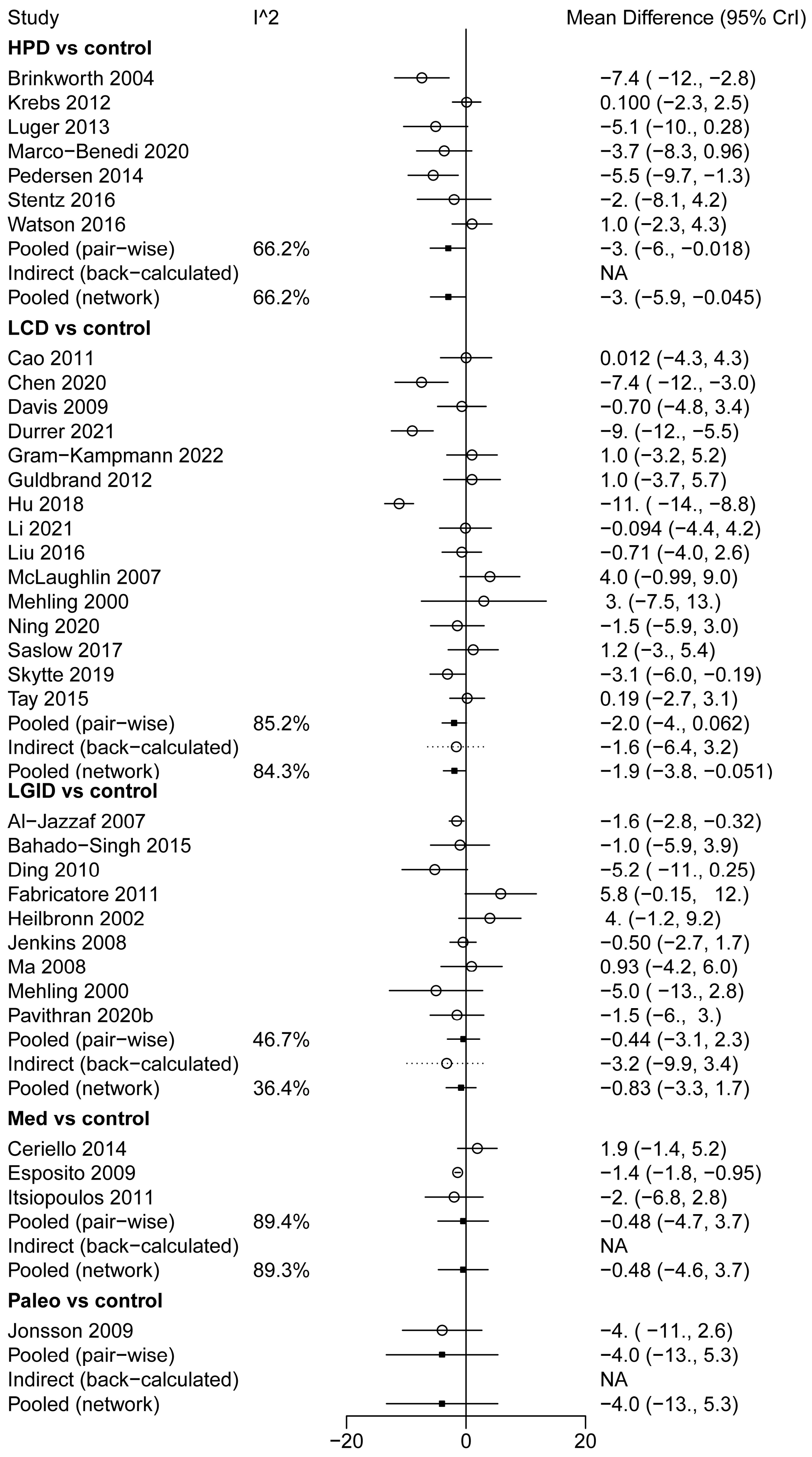
Study
I^2
Mean Difference (95% CrI)
HPD vs control
Brinkworth 2004 −7.4 ( −12., −2.8)
Krebs 2012 0.100 (−2.3, 2.5)
Luger 2013 −5.1 (−10., 0.28)
Marco−Benedi 2020 −3.7 (−8.3, 0.96)
Pedersen 2014 −5.5 (−9.7, −1.3)
Stentz 2016 −2. (−8.1, 4.2)
Watson 2016 1.0 (−2.3, 4.3)
Pooled (pair−wise) 66.2% −3. (−6., −0.018)
Indirect (back−calculated) NA
Pooled (network) 66.2% −3. (−5.9, −0.045)
LCD vs control
Cao 2011 0.012 (−4.3, 4.3)
Chen 2020 −7.4 ( −12., −3.0)
Davis 2009 −0.70 (−4.8, 3.4)
Durrer 2021 −9. (−12., −5.5)
Gram−Kampmann 2022 1.0 (−3.2, 5.2)
Guldbrand 2012 1.0 (−3.7, 5.7)
Hu 2018 −11. ( −14., −8.8)
Li 2021 −0.094 (−4.4, 4.2)
Liu 2016 −0.71 (−4.0, 2.6)
McLaughlin 2007 4.0 (−0.99, 9.0)
Mehling 2000 3. (−7.5, 13.)
Ning 2020 −1.5 (−5.9, 3.0)
Saslow 2017 1.2 (−3., 5.4)
Skytte 2019 −3.1 (−6.0, −0.19)
Tay 2015 0.19 (−2.7, 3.1)
Pooled (pair−wise) 85.2% −2.0 (−4., 0.062)
Indirect (back−calculated) −1.6 (−6.4, 3.2)
Pooled (network) 84.3% −1.9 (−3.8, −0.051)
LGID vs control
Al−Jazzaf 2007 −1.6 (−2.8, −0.32)
Bahado−Singh 2015 −1.0 (−5.9, 3.9)
Ding 2010 −5.2 ( −11., 0.25)
Fabricatore 2011 5.8 (−0.15, 12.)
Heilbronn 2002 4. (−1.2, 9.2)
Jenkins 2008 −0.50 (−2.7, 1.7)
Ma 2008 0.93 (−4.2, 6.0)
Mehling 2000 −5.0 ( −13., 2.8)
Pavithran 2020b −1.5 (−6., 3.)
Pooled (pair−wise) 46.7% −0.44 (−3.1, 2.3)
Indirect (back−calculated) −3.2 (−9.9, 3.4)
Pooled (network) 36.4% −0.83 (−3.3, 1.7)
Med vs control
Ceriello 2014 1.9 (−1.4, 5.2)
Esposito 2009 −1.4 (−1.8, −0.95)
Itsiopoulos 2011 −2. (−6.8, 2.8)
Pooled (pair−wise) 89.4% −0.48 (−4.7, 3.7)
Indirect (back−calculated) NA
Pooled (network) 89.3% −0.48 (−4.6, 3.7)
Paleo vs control
Jonsson 2009 −4. ( −11., 2.6)
Pooled (pair−wise) −4.0 (−13., 5.3)
Indirect (back−calculated) NA
Pooled (network) −4.0 (−13., 5.3)
−20
0
20

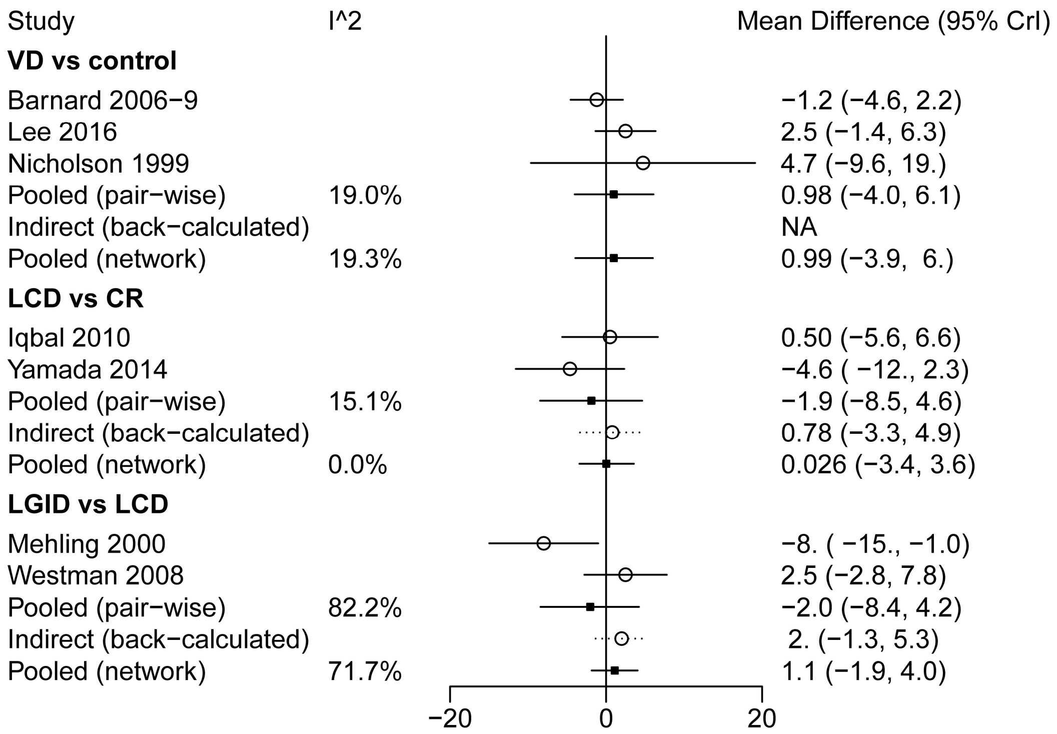
Study
I^2
Mean Difference (95% CrI)
VD vs control
Barnard 2006−9
−1.2 (−4.6, 2.2)
Lee 2016
2.5 (−1.4, 6.3)
Nicholson 1999
4.7 (−9.6, 19.)
Pooled (pair−wise)
19.0%
0.98 (−4.0, 6.1)
Indirect (back−calculated)
NA
Pooled (network)
19.3%
0.99 (−3.9, 6.)
LCD vs CR
Iqbal 2010
0.50 (−5.6, 6.6)
Yamada 2014
−4.6 ( −12., 2.3)
Pooled (pair−wise)
15.1%
−1.9 (−8.5, 4.6)
Indirect (back−calculated)
0.78 (−3.3, 4.9)
Pooled (network)
0.0%
0.026 (−3.4, 3.6)
LGID vs LCD
Mehling 2000
−8. ( −15., −1.0)
Westman 2008
2.5 (−2.8, 7.8)
Pooled (pair−wise)
82.2%
−2.0 (−8.4, 4.2)
Indirect (back−calculated)
2. (−1.3, 5.3)
Pooled (network)
71.7%
1.1 (−1.9, 4.0)
−20
0
20

**S9.10 Forest plots of TG (*MD*, mmol/L)**

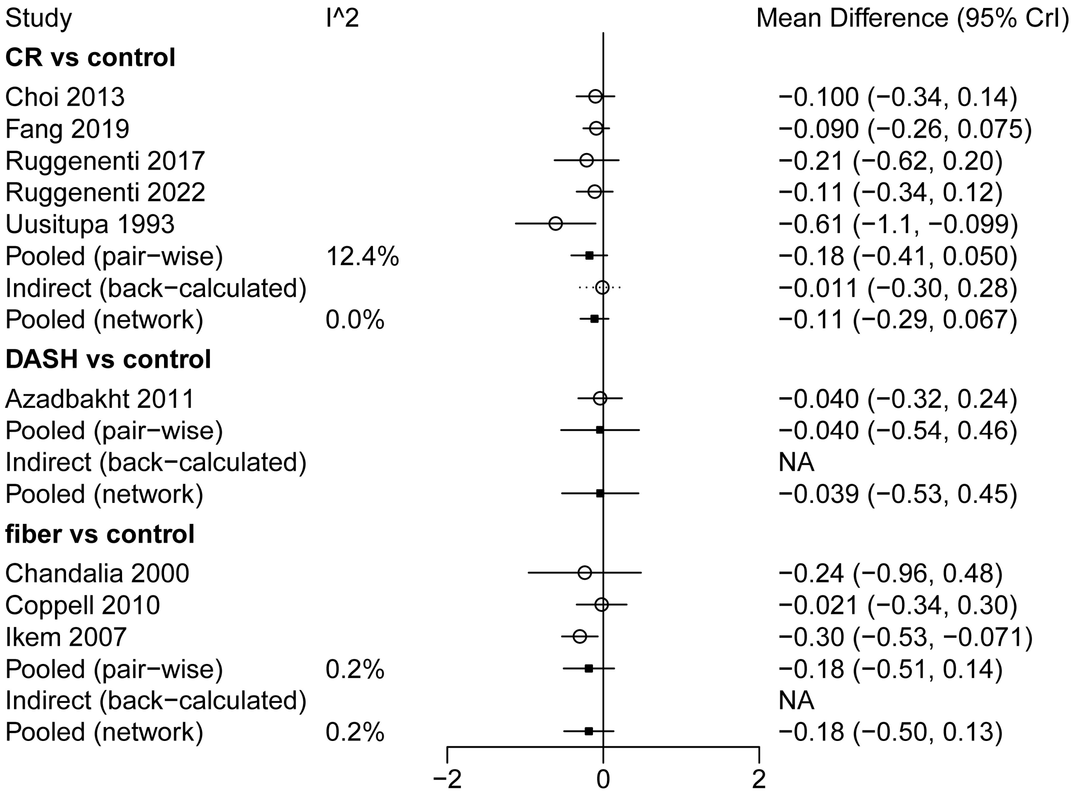

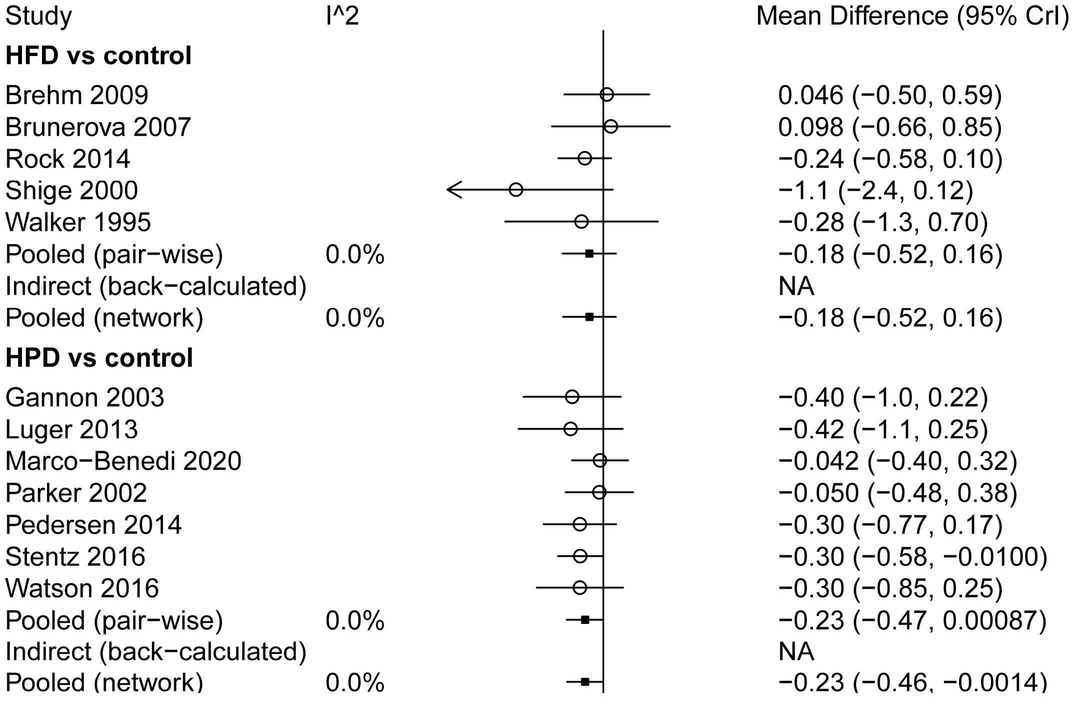

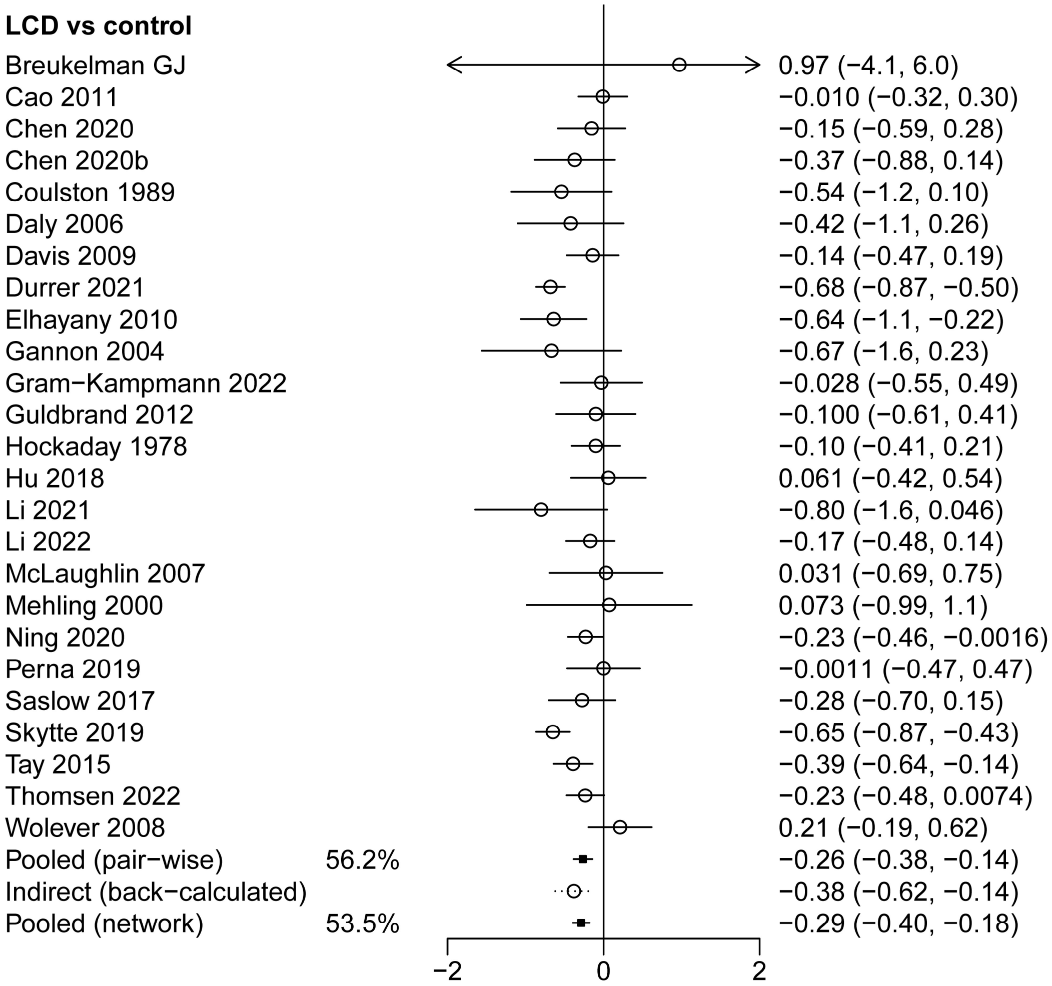
LCD vs control
Breukelman GJ 0.97 (−4.1, 6.0)
Cao 2011 −0.010 (−0.32, 0.30)
Chen 2020 −0.15 (−0.59, 0.28)
Chen 2020b −0.37 (−0.88, 0.14)
Coulston 1989 −0.54 (−1.2, 0.10)
Daly 2006 −0.42 (−1.1, 0.26)
Davis 2009 −0.14 (−0.47, 0.19)
Durrer 2021 −0.68 (−0.87, −0.50)
Elhayany 2010 −0.64 (−1.1, −0.22)
Gannon 2004 −0.67 (−1.6, 0.23)
Gram−Kampmann 2022 −0.028 (−0.55, 0.49)
Guldbrand 2012 −0.100 (−0.61, 0.41)
Hockaday 1978 −0.10 (−0.41, 0.21)
Hu 2018 0.061 (−0.42, 0.54)
Li 2021 −0.80 (−1.6, 0.046)
Li 2022 −0.17 (−0.48, 0.14)
McLaughlin 2007 0.031 (−0.69, 0.75)
Mehling 2000 0.073 (−0.99, 1.1)
Ning 2020 −0.23 (−0.46, −0.0016)
Perna 2019 −0.0011 (−0.47, 0.47)
Saslow 2017 −0.28 (−0.70, 0.15)
Skytte 2019 −0.65 (−0.87, −0.43)
Tay 2015 −0.39 (−0.64, −0.14)
Thomsen 2022 −0.23 (−0.48, 0.0074)
Wolever 2008 0.21 (−0.19, 0.62)
Pooled (pair−wise) 56.2% −0.26 (−0.38, −0.14)
Indirect (back−calculated) −0.38 (−0.62, −0.14)
Pooled (network) 53.5% −0.29 (−0.40, −0.18)
−2
0
2

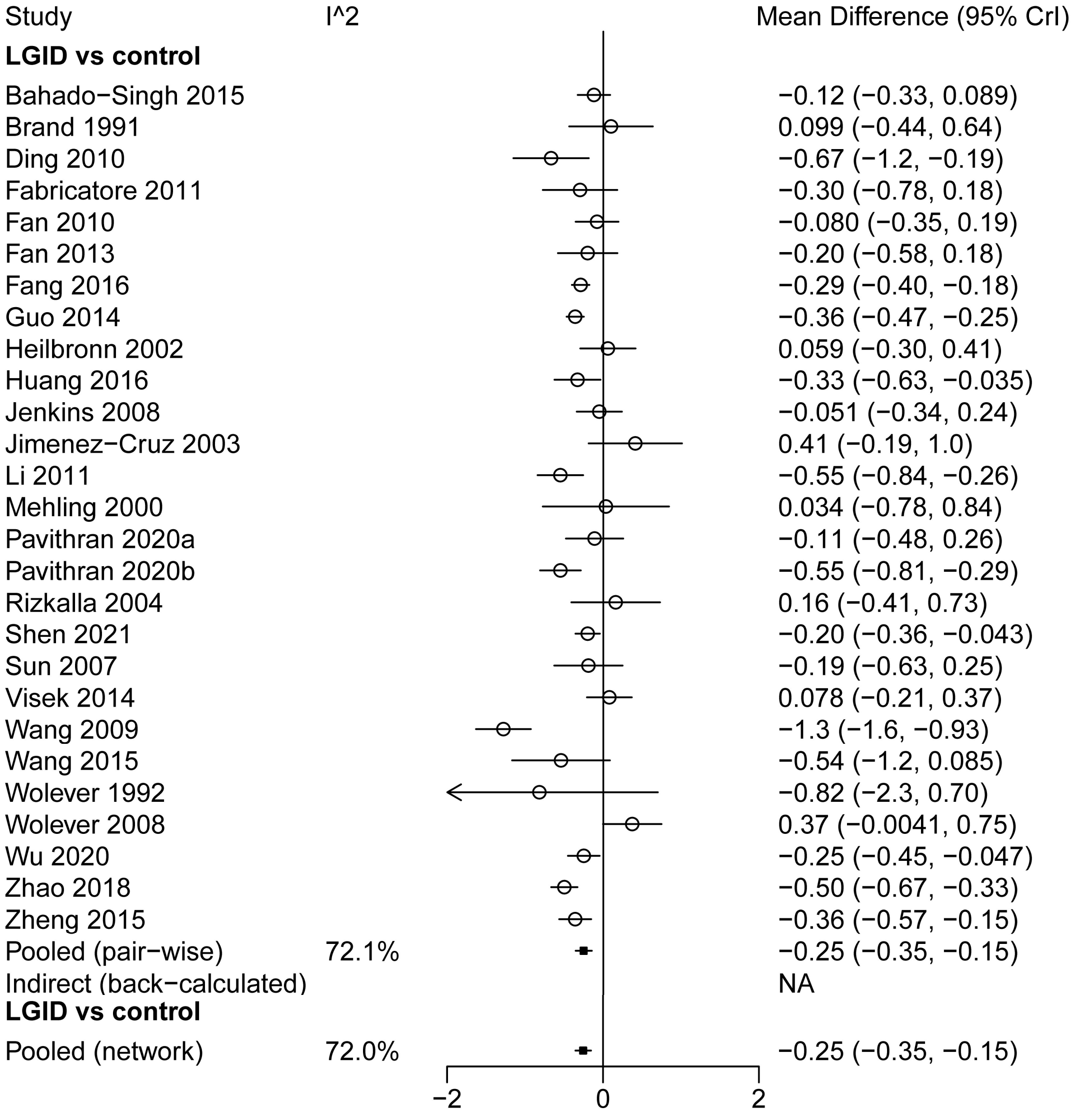
Study
I^2
Mean Difference (95% CrI)
LGID vs control
Bahado−Singh 2015 −0.12 (−0.33, 0.089)
Brand 1991 0.099 (−0.44, 0.64)
Ding 2010 −0.67 (−1.2, −0.19)
Fabricatore 2011 −0.30 (−0.78, 0.18)
Fan 2010 −0.080 (−0.35, 0.19)
Fan 2013 −0.20 (−0.58, 0.18)
Fang 2016 −0.29 (−0.40, −0.18)
Guo 2014 −0.36 (−0.47, −0.25)
Heilbronn 2002 0.059 (−0.30, 0.41)
Huang 2016 −0.33 (−0.63, −0.035)
Jenkins 2008 −0.051 (−0.34, 0.24)
Jimenez−Cruz 2003 0.41 (−0.19, 1.0)
Li 2011 −0.55 (−0.84, −0.26)
Mehling 2000 0.034 (−0.78, 0.84)
Pavithran 2020a −0.11 (−0.48, 0.26)
Pavithran 2020b −0.55 (−0.81, −0.29)
Rizkalla 2004 0.16 (−0.41, 0.73)
Shen 2021 −0.20 (−0.36, −0.043)
Sun 2007 −0.19 (−0.63, 0.25)
Visek 2014 0.078 (−0.21, 0.37)
Wang 2009 −1.3 (−1.6, −0.93)
Wang 2015 −0.54 (−1.2, 0.085)
Wolever 1992 −0.82 (−2.3, 0.70)
Wolever 2008 0.37 (−0.0041, 0.75)
Wu 2020 −0.25 (−0.45, −0.047)
Zhao 2018 −0.50 (−0.67, −0.33)
Zheng 2015 −0.36 (−0.57, −0.15)
Pooled (pair−wise) 72.1% −0.25 (−0.35, −0.15)
Indirect (back−calculated) NA
LGID vs control
Pooled (network) 72.0% −0.25 (−0.35, −0.15)
−2
0
2

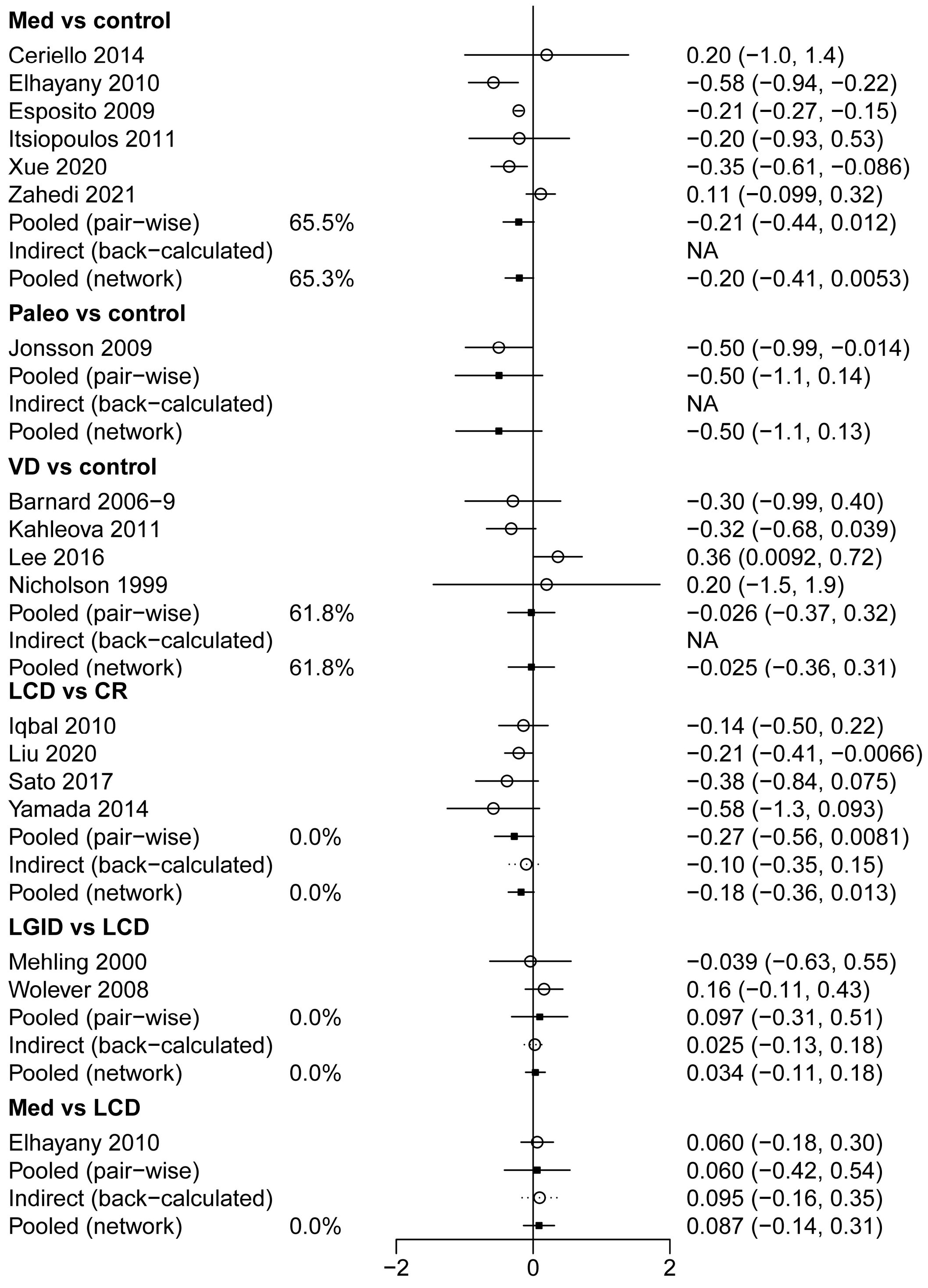
Med vs control
Ceriello 2014
0.20 (−1.0, 1.4)
Elhayany 2010
−0.58 (−0.94, −0.22)
Esposito 2009
−0.21 (−0.27, −0.15)
Itsiopoulos 2011
−0.20 (−0.93, 0.53)
Xue 2020
−0.35 (−0.61, −0.086)
Zahedi 2021
0.11 (−0.099, 0.32)
Pooled (pair−wise)
65.5%
−0.21 (−0.44, 0.012)
Indirect (back−calculated)
NA
Pooled (network)
65.3%
−0.20 (−0.41, 0.0053)
Paleo vs control
Jonsson 2009
−0.50 (−0.99, −0.014)
Pooled (pair−wise)
−0.50 (−1.1, 0.14)
Indirect (back−calculated)
NA
Pooled (network)
−0.50 (−1.1, 0.13)
VD vs control
Barnard 2006−9
−0.30 (−0.99, 0.40)
Kahleova 2011
−0.32 (−0.68, 0.039)
Lee 2016
0.36 (0.0092, 0.72)
Nicholson 1999
0.20 (−1.5, 1.9)
Pooled (pair−wise)
61.8%
−0.026 (−0.37, 0.32)
Indirect (back−calculated)
NA
Pooled (network)
61.8%
−0.025 (−0.36, 0.31)
LCD vs CR
Iqbal 2010
−0.14 (−0.50, 0.22)
Liu 2020
−0.21 (−0.41, −0.0066)
Sato 2017
−0.38 (−0.84, 0.075)
Yamada 2014
−0.58 (−1.3, 0.093)
Pooled (pair−wise)
0.0%
−0.27 (−0.56, 0.0081)
Indirect (back−calculated)
−0.10 (−0.35, 0.15)
Pooled (network)
0.0%
−0.18 (−0.36, 0.013)
LGID vs LCD
Mehling 2000
−0.039 (−0.63, 0.55)
Wolever 2008
0.16 (−0.11, 0.43)
Pooled (pair−wise)
0.0%
0.097 (−0.31, 0.51)
Indirect (back−calculated)
0.025 (−0.13, 0.18)
Pooled (network)
0.0%
0.034 (−0.11, 0.18)
Med vs LCD
Elhayany 2010
0.060 (−0.18, 0.30)
Pooled (pair−wise)
0.060 (−0.42, 0.54)
Indirect (back−calculated)
0.095 (−0.16, 0.35)
Pooled (network)
0.0%
0.087 (−0.14, 0.31)
−2
0
2

**S9.11 Forest plots of TC (*MD*, mmol/L)**

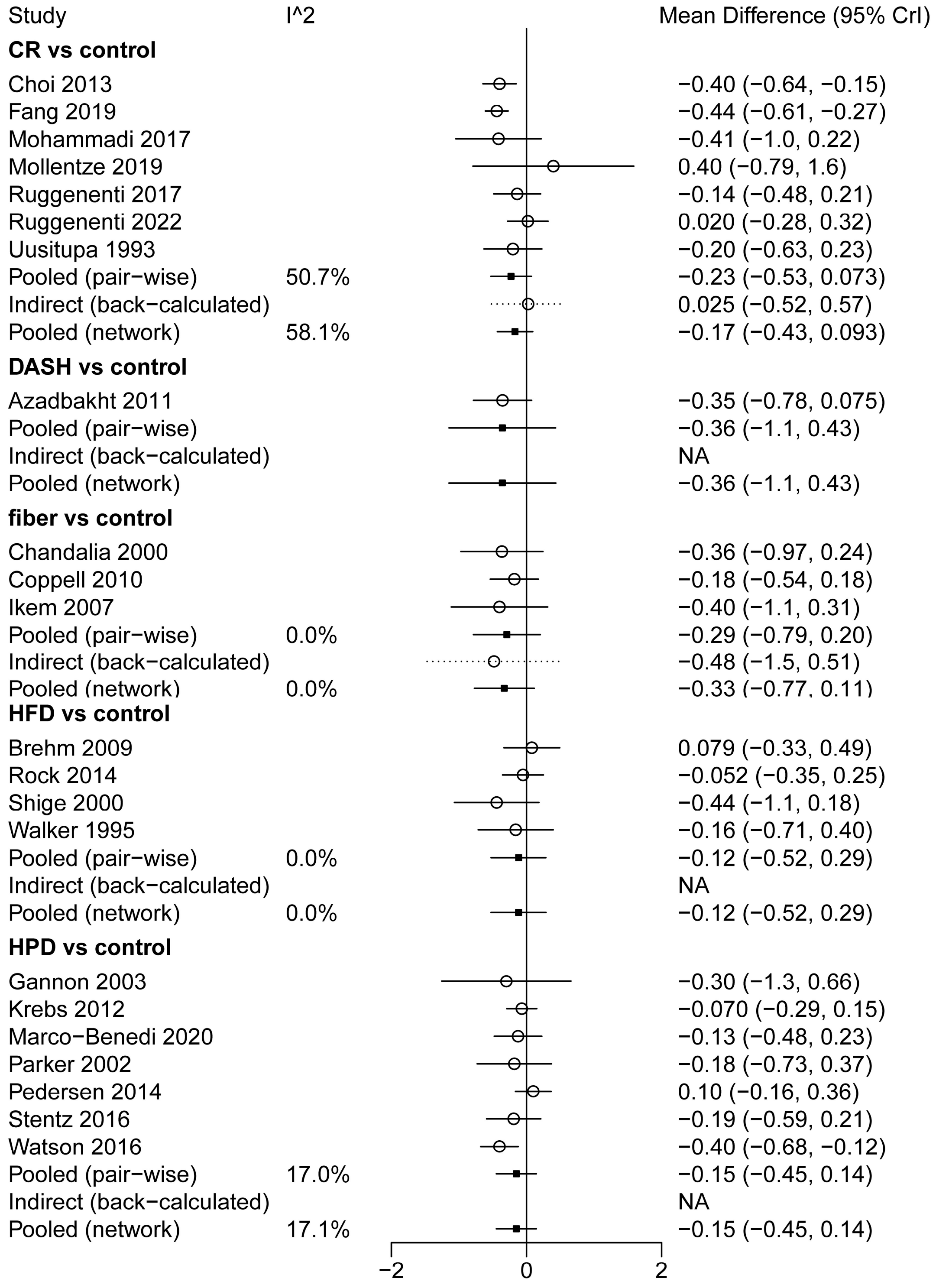

| Study | I^2 | Mean Difference (95% CrI) |
|---|---|---|
| **LCD vs control** | | |
| Breukelman GJ | | −0.15 (−8.4, 8.1) |
| Cao 2011 | | −0.10 (−0.40, 0.20) |
| Chen 2020 | | −0.0077 (−0.42, 0.40) |
| Chen 2020b | | −0.099 (−0.50, 0.30) |
| Coulston 1989 | | −0.051 (−1.2, 1.1) |
| Davis 2009 | | 0.23 (−0.049, 0.51) |
| Durrer 2021 | | −0.070 (−0.35, 0.21) |
| Elhayany 2010 | | −0.020 (−0.41, 0.37) |
| Gannon 2004 | | −0.0028 (−0.90, 0.90) |
| Gram−Kampmann 2022 | | 0.40 (−0.029, 0.83) |
| Guldbrand 2012 | | 0.20 (−0.26, 0.66) |
| Hockaday 1978 | | 0.40 (−0.14, 0.94) |
| Hu 2018 | | −0.17 (−0.61, 0.27) |
| Li 2021 | | −1.3 (−1.8, −0.83) |
| Li 2022 | | −0.19 (−0.50, 0.12) |
| McLaughlin 2007 | | −0.13 (−0.82, 0.56) |
| Mehling 2000 | | 0.069 (−1.2, 1.3) |
| Ning 2020 | | −0.59 (−1.0, −0.16) |
| Perna 2019 | | −0.097 (−0.87, 0.67) |
| Skytte 2019 | | −0.44 (−0.72, −0.16) |
| Tay 2015 | | 0.00039 (−0.28, 0.28) |
| Thomsen 2022 | | −0.098 (−0.39, 0.19) |
| Wang 2016 | | −0.44 (−0.95, 0.071) |
| Wolever 2008 | | −0.20 (−0.63, 0.23) |
| Pooled (pair−wise) | 57.3% | −0.12 (−0.30, 0.044) |
| Indirect (back−calculated) | | −0.36 (−0.74, 0.013) |
| Pooled (network) | 57.4% | −0.17 (−0.32, −0.0099) |
| **LGID vs control** | | |
| Al−Jazzaf 2007 | | 0.33 (−0.22, 0.88) |
| Bahado−Singh 2015 | | −0.032 (−0.62, 0.56) |
| Brand 1991 | | −0.011 (−0.67, 0.65) |
| Ding 2010 | | −0.27 (−0.80, 0.26) |
| Fabricatore 2011 | | −0.37 (−0.94, 0.21) |
| Fan 2010 | | −0.24 (−0.71, 0.23) |
| Fan 2013 | | 0.0013 (−0.72, 0.72) |
| Fang 2016 | | −1.1 (−1.4, −0.86) |
| Heilbronn 2002 | | −0.17 (−0.66, 0.32) |
| Huang 2016 | | 0.0012 (−0.60, 0.60) |
| Jimenez−Cruz 2003 | | 0.10 (−0.52, 0.72) |
| Li 2011 | | −1.8 (−2.3, −1.3) |
| Liu 2011 | | −1.4 (−2.6, −0.26) |
| Ma 2008 | | 0.43 (−0.23, 1.1) |
| Mehling 2000 | | −0.041 (−0.82, 0.74) |
| Pavithran 2020a | | −0.48 (−1.1, 0.11) |
| Pavithran 2020b | | −0.075 (−0.47, 0.32) |
| Rizkalla 2004 | | −0.98 (−1.8, −0.15) |
| Shen 2021 | | −0.68 (−1.0, −0.36) |
| Sun 2007 | | 0.14 (−0.24, 0.52) |
| Wang 2009 | | −1.8 (−2.4, −1.2) |
| Wang 2015 | | −1.5 (−1.8, −1.1) |
| Wolever 1992 | | −0.76 (−2.8, 1.3) |
| Wolever 2008 | | −0.23 (−0.66, 0.20) |
| Wu 2020 | | −0.37 (−0.57, −0.17) |
| Zhao 2018 | | −0.70 (−1.1, −0.31) |
| Zheng 2015 | | −0.88 (−1.3, −0.46) |
| Pooled (pair−wise) | 83.5% | −0.48 (−0.64, −0.31) |
| Indirect (back−calculated) | | −0.14 (−0.80, 0.52) |

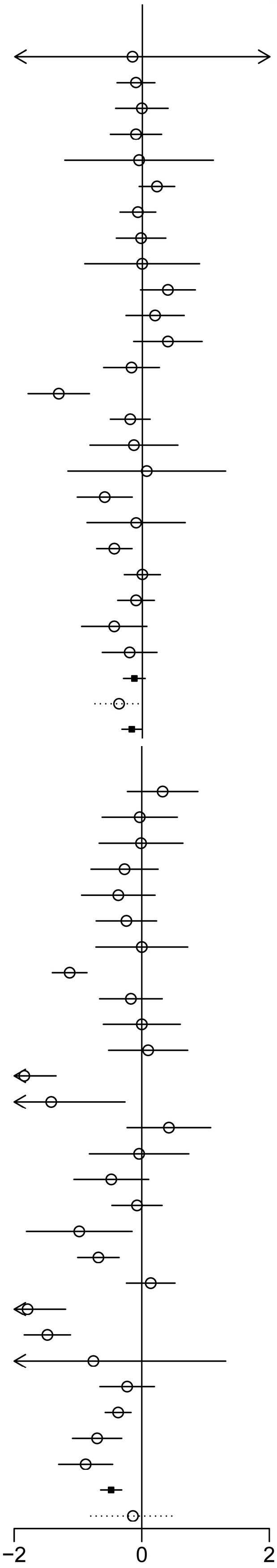

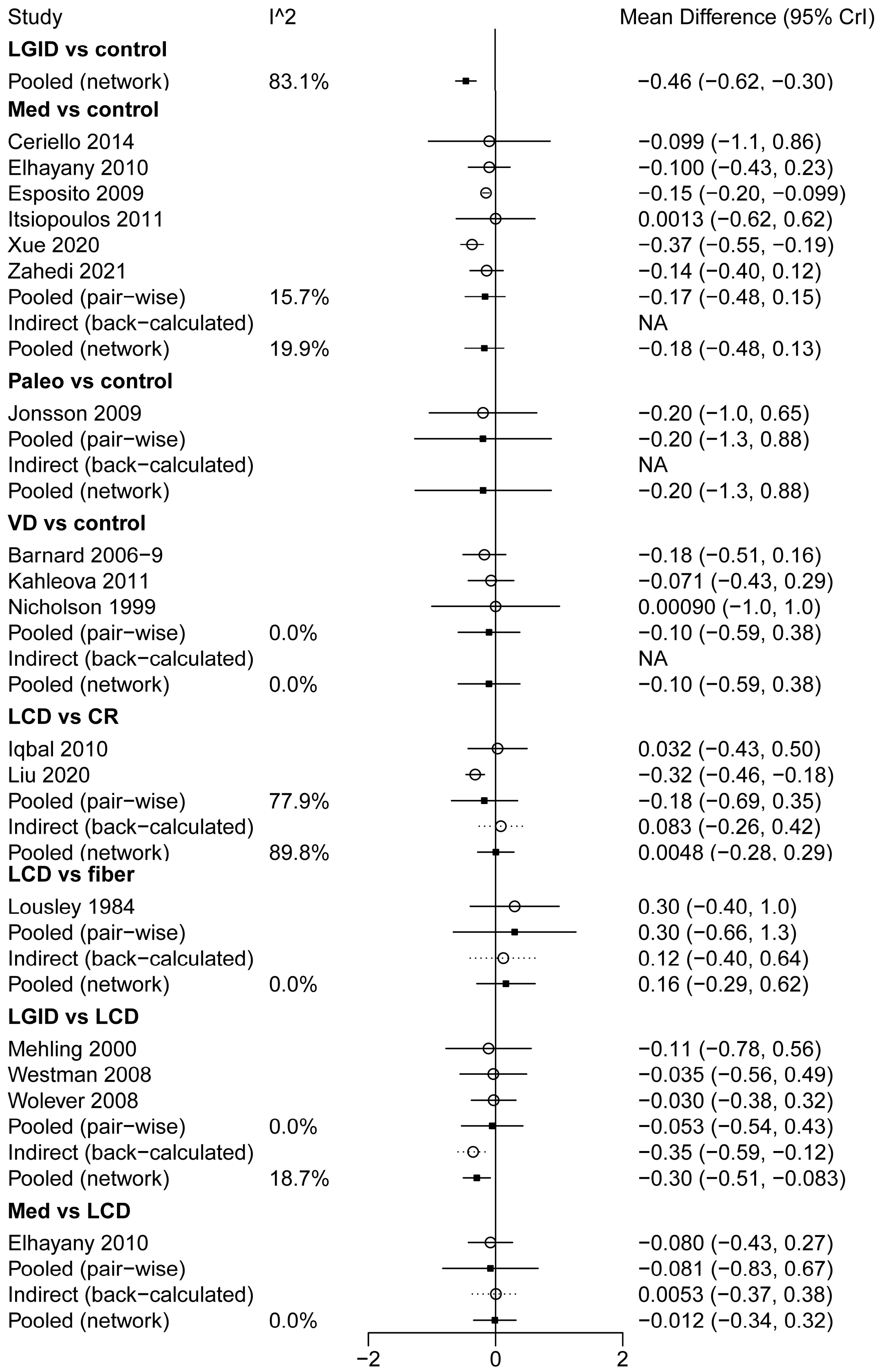
Study | I^2 | Mean Difference (95% CrI)
LGID vs control
Pooled (network) 83.1% −0.46 (−0.62, −0.30)
Med vs control
Ceriello 2014 −0.099 (−1.1, 0.86)
Elhayany 2010 −0.100 (−0.43, 0.23)
Esposito 2009 −0.15 (−0.20, −0.099)
Itsiopoulos 2011 0.0013 (−0.62, 0.62)
Xue 2020 −0.37 (−0.55, −0.19)
Zahedi 2021 −0.14 (−0.40, 0.12)
Pooled (pair−wise) 15.7% −0.17 (−0.48, 0.15)
Indirect (back−calculated) NA
Pooled (network) 19.9% −0.18 (−0.48, 0.13)
Paleo vs control
Jonsson 2009 −0.20 (−1.0, 0.65)
Pooled (pair−wise) −0.20 (−1.3, 0.88)
Indirect (back−calculated) NA
Pooled (network) −0.20 (−1.3, 0.88)
VD vs control
Barnard 2006−9 −0.18 (−0.51, 0.16)
Kahleova 2011 −0.071 (−0.43, 0.29)
Nicholson 1999 0.00090 (−1.0, 1.0)
Pooled (pair−wise) 0.0% −0.10 (−0.59, 0.38)
Indirect (back−calculated) NA
Pooled (network) 0.0% −0.10 (−0.59, 0.38)
LCD vs CR
Iqbal 2010 0.032 (−0.43, 0.50)
Liu 2020 −0.32 (−0.46, −0.18)
Pooled (pair−wise) 77.9% −0.18 (−0.69, 0.35)
Indirect (back−calculated) 0.083 (−0.26, 0.42)
Pooled (network) 89.8% 0.0048 (−0.28, 0.29)
LCD vs fiber
Lousley 1984 0.30 (−0.40, 1.0)
Pooled (pair−wise) 0.30 (−0.66, 1.3)
Indirect (back−calculated) 0.12 (−0.40, 0.64)
Pooled (network) 0.0% 0.16 (−0.29, 0.62)
LGID vs LCD
Mehling 2000 −0.11 (−0.78, 0.56)
Westman 2008 −0.035 (−0.56, 0.49)
Wolever 2008 −0.030 (−0.38, 0.32)
Pooled (pair−wise) 0.0% −0.053 (−0.54, 0.43)
Indirect (back−calculated) −0.35 (−0.59, −0.12)
Pooled (network) 18.7% −0.30 (−0.51, −0.083)
Med vs LCD
Elhayany 2010 −0.080 (−0.43, 0.27)
Pooled (pair−wise) −0.081 (−0.83, 0.67)
Indirect (back−calculated) 0.0053 (−0.37, 0.38)
Pooled (network) 0.0% −0.012 (−0.34, 0.32)
−2
0
2

**S9.12 Forest plots of LDL (*MD*, mmol/L)**

| Study | I^2 | Mean Difference (95% CrI) |
|---|---|---|
| **CR vs control** | | |
| Fang 2019 | | −0.82 (−0.98, −0.66) |
| Mohammadi 2017 | | −1.0 (−1.8, −0.24) |
| Mollentze 2019 | | 0.14 (−0.71, 0.99) |
| Ruggenenti 2017 | | −0.12 (−0.46, 0.22) |
| Ruggenenti 2022 | | −0.010 (−0.27, 0.25) |
| Uusitupa 1993 | | −0.40 (−0.83, 0.029) |
| Pooled (pair−wise) | 88.2% | −0.39 (−0.62, −0.15) |
| Indirect (back−calculated) | | 0.20 (−0.21, 0.61) |
| Pooled (network) | 90.5% | −0.24 (−0.44, −0.039) |
| **DASH vs control** | | |
| Azadbakht 2011 | | −0.37 (−0.67, −0.071) |
| Pooled (pair−wise) | | −0.37 (−0.88, 0.13) |
| Indirect (back−calculated) | | NA |
| Pooled (network) | | −0.37 (−0.90, 0.15) |
| **fiber vs control** | | |
| Chandalia 2000 | | −0.23 (−0.76, 0.30) |
| Coppell 2010 | | −0.19 (−0.49, 0.11) |
| Ikem 2007 | | −0.40 (−1.2, 0.35) |
| Pooled (pair−wise) | 0.0% | −0.24 (−0.60, 0.12) |
| Indirect (back−calculated) | | −0.21 (−1.2, 0.81) |
| Pooled (network) | 0.0% | −0.24 (−0.58, 0.11) |
| **HFD vs control** | | |
| Brehm 2009 | | 0.00019 (−0.38, 0.38) |
| Rock 2014 | | −0.055 (−0.34, 0.23) |
| Shige 2000 | | −0.30 (−0.86, 0.26) |
| Walker 1995 | | −0.011 (−0.29, 0.27) |
| Pooled (pair−wise) | 0.0% | −0.066 (−0.34, 0.20) |
| Indirect (back−calculated) | | NA |
| Pooled (network) | 0.0% | −0.066 (−0.35, 0.21) |
| **HPD vs control** | | |
| Gannon 2003 | | 0.0017 (−0.76, 0.77) |
| Krebs 2012 | | 0.030 (−0.16, 0.22) |
| Luger 2013 | | 0.021 (−0.49, 0.53) |
| Marco−Benedi 2020 | | −0.13 (−0.44, 0.19) |
| Parker 2002 | | −0.19 (−0.65, 0.27) |
| Pedersen 2014 | | 0.100 (−0.16, 0.36) |
| Stentz 2016 | | −0.52 (−0.87, −0.17) |
| Watson 2016 | | −0.20 (−0.48, 0.078) |
| Pooled (pair−wise) | 34.5% | −0.11 (−0.30, 0.082) |
| Indirect (back−calculated) | | NA |
| Pooled (network) | 34.7% | −0.11 (−0.31, 0.089) |

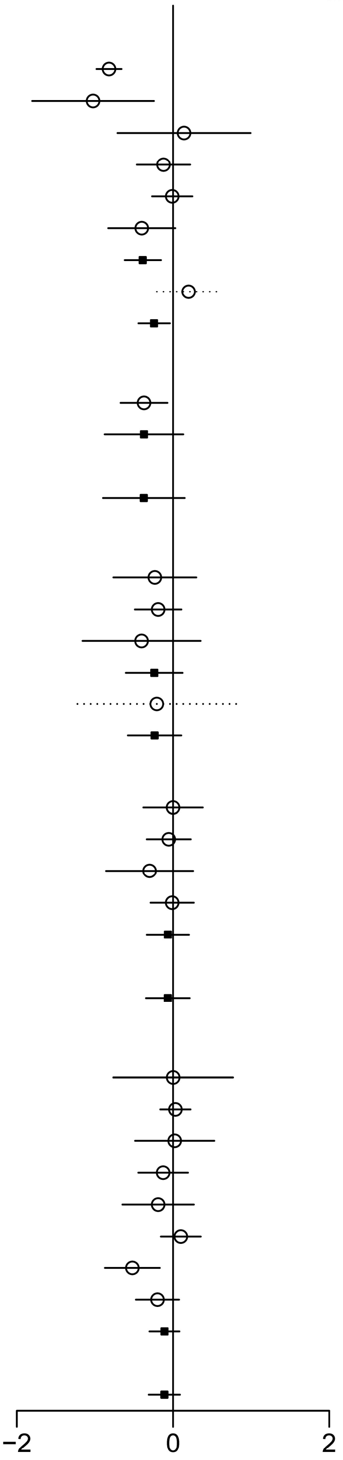

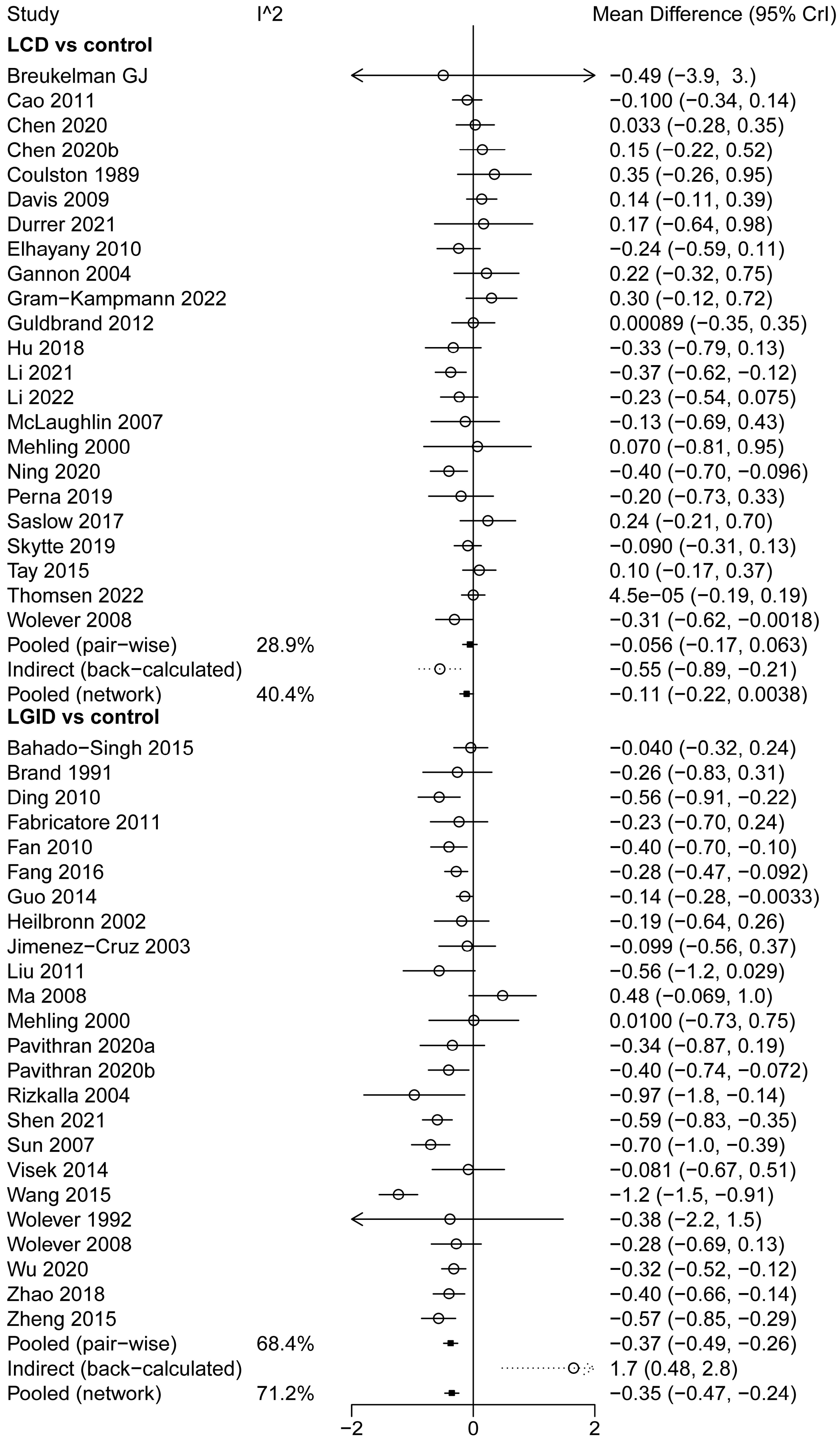
Study
I^2
Mean Difference (95% CrI)
LCD vs control
Breukelman GJ −0.49 (−3.9, 3.)
Cao 2011 −0.100 (−0.34, 0.14)
Chen 2020 0.033 (−0.28, 0.35)
Chen 2020b 0.15 (−0.22, 0.52)
Coulston 1989 0.35 (−0.26, 0.95)
Davis 2009 0.14 (−0.11, 0.39)
Durrer 2021 0.17 (−0.64, 0.98)
Elhayany 2010 −0.24 (−0.59, 0.11)
Gannon 2004 0.22 (−0.32, 0.75)
Gram−Kampmann 2022 0.30 (−0.12, 0.72)
Guldbrand 2012 0.00089 (−0.35, 0.35)
Hu 2018 −0.33 (−0.79, 0.13)
Li 2021 −0.37 (−0.62, −0.12)
Li 2022 −0.23 (−0.54, 0.075)
McLaughlin 2007 −0.13 (−0.69, 0.43)
Mehling 2000 0.070 (−0.81, 0.95)
Ning 2020 −0.40 (−0.70, −0.096)
Perna 2019 −0.20 (−0.73, 0.33)
Saslow 2017 0.24 (−0.21, 0.70)
Skytte 2019 −0.090 (−0.31, 0.13)
Tay 2015 0.10 (−0.17, 0.37)
Thomsen 2022 4.5e−05 (−0.19, 0.19)
Wolever 2008 −0.31 (−0.62, −0.0018)
Pooled (pair−wise) 28.9% −0.056 (−0.17, 0.063)
Indirect (back−calculated) −0.55 (−0.89, −0.21)
Pooled (network) 40.4% −0.11 (−0.22, 0.0038)
LGID vs control
Bahado−Singh 2015 −0.040 (−0.32, 0.24)
Brand 1991 −0.26 (−0.83, 0.31)
Ding 2010 −0.56 (−0.91, −0.22)
Fabricatore 2011 −0.23 (−0.70, 0.24)
Fan 2010 −0.40 (−0.70, −0.10)
Fang 2016 −0.28 (−0.47, −0.092)
Guo 2014 −0.14 (−0.28, −0.0033)
Heilbronn 2002 −0.19 (−0.64, 0.26)
Jimenez−Cruz 2003 −0.099 (−0.56, 0.37)
Liu 2011 −0.56 (−1.2, 0.029)
Ma 2008 0.48 (−0.069, 1.0)
Mehling 2000 0.0100 (−0.73, 0.75)
Pavithran 2020a −0.34 (−0.87, 0.19)
Pavithran 2020b −0.40 (−0.74, −0.072)
Rizkalla 2004 −0.97 (−1.8, −0.14)
Shen 2021 −0.59 (−0.83, −0.35)
Sun 2007 −0.70 (−1.0, −0.39)
Visek 2014 −0.081 (−0.67, 0.51)
Wang 2015 −1.2 (−1.5, −0.91)
Wolever 1992 −0.38 (−2.2, 1.5)
Wolever 2008 −0.28 (−0.69, 0.13)
Wu 2020 −0.32 (−0.52, −0.12)
Zhao 2018 −0.40 (−0.66, −0.14)
Zheng 2015 −0.57 (−0.85, −0.29)
Pooled (pair−wise) 68.4% −0.37 (−0.49, −0.26)
Indirect (back−calculated) 1.7 (0.48, 2.8)
Pooled (network) 71.2% −0.35 (−0.47, −0.24)
−2
0
2

| Study | I^2 | Mean Difference (95% Crl) |
|---|---|---|
| **Med vs control** | | |
| Ceriello 2014 | | −3.3e−05 (−1.1, 1.1) |
| Elhayany 2010 | | −0.17 (−0.50, 0.16) |
| Itsiopoulos 2011 | | −0.0011 (−0.41, 0.41) |
| Xue 2020 | | −0.38 (−0.62, −0.13) |
| Zahedi 2021 | | 0.36 (0.16, 0.56) |
| Pooled (pair−wise) | 82.3% | −0.033 (−0.28, 0.21) |
| Indirect (back−calculated) | | NA |
| Pooled (network) | 82.2% | −0.028 (−0.27, 0.21) |
| **Paleo vs control** | | |
| Jonsson 2009 | | −0.10 (−0.80, 0.60) |
| Pooled (pair−wise) | | −0.098 (−0.90, 0.71) |
| Indirect (back−calculated) | | NA |
| Pooled (network) | | −0.099 (−0.92, 0.73) |
| **VD vs control** | | |
| Barnard 2006−9 | | −0.11 (−0.42, 0.21) |
| Kahleova 2011 | | −0.030 (−0.34, 0.28) |
| Lee 2016 | | −0.049 (−0.32, 0.22) |
| Pooled (pair−wise) | 0.0% | −0.061 (−0.35, 0.23) |
| Indirect (back−calculated) | | NA |
| Pooled (network) | 0.0% | −0.061 (−0.36, 0.24) |
| **LCD vs CR** | | |
| Iqbal 2010 | | −0.060 (−0.50, 0.38) |
| Liu 2020 | | −0.35 (−0.74, 0.041) |
| Yamada 2014 | | −0.078 (−0.55, 0.39) |
| Pooled (pair−wise) | 0.0% | −0.17 (−0.51, 0.17) |
| Indirect (back−calculated) | | 0.33 (0.057, 0.60) |
| Pooled (network) | 68.2% | 0.14 (−0.078, 0.35) |
| **LCD vs fiber** | | |
| Lousley 1984 | | 0.10 (−0.62, 0.82) |
| Pooled (pair−wise) | | 0.10 (−0.74, 0.94) |
| Indirect (back−calculated) | | 0.13 (−0.26, 0.52) |
| Pooled (network) | 0.0% | 0.13 (−0.23, 0.48) |
| **LGID vs LCD** | | |
| Mehling 2000 | | −0.060 (−0.63, 0.51) |
| Westman 2008 | | −0.11 (−0.57, 0.35) |
| Wolever 2008 | | 0.030 (−0.28, 0.34) |
| Pooled (pair−wise) | 0.0% | −0.034 (−0.38, 0.31) |
| Indirect (back−calculated) | | −0.30 (−0.47, −0.13) |
| Pooled (network) | 25.2% | −0.24 (−0.40, −0.092) |
| **Med vs LCD** | | |
| Elhayany 2010 | | 0.070 (−0.21, 0.35) |
| Pooled (pair−wise) | | 0.070 (−0.42, 0.56) |
| Indirect (back−calculated) | | 0.084 (−0.21, 0.38) |
| Pooled (network) | 0.0% | 0.080 (−0.18, 0.33) |

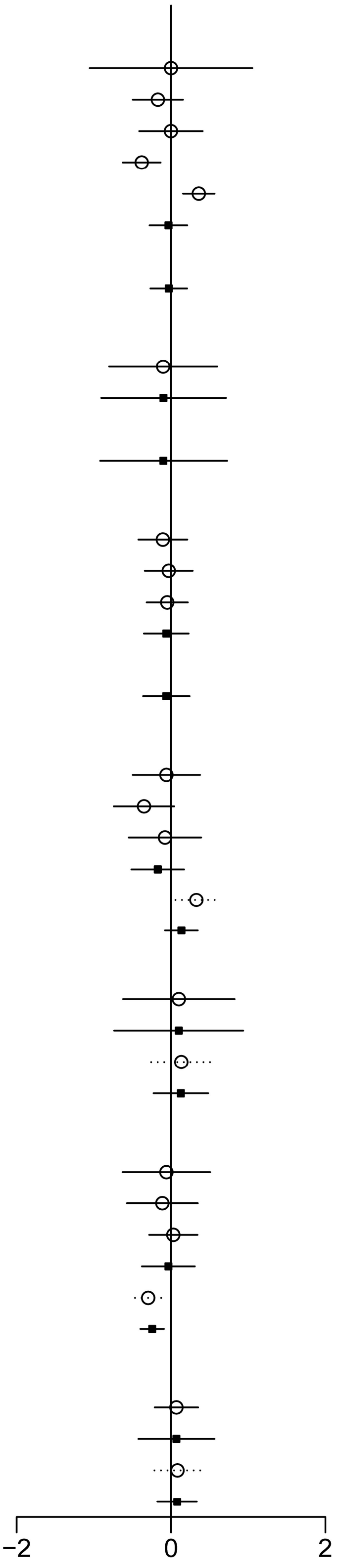

**S9.13 Forest plots of HDL (*MD*, mmol/L)**

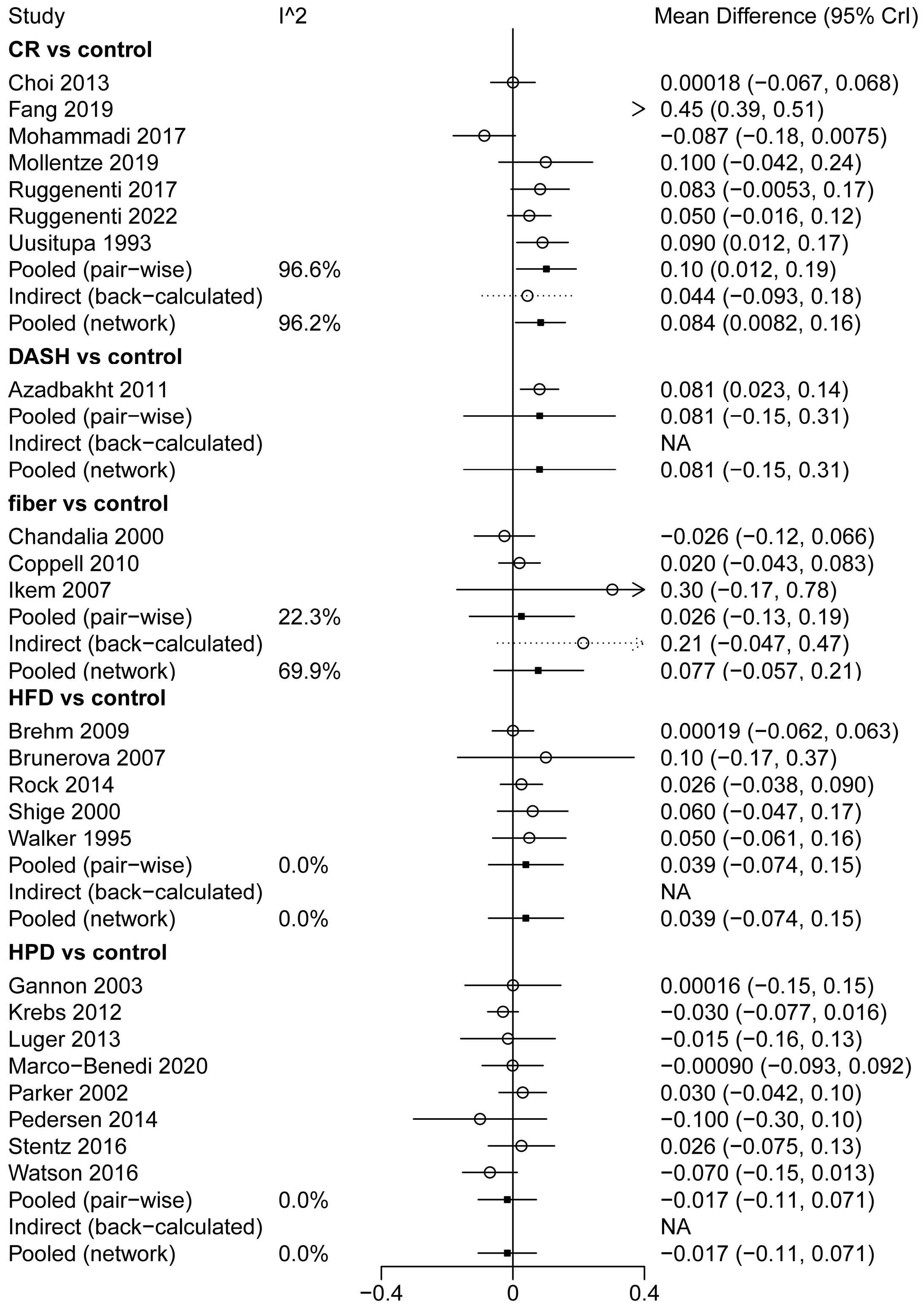

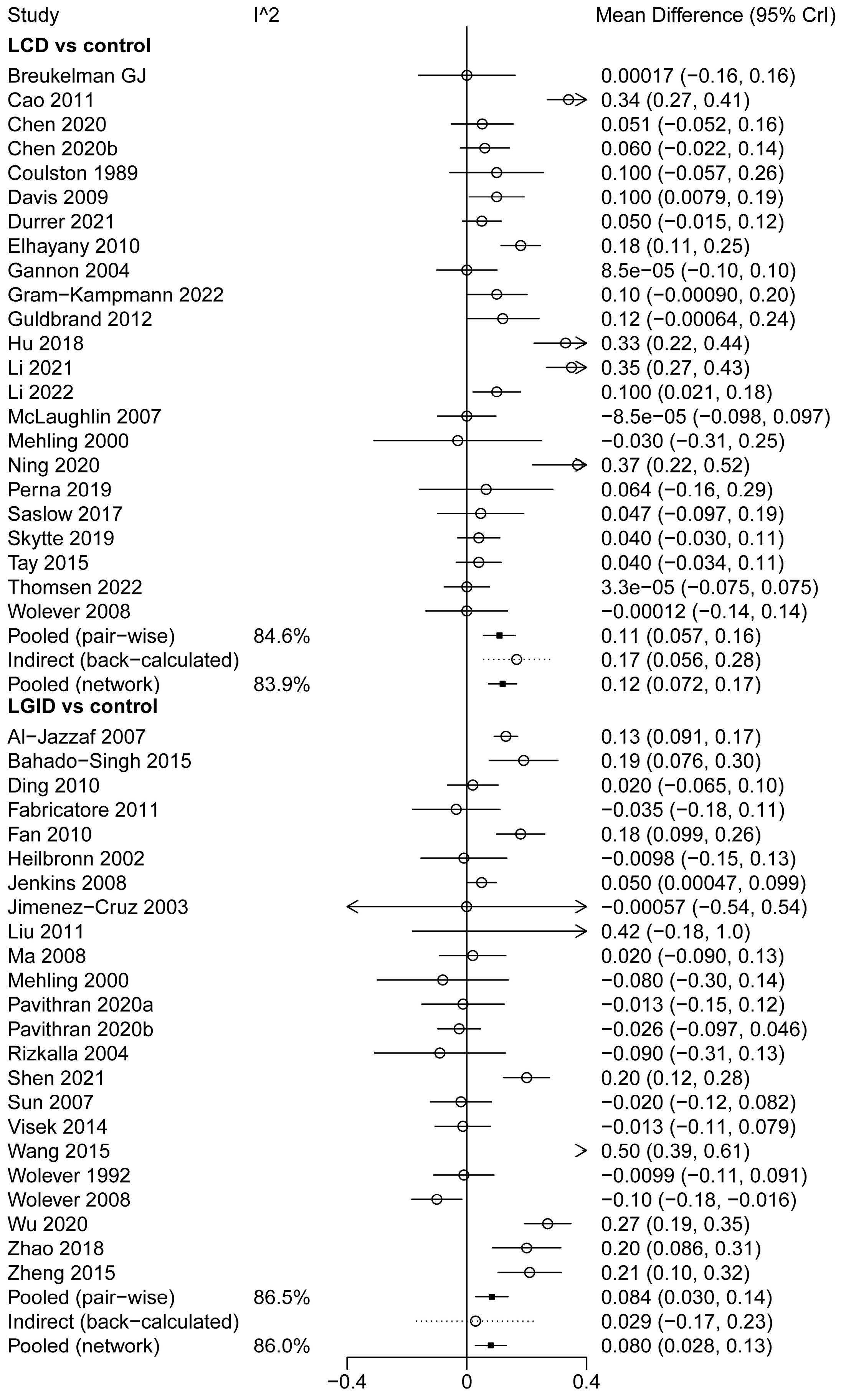
Study
I^2
Mean Difference (95% CrI)
LCD vs control
Breukelman GJ
0.00017 (−0.16, 0.16)
Cao 2011
0.34 (0.27, 0.41)
Chen 2020
0.051 (−0.052, 0.16)
Chen 2020b
0.060 (−0.022, 0.14)
Coulston 1989
0.100 (−0.057, 0.26)
Davis 2009
0.100 (0.0079, 0.19)
Durrer 2021
0.050 (−0.015, 0.12)
Elhayany 2010
0.18 (0.11, 0.25)
Gannon 2004
8.5e−05 (−0.10, 0.10)
Gram−Kampmann 2022
0.10 (−0.00090, 0.20)
Guldbrand 2012
0.12 (−0.00064, 0.24)
Hu 2018
0.33 (0.22, 0.44)
Li 2021
0.35 (0.27, 0.43)
Li 2022
0.100 (0.021, 0.18)
McLaughlin 2007
−8.5e−05 (−0.098, 0.097)
Mehling 2000
−0.030 (−0.31, 0.25)
Ning 2020
0.37 (0.22, 0.52)
Perna 2019
0.064 (−0.16, 0.29)
Saslow 2017
0.047 (−0.097, 0.19)
Skytte 2019
0.040 (−0.030, 0.11)
Tay 2015
0.040 (−0.034, 0.11)
Thomsen 2022
3.3e−05 (−0.075, 0.075)
Wolever 2008
−0.00012 (−0.14, 0.14)
Pooled (pair−wise)
84.6%
0.11 (0.057, 0.16)
Indirect (back−calculated)
0.17 (0.056, 0.28)
Pooled (network)
83.9%
0.12 (0.072, 0.17)
LGID vs control
Al−Jazzaf 2007
0.13 (0.091, 0.17)
Bahado−Singh 2015
0.19 (0.076, 0.30)
Ding 2010
0.020 (−0.065, 0.10)
Fabricatore 2011
−0.035 (−0.18, 0.11)
Fan 2010
0.18 (0.099, 0.26)
Heilbronn 2002
−0.0098 (−0.15, 0.13)
Jenkins 2008
0.050 (0.00047, 0.099)
Jimenez−Cruz 2003
−0.00057 (−0.54, 0.54)
Liu 2011
0.42 (−0.18, 1.0)
Ma 2008
0.020 (−0.090, 0.13)
Mehling 2000
−0.080 (−0.30, 0.14)
Pavithran 2020a
−0.013 (−0.15, 0.12)
Pavithran 2020b
−0.026 (−0.097, 0.046)
Rizkalla 2004
−0.090 (−0.31, 0.13)
Shen 2021
0.20 (0.12, 0.28)
Sun 2007
−0.020 (−0.12, 0.082)
Visek 2014
−0.013 (−0.11, 0.079)
Wang 2015
0.50 (0.39, 0.61)
Wolever 1992
−0.0099 (−0.11, 0.091)
Wolever 2008
−0.10 (−0.18, −0.016)
Wu 2020
0.27 (0.19, 0.35)
Zhao 2018
0.20 (0.086, 0.31)
Zheng 2015
0.21 (0.10, 0.32)
Pooled (pair−wise)
86.5%
0.084 (0.030, 0.14)
Indirect (back−calculated)
0.029 (−0.17, 0.23)
Pooled (network)
86.0%
0.080 (0.028, 0.13)
−0.4
0
0.4

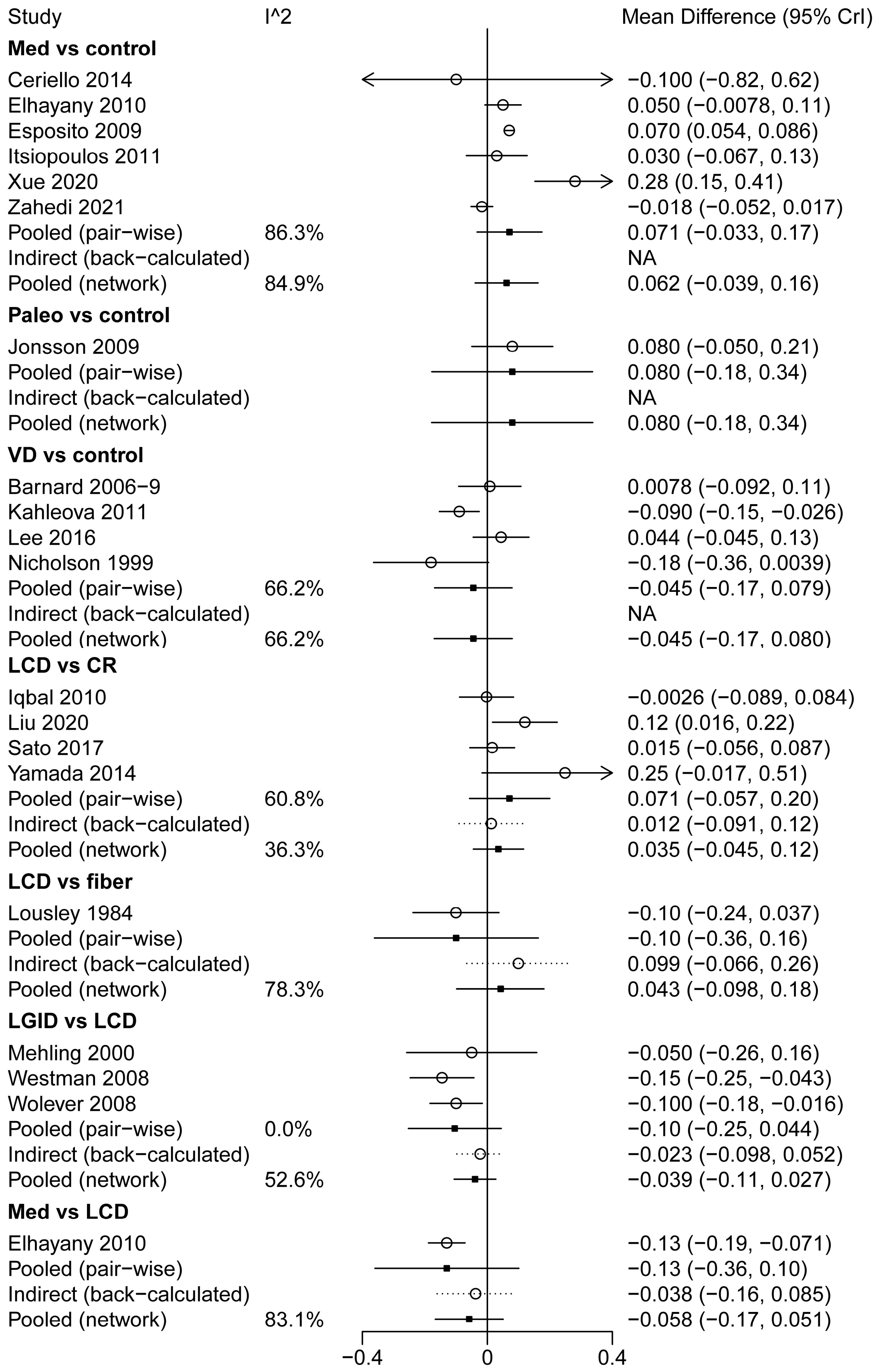

Study
I^2
Mean Difference (95% CrI)
Med vs control
Ceriello 2014 −0.100 (−0.82, 0.62)
Elhayany 2010 0.050 (−0.0078, 0.11)
Esposito 2009 0.070 (0.054, 0.086)
Itsiopoulos 2011 0.030 (−0.067, 0.13)
Xue 2020 0.28 (0.15, 0.41)
Zahedi 2021 −0.018 (−0.052, 0.017)
Pooled (pair−wise) 86.3% 0.071 (−0.033, 0.17)
Indirect (back−calculated) NA
Pooled (network) 84.9% 0.062 (−0.039, 0.16)
Paleo vs control
Jonsson 2009 0.080 (−0.050, 0.21)
Pooled (pair−wise) 0.080 (−0.18, 0.34)
Indirect (back−calculated) NA
Pooled (network) 0.080 (−0.18, 0.34)
VD vs control
Barnard 2006−9 0.0078 (−0.092, 0.11)
Kahleova 2011 −0.090 (−0.15, −0.026)
Lee 2016 0.044 (−0.045, 0.13)
Nicholson 1999 −0.18 (−0.36, 0.0039)
Pooled (pair−wise) 66.2% −0.045 (−0.17, 0.079)
Indirect (back−calculated) NA
Pooled (network) 66.2% −0.045 (−0.17, 0.080)
LCD vs CR
Iqbal 2010 −0.0026 (−0.089, 0.084)
Liu 2020 0.12 (0.016, 0.22)
Sato 2017 0.015 (−0.056, 0.087)
Yamada 2014 0.25 (−0.017, 0.51)
Pooled (pair−wise) 60.8% 0.071 (−0.057, 0.20)
Indirect (back−calculated) 0.012 (−0.091, 0.12)
Pooled (network) 36.3% 0.035 (−0.045, 0.12)
LCD vs fiber
Lousley 1984 −0.10 (−0.24, 0.037)
Pooled (pair−wise) −0.10 (−0.36, 0.16)
Indirect (back−calculated) 0.099 (−0.066, 0.26)
Pooled (network) 78.3% 0.043 (−0.098, 0.18)
LGID vs LCD
Mehling 2000 −0.050 (−0.26, 0.16)
Westman 2008 −0.15 (−0.25, −0.043)
Wolever 2008 −0.100 (−0.18, −0.016)
Pooled (pair−wise) 0.0% −0.10 (−0.25, 0.044)
Indirect (back−calculated) −0.023 (−0.098, 0.052)
Pooled (network) 52.6% −0.039 (−0.11, 0.027)
Med vs LCD
Elhayany 2010 −0.13 (−0.19, −0.071)
Pooled (pair−wise) −0.13 (−0.36, 0.10)
Indirect (back−calculated) −0.038 (−0.16, 0.085)
Pooled (network) 83.1% −0.058 (−0.17, 0.051)
−0.4
0
0.4

**S9.14 Forest plots of Attrition Rate (*RR*)**

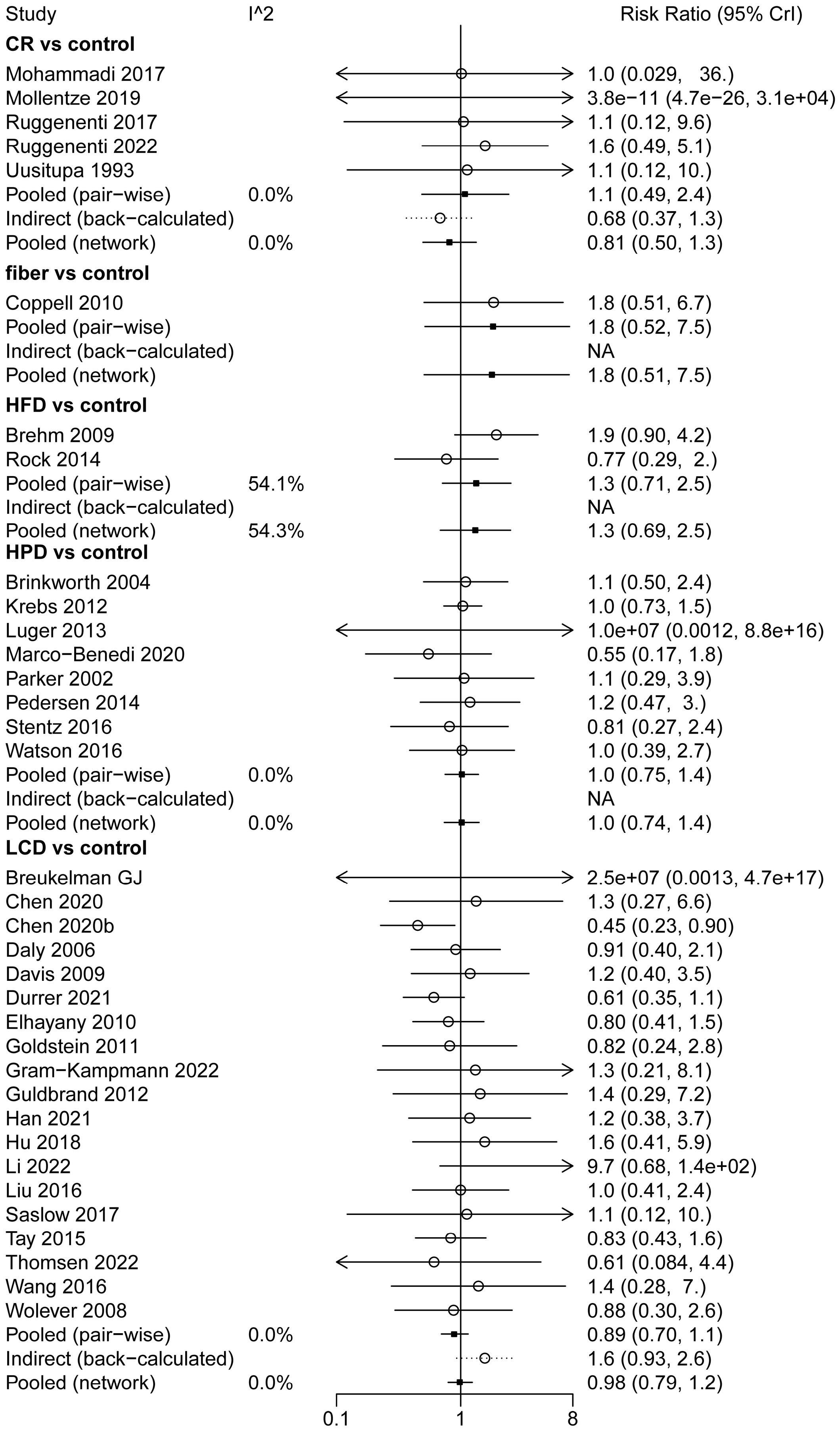

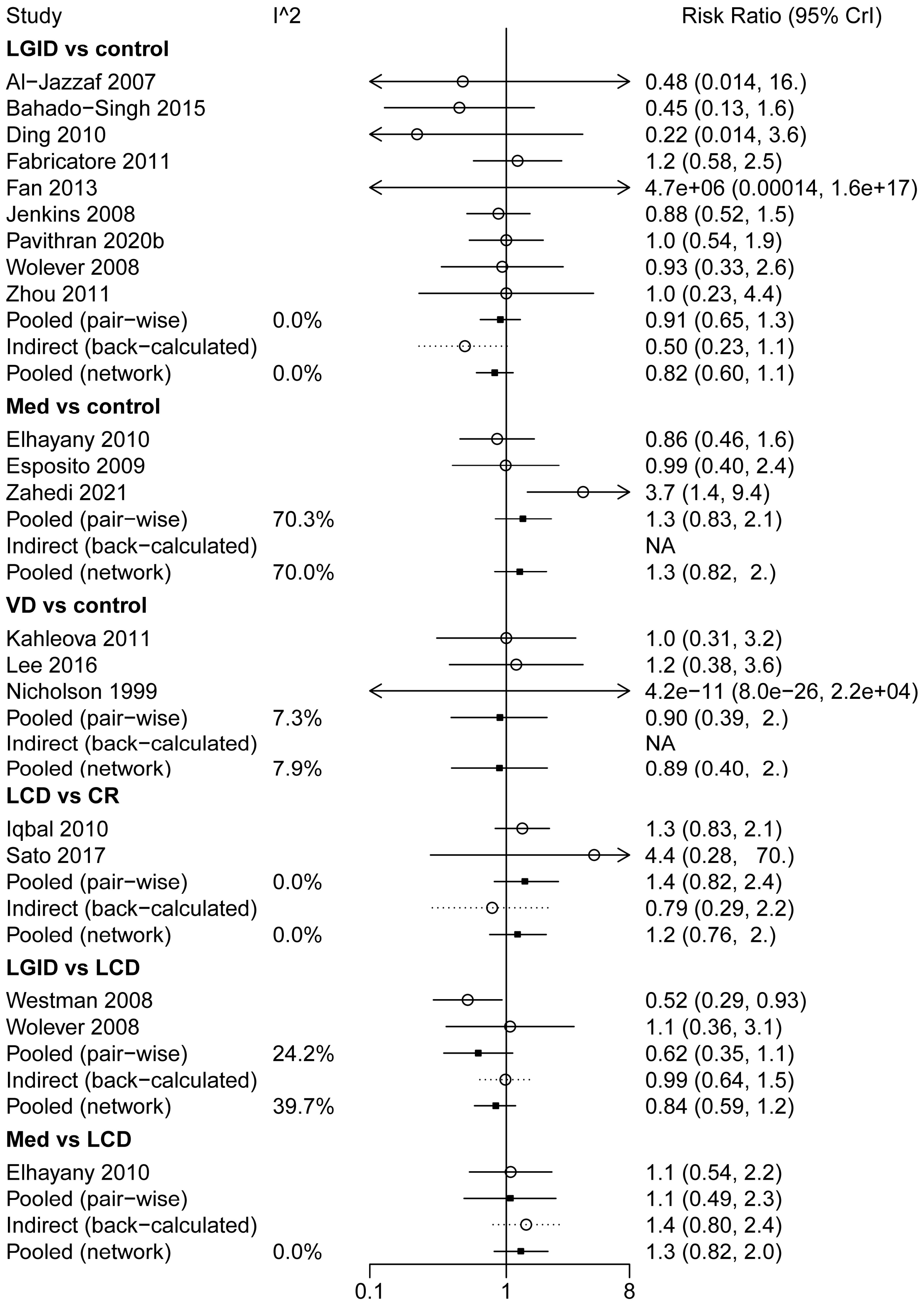
Study
I^2
Risk Ratio (95% CrI)
LGID vs control
Al−Jazzaf 2007
0.48 (0.014, 16.)
Bahado−Singh 2015
0.45 (0.13, 1.6)
Ding 2010
0.22 (0.014, 3.6)
Fabricatore 2011
1.2 (0.58, 2.5)
Fan 2013
4.7e+06 (0.00014, 1.6e+17)
Jenkins 2008
0.88 (0.52, 1.5)
Pavithran 2020b
1.0 (0.54, 1.9)
Wolever 2008
0.93 (0.33, 2.6)
Zhou 2011
1.0 (0.23, 4.4)
Pooled (pair−wise)
0.0%
0.91 (0.65, 1.3)
Indirect (back−calculated)
0.50 (0.23, 1.1)
Pooled (network)
0.0%
0.82 (0.60, 1.1)
Med vs control
Elhayany 2010
0.86 (0.46, 1.6)
Esposito 2009
0.99 (0.40, 2.4)
Zahedi 2021
3.7 (1.4, 9.4)
Pooled (pair−wise)
70.3%
1.3 (0.83, 2.1)
Indirect (back−calculated)
NA
Pooled (network)
70.0%
1.3 (0.82, 2.)
VD vs control
Kahleova 2011
1.0 (0.31, 3.2)
Lee 2016
1.2 (0.38, 3.6)
Nicholson 1999
4.2e−11 (8.0e−26, 2.2e+04)
Pooled (pair−wise)
7.3%
0.90 (0.39, 2.)
Indirect (back−calculated)
NA
Pooled (network)
7.9%
0.89 (0.40, 2.)
LCD vs CR
Iqbal 2010
1.3 (0.83, 2.1)
Sato 2017
4.4 (0.28, 70.)
Pooled (pair−wise)
0.0%
1.4 (0.82, 2.4)
Indirect (back−calculated)
0.79 (0.29, 2.2)
Pooled (network)
0.0%
1.2 (0.76, 2.)
LGID vs LCD
Westman 2008
0.52 (0.29, 0.93)
Wolever 2008
1.1 (0.36, 3.1)
Pooled (pair−wise)
24.2%
0.62 (0.35, 1.1)
Indirect (back−calculated)
0.99 (0.64, 1.5)
Pooled (network)
39.7%
0.84 (0.59, 1.2)
Med vs LCD
Elhayany 2010
1.1 (0.54, 2.2)
Pooled (pair−wise)
1.1 (0.49, 2.3)
Indirect (back−calculated)
1.4 (0.80, 2.4)
Pooled (network)
0.0%
1.3 (0.82, 2.0)
0.1
1
8

#### File S10: Heterogeneity and inconsistency test

**Table S10.1** Heterogeneity and inconsistency test for FPG

| Pairs | Number of Comparisons | Heterogeneity ($I^2$) | | Inconsistency test (*P* value) | |
|---|---|---|---|---|---|
| | | Paired | Network | Model | Node-splitting |
| CR vs control | 7 | 72.3%* | 77.8%* | 0.032* | 0.043* |
| DASH vs control | 1 | - | - | - | - |
| fiber vs control | 3 | 0.0% | 0.0% | 0.473 | 0.531 |
| HFD vs control | 5 | 57.0%* | 57.0%* | - | - |
| HPD vs control | 8 | 47.1% | 47.1% | - | - |
| LCD vs control | 24 | 87.7%* | 87.9%* | 0.029* | 0.085 |
| LGID vs control | 30 | 58.3%* | 58.3%* | - | 0.810 |
| Med vs control | 6 | 58.5%* | 65.2%* | - | - |
| Paleo vs control | 1 | - | 0.0% | - | - |
| VD vs control | 4 | 0.0% | 0.0% | - | - |
| LCD vs CR | 3 | 34.3% | 78.8%* | 0.040* | 0.045* |
| LCD vs fiber | 1 | - | 0.0% | 0.530 | 0.553 |
| LGID vs LCD | 2 | 0.0% | 0.0% | 0.955 | 0.967 |
| Med vs LCD | 1 | - | 26.3% | 0.513 | 0.515 |

* $I^2 > 50\%$ or *P* value < 0.05.

**Table S10.2** Heterogeneity and inconsistency test for $HbA_{1c}$

| Pairs | Number of Comparisons | Heterogeneity ($I^2$) | | Inconsistency test (*P* value) | |
|---|---|---|---|---|---|
| | | Paired | Network | Model | Node-splitting |
| CR vs control | 5 | 0.0% | 35.2% | 0.344 | 0.346 |
| DASH vs control | 1 | - | - | - | - |
| fiber vs control | 2 | 0.0% | 59.47%* | 0.050* | 0.058 |
| HFD vs control | 5 | 0.0% | 0.0% | - | - |
| HPD vs control | 8 | 0.0% | 0.0% | - | - |
| LCD vs control | 26 | 92.8%* | 93.02%* | 0.309 | 0.352 |
| LGID vs control | 28 | 75.1%* | 74.6%* | 0.134 | 0.132 |
| Med vs control | 6 | 46.2% | 47.5% | - | - |
| Paleo vs control | 1 | - | - | - | - |
| VD vs control | 4 | 0.0% | 0.0% | - | - |
| LCD vs CR | 4 | 52.3%* | 48.1% | 0.345 | 0.356 |
| LCD vs fiber | 1 | - | 77.4%* | 0.058 | 0.059 |
| LGID vs LCD | 2 | 0.0% | 51.4%* | 0.176 | 0.186 |
| Med vs LCD | 1 | - | 0.0% | 0.992 | 0.955 |

* $I^2 > 50\%$ or *P* value < 0.05.

**Table S10.3** Heterogeneity and inconsistency test for FIns

| Pairs | Number of Comparisons | Heterogeneity ($I^2$) | | Inconsistency test ($P$ value) | |
|---|---|---|---|---|---|
| | | Paired | Network | Model | Node-splitting |
| fiber vs control | 1 | - | 0.0% | 0.905 | 0.896 |
| HFD vs control | 3 | 56.0%* | 56.0%* | - | - |
| HPD vs control | 5 | 11.1% | 10.7% | - | - |
| LCD vs control | 13 | 89.8%* | 88.5%* | 0.531 | 0.324 |
| LGID vs control | 7 | 80.9%* | 76.2%* | 0.372 | 0.292 |
| Med vs control | 3 | 0.0% | 87.1%* | - | - |
| Paleo vs control | 1 | - | - | - | - |
| VD vs control | 1 | - | - | - | - |
| LCD vs fiber | 1 | - | 0.0% | 0.888 | 0.884 |
| LGID vs LCD | 1 | - | 64.2% | 0.310 | 0.289 |
| Med vs LCD | 1 | - | 25.5% | 0.441 | 0.483 |

* $I^2 > 50\%$ or $P$ value < 0.05.

**Table S10.4** Heterogeneity and inconsistency test for IR

| Pairs | Number of Comparisons | Heterogeneity ($I^2$) | | Inconsistency test ($P$ value) | |
|---|---|---|---|---|---|
| | | Paired | Network | Model | Node-splitting |
| HFD vs control | 1 | - | - | - | - |
| HPD vs control | 4 | 38.2% | 38.2% | - | - |
| LCD vs control | 7 | 33.7% | 33.1% | - | - |
| LGID vs control | 5 | 8.3% | 8.3% | - | - |
| Med vs control | 4 | 0.0% | 0.0% | - | - |
| Paleo vs control | 1 | - | - | - | - |
| Med vs LCD | 1 | - | 0.0% | 0.698 | 0.613 |

* $I^2 > 50\%$ or $P$ value < 0.05.

**Table S10.5** Heterogeneity and inconsistency test for weight

| Pairs | Number of Comparisons | Heterogeneity ($I^2$) | | Inconsistency test ($P$ value) | |
|---|---|---|---|---|---|
| | | Paired | Network | Model | Node-splitting |
| CR vs control | 6 | 99.5%* | 99.6%* | 0.007* | 0.012* |
| DASH vs control | 1 | - | - | - | - |
| fiber vs control | 2 | 0.0% | 0.0% | - | - |
| HFD vs control | 5 | 56.6%* | 56.8%* | - | - |
| HPD vs control | 8 | 71.9%* | 71.9%* | - | - |
| LCD vs control | 22 | 99.8%* | 99.7%* | 0.006* | 0.010* |
| LGID vs control | 15 | 82.8%* | 82.2%* | 0.715 | 0.523 |
| Med vs control | 3 | 1.3% | 0.0% | - | - |
| Paleo vs control | 1 | - | - | - | - |
| VD vs control | 2 | 0.0% | 0.0% | - | - |
| LCD vs CR | 4 | 0.0% | 88.1%* | 0.010* | 0.012* |
| LGID vs LCD | 3 | 51.6% | 28.3% | 0.895 | 0.863 |
| Med vs LCD | 1 | - | 45.2% | 0.693 | 0.778 |

* $I^2 > 50\%$ or $P$ value < 0.05.

**Table S10.6** Heterogeneity and inconsistency test for BMI

| Pairs | Number of Comparisons | Heterogeneity ($I^2$) | | Inconsistency test ($P$ value) | |
|---|---|---|---|---|---|
| | | Paired | Network | Model | Node-splitting |
| CR vs control | 6 | 83.2%* | 82.0%* | 0.583 | 0.565 |
| fiber vs control | 2 | 34.9% | 35.1% | - | - |
| HFD vs control | 2 | 0.0% | 0.0% | - | - |
| HPD vs control | 5 | 7.4% | 7.3% | - | - |
| LCD vs control | 20 | 99.8%* | 99.8%* | 0.444 | 0.370 |
| LGID vs control | 15 | 81.6%* | 80.2%* | 0.919 | 0.571 |
| Med vs control | 7 | 92.7%* | 92.7%* | - | - |
| Paleo vs control | 1 | - | - | - | - |
| VD vs control | 3 | 59.1%* | 59.2%* | - | - |
| LCD vs CR | 2 | 0.0% | 62.7%* | 0.579 | 0.563 |
| LGID vs LCD | 2 | 43.2% | 1.2% | 0.845 | 0.846 |
| Med vs LCD | 1 | - | 0.0% | 0.878 | 0.832 |

* $I^2 > 50\%$ or $P$ value < 0.05.

**Table S10.7** Heterogeneity and inconsistency test for WC

| Pairs | Number of Comparisons | Heterogeneity ($I^2$) | | Inconsistency test ($P$ value) | |
|---|---|---|---|---|---|
| | | Paired | Network | Model | Node-splitting |
| CR vs control | 5 | 82.3%* | 82.4%* | - | - |
| DASH vs control | 1 | - | - | - | - |
| fiber vs control | 2 | 0.0% | 0.0% | - | - |
| HFD vs control | 2 | 0.0% | 0.0% | - | - |
| HPD vs control | 3 | 70.7%* | 70.6%* | - | - |
| LCD vs control | 13 | 92.4%* | 91.7%* | 0.791 | 0.715 |
| LGID vs control | 11 | 46.1% | 41.2% | 0.819 | 0.805 |
| Med vs control | 2 | 0.0% | 58.4%* | - | - |
| Paleo vs control | 1 | - | - | - | - |
| VD vs control | 2 | 0.0% | 0.0% | - | - |
| LGID vs LCD | 2 | 0.0% | 0.0% | 0.683 | 0.638 |
| Med vs LCD | 1 | - | 36.0% | 0.604 | 0.680 |

* $I^2 > 50\%$ or $P$ value < 0.05.

**Table S10.8** Heterogeneity and inconsistency test for SBP

| Pairs | Number of Comparisons | Heterogeneity ($I^2$) | | Inconsistency test ($P$ value) | |
|---|---|---|---|---|---|
| | | Paired | Network | Model | Node-splitting |
| CR vs control | 5 | 66.4%* | 57.2%* | 0.670 | 0.645 |
| DASH vs control | 2 | 0.0% | 0.0% | - | - |
| fiber vs control | 1 | - | - | - | - |
| HFD vs control | 3 | 0.0% | 0.0% | - | - |
| HPD vs control | 7 | 53.6%* | 53.6%* | - | - |
| LCD vs control | 17 | 66.0%* | 63.9%* | 0.484 | 0.312 |
| LGID vs control | 10 | 60.8%* | 58.4%* | 0.753 | 0.404 |
| Med vs control | 3 | 68.0%* | 68.6%* | - | - |

| Pairs | Number of Comparisons | Heterogeneity ($I^2$) | | Inconsistency test ($P$ value) | |
|---|---|---|---|---|---|
| | | Paired | Network | Model | Node-splitting |
| Paleo vs control | 1 | - | - | - | - |
| VD vs control | 3 | 34.1% | 33.9% | - | - |
| LCD vs CR | 2 | 43.6% | 0.0% | 0.653 | 0.647 |
| LGID vs LCD | 3 | 30.1% | 10.8% | 0.663 | 0.631 |

* $I^2$ > 50% or $P$ value < 0.05.

**Table S10.9** Heterogeneity and inconsistency test for DBP

| Pairs | Number of Comparisons | Heterogeneity ($I^2$) | | Inconsistency test ($P$ value) | |
|---|---|---|---|---|---|
| | | Paired | Network | Model | Node-splitting |
| CR vs control | 5 | 69.0%* | 59.5%* | 0.532 | 0.484 |
| DASH vs control | 2 | 64.7%* | 64.6%* | - | - |
| fiber vs control | 1 | - | - | - | - |
| HFD vs control | 3 | 17.5% | 17.7% | - | - |
| HPD vs control | 7 | 66.2%* | 66.2%* | - | - |
| LCD vs control | 15 | 85.2%* | 84.3%* | 0.887 | 0.568 |
| LGID vs control | 9 | 46.7% | 36.4% | 0.445 | 0.874 |
| Med vs control | 3 | 89.4%* | 89.3%* | - | - |
| Paleo vs control | 1 | - | - | - | - |
| VD vs control | 3 | 19.0% | 19.3% | - | - |
| LCD vs CR | 2 | 15.1% | 0.0% | 0.499 | 0.488 |
| LGID vs LCD | 2 | 82.2%* | 71.7%* | 0.269 | 0.197 |

* $I^2$ > 50% or $P$ value < 0.05.

**Table S10.10** Heterogeneity and inconsistency test for TG

| Pairs | Number of Comparisons | Heterogeneity ($I^2$) | | Inconsistency test ($P$ value) | |
|---|---|---|---|---|---|
| | | Paired | Network | Model | Node-splitting |
| CR vs control | 5 | 11.9% | 0.0% | 0.378 | 0.346 |
| DASH vs control | 1 | - | - | - | - |
| fiber vs control | 3 | 2.0% | 1.9% | - | - |
| HFD vs control | 5 | 0.0% | 0.0% | - | - |
| HPD vs control | 7 | 0.0% | 0.0% | - | - |
| LCD vs control | 25 | 56.1%* | 53.5%* | 0.393 | 0.324 |
| LGID vs control | 27 | 72.2%* | 72.1%* | - | - |
| Med vs control | 6 | 65.7%* | 65.5%* | - | - |
| Paleo vs control | 1 | - | - | - | - |
| VD vs control | 4 | 61.7%* | 61.7%* | - | - |
| LCD vs CR | 4 | 0.0% | 0.0% | 0.358 | 0.346 |
| LGID vs LCD | 2 | 0.0% | 0.0% | 0.747 | 0.722 |
| Med vs LCD | 1 | - | 0.0% | 0.898 | 0.771 |

* $I^2$ > 50% or $P$ value < 0.05.

**Table S10.11** Heterogeneity and inconsistency test for TC

| Pairs | Number of Comparisons | Heterogeneity ($I^2$) | | Inconsistency test ($P$ value) | |
|---|---|---|---|---|---|
| | | Paired | Network | Model | Node-splitting |
| CR vs control | 7 | 50.7%* | 58.1%* | 0.423 | 0.413 |
| DASH vs control | 1 | - | - | - | - |
| fiber vs control | 3 | 0.0% | 0.0% | 0.737 | 0.748 |
| HFD vs control | 4 | 0.0% | 0.0% | - | - |
| HPD vs control | 7 | 17.0% | 17.1% | - | - |
| LCD vs control | 24 | 57.3%* | 57.4%* | 0.256 | 0.356 |
| LGID vs control | 27 | 83.5%* | 83.1%* | 0.326 | 0.506 |
| Med vs control | 6 | 15.7% | 19.9% | - | - |
| Paleo vs control | 1 | - | - | - | - |
| VD vs control | 3 | 0.0% | 0.0% | - | - |
| LCD vs CR | 2 | 77.9%* | 89.8%* | 0.411 | 0.418 |
| LCD vs fiber | 1 | - | 0.0% | 0.751 | 0.758 |
| LGID vs LCD | 3 | 0.0% | 18.7% | 0.272 | 0.257 |
| Med vs LCD | 1 | - | 0.0% | 0.840 | 0.861 |

* $I^2 > 50\%$ or $P$ value $< 0.05$.

**Table S10.12** Heterogeneity and inconsistency test for LDL

| Pairs | Number of Comparisons | Heterogeneity ($I^2$) | | Inconsistency test ($P$ value) | |
|---|---|---|---|---|---|
| | | Paired | Network | Model | Node-splitting |
| CR vs control | 6 | 88.3%* | 90.5%* | 0.013* | 0.026* |
| DASH vs control | 1 | - | - | - | - |
| fiber vs control | 3 | 0.0% | 0.0% | 0.945 | 0.947 |
| HFD vs control | 4 | 0.0% | 0.0% | - | - |
| HPD vs control | 8 | 35.1% | 35.1% | - | - |
| LCD vs control | 23 | 28.8% | 41.0% | 0.015* | 0.038* |
| LGID vs control | 24 | 68.4%* | 79.8%* | 0.005* | 0.599 |
| Med vs control | 5 | 82.3%* | 82.2%* | - | - |
| Paleo vs control | 1 | - | - | - | - |
| VD vs control | 3 | 0.0% | 0.0% | - | - |
| LCD vs CR | 3 | 0.0% | 68.1%* | 0.023* | 0.027* |
| LCD vs fiber | 1 | - | 0.0% | 0.941 | 0.941 |
| LGID vs LCD | 3 | 0.0% | 24.7% | 0.184 | 0.166 |
| Med vs LCD | 1 | - | 0.0% | 0.961 | 0.902 |

* $I^2 > 50\%$ or $P$ value $< 0.05$.

**Table S10.13** Heterogeneity and inconsistency test for HDL

| Pairs | Number of Comparisons | Heterogeneity ($I^2$) | | Inconsistency test ($P$ value) | |
|---|---|---|---|---|---|
| | | Paired | Network | Model | Node-splitting |
| CR vs control | 7 | 96.6%* | 96.2%* | 0.494 | 0.487 |
| DASH vs control | 1 | - | - | - | - |
| fiber vs control | 3 | 22.3% | 69.9%* | 0.228 | 0.207 |
| HFD vs control | 5 | 0.0% | 0.0% | - | - |

| Pairs | Number of Comparisons | Heterogeneity ($I^2$) | | Inconsistency test ($P$ value) | |
|---|---|---|---|---|---|
| | | Paired | Network | Model | Node-splitting |
| HPD vs control | 8 | 0.0% | 0.0% | - | - |
| LCD vs control | 23 | 84.6%* | 83.9%* | 0.356 | 0.534 |
| LGID vs control | 23 | 86.5%* | 86.0%* | 0.599 | 0.407 |
| Med vs control | 6 | 86.3%* | 84.9%* | - | - |
| Paleo vs control | 1 | - | - | - | - |
| VD vs control | 4 | 66.2%* | 66.2%* | - | - |
| LCD vs CR | 4 | 60.8%* | 36.3% | 0.488 | 0.494 |
| LCD vs fiber | 1 | - | 78.3%* | 0.208 | 0.203 |
| LGID vs LCD | 3 | 0.0% | 52.6%* | 0.336 | 0.310 |
| Med vs LCD | 1 | - | 83.1%* | 0.489 | 0.497 |

* $I^2 > 50\%$ or $P$ value < 0.05.

**Table S10.14** Heterogeneity and inconsistency test for Attrition Rate

| Pairs | Number of Comparisons | Heterogeneity ($I^2$) | | Inconsistency test ($P$ value) | |
|---|---|---|---|---|---|
| | | Paired | Network | Model | Node-splitting |
| CR vs control | 5 | 0.0% | 0.0% | 0.387 | 0.468 |
| fiber vs control | 1 | - | - | - | - |
| HFD vs control | 2 | 54.0%* | 53.9%* | - | - |
| HPD vs control | 8 | 0.0% | 0.0% | - | - |
| LCD vs control | 19 | 0.0% | 0.0% | 0.031* | 0.040* |
| LGID vs control | 9 | 0.0% | 0.0% | 0.093 | 0.114 |
| Med vs control | 3 | 70.4%* | 70.2%* | - | - |
| VD vs control | 3 | 6.1% | 8.1% | - | - |
| LCD vs CR | 2 | 0.0% | 0.0% | 0.343 | 0.409 |
| LGID vs LCD | 2 | 22.7% | 38.2% | 0.210 | 0.332 |
| Med vs LCD | 1 | - | 0.0% | 0.675 | 0.238 |

* $I^2 > 50\%$ or $P$ value < 0.05.

#### File S11: Meta-regression

We tested four groups of covariates separately, i.e., basic study information (publication year, origin, single/multicenter, parallel/crossover), study design (sample size, duration of the intervention, intensity, extra prescribed caloric restriction, additional use of antihyperglycemic medication, insulin and extra advice on exercise), baseline characteristics (sex ratio, mean age, weight, BMI, duration of T2DM/PreD), and macronutrients intake (TEI, protein, fat and carbohydrate). Meta-regression was conducted using R package "gemtc" with a linear model, and coefficients were set as exchangeable.
Covariates with significant coefficients (95% *CrI* did not contain the null value) were displayed below:
(see the next page)

**Table S11.1 Overview of Meta-regression**

| Outcome variables | Covariates* | | | | | | | | | | | | | | | | | | | | |
|---|---|---|---|---|---|---|---|---|---|---|---|---|---|---|---|---|---|---|---|---|---|
| | size | weeks | center | design | CR | intensity | medication | insulin | exercise | year | origin | sex ratio | baseline | age | weight | BMI | duration | energy | protein | fat | carbohydrate |
| FPG | a+ | | | | | | | | a+ | f+ | fg- | | afghk+ | | | | | | fg+ | | |
| $HbA_{1c}$ | | | | | | | | | | | g- | | f+ | | | | | | f+ | | |
| FIns | | | | | | | | | | | | | | f- | | | | | | f+ | |
| IR | | | | | | | | | | | | | | | | | | | | | |
| Weight | af+ | | | | f+ | | | | | f+ | a- | | | | NA | | | f- | a+ | | a- |
| BMI | | | | | | | | | | | | | f+ | | | NA | | | f+ | | |
| WC | | | | | | | | | | | | | | | | | | f- | | | f- |
| SBP | | | | | | | | | | | | | | | | | | | f+ | | |
| DBP | | | | | | | | | f+ | | | | a+ | | | | | | | | |
| TG | | | | | | | | | | | g- | | gh+ | | | | | | f+ | | g+ |
| TC | g+ | | | | | g- | | | | | fg- | | | | | g- | | | | g- | |
| LDL | a+ | | | | | | | | | | afghk- | | afk+ | a-g+ | g- | | | | | a- | a+ |
| HDL | a+ | | | | | fg- | | | a+ | g- | afghk- | | | | g- | gk- | | | | | g+ |

a. CR; b. DASH; c. fiber; d. HFD; e. HPD; f. LCD; g. LGID; h. Med; i, Paleo; j, VD; k, coefficient of all comparisons; +, positively correlated with the efficacy; -, negatively correlated with the efficacy; NA, not applicable. All data were for intervention vs control.

* size: sample size of the intervention arm; weeks: duration of the intervention in weeks; center: dichotomous, single center = 0, multicenter = 1; design: dichotomous, parallel = 0, crossover = 1; CR: ordinal, no extra caloric restriction = 0, mild restriction (500 to 600-kcal negative balance) = 1, moderate restriction (600 to 900) = 2, severe restriction (greater than 900) = 3; intensity: ordinal, no intervention = 1, only consultation = 2, providing menus and/or reported 24-hour recall of diets = 3, providing prepackaged or prepared food = 4, metabolic ward = 5; medication/insulin/exercise: dichotomous, no/not reported = 0, reported = 1; year: year of publication; origin: dichotomous, from China = 0, from other countries = 1; sex ratio: female percentage; baseline: baseline of the outcome variable; age: mean, in years old; weight: mean, kg; BMI: mean, $kg/m^2$; duration: duration of T2DM/PreD, years; energy: total energy intake as prescribed or reported, kcal/d; protein/fat/carbohydrate: as prescribed or reported, TEI%.

### File S12: Sensitivity analysis

#### S12.1 Sensitivity analysis

(Only effect sizes that were not robust were listed below)

**Table S12.1** Sensitivity analysis 1

| Outcome | Excluded study | Arms of the study | | | Adjusted Effect Size | |
|---|---|---|---|---|---|---|
| | | Arm 1 | Arm 2 | Arm 3 | Comparisons | *MD* [95% *CrI*] |
| FPG | Fang 2019 | CR | control | - | CR vs control | -0.523 [-1.110, 0.064] |
| | Ikem 2007 | fiber | control | - | fiber vs control | -0.984 [-2.223, 0.253] |
| $HbA_{1c}$ | Liu 2020 | LCD | CR | - | CR vs control | -0.425 [-0.845, -0.004] |
| | Chandalia 2000 | fiber | control | - | fiber vs control | -0.950 [-1.919, -0.001] |
| | Wang 2009b | LCD | control | - | fiber vs control | -0.672 [-1.346, -0.003] |
| | Elhayany 2010 | LCD | Med | control | Med vs control | -0.507 [-1.019, 0.010] |
| | Esposito 2009 | Med | control | - | Med vs control | -0.476 [-0.972, 0.022] |
| | Xue 2020 | Med | control | - | Med vs control | -0.463 [-0.953, 0.026] |
| | Zahedi 2021 | Med | control | - | Med vs control | -0.351 [-0.835, 0.133] |
| FIns | Li 2022 | LCD | control | - | fiber vs control | -0.205 [-0.379, -0.026] |
| | | | | | LCD vs control | -0.068 [-0.145, 0.008] |
| | Bahado-Singh 2015 | LGID | control | | LGID vs control | -0.101 [-0.237, 0.035] |
| | Liu 2011 | LGID | control | - | LGID vs control | -0.114 [-0.260, 0.030] |
| | Pavithran 2020b | LGID | control | - | LGID vs control | -0.123 [-0.251, 0.009] |
| IR | Marco-Benedí 2020 | HPD | control | - | HPD vs control | -0.142 [-0.351, 0.062] |
| | Skytte 2019 | LCD | control | - | LCD vs control | -0.122 [-0.210, -0.026] |
| | | | | | Med vs control | -0.102 [-0.185, -0.005] |
| | Watson 2016 | HPD | control | - | LCD vs control | -0.086 [-0.166, -0.004] |
| | Guo 2014 | LGID | control | - | LGID vs control | -0.152 [-0.352, 0.039] |
| | Liu 2016 | LCD | control | | Med vs control | -0.096 [-0.175, -0.001] |
| WC | Durrer 2021 | LCD | control | - | DASH vs control | -4.800 [-9.190, -0.423] |
| | He 2017 | LGID | control | - | LGID vs control | -1.889 [-3.809, 0.034] |
| | Wang 2015 | LGID | control | - | LGID vs control | -1.657 [-3.498, 0.187] |
| SBP* | | | | | | |
| DBP* | | | | | | |
| TG | Ceriello 2014 | Med | control | - | Med vs control | -0.212 [-0.422, -0.003] |
| | Wang 2009a | LGID | control | - | Med vs control | -0.201 [-0.381, -0.021] |
| | Wolever 2008 | LGID | LCD | control | Med vs control | -0.203 [-0.398, -0.007] |
| | Zahedi 2021 | Med | control | - | Med vs control | -0.283 [-0.509, -0.056] |
| TC | Li 2021 | LCD | control | - | LCD vs control | -0.124 [-0.272, 0.025] |
| | Liu 2020 | LCD | CR | - | LCD vs control | -0.149 [-0.304, 0.005] |
| | Ning 2020 | LCD | control | - | LCD vs control | -0.148 [-0.302, 0.008] |
| | Skytte 2019 | LCD | control | - | LCD vs control | -0.152 [-0.308, 0.005] |
| | Wang 2018 | LCD | control | - | LCD vs control | -0.155 [-0.311, 0.001] |
| | Wolever 2008 | LCD | LGID | control | LCD vs control | -0.156 [-0.313, 0.002] |
| LDL | Fang 2019 | CR | control | - | CR vs control | -0.094 [-0.304, 0.114] |
| | Mohammadi 2017 | CR | control | - | CR vs control | -0.201 [-0.406, 0.010] |

| Outcome | Excluded study | Arms of the study | | | Adjusted Effect Size | |
|---|---|---|---|---|---|---|
| | | Arm 1 | Arm 2 | Arm 3 | Comparisons | *MD* [95% *CrI*] |
| | Chen 2020a | LCD | control | - | LCD vs control | -0.114 [-0.228, -0.000] |
| | Chen 2020b | LCD | control | - | LCD vs control | -0.119 [-0.231, -0.005] |
| | Coulston 1989 | LCD | control | - | LCD vs control | -0.119 [-0.230, -0.007] |
| | Davis 2009 | LCD | control | - | LCD vs control | -0.121 [-0.234, -0.007] |
| | Durrer 2021 | LCD | control | - | LCD vs control | -0.112 [-0.223, -0.001] |
| | Gannon 2004 | LCD | control | - | LCD vs control | -0.117 [-0.228, -0.004] |
| | Gram-Kampmann 2022 | LCD | control | - | LCD vs control | -0.123 [-0.235, -0.011] |
| | Ma 2008 | LGID | control | - | LCD vs control | -0.111 [-0.218, -0.002] |
| | Ruggenenti 2022 | CR | control | - | LCD vs control | -0.113 [-0.223, -0.002] |
| | Saslow 2014/7 | LCD | control | - | LCD vs control | -0.120 [-0.231, -0.007] |
| | Tay 2015 | LCD | control | - | LCD vs control | -0.119 [-0.232, -0.004] |
| | Wang 2015 | LGID | control | - | LCD vs control | -0.106 [-0.208, -0.003] |
| | Zahedi 2021 | Med | control | - | LCD vs control | -0.114 [-0.221, -0.005] |
| HDL | Fang 2019 | CR | control | - | CR vs control | 0.041 [-0.032, 0.113] |

* The effect size was not robust in nearly all comparisons, and relevant studies were of a large number.
Blue cells: the intervention was of significant beneficial effect before adjustment; Red cells: the intervention did not show significant beneficial effect before adjustment.

**S12.2 Models**

We tested a consistency model and an unrelated study effect (USE) model, both with random and fixed effect models. Results were presented below: (see the next page)

**Table S12.2** Models

| Intervention compared to control | Consistent Random Effect Model | | | Consistent Fixed Effect Model | | | USE Random Effect Model | | | USE Fixed Effect Model | | |
|---|---|---|---|---|---|---|---|---|---|---|---|---|
| | Mean | Lower 95% Limit | Upper 95% Limit | Mean | Lower 95% Limit | Upper 95% Limit | Mean | Lower 95% Limit | Upper 95% Limit | Mean | Lower 95% Limit | Upper 95% Limit |
| FPG (*MD*, mmol/L) | | | | | | | | | | | | |
| CR | -0.811 | -1.368 | -0.251 | -0.913 | -1.209 | -0.618 | -0.918 | -1.918 | 0.083 | -0.921 | -1.928 | 0.086 |
| DASH | -0.923 | -2.539 | 0.689 | -0.922 | -1.921 | 0.079 | -1.367 | -2.410 | -0.328 | -1.369 | -2.405 | -0.332 |
| fiber | -1.257 | -2.320 | -0.203 | -1.175 | -1.948 | -0.401 | -0.307 | -1.483 | 0.867 | -0.306 | -1.479 | 0.870 |
| HFD | -0.005 | -0.785 | 0.779 | 0.017 | -0.497 | 0.531 | -0.104 | -1.780 | 1.571 | -0.098 | -1.778 | 1.571 |
| HPD | -0.294 | -0.884 | 0.291 | -0.312 | -0.615 | -0.008 | -0.165 | -1.257 | 0.925 | -0.167 | -1.258 | 0.924 |
| LCD | -0.817 | -1.126 | -0.510 | -0.707 | -0.832 | -0.582 | -1.071 | -4.603 | 2.406 | -1.082 | -4.613 | 2.483 |
| LGID | -0.936 | -1.219 | -0.651 | -0.931 | -1.057 | -0.805 | -1.095 | -2.918 | 0.729 | -1.099 | -2.926 | 0.729 |
| Med | -0.454 | -1.063 | 0.156 | -0.668 | -0.876 | -0.460 | -0.197 | -1.472 | 1.073 | -0.199 | -1.470 | 1.069 |
| Paleo | -0.505 | -2.182 | 1.168 | -0.500 | -1.602 | 0.601 | -0.668 | -2.387 | 1.067 | -0.668 | -2.398 | 1.061 |
| VD | -0.636 | -1.560 | 0.290 | -0.599 | -1.220 | 0.020 | -0.952 | -2.072 | 0.174 | -0.951 | -2.078 | 0.178 |
| $HbA_{1c}$ (*MD*, %) | | | | | | | | | | | | |
| CR | -0.342 | -0.755 | 0.072 | -0.384 | -0.558 | -0.211 | -1.200 | -1.400 | -1.000 | -1.200 | -1.399 | -1.001 |
| DASH | -1.200 | -2.174 | -0.227 | -1.200 | -1.401 | -1.000 | -0.491 | -0.965 | -0.015 | -0.490 | -0.966 | -0.013 |
| fiber | -0.736 | -1.519 | 0.040 | -0.562 | -0.996 | -0.129 | -0.201 | -0.701 | 0.301 | -0.200 | -0.699 | 0.299 |
| HFD | -0.107 | -0.628 | 0.415 | -0.166 | -0.434 | 0.103 | -0.901 | -2.195 | 0.392 | -0.897 | -2.195 | 0.399 |
| HPD | -0.182 | -0.570 | 0.205 | -0.134 | -0.274 | 0.004 | 0.099 | -0.518 | 0.717 | 0.100 | -0.514 | 0.719 |
| LCD | -0.668 | -0.880 | -0.460 | -0.372 | -0.429 | -0.315 | -0.464 | -7.531 | 6.894 | -0.442 | -8.199 | 7.086 |
| LGID | -0.712 | -0.930 | -0.493 | -0.623 | -0.710 | -0.535 | -0.303 | -1.158 | 0.549 | -0.300 | -1.152 | 0.547 |
| Med | -0.463 | -0.900 | -0.025 | -0.416 | -0.528 | -0.303 | -0.399 | -1.084 | 0.287 | -0.399 | -1.086 | 0.286 |
| Paleo | -0.398 | -1.500 | 0.708 | -0.400 | -0.957 | 0.157 | -0.100 | -0.668 | 0.468 | -0.101 | -0.669 | 0.467 |
| VD | -0.324 | -0.897 | 0.245 | -0.313 | -0.539 | -0.087 | -0.099 | -1.467 | 1.267 | -0.103 | -1.469 | 1.256 |
| FIns (*PMD*) | | | | | | | | | | | | |
| fiber | -0.206 | -0.462 | 0.052 | -0.216 | -0.286 | -0.147 | -0.355 | -0.533 | -0.176 | -0.354 | -0.533 | -0.176 |
| HFD | -0.049 | -0.258 | 0.159 | -0.043 | -0.171 | 0.085 | -0.100 | -0.314 | 0.115 | -0.101 | -0.315 | 0.111 |
| HPD | -0.091 | -0.261 | 0.082 | -0.130 | -0.226 | -0.033 | -0.178 | -3.392 | 3.023 | -0.182 | -3.397 | 3.028 |
| LCD | -0.119 | -0.215 | -0.021 | -0.195 | -0.235 | -0.155 | -0.110 | -0.493 | 0.273 | -0.109 | -0.494 | 0.276 |
| LGID | -0.138 | -0.266 | -0.010 | -0.131 | -0.165 | -0.097 | -0.216 | -0.286 | -0.146 | -0.216 | -0.286 | -0.145 |

| Intervention compared to control | Consistent Random Effect Model | | | Consistent Fixed Effect Model | | | USE Random Effect Model | | | USE Fixed Effect Model | | |
|---|---|---|---|---|---|---|---|---|---|---|---|---|
| | Mean | Lower 95% Limit | Upper 95% Limit | Mean | Lower 95% Limit | Upper 95% Limit | Mean | Lower 95% Limit | Upper 95% Limit | Mean | Lower 95% Limit | Upper 95% Limit |
| Med | -0.075 | -0.255 | 0.104 | -0.043 | -0.060 | -0.026 | -0.160 | -0.412 | 0.091 | -0.159 | -0.410 | 0.092 |
| Paleo | 0.021 | -0.364 | 0.407 | 0.020 | -0.241 | 0.281 | 0.112 | -0.049 | 0.275 | 0.112 | -0.050 | 0.274 |
| VD | -0.066 | -0.456 | 0.325 | -0.066 | -0.335 | 0.205 | 0.037 | -0.118 | 0.192 | 0.037 | -0.118 | 0.191 |
| IR (*PMD*) | | | | | | | | | | | | |
| HFD | -0.153 | -0.437 | 0.130 | -0.152 | -0.396 | 0.091 | -0.192 | -0.730 | 0.346 | -0.191 | -0.730 | 0.346 |
| HPD | -0.217 | -0.366 | -0.067 | -0.221 | -0.343 | -0.099 | -0.152 | -0.396 | 0.093 | -0.152 | -0.396 | 0.093 |
| LCD | -0.085 | -0.173 | 0.002 | -0.093 | -0.147 | -0.039 | -0.257 | -0.741 | 0.226 | -0.258 | -0.744 | 0.225 |
| LGID | -0.156 | -0.278 | -0.041 | -0.155 | -0.208 | -0.101 | -0.031 | -0.214 | 0.153 | -0.032 | -0.214 | 0.152 |
| Med | -0.095 | -0.191 | 0.010 | -0.111 | -0.150 | -0.072 | 0.005 | -0.164 | 0.174 | 0.005 | -0.163 | 0.173 |
| Paleo | -0.001 | -0.287 | 0.287 | 0.000 | -0.249 | 0.249 | -0.119 | -0.161 | -0.076 | -0.119 | -0.161 | -0.077 |
| weight (*MD*, kg) | | | | | | | | | | | | |
| CR | -4.083 | -6.131 | -2.012 | -8.559 | -8.912 | -8.208 | -1.913 | -3.199 | -0.627 | -1.908 | -3.195 | -0.617 |
| DASH | -2.991 | -9.053 | 3.044 | -3.002 | -4.860 | -1.141 | -2.999 | -4.856 | -1.132 | -3.003 | -4.860 | -1.140 |
| fiber | -0.871 | -5.255 | 3.509 | -1.026 | -2.595 | 0.534 | -1.401 | -3.761 | 0.951 | -1.398 | -3.772 | 0.966 |
| HFD | -0.621 | -3.354 | 2.119 | -0.228 | -0.937 | 0.479 | -0.107 | -2.684 | 2.458 | -0.101 | -2.662 | 2.458 |
| HPD | -0.493 | -2.722 | 1.729 | 0.299 | -0.461 | 1.059 | -0.199 | -2.042 | 1.639 | -0.198 | -2.030 | 1.630 |
| LCD | -3.001 | -4.248 | -1.744 | -6.659 | -6.756 | -6.562 | -0.703 | -3.744 | 2.363 | -0.696 | -3.738 | 2.348 |
| LGID | -1.117 | -2.686 | 0.468 | -1.929 | -2.253 | -1.603 | -0.794 | -3.587 | 2.019 | -0.794 | -3.582 | 2.008 |
| Med | -0.584 | -3.792 | 2.628 | -0.886 | -1.279 | -0.494 | 0.002 | -1.804 | 1.811 | 0.000 | -1.808 | 1.802 |
| Paleo | -3.003 | -9.969 | 3.968 | -2.999 | -6.909 | 0.919 | -0.195 | -2.778 | 2.394 | -0.196 | -2.769 | 2.384 |
| VD | -2.068 | -7.112 | 2.974 | -1.639 | -3.854 | 0.565 | -2.048 | -4.060 | -0.043 | -2.049 | -4.051 | -0.053 |
| BMI (*MD*, kg/m$^2$) | | | | | | | | | | | | |
| CR | -1.131 | -1.928 | -0.339 | -1.594 | -1.771 | -1.417 | -0.684 | -2.049 | 0.686 | -0.687 | -2.050 | 0.674 |
| fiber | -0.293 | -1.829 | 1.250 | -0.233 | -0.706 | 0.241 | 0.011 | -0.680 | 0.701 | 0.011 | -0.678 | 0.700 |
| HFD | -0.350 | -1.854 | 1.160 | -0.326 | -0.660 | 0.009 | -0.500 | -1.329 | 0.335 | -0.502 | -1.333 | 0.329 |
| HPD | -0.430 | -1.449 | 0.593 | -0.502 | -0.896 | -0.107 | -0.301 | -1.386 | 0.776 | -0.298 | -1.374 | 0.780 |
| LCD | -1.207 | -1.674 | -0.737 | -3.235 | -3.294 | -3.176 | -0.710 | -1.519 | 0.100 | -0.709 | -1.516 | 0.099 |
| LGID | -0.732 | -1.277 | -0.185 | -0.766 | -0.876 | -0.657 | -0.501 | -0.959 | -0.044 | -0.502 | -0.958 | -0.043 |
| Med | -0.657 | -1.444 | 0.134 | -0.862 | -0.976 | -0.747 | -1.700 | -1.970 | -1.431 | -1.700 | -1.970 | -1.431 |
| Paleo | -1.002 | -3.818 | 1.811 | -0.998 | -2.918 | 0.911 | -0.599 | -1.353 | 0.158 | -0.598 | -1.354 | 0.156 |

| Intervention compared to control | Consistent Random Effect Model | | | Consistent Fixed Effect Model | | | USE Random Effect Model | | | USE Fixed Effect Model | | |
|---|---|---|---|---|---|---|---|---|---|---|---|---|
| | Mean | Lower 95% Limit | Upper 95% Limit | Mean | Lower 95% Limit | Upper 95% Limit | Mean | Lower 95% Limit | Upper 95% Limit | Mean | Lower 95% Limit | Upper 95% Limit |
| VD | -0.690 | -1.944 | 0.570 | -0.498 | -0.773 | -0.223 | -1.110 | -1.726 | -0.493 | -1.109 | -1.725 | -0.494 |
| WC (*MD*, cm) | | | | | | | | | | | | |
| CR | -4.508 | -7.349 | -1.765 | -3.728 | -4.656 | -2.796 | -5.137 | -12.107 | 1.819 | -5.149 | -12.127 | 1.858 |
| DASH | -4.790 | -10.655 | 1.099 | -4.802 | -7.280 | -2.313 | -4.800 | -7.269 | -2.324 | -4.800 | -7.275 | -2.310 |
| fiber | -1.085 | -5.334 | 3.175 | -1.159 | -3.114 | 0.802 | -2.393 | -4.904 | 0.113 | -2.407 | -4.912 | 0.098 |
| HFD | -2.396 | -6.623 | 1.837 | -2.357 | -4.125 | -0.587 | -3.829 | -6.971 | -0.678 | -3.824 | -6.968 | -0.689 |
| HPD | 0.530 | -2.832 | 3.895 | 1.090 | -0.080 | 2.263 | -5.000 | -6.373 | -3.633 | -4.998 | -6.372 | -3.628 |
| LCD | -3.008 | -4.682 | -1.325 | -2.634 | -3.249 | -2.019 | -1.903 | -4.504 | 0.698 | -1.898 | -4.491 | 0.698 |
| LGID | -2.074 | -3.862 | -0.274 | -2.137 | -2.859 | -1.420 | -0.861 | -2.534 | 0.813 | -0.859 | -2.532 | 0.816 |
| Med | -0.766 | -4.375 | 2.869 | -0.458 | -0.927 | 0.007 | -11.397 | -13.095 | -9.694 | -11.403 | -13.107 | -9.702 |
| Paleo | -4.002 | -11.603 | 3.559 | -3.993 | -9.422 | 1.421 | -1.303 | -3.062 | 0.454 | -1.299 | -3.059 | 0.461 |
| VD | -2.355 | -6.445 | 1.723 | -2.334 | -3.868 | -0.801 | -0.202 | -1.835 | 1.430 | -0.199 | -1.839 | 1.442 |
| SBP (*MD*, mmHg) | | | | | | | | | | | | |
| CR | -1.773 | -5.667 | 2.044 | -1.341 | -3.613 | 0.938 | -5.077 | -7.280 | -2.865 | -5.075 | -7.293 | -2.873 |
| DASH | -7.584 | -14.930 | -0.289 | -7.368 | -12.869 | -1.858 | -10.497 | -19.173 | -1.807 | -10.502 | -19.141 | -1.868 |
| fiber | -1.575 | -10.445 | 7.308 | -1.614 | -7.305 | 4.107 | -5.993 | -14.612 | 2.610 | -5.985 | -14.614 | 2.661 |
| HFD | 1.156 | -4.120 | 6.420 | 1.048 | -2.307 | 4.429 | -3.703 | -9.153 | 1.753 | -3.704 | -9.142 | 1.697 |
| HPD | -2.744 | -6.293 | 0.710 | -1.906 | -4.150 | 0.340 | -0.999 | -6.510 | 4.517 | -1.002 | -6.529 | 4.520 |
| LCD | -2.211 | -4.289 | -0.101 | -2.549 | -3.777 | -1.319 | -8.285 | -15.854 | -0.685 | -8.294 | -15.920 | -0.722 |
| LGID | -0.746 | -3.657 | 2.234 | -2.518 | -4.100 | -0.933 | -0.008 | -6.912 | 6.910 | 0.000 | -6.925 | 6.919 |
| Med | -0.816 | -5.421 | 3.813 | -1.465 | -1.983 | -0.946 | -0.310 | -3.623 | 2.987 | -0.295 | -3.596 | 2.998 |
| Paleo | -8.948 | -24.228 | 6.354 | -8.968 | -22.556 | 4.660 | -9.873 | -15.836 | -3.893 | -9.866 | -15.872 | -3.886 |
| VD | -0.085 | -6.170 | 6.108 | -0.838 | -4.990 | 3.324 | -1.596 | -7.275 | 4.106 | -1.595 | -7.320 | 4.129 |
| DBP (*MD*, mmHg) | | | | | | | | | | | | |
| CR | -1.968 | -5.198 | 1.226 | -1.645 | -3.257 | -0.042 | -1.564 | -2.799 | -0.331 | -1.564 | -2.802 | -0.325 |
| DASH | -3.730 | -10.331 | 2.814 | -3.018 | -7.575 | 1.542 | -8.778 | -16.988 | -0.541 | -8.769 | -16.987 | -0.580 |
| fiber | -0.723 | -8.166 | 6.747 | -0.706 | -4.408 | 2.982 | -1.013 | -5.918 | 3.890 | -0.980 | -5.883 | 3.925 |
| HFD | 0.761 | -3.696 | 5.220 | 0.978 | -1.371 | 3.324 | -1.201 | -4.545 | 2.151 | -1.205 | -4.556 | 2.138 |
| HPD | -2.972 | -5.920 | -0.068 | -1.993 | -3.435 | -0.555 | -1.005 | -4.856 | 2.844 | -1.014 | -4.864 | 2.849 |
| LCD | -1.944 | -3.783 | -0.069 | -2.895 | -3.829 | -1.963 | -7.388 | -11.972 | -2.840 | -7.391 | -11.948 | -2.833 |

| Intervention compared to control | Consistent Random Effect Model | | | Consistent Fixed Effect Model | | | USE Random Effect Model | | | USE Fixed Effect Model | | |
|---|---|---|---|---|---|---|---|---|---|---|---|---|
| | Mean | Lower 95% Limit | Upper 95% Limit | Mean | Lower 95% Limit | Upper 95% Limit | Mean | Lower 95% Limit | Upper 95% Limit | Mean | Lower 95% Limit | Upper 95% Limit |
| LGID | -0.827 | -3.310 | 1.664 | -1.198 | -2.119 | -0.276 | -0.005 | -4.286 | 4.304 | 0.002 | -4.299 | 4.293 |
| Med | -0.477 | -4.601 | 3.660 | -1.347 | -1.788 | -0.908 | 1.907 | -1.396 | 5.207 | 1.899 | -1.403 | 5.197 |
| Paleo | -3.980 | -13.220 | 5.260 | -3.986 | -10.631 | 2.649 | -7.450 | -11.881 | -2.991 | -7.443 | -11.894 | -3.018 |
| VD | 0.989 | -3.935 | 5.946 | 0.529 | -1.958 | 3.015 | -0.704 | -4.398 | 2.991 | -0.693 | -4.390 | 3.012 |
| TG (*MD*, mmol/L) | | | | | | | | | | | | |
| CR | -0.113 | -0.293 | 0.066 | -0.117 | -0.210 | -0.024 | -0.040 | -0.319 | 0.240 | -0.041 | -0.321 | 0.239 |
| DASH | -0.040 | -0.530 | 0.450 | -0.039 | -0.319 | 0.239 | -0.120 | -0.328 | 0.089 | -0.120 | -0.328 | 0.091 |
| fiber | -0.186 | -0.501 | 0.131 | -0.208 | -0.388 | -0.027 | -0.293 | -0.986 | 0.401 | -0.296 | -0.992 | 0.406 |
| HFD | -0.176 | -0.515 | 0.162 | -0.177 | -0.430 | 0.077 | 0.098 | -0.439 | 0.634 | 0.101 | -0.435 | 0.638 |
| HPD | -0.233 | -0.464 | -0.003 | -0.224 | -0.386 | -0.062 | 0.045 | -0.498 | 0.590 | 0.045 | -0.497 | 0.590 |
| LCD | -0.288 | -0.396 | -0.179 | -0.328 | -0.391 | -0.266 | 0.850 | -3.860 | 5.559 | 0.856 | -3.850 | 5.544 |
| LGID | -0.254 | -0.353 | -0.154 | -0.285 | -0.331 | -0.238 | 0.100 | -0.655 | 0.852 | 0.098 | -0.657 | 0.854 |
| Med | -0.201 | -0.408 | 0.005 | -0.206 | -0.257 | -0.155 | -0.010 | -0.325 | 0.305 | -0.010 | -0.324 | 0.304 |
| Paleo | -0.500 | -1.128 | 0.129 | -0.499 | -0.987 | -0.011 | 0.202 | -0.991 | 1.396 | 0.199 | -0.994 | 1.389 |
| VD | -0.024 | -0.360 | 0.310 | -0.007 | -0.242 | 0.227 | -0.237 | -0.959 | 0.483 | -0.236 | -0.954 | 0.484 |
| TC (*MD*, mmol/L) | | | | | | | | | | | | |
| CR | -0.170 | -0.426 | 0.089 | -0.158 | -0.250 | -0.067 | 0.327 | -0.227 | 0.878 | 0.328 | -0.224 | 0.882 |
| DASH | -0.357 | -1.134 | 0.420 | -0.356 | -0.786 | 0.073 | -0.357 | -0.789 | 0.074 | -0.356 | -0.785 | 0.076 |
| fiber | -0.328 | -0.764 | 0.107 | -0.290 | -0.554 | -0.029 | -0.031 | -0.622 | 0.555 | -0.031 | -0.615 | 0.556 |
| HFD | -0.116 | -0.516 | 0.282 | -0.077 | -0.287 | 0.133 | -0.175 | -0.510 | 0.158 | -0.177 | -0.512 | 0.158 |
| HPD | -0.150 | -0.440 | 0.141 | -0.125 | -0.247 | -0.003 | -0.011 | -0.666 | 0.647 | -0.010 | -0.666 | 0.647 |
| LCD | -0.165 | -0.318 | -0.012 | -0.212 | -0.283 | -0.142 | 0.078 | -0.337 | 0.491 | 0.078 | -0.336 | 0.491 |
| LGID | -0.461 | -0.619 | -0.304 | -0.504 | -0.587 | -0.422 | -0.003 | -6.926 | 6.916 | -0.266 | -7.298 | 6.696 |
| Med | -0.177 | -0.474 | 0.122 | -0.166 | -0.213 | -0.119 | -0.100 | -0.398 | 0.196 | -0.099 | -0.397 | 0.198 |
| Paleo | -0.198 | -1.252 | 0.867 | -0.199 | -1.040 | 0.646 | -0.100 | -1.053 | 0.857 | -0.103 | -1.056 | 0.853 |
| VD | -0.104 | -0.581 | 0.372 | -0.119 | -0.358 | 0.118 | -0.362 | -0.969 | 0.244 | -0.362 | -0.968 | 0.243 |
| LDL (*MD*, mmol/L) | | | | | | | | | | | | |
| CR | -0.242 | -0.443 | -0.039 | -0.410 | -0.518 | -0.302 | -0.372 | -0.673 | -0.071 | -0.371 | -0.674 | -0.069 |
| DASH | -0.371 | -0.891 | 0.152 | -0.373 | -0.674 | -0.071 | -0.040 | -0.318 | 0.239 | -0.040 | -0.319 | 0.238 |
| fiber | -0.235 | -0.578 | 0.106 | -0.221 | -0.454 | 0.012 | -0.106 | -0.423 | 0.212 | -0.107 | -0.425 | 0.209 |

| Intervention compared to control | Consistent Random Effect Model | | | Consistent Fixed Effect Model | | | USE Random Effect Model | | | USE Fixed Effect Model | | |
|---|---|---|---|---|---|---|---|---|---|---|---|---|
| | Mean | Lower 95% Limit | Upper 95% Limit | Mean | Lower 95% Limit | Upper 95% Limit | Mean | Lower 95% Limit | Upper 95% Limit | Mean | Lower 95% Limit | Upper 95% Limit |
| HFD | -0.067 | -0.347 | 0.212 | -0.050 | -0.217 | 0.118 | -0.261 | -0.827 | 0.307 | -0.259 | -0.828 | 0.305 |
| HPD | -0.110 | -0.309 | 0.088 | -0.079 | -0.189 | 0.030 | 0.000 | -0.380 | 0.381 | 0.000 | -0.381 | 0.382 |
| LCD | -0.108 | -0.219 | 0.004 | -0.119 | -0.182 | -0.055 | -0.470 | -3.810 | 2.864 | -0.463 | -3.794 | 2.866 |
| LGID | -0.352 | -0.466 | -0.237 | -0.332 | -0.393 | -0.271 | -0.100 | -0.339 | 0.140 | -0.099 | -0.340 | 0.141 |
| Med | -0.029 | -0.268 | 0.209 | 0.000 | -0.120 | 0.120 | 0.000 | -1.051 | 1.051 | -0.001 | -1.056 | 1.048 |
| Paleo | -0.102 | -0.920 | 0.716 | -0.100 | -0.801 | 0.598 | -0.233 | -0.762 | 0.298 | -0.232 | -0.762 | 0.300 |
| VD | -0.060 | -0.360 | 0.240 | -0.060 | -0.230 | 0.110 | 0.032 | -0.280 | 0.346 | 0.033 | -0.278 | 0.344 |
| HDL (*MD*, mmol/L) | | | | | | | | | | | | |
| CR | 0.084 | 0.008 | 0.160 | 0.130 | 0.105 | 0.155 | 0.131 | 0.091 | 0.170 | 0.131 | 0.091 | 0.170 |
| DASH | 0.081 | -0.150 | 0.312 | 0.081 | 0.023 | 0.139 | 0.081 | 0.023 | 0.139 | 0.081 | 0.023 | 0.139 |
| fiber | 0.078 | -0.059 | 0.215 | 0.036 | -0.012 | 0.084 | 0.190 | 0.075 | 0.304 | 0.190 | 0.076 | 0.305 |
| HFD | 0.039 | -0.074 | 0.153 | 0.025 | -0.013 | 0.063 | 0.008 | -0.092 | 0.109 | 0.007 | -0.093 | 0.107 |
| HPD | -0.017 | -0.105 | 0.072 | -0.017 | -0.046 | 0.013 | 0.000 | -0.062 | 0.063 | 0.000 | -0.063 | 0.063 |
| LCD | 0.120 | 0.073 | 0.167 | 0.131 | 0.114 | 0.147 | -0.001 | -0.161 | 0.160 | 0.000 | -0.161 | 0.161 |
| LGID | 0.080 | 0.028 | 0.132 | 0.087 | 0.070 | 0.105 | 0.100 | -0.170 | 0.366 | 0.099 | -0.170 | 0.367 |
| Med | 0.062 | -0.039 | 0.162 | 0.054 | 0.041 | 0.067 | 0.340 | 0.269 | 0.411 | 0.340 | 0.270 | 0.411 |
| Paleo | 0.080 | -0.178 | 0.338 | 0.080 | -0.050 | 0.210 | -0.099 | -0.819 | 0.620 | -0.098 | -0.817 | 0.626 |
| VD | -0.045 | -0.170 | 0.079 | -0.042 | -0.087 | 0.003 | -0.026 | -0.117 | 0.065 | -0.026 | -0.117 | 0.065 |

Blue cells: the upper limit of 95% *CrI* smaller than the null value; Red cells: the lower limit of 95% *CrI* greater than the null value.

**Table S12.3** Model Fit

| Outcomes | Data Points | Consistent Random Effect Model | | | Consistent Fixed Effect Model | | | USE Random Effect Model | | | USE Fixed Effect Model | | |
|---|---|---|---|---|---|---|---|---|---|---|---|---|---|
| | | Dbar | pD | DIC | Dbar | pD | DIC | Dbar | pD | DIC | Dbar | pD | DIC |
| FPG | 186 | 177.87 | 151.56 | 329.43 | 439.04 | 102.01 | 541.05 | 185.92 | 185.92 | 371.84 | 185.98 | 185.98 | 371.96 |
| $HbA_{1c}$ | 182 | 177.35 | 155.49 | 332.84 | 508.48 | 100.02 | 608.50 | 181.97 | 181.97 | 363.94 | 182.00 | 182.00 | 364.00 |
| FIns | 71 | 65.78 | 59.21 | 124.99 | 184.08 | 42.97 | 227.05 | 71.02 | 71.02 | 142.04 | 71.00 | 71.00 | 142.00 |
| IR | 43 | 41.16 | 31.57 | 72.73 | 46.94 | 27.00 | 73.93 | 43.02 | 43.02 | 86.05 | 43.01 | 43.01 | 86.02 |
| weight | 137 | 135.02 | 126.78 | 261.80 | 3373.73 | 77.00 | 3450.73 | 136.97 | 136.97 | 273.95 | 136.98 | 136.97 | 273.95 |
| BMI | 126 | 124.81 | 118.62 | 243.43 | 5556.90 | 71.03 | 5627.93 | 125.97 | 125.97 | 251.94 | 125.94 | 125.94 | 251.89 |
| WC | 84 | 83.00 | 75.19 | 158.19 | 254.85 | 51.02 | 305.87 | 84.06 | 84.06 | 168.12 | 84.01 | 84.01 | 168.02 |
| SBP | 108 | 109.21 | 85.56 | 194.76 | 163.13 | 63.03 | 226.16 | 107.94 | 107.94 | 215.87 | 108.00 | 108.00 | 216.00 |
| DBP | 103 | 105.01 | 88.73 | 193.73 | 213.00 | 60.94 | 273.94 | 102.93 | 102.93 | 205.86 | 102.92 | 102.92 | 205.85 |
| TG | 173 | 169.72 | 134.01 | 303.73 | 275.57 | 94.97 | 370.54 | 172.59 | 172.59 | 345.18 | 172.68 | 172.67 | 345.35 |
| TC | 171 | 171.73 | 142.96 | 314.69 | 359.82 | 93.98 | 453.80 | 170.64 | 170.63 | 341.27 | 170.62 | 170.62 | 341.24 |
| LDL | 163 | 157.01 | 129.34 | 286.35 | 288.36 | 89.98 | 378.35 | 162.79 | 162.79 | 325.58 | 162.71 | 162.71 | 325.42 |
| HDL | 171 | 169.04 | 153.88 | 322.92 | 645.87 | 93.96 | 739.83 | 170.91 | 170.91 | 341.82 | 170.98 | 170.98 | 341.96 |

Dbar, the posterior mean of the deviance; pD, the effective number of parameters; DIC, Deviance Information Criterion.

### File S13: Publication Bias

#### S13.1 Comparison-adjusted funnel plots

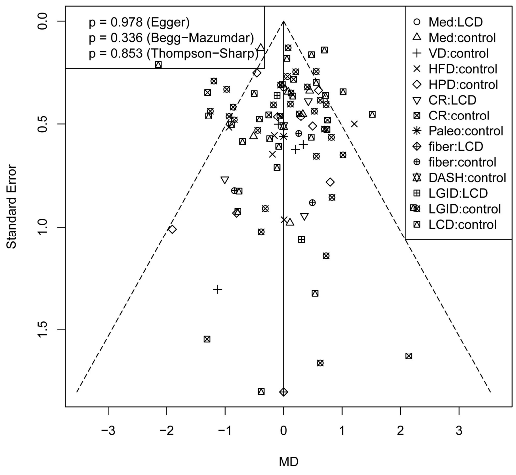

(A) FPG

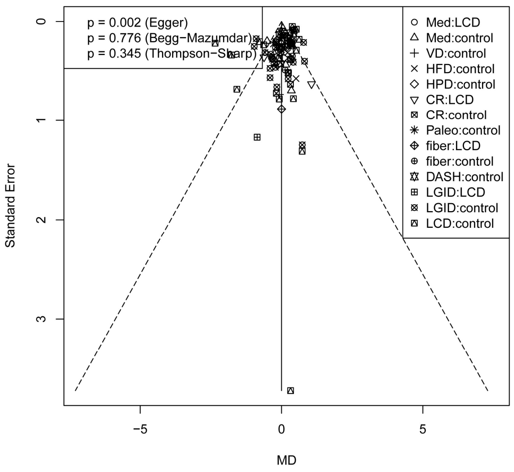

(B) $HbA_{1c}$

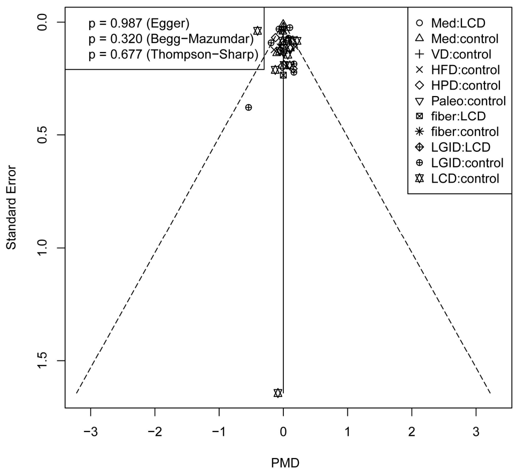

(C) FIns

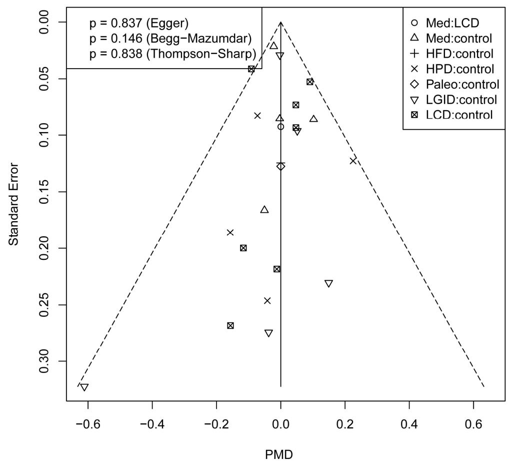

(D) IR

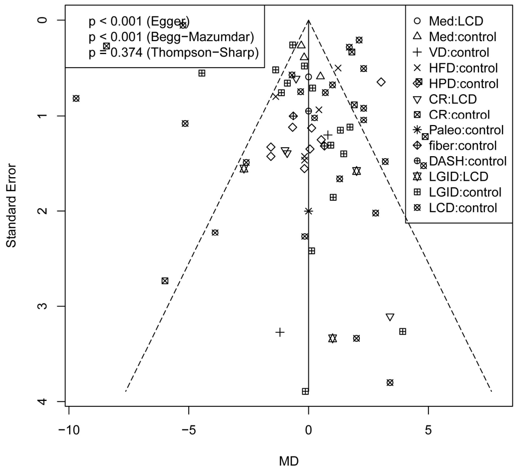

(E) Weight

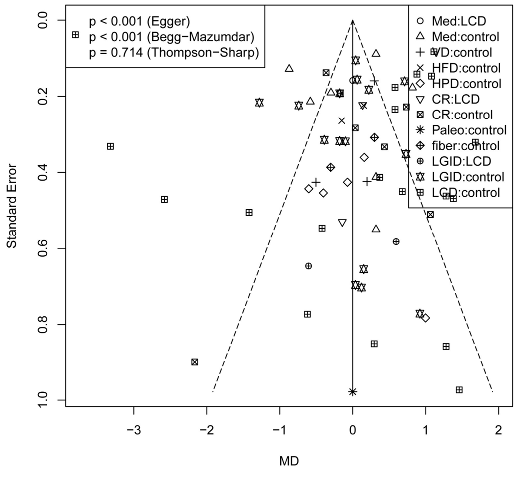

(F) BMI

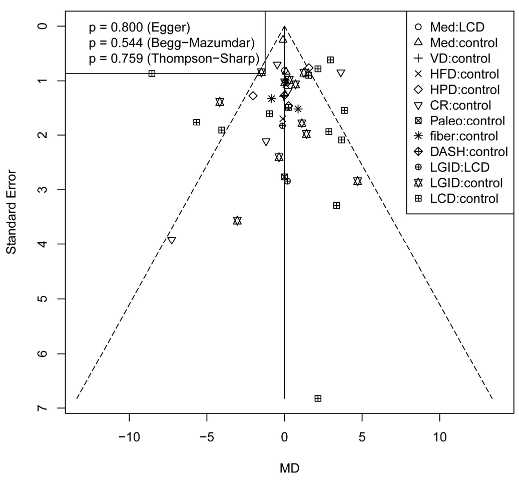

(G) WC

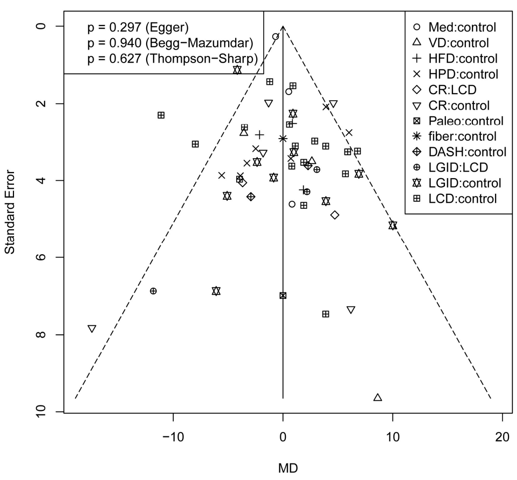

(H) SBP

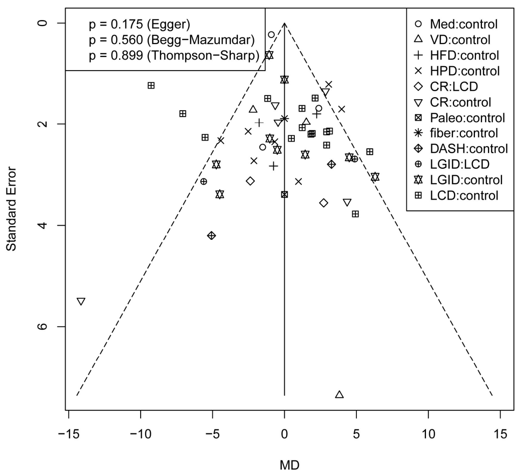

(I) DBP

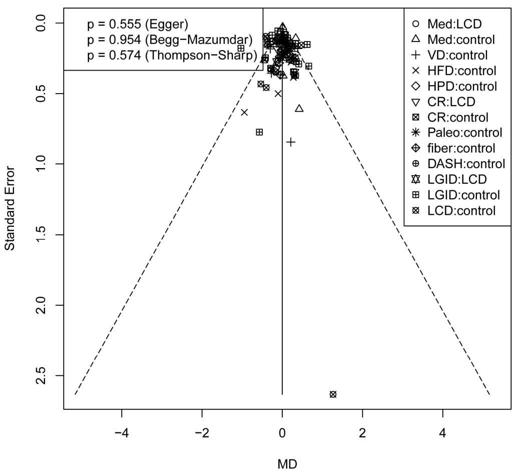

(J) TG

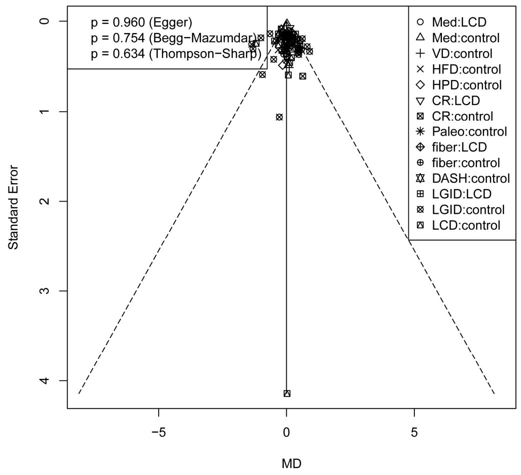

(K) TC

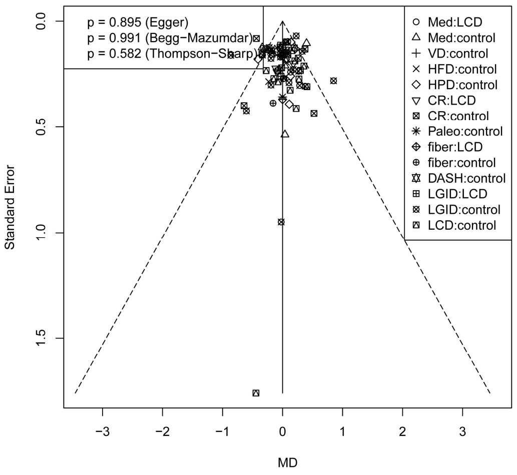

(L) LDL

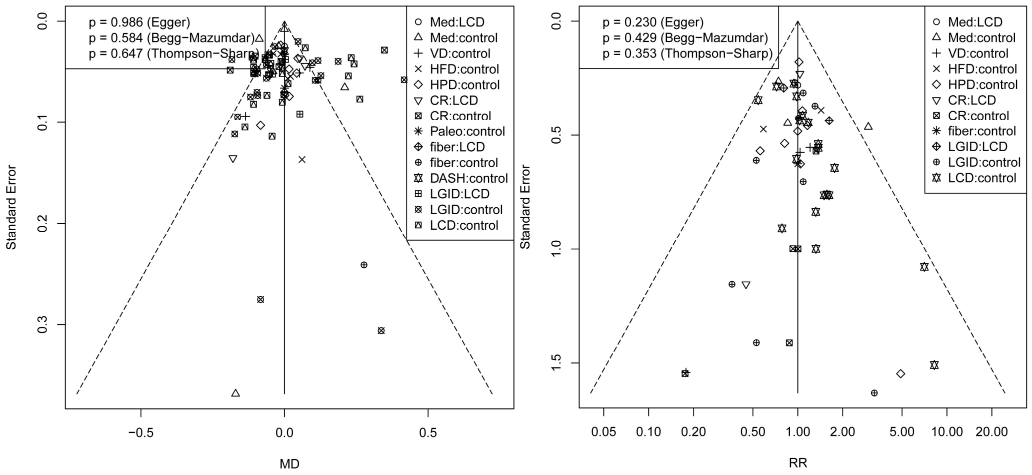

(M) HDL (N) Attrition

**Figure S13.1** Comparison-adjusted funnel plots

(A) FPG; (B) $HbA_{1c}$; (C) FIns; (D) IR; (E) weight; (F) BMI; (G) WC; (H) SBP; (I) DBP; (J) TG; (I) TC; (L) LDL; (M) HDL; (N) Attrition.

#### S13.2 Hypothesis testing

**Table S13.1** *P* values of hypothesis testing

| Outcome | *P* value | | |
|---|---|---|---|
| | Egger's | Begg's | Thompson-Sharp |
| FPG | 0.978 | 0.336 | 0.853 |
| $HbA_{1c}$ | 0.002* | 0.776 | 0.345 |
| FIns | 0.987 | 0.320 | 0.677 |
| IR | 0.837 | 0.146 | 0.838 |
| weight | <0.001* | <0.001* | 0.374 |
| BMI | <0.001* | <0.001* | 0.714 |
| WC | 0.800 | 0.544 | 0.759 |
| SBP | 0.297 | 0.940 | 0.627 |
| DBP | 0.175 | 0.560 | 0.899 |
| TG | 0.555 | 0.954 | 0.574 |
| TC | 0.960 | 0.754 | 0.634 |
| LDL | 0.895 | 0.991 | 0.582 |
| HDL | 0.986 | 0,584 | 0.647 |
| Attrition | 0.230 | 0.429 | 0.353 |

* *P* value < 0.05.

#### File S14: Minimal clinically important difference and thresholds for effects

MCIDs were identified based on the following criteria and the previous studies (see the main text Section 3.8):

a. Approx. 5% of the upper limit of the normal value of glycemic indicators and lipid profiles;

b. Approx. 5% of the baseline of anthropometric indicators;

Thresholds for small to moderate and moderate to large were estimated as 1.5 to 3 times of MCID, or 10% to 25% of the upper limit of the normal value.

**Table S14.1** MCID and thresholds for effects

| Outcome | Type | Unit | Thresholds (Absolute Value) | | |
|---|---|---|---|---|---|
| | | | MCID | Small to Moderate | Moderate to Large |
| FPG | *MD* | mmol/L | 0.80 | 1.40 | 1.80 |
| $HbA_{1c}$ | *MD* | % | 0.50 | 0.90 | 1.40 |
| FIns | *PMD* | - | 8% | 12% | 16% |
| IR | *PMD* | - | 5% | 8% | 12% |
| weight | *MD* | kg | 3.00 | 5.00 | 7.00 |
| BMI | *MD* | $kg/m^2$ | 1.05 | 1.55 | 1.85 |
| WC | *MD* | cm | 4.50 | 7.00 | 12.00 |
| SBP | *MD* | mmHg | 6.00 | 10.00 | 15.00 |
| DBP | *MD* | mmHg | 3.50 | 7.00 | 10.00 |
| TG | *MD* | mmol/L | 0.09 | 0.15 | 0.25 |
| TC | *MD* | mmol/L | 0.26 | 0.40 | 0.52 |
| LDL | *MD* | mmol/L | 0.10 | 0.25 | 0.40 |
| HDL | *MD* | mmol/L | 0.10 | 0.15 | 0.20 |
